# Cardiovascular magnetic resonance reference ranges and centile-based categories for atrial and ventricular cardiac structure and function: 2026 Update from the Healthy Hearts Consortium

**DOI:** 10.1101/2025.04.07.25325351

**Authors:** Liliana Szabo, Dorina-Gabriela Condurache, Janek Salatzki, Robin Bülow, Giovanni Donato Aquaro, Florian Andre, Le Thu-Thao, Dominika Suchá, Ahmed M. Salih, Roman Roy, Nay Aung, Sucharitha Chadalavada, Kerrik Hesse, Leo Westbury, Nicholas C. Harvey, Tim Leiner, Calvin W.L. Chin, Matthias G. Friedrich, Andrea Barison, Marcus Dörr, Zahra Raisi-Estabragh, Steffen E. Petersen

**Affiliations:** William Harvey Research Institute, NIHR Barts Biomedical Research Centre, Queen Mary University of London, Charterhouse Square, London, EC1M 6BQ, UK; Semmelweis University, Heart and Vascular Center, Budapest, Hungary; Barts Heart Centre, St Bartholomew’s Hospital, Barts Health NHS Trust, West Smithfield, London, EC1A 7BE, UK; Department of Cardiology, Angiology and Pneumology, University of Heidelberg, Im Neuenheimer Feld 410, Heidelberg, 69120, Germany; Institute for Diagnostic Radiology and Neuroradiology, University Medicine, Ernst Moritz Arndt University Greifswald, Ferdinand-Sauerbruch-Straße 1, 17475 Greifswald, Germany; Academic Radiology, Department of Surgical, Medical and Molecular Pathology and of critical area, University of Pisa, Italy; National Heart Centre Singapore, 5 Hospital Drive, Singapore 169609, Singapore; Cardiovascular ACP, Duke-NUS Medical School, Singapore; University Medical Center Utrecht, Department of Radiology and Nuclear Medicine, Utrecht, the Netherlands; Division of Cardiovascular Sciences, University of Leicester, British Heart Foundation (BHF) Leicester Centre of Research Excellence, University of Leicester, UK; Lifecourse Epidemiology Centre, Human Development and Health, University of Southampton, Southampton, SO16 6YD, UK; NIHR Southampton Biomedical Research Centre, University of Southampton and University Hospital Southampton NHS Foundation Trust, Southampton, SO16 6YD, UK; Mayo Clinic, Department of Radiology, Rochester, Minnesota, USA; Department of Medicine and Diagnostic Radiology, McGill University, Montreal, Quebec, Canada; Cardiology and Cardiovascular Medicine, Fondazione Toscana Gabriele Monasterio, Pisa, Italy; Institute for Community Medicine, SHIP/KEF, University Medicine Greifswald,17475 Greifswald, Germany; DZHK (German Centre for Cardiovascular Research), partner site North, Germany; Ambulantes Herz-Kreislauf-Zentrum, Zentrum für kardiovaskuläre Telemedizin, 17309 Pasewalk, Germany

**Keywords:** Cardiovascular magnetic resonance, centile-based reference ranges, healthy adults, automated image analysis, multicentre study

## Abstract

**Background:** Cardiovascular magnetic resonance (CMR) provides accurate, reproducible quantification of cardiac chamber size and function. Population-specific reference ranges, standardized categories for grading degree of deviation, and evidence for their prognostic relevance are needed to enhance clinical utility. Using the Healthy Hearts Consortium (HHC), we derived centile-based reference ranges and deviation categories for ventricular and atrial CMR measures across multiple demographic groups and two software platforms, and evaluated their prognostic relevance in a large longitudinal cohort.

**Methods:** We analyzed 9,059 CMR studies from the HHC, comprising healthy adults aged 18–83 years from six international cohorts. Images were processed automatically with cvi42 (Circle Cardiovascular Imaging) and suiteHEART (NeoSoft) to derive left and right ventricular volumes, ejection fraction and mass, and atrial volumes and function. Age- and sex-specific centile curves were estimated using generalized additive models for location, scale, and shape. Reference ranges were defined by the 2.5th–97.5th percentiles; values in the 0.5th–2.5th or 97.5th–99.5th percentiles were classified as mild deviation, and those beyond the 0.5th or 99.5th percentiles as at least moderate deviation. Associations of these categories with incident cardiovascular events over a median 4-year follow-up were evaluated in 59,916 UK Biobank participants.

**Results:** The reference cohort included 4,624 women (51.0%) and 4,435 men (49.0%), mean age 61±13 years. Cardiac anatomy and function varied systematically with age and sex, enabling categories beyond conventional reference limits. Measurements from cvi42 and suiteHEART were comparable for ventricular parameters. In the UK Biobank Imaging Substudy, progression from within reference range to mildly and at least moderately out of range showed stepwise increases in cardiovascular risk across left and right ventricular metrics. Associations were strongest for heart failure and major cardiovascular disease; an at least moderately out-of-range LVEF conferred higher risk of incident heart failure (HR 8.81, 95% CI 7.08–10.97) and major cardiovascular disease (HR 2.74, 95% CI 2.32–3.23).

**Conclusions:** We present reference ranges and centile-based categories for ventricular and atrial CMR measures from a large multi-ethnic reference population. These categories had demonstrable prognostic significance, identifying individuals at higher risk of major cardiovascular outcomes, and could support more standardized, clinically meaningful CMR reporting.

**Graphical abstract:** 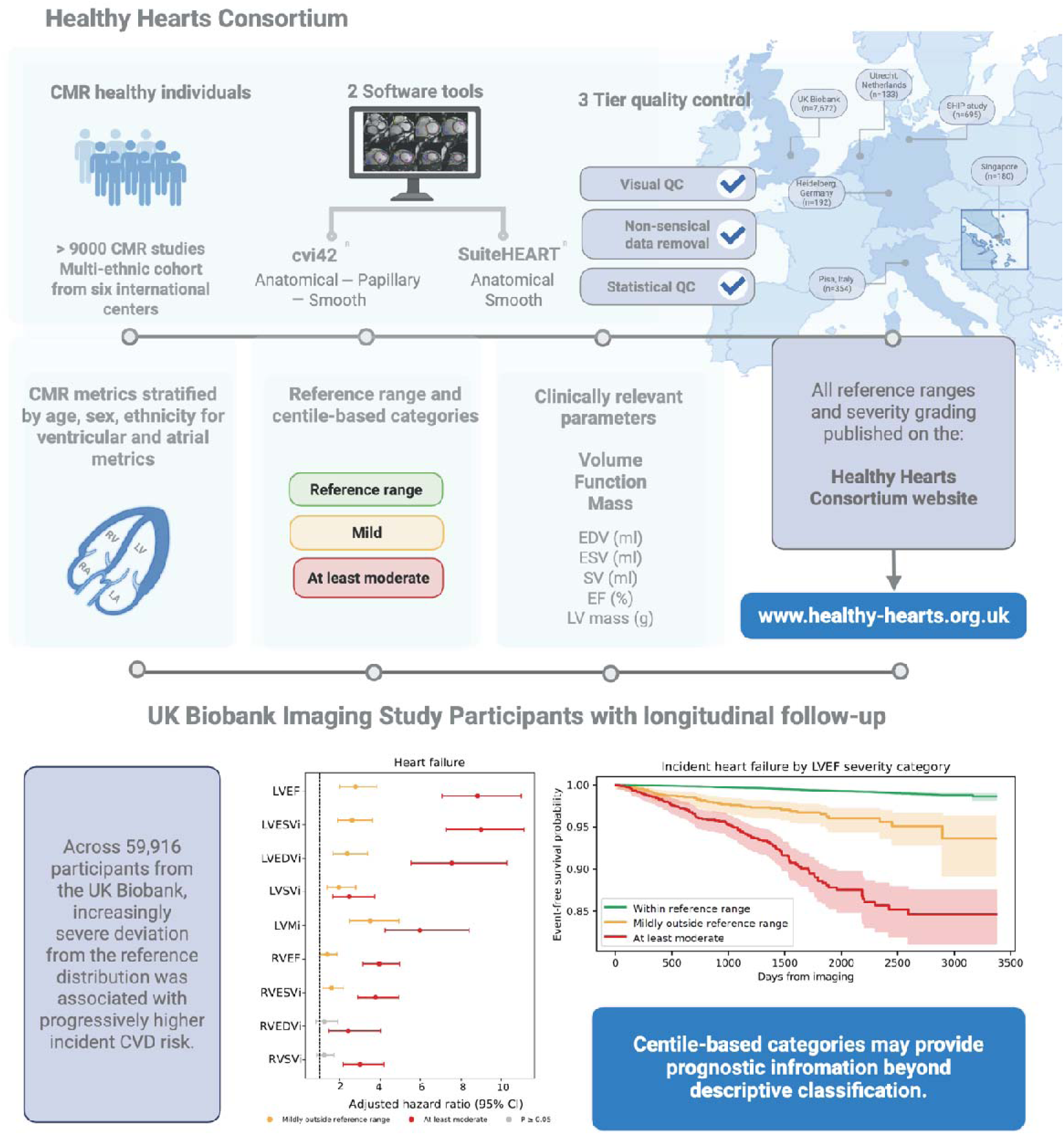

**Footnote**: In this study, we established CMR reference ranges and centile-based categories for cardiac structure and function using data from over 9,000 healthy individuals in the Healthy Hearts Consortium, and demonstrated in the UK Biobank that greater deviation from the reference distribution was associated with progressively higher incident cardiovascular risk.

## Introduction

Cardiovascular magnetic resonance (CMR) imaging is the reference standard for quantifying cardiac chamber size, function, and tissue composition [1, 2]. Because clinical decisions rest on comparing an individual’s results against expected values, the usefulness of CMR depends directly on the quality of its reference ranges. Yet existing reference ranges remain limited: they are derived from heterogeneous populations, generated under varying technical and post-processing conditions, and typically expressed as fixed normal/abnormal thresholds that indicate whether a value is atypical but not how far it departs from the healthy distribution. Standardized approaches for quantifying this degree of deviation in CMR are still lacking [3, 4].

Efforts to establish CMR reference values have a substantial history. Society-endorsed normal values and several single- and multi-centre cohort studies have provided widely used reference data[1], and large population studies such as UK Biobank or Canadian Alliance for Healthy Hearts and Minds have generated sex- and age-specific ranges from thousands of participants [5, 6]. Methodologically, most of these ranges rest on descriptive statistics, typically means and standard deviations within age and sex strata [1], rather than formal modelling of how the healthy distribution itself varies across continuous factors such as age.

Addressing these gaps requires large-scale data able to model the healthy distribution robustly rather than merely describe it. Having previously shown that published CMR reference ranges vary substantially with biological and technical differences [4] we established the Healthy Hearts Consortium (HHC) to assemble a healthy, demographically diverse dataset large enough to capture the key population drivers of cardiac variation. The HHC includes more than 9,000 CMR examinations from verified healthy adults spanning a broad age range and diverse ethnic backgrounds, providing a unique basis for robust reference modelling. Advances in automated deep learning-based segmentation now allow this volume of CMR data to be measured reproducibly and with minimal operator dependence [7], which manual analysis could not support in the past.

Here we present a substantial extension of our earlier work [8], drawing on the same resource. We aimed to: (1) derive continuous, centile-based reference ranges for key CMR metrics using generalized additive models for location, scale, and shape (GAMLSS); (2) generate these ranges across two widely used post-processing platforms; and (3) test the clinical value of this approach in UK Biobank participants, examining whether deviation from the age-, sex-, and ethnicity-matched reference range predicts subsequent cardiovascular outcomes. By matching patients to the most appropriate reference ranges and centile-based categories for their age, sex, and ethnicity, this work aims to make quantitative CMR more interpretable and to support better clinical decisions.

## Methods

### Healthy Hearts Consortium

The HHC is an international collaborative comprising over 9,000 CMR examinations from healthy individuals. The consortium, described in detail elsewhere [8], includes participants from six contributing cohorts: Utrecht (The Netherlands) [9], Pisa (Italy) [10], Heidelberg (Germany) [11], Singapore [12], the Study of Health in Pomerania (SHIP, Germany) [13], and the UK Biobank [6]. Further information on the HHC resource is available in **Table S1** and on the linked website: https://www.healthy-hearts.org.uk.

### CMR image segmentation

We used two commercially available software solutions with batch processing capabilities: cvi42 (version 5.11.2, Circle Cardiovascular Imaging, Calgary, Alberta, Canada) and suiteHEART (version 5.0.0, Neosoft, Pewaukee, Wisconsin, USA). Both software packages generated comprehensive measurements of left and right ventricular and atrial volumes and function. We employed a fully automated segmentation workflow to ensure reproducibility without manual adjustments. The segmentation algorithms delineated the endocardial and epicardial contours in standard short-axis and long-axis (2-, 3- and 4-chamber) cine views.

Short-axis images covering the left and right ventricles were processed according to predefined protocols endorsed by the Society for Cardiovascular Magnetic Resonance (SCMR) [14]. In cvi42, three segmentation methods were used to contour the endocardial and epicardial borders of the ventricles: 1) Smooth: endocardial contours excluding papillary muscles and trabeculation from the left ventricular (LV) mass (included in the blood pool). 2) Papillary: smooth endocardial contours including papillary muscles (excluding trabecular tissue) in the LV mass. 3) Anatomical: papillary muscles and trabecular tissue are included in the LV mass. Only smooth and anatomical segmentation approaches are supported by suiteHEART. Atrial segmentation was uniformly performed on long-axis images.

### Quality control

We implemented a multistep quality control (QC) approach, including visual and statistical QC. First, we conducted manual visual QC using a previously described scoring system in the whole HHC cohort [8]. Each chamber’s image acquisition, planning, and segmentation quality was assigned one of three scores: 1 = ‘perfect’, 2 = ‘satisfactory’, and 3 = ‘unacceptable for clinical use’. All metrics derived from chamber views receiving a score of 3 were excluded from further analysis. Following visual QC, nonsensical values were removed based on objective criteria (detailed in **Figures S1 and S2**). Finally, statistical QC was applied to remove biologically implausible values, using the 3-SD method to identify and remove extreme values across the dataset.

### Derivation of CMR metrics

LV and right ventricular (RV) parameters were derived from the full short-axis stack. Ventricular volumes were determined using the ’sum of discs’ method. Atrial parameters were obtained using the biplane method based on long-axis imaging, deriving left atrial (LA) metrics from the 2- and 4-chamber views with the area-length method, and right atrial (RA) metrics from the 4-chamber view.

We report the following metrics as per their availability in the software solutions: LV cardiac output (LVCO), LV end-diastolic volume (LVEDV), LV end-systolic volume (LVESV), LV stroke volume (LVSV), LV ejection fraction (LVEF), LV myocardial mass in end-diastole (LVM), LVM/LVEDV ratio, RV cardiac output (RVCO), RV end-diastolic volume (RVEDV), RV end-systolic volume (RVESV), RV stroke volume (RVSV), RV ejection fraction (RVEF), LA end-systolic volume (LAESV), LA ejection fraction (LAEF), LA maximal volume (LAV), RA end-systolic volume (RAESV), RA ejection fraction (RAEF), and RA maximal volume (RAV).

### Body size adjustments

We present volumetric and mass measurements indexed to both body surface area (BSA) and height, consistent with previous work. Height and weight, measured at the time of imaging, were used to calculate BSA using the Mosteller formula (BSA = √[height (cm) × weight (kg) / 3600]). The main manuscript presents BSA-indexed values, while BSA- and height-indexed reference ranges are provided in the **Supplemental material** and on our project website: https://www.healthy-hearts.org.uk.

### Stratification data

Each HHC contributor provided details on the CMR scanner vendor and magnetic field strength (1.5 Tesla or 3.0 Tesla). Participant age was recorded at the time of imaging. Sex and ethnicity were self-reported. The cohorts from Utrecht, Heidelberg, and SHIP comprised individuals of White/Caucasian ethnicity. The Pisa cohort included one individual of Black ethnicity, and the remainder were of White/Caucasian ethnic backgrounds. The Singapore cohort contained only individuals of Chinese descent. Participants in the UK Biobank were assigned to the following ethnic categories: “White,” “Black or Black British,” “Asian or Asian British,” “Chinese,” “Mixed,” and “Other”. According to UK government guidance, “Asian or Asian British” encompasses Indian, Pakistani, Bangladeshi, or “any other Asian” backgrounds; for simplicity, we label this group “South Asian”[15].

### Incident outcomes associations in UK Biobank

Among UK Biobank participants with available CMR imaging, we performed a complete-case analysis, retaining only those with cvi42-derived measurements (smooth segmentation method), complete covariate data, and linked outcome data. This yielded a final validation cohort of 59,916 participants. We evaluated the following set of available ventricular CMR measures: LVEDV, LVESV, LVSV, LVM, LVEF, RVEDV, RVESV, RVSV, and RVEF. We applied BSA indexation to volumes and mass. Age- and sex-specific reference ranges and percentile-based thresholds were taken from the previously derived cvi42 smooth reference tables. Participants were assigned to the relevant age stratum and sex category, and each observed CMR value was mapped to the corresponding reference interval described in the statistical methods section. Cardiovascular events were ascertained through linked hospital inpatient data (based on relevant ICD-10 codes). Outcomes included heart failure, myocardial infarction, ischemic heart disease, valvular heart disease, atrial fibrillation, and non-ischemic cardiomyopathy. A composite major adverse cardiovascular events (MACE) endpoint was also defined from these component outcomes. We also defined all-cause mortality and cardiovascular mortality. Baseline was defined as the imaging assessment date. For each endpoint, the first recorded occurrence date was used. Participants with evidence of the relevant outcome on or before the imaging date were excluded from the corresponding incident outcome analysis. Follow-up was until the first occurrence of the endpoint of interest, death, or administrative censoring on 31 March 2023.

## Statistical analysis

Statistical analysis was conducted using R version 4.5.1 and Python version 3.14. Descriptive statistics, including mean, standard deviation (SD), and range, were used to summarize the dataset. Age was categorized into the following groups: 18-29, 30-39, 40-49, 50-59, 60-69, and over 70 years (ages inclusive). To examine associations between ethnicity and CMR metrics, we fitted a series of linear regression models for each variable. This exemplary analysis was restricted to results derived using smooth segmentation and adjusted for BSA for simplicity; however, other segmentation types are expected to yield comparable results. The outcome variable was standardized (z-score) within the full dataset to enable interpretation of effect sizes across metrics. Each model included ethnicity (categorical, with White individuals as the reference group), age (standardized), and sex as covariates. Confidence intervals (CI) were calculated at the 95% level, and p-values were adjusted for multiple comparisons across all models using the Benjamini-Hochberg false discovery rate (FDR) method. Statistical significance was defined as an FDR-adjusted *p*-value < 0.05.

Agreement between cvi42 and suiteHEART was assessed using Bland–Altman analysis of paired measurements from the same subjects, analyzed with both software packages. For each cardiac metric, the difference (suiteHEART − cvi42) was plotted against the mean of the two measurements. The bias was calculated as the mean of the paired differences, and the 95% limits of agreement as the bias ± 1.96·SD of the differences, assuming approximately normally distributed differences. Ventricular metrics were stratified by segmentation method (smooth, anatomical). Atrial metrics were tested using only the smooth segmentation method, as atrial segmentation does not depend on the papillary/trabecular handling used in the ventricular methods.

Age-specific reference ranges were estimated using GAMLSS [16]. For each cardiac measurement, sex-specific models were fitted using the Box-Cox Cole and Green (BCCG) distribution to account for positive support and distributional asymmetry. The location parameter (μ) was modeled as a smooth function of age using penalized B-splines. Skewness (ν) was fixed to be constant across age in all models, reflecting the assumption that the overall asymmetry of the measurement distribution is a global property of the metric and does not vary markedly with age. The scale parameter (σ), representing variability around the median, was initially allowed to vary smoothly with age. However, when age-varying σ resulted in model instability defined a priori as non-convergence or implausible behavior at extreme centiles (e.g., abrupt local excursions or unrealistically wide upper tails not supported by the empirical data), σ was fixed to a constant to regularize the model and ensure smooth, clinically interpretable reference ranges. Centiles (0.5th, 2.5th, 97.5th, and 99.5th) were derived from the fitted models within the observed age range and summarized within decade-based age groups. For a given centile, metric values were calculated at each integer age within the specified age interval, with the median of these values then used to represent the centile value for that metric within the age interval. The interval between the 2.5th and 97.5th centiles was defined as the ‘reference range’. Values between the 0.5th and 2.5th centiles, and between the 97.5th and 99.5th centiles, were classified as mildly outside the reference range and shown in the table as ‘mild’. Values below the 0.5th percentile or above the 99.5th percentile were considered to represent ‘at least moderate’ deviation from the expected range.

Directionality of deviation was defined according to the clinical interpretation of each metric. For volumetric and mass-based measures (including atrial and ventricular end-diastolic and end-systolic volumes and myocardial mass), higher values were considered to represent deviation from the reference range. For functional measures and stroke volume, including ejection fraction, lower values were considered to represent deviation from the reference range.

In the main manuscript, we present reference ranges and centile-based categories derived from all non-ethnicity-stratified participants, using smooth segmentation and BSA adjustment. This group was selected due to its large sample size; feedback from the imaging community on our previous work using the HHC included remarks that ethnicity is infrequently captured, and that clinical use of smooth segmentation and BSA adjustment is common. Results for all other ethnicities and segmentation types are provided in the **Supplementary material** and on the project website (https://www.healthy-hearts.org.uk) for reference.

Incident outcome associations using percentile-based ranges were examined in a subset of UK Biobank participants. For each endpoint, we first calculated crude event counts, competing deaths, cumulative risk, total follow-up time, and incidence rates per 1,000 person-years across centile-based categories. We then used Cox proportional hazards regression to evaluate the association between CMR severity category and incident outcomes among participants free of the relevant disease at baseline. Hazard ratios (HRs) and 95% CIs were estimated using the Normal category as the reference. Primary models were adjusted for age, sex, and body mass index at imaging. Event-free survival was displayed using Kaplan-Meier curves stratified by percentile-based category. Adjusted associations between CMR severity categories and incident outcomes were summarized using horizontal forest plots of HRs and 95% CIs.

## Results

### Quality control

**Figure S1 and S2** illustrate QC results for cvi42 and suiteHEART, respectively, indicating the number of excluded cases at each stage. The datasets were derived from the UK Biobank healthy subset (n=7,672), the SHIP study healthy subset (n=695), Heidelberg, Germany (n=192), Utrecht, the Netherlands (n=133), Pisa, Italy (n=354), and Singapore (n=180). The largest number of values were removed from LA (cvi42 n=1,502; suiteHEART n=1,466) and RA (n=706; n=2,301) metrics, primarily due to foreshortening. The statistical QC step resulted in the removal of fewer than 1% (cvi42 LV n=96, RV n= 72; suiteHEART LV n =107, RV n=87) of cases.

### Population characteristics and ethnicity trends

After QC the final cohort comprised 9,059 CMR studies, including 4,624 women (51.0%) and 4,435 men (49.0%), with a mean age of 61 ± 13 years (range, 18–83 years). Most participants were of White ethnic background (n=7,396; 81.6%), with smaller representations from South Asian (n=509; 5.6%), Mixed/Other (n=478; 5.3%), Black (n=341; 3.8%), and Chinese (n=335; 3.7%) populations (**Table 1**).

**Table 1.**
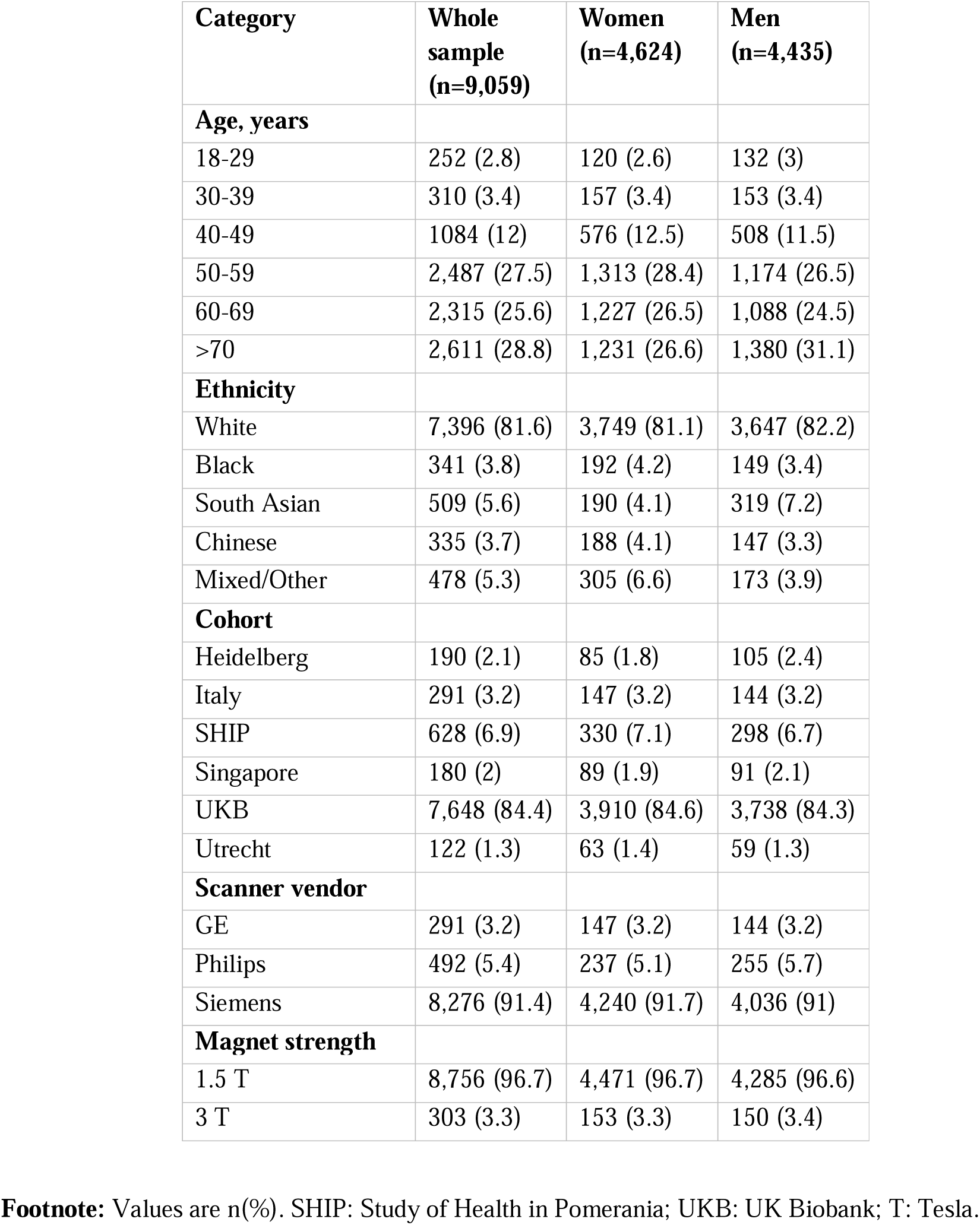
Baseline characteristics.

| **Category** | **Whole sample**<br>**(n=9,059)** | **Women**<br>**(n=4,624)** | **Men**<br>**(n=4,435)** |
| --- | --- | --- | --- |
| **Age, years** |  |  |  |
| 18-29 | 252 (2.8) | 120 (2.6) | 132 (3) |
| 30-39 | 310 (3.4) | 157 (3.4) | 153 (3.4) |
| 40-49 | 1084 (12) | 576 (12.5) | 508 (11.5) |
| 50-59 | 2,487 (27.5) | 1,313 (28.4) | 1,174 (26.5) |
| 60-69 | 2,315 (25.6) | 1,227 (26.5) | 1,088 (24.5) |
| >70 | 2,611 (28.8) | 1,231 (26.6) | 1,380 (31.1) |
| **Ethnicity** |  |  |  |
| White | 7,396 (81.6) | 3,749 (81.1) | 3,647 (82.2) |
| Black | 341 (3.8) | 192 (4.2) | 149 (3.4) |
| South Asian | 509 (5.6) | 190 (4.1) | 319 (7.2) |
| Chinese | 335 (3.7) | 188 (4.1) | 147 (3.3) |
| Mixed/Other | 478 (5.3) | 305 (6.6) | 173 (3.9) |
| **Cohort** |  |  |  |
| Heidelberg | 190 (2.1) | 85 (1.8) | 105 (2.4) |
| Italy | 291 (3.2) | 147 (3.2) | 144 (3.2) |
| SHIP | 628 (6.9) | 330 (7.1) | 298 (6.7) |
| Singapore | 180 (2) | 89 (1.9) | 91 (2.1) |
| UKB | 7,648 (84.4) | 3,910 (84.6) | 3,738 (84.3) |
| Utrecht | 122 (1.3) | 63 (1.4) | 59 (1.3) |
| **Scanner vendor** |  |  |  |
| GE | 291 (3.2) | 147 (3.2) | 144 (3.2) |
| Philips | 492 (5.4) | 237 (5.1) | 255 (5.7) |
| Siemens | 8,276 (91.4) | 4,240 (91.7) | 4,036 (91) |
| **Magnet strength** |  |  |  |
| 1.5 T | 8,756 (96.7) | 4,471 (96.7) | 4,285 (96.6) |
| 3 T | 303 (3.3) | 153 (3.3) | 150 (3.4) |

**Footnote:** Values are n(%). SHIP: Study of Health in Pomerania; UKB: UK Biobank; T: Tesla.

**Table 2:**
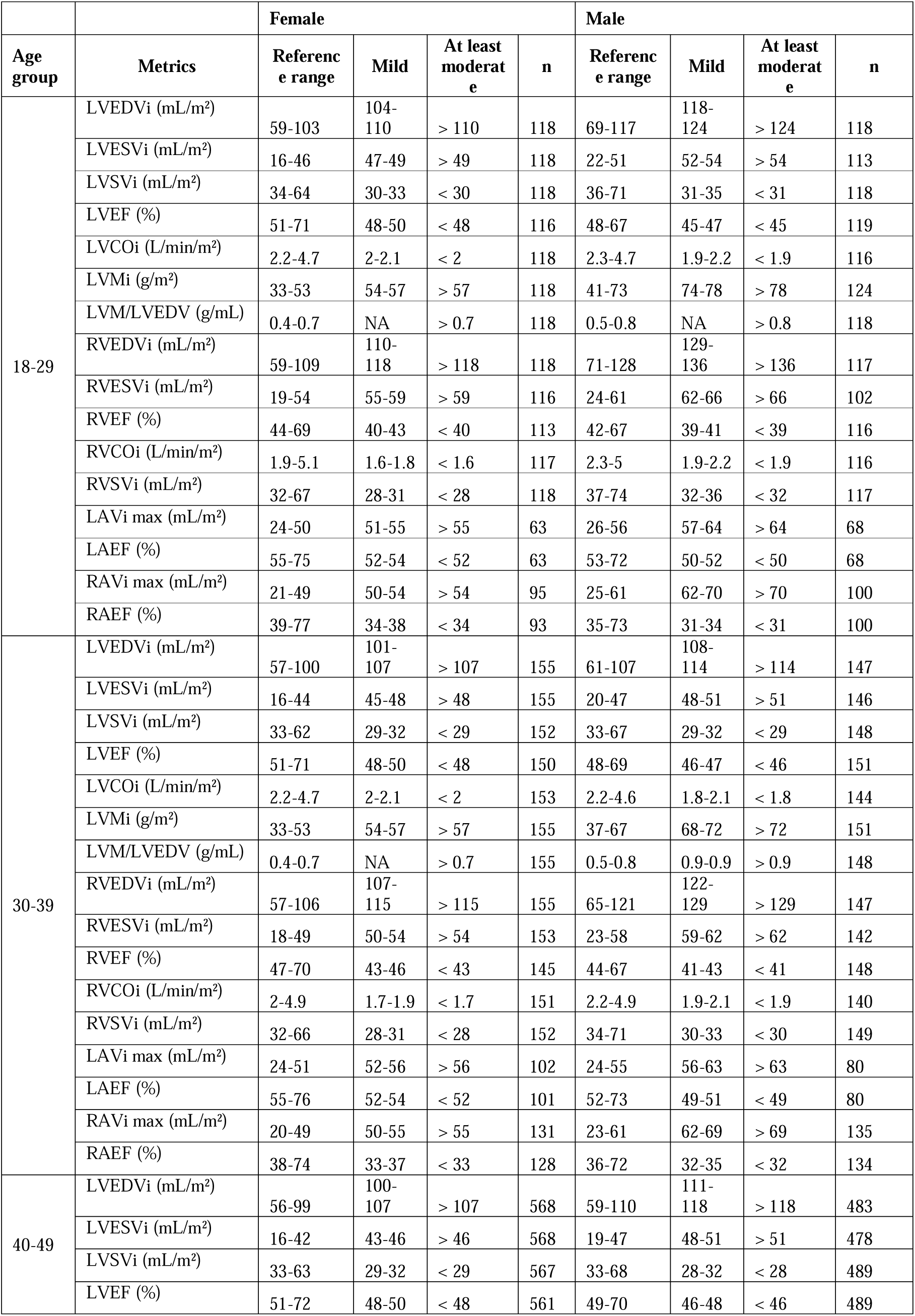
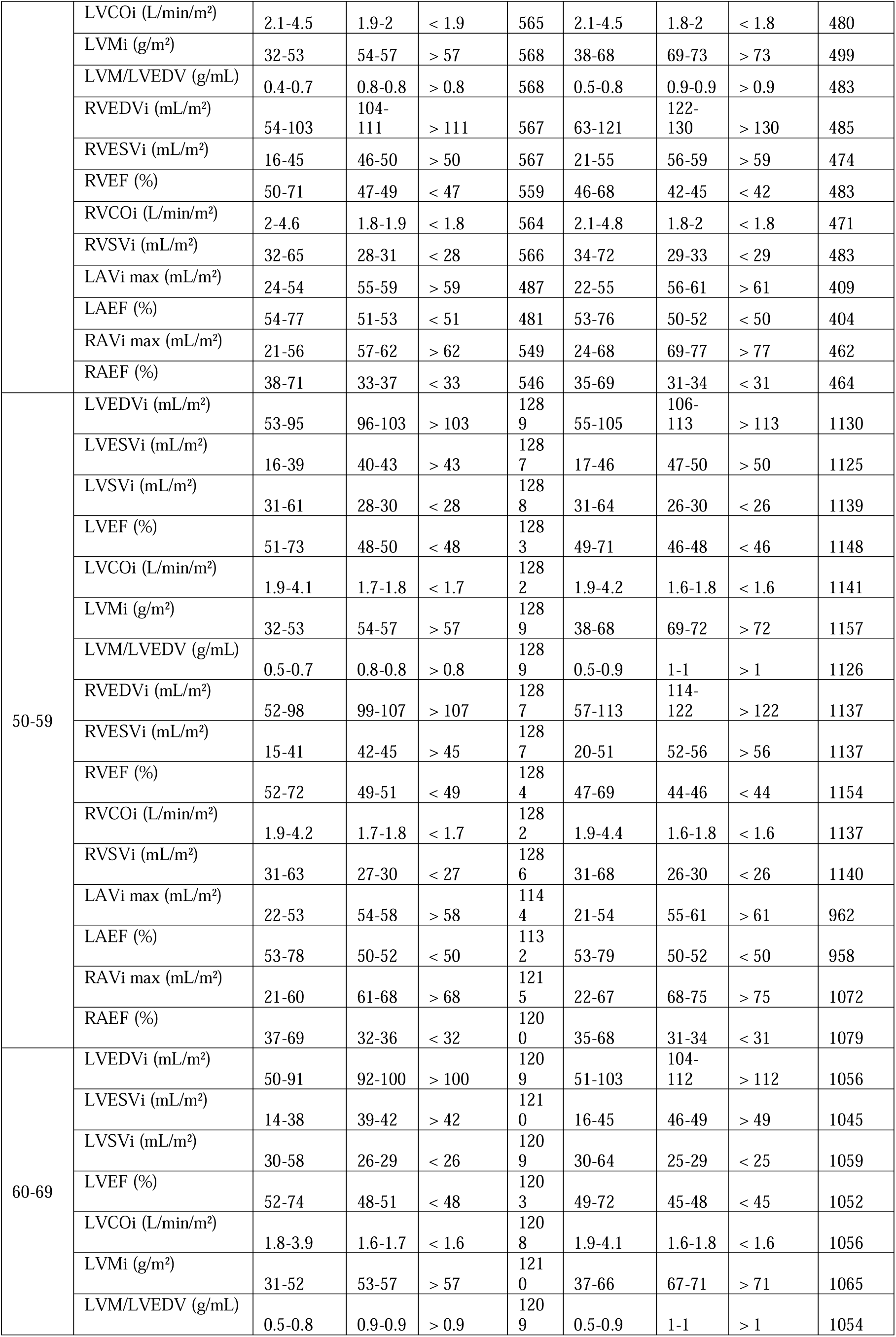
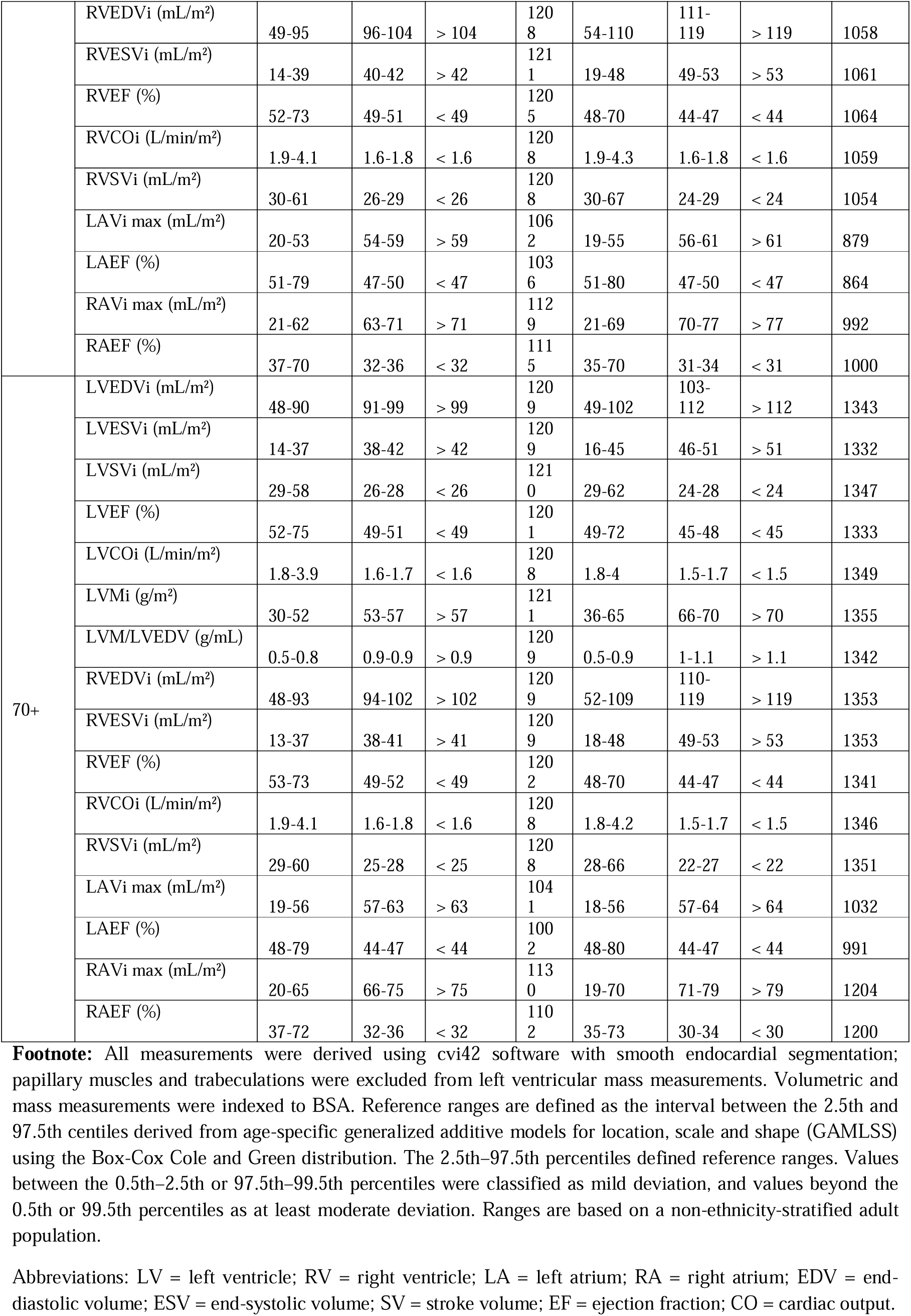
Reference ranges and centile-based categories for cardiac chamber measurements derived using cvi42 software (smooth segmentation), indexed to BSA, in an adult population.

**Table 3:** Reference ranges and centile-based categories for cardiac chamber measurements derived using suiteHEART software (smooth segmentation), indexed to BSA, in an adult population.

| Age group | Metrics | Female |  |  |  | Male |  |  |  |
| --- | --- | --- | --- | --- | --- | --- | --- | --- | --- |
|  |  | Reference range | Mild | At least moderate | n | Reference range | Mild | At least moderate | n |
| 18-29 | LVEDVi (mL/m<sup>2</sup>) | 63-108 | 109-116 | > 116 | 117 | 73-121 | 122-128 | > 128 | 114 |
|  | LVESVi (mL/m<sup>2</sup>) | 23-48 | 49-53 | > 53 | 117 | 27-52 | 53-56 | > 56 | 102 |
|  | LVSVi (mL/m<sup>2</sup>) | 36-65 | 31-35 | < 31 | 117 | 40-73 | 36-39 | < 36 | 119 |
|  | LVEF (%) | 50-69 | 47-49 | < 47 | 117 | 47-67 | 45-46 | < 45 | 123 |
|  | LVMi (g/m<sup>2</sup>) | 32-55 | 56-60 | > 60 | 117 | 43-77 | 78-85 | > 85 | 123 |
|  | LVM/LVEDV (g/mL) | 0.4-0.7 | 0.8-0.8 | > 0.8 | 117 | 0.5-0.8 | 0.9-0.9 | > 0.9 | 114 |
|  | RVEDVi (mL/m<sup>2</sup>) | 61-118 | 119-129 | > 129 | 117 | 76-133 | 134-143 | > 143 | 112 |
|  | RVESVi (mL/m<sup>2</sup>) | 25-54 | 55-61 | > 61 | 116 | 30-59 | 60-64 | > 64 | 109 |
|  | RVEF (%) | 46-68 | 43-45 | < 43 | 115 | 45-66 | 42-44 | < 42 | 121 |
|  | RVSVi (mL/m<sup>2</sup>) | 33-71 | 30-32 | < 30 | 116 | 40-76 | 35-39 | < 35 | 116 |
|  | LAESVi (mL/m<sup>2</sup>) | 18-43 | 44-48 | > 48 | 77 | 19-49 | 50-56 | > 56 | 79 |
|  | LAEF (%) | 56-84 | 51-55 | < 51 | 77 | 54-79 | 51-53 | < 51 | 77 |
|  | RAESVi (mL/m<sup>2</sup>) | 23-48 | 49-53 | > 53 | 88 | 23-59 | 60-66 | > 66 | 102 |
|  | RAEF (%) | 42-75 | 38-41 | < 38 | 83 | 36-71 | 31-35 | < 31 | 99 |
| 30-39 | LVEDVi (mL/m<sup>2</sup>) | 60-106 | 107-114 | > 114 | 149 | 65-111 | 112-119 | > 119 | 144 |
|  | LVESVi (mL/m<sup>2</sup>) | 22-47 | 48-52 | > 52 | 149 | 25-50 | 51-54 | > 54 | 140 |
|  | LVSVi (mL/m<sup>2</sup>) | 35-63 | 30-34 | < 30 | 148 | 36-67 | 32-35 | < 32 | 145 |
|  | LVEF (%) | 50-69 | 47-49 | < 47 | 149 | 48-68 | 45-47 | < 45 | 146 |
|  | LVMi (g/m<sup>2</sup>) | 32-56 | 57-61 | > 61 | 149 | 42-76 | 77-83 | > 83 | 145 |
|  | LVM/LVEDV (g/mL) | 0.4-0.7 | 0.8-0.9 | > 0.9 | 149 | 0.5-0.9 | 1-1 | > 1 | 145 |
|  | RVEDVi (mL/m<sup>2</sup>) | 58-112 | 113-123 | > 123 | 149 | 68-123 | 124-133 | > 133 | 142 |
|  | RVESVi (mL/m<sup>2</sup>) | 23-50 | 51-57 | > 57 | 149 | 27-57 | 58-62 | > 62 | 143 |
|  | RVEF (%) | 48-69 | 45-47 | < 45 | 149 | 45-67 | 42-44 | < 42 | 145 |
|  | RVSVi (mL/m<sup>2</sup>) | 32-70 | 28-31 | < 28 | 149 | 36-72 | 31-35 | < 31 | 142 |
|  | LAESVi (mL/m<sup>2</sup>) | 19-46 | 47-51 | > 51 | 111 | 18-48 | 49-55 | > 55 | 103 |
|  | LAEF (%) | 57-83 | 52-56 | < 52 | 111 | 55-80 | 51-54 | < 51 | 104 |
|  | RAESVi (mL/m<sup>2</sup>) | 22-50 | 51-55 | > 55 | 129 | 23-60 | 61-67 | > 67 | 121 |
|  | RAEF (%) | 42-75 | 37-41 | < 37 | 122 | 36-71 | 32-35 | < 32 | 120 |
| 40-49 | LVEDVi (mL/m<sup>2</sup>) | 58-104 | 105-113 | > 113 | 571 | 63-114 | 115-122 | > 122 | 478 |
|  | LVESVi (mL/m<sup>2</sup>) | 20-45 | 46-51 | > 51 | 571 | 23-50 | 51-54 | > 54 | 478 |
|  | LVSVi (mL/m<sup>2</sup>) | 34-63 | 30-33 | < 30 | 571 | 35-69 | 31-34 | < 31 | 485 |
|  | LVEF (%) | 51-70 | 48-50 | < 48 | 570 | 49-69 | 46-48 | < 46 | 499 |
|  | LVMi (g/m<sup>2</sup>) | 32-57 | 58-62 | > 62 | 571 | 41-74 | 75-82 | > 82 | 497 |
|  | LVM/LVEDV (g/mL) | 0.4-0.8 | 0.9-0.9 | > 0.9 | 571 | 0.5-0.9 | 1-1.1 | > 1.1 | 475 |
|  | RVEDVi (mL/m<sup>2</sup>) | 56-107 | 108-117 | > 117 | 571 | 67-125 | 126-135 | > 135 | 484 |
|  | RVESVi (mL/m<sup>2</sup>) | 20-47 | 48-53 | > 53 | 571 | 25-56 | 57-61 | > 61 | 478 |
|  | RVEF (%) | 49-71 | 46-48 | < 46 | 568 | 46-69 | 42-45 | < 42 | 498 |
|  | RVSVi (mL/m<sup>2</sup>) | 32-67 | 28-31 | < 28 | 569 | 35-73 | 30-34 | < 30 | 478 |
|  | LAESVi (mL/m<sup>2</sup>) | 20-48 | 49-53 | > 53 | 498 | 18-48 | 49-55 | > 55 | 425 |
|  | LAEF (%) | 57-83 | 53-56 | < 53 | 498 | 55-81 | 51-54 | < 51 | 426 |
|  | RAESVi (mL/m<sup>2</sup>) | 22-52 | 53-58 | > 58 | 498 | 23-63 | 64-70 | > 70 | 414 |
|  | RAEF (%) | 41-74 | 36-40 | < 36 | 489 | 37-72 | 32-36 | < 32 | 411 |
| 50-59 | LVEDVi (mL/m<sup>2</sup>) | 55-99 | 100-108 | > 108 | 1290 | 58-109 | 110-118 | > 118 | 1125 |
|  | LVESVi (mL/m<sup>2</sup>) | 18-42 | 43-47 | > 47 | 1289 | 20-47 | 48-52 | > 52 | 1121 |
|  | LVSVi (mL/m<sup>2</sup>) | 33-62 | 29-32 | < 29 | 1289 | 34-68 | 29-33 | < 29 | 1142 |
|  | LVEF (%) | 53-72 | 49-52 | < 49 | 1290 | 50-71 | 47-49 | < 47 | 1155 |
|  | LVMi (g/m<sup>2</sup>) | 32-57 | 58-63 | > 63 | 1290 | 41-73 | 74-80 | > 80 | 1146 |
|  | LVM/LVEDV (g/mL) | 0.4-0.8 | 0.9-1 | > 1 | 1290 | 0.5-1 | 1.1-1.1 | > 1.1 | 1112 |
|  | RVEDVi (mL/m<sup>2</sup>) | 53-102 | 103-112 | > 112 | 1288 | 60-115 | 116-125 | > 125 | 1133 |
|  | RVESVi (mL/m<sup>2</sup>) | 18-43 | 44-49 | > 49 | 1288 | 22-52 | 53-58 | > 58 | 1123 |
|  | RVEF (%) | 50-73 | 46-49 | < 46 | 1286 | 46-70 | 42-45 | < 42 | 1144 |
|  | RVSVi (mL/m<sup>2</sup>) | 31-65 | 27-30 | < 27 | 1283 | 32-70 | 26-31 | < 26 | 1140 |
|  | LAESVi (mL/m<sup>2</sup>) | 19-48 | 49-53 | > 53 | 1171 | 17-48 | 49-55 | > 55 | 986 |
|  | LAEF (%) | 56-82 | 52-55 | < 52 | 1168 | 55-82 | 50-54 | < 50 | 991 |
|  | RAESVi (mL/m<sup>2</sup>) | 21-54 | 55-61 | > 61 | 1099 | 21-60 | 61-68 | > 68 | 884 |
|  | RAEF (%) | 40-74 | 35-39 | < 35 | 1089 | 37-73 | 32-36 | < 32 | 880 |
| 60-69 | LVEDVi (mL/m<sup>2</sup>) | 51-95 | 96-105 | > 105 | 1210 | 54-108 | 109-117 | > 117 | 1051 |
|  | LVESVi (mL/m<sup>2</sup>) | 16-39 | 40-45 | > 45 | 1208 | 18-46 | 47-51 | > 51 | 1045 |
|  | LVSVi (mL/m<sup>2</sup>) | 32-60 | 28-31 | < 28 | 1210 | 32-66 | 27-31 | < 27 | 1054 |
|  | LVEF (%) | 53-73 | 49-52 | < 49 | 1207 | 51-71 | 47-50 | < 47 | 1059 |
|  | LVMi (g/m<sup>2</sup>) | 33-58 | 59-65 | > 65 | 1210 | 40-72 | 73-79 | > 79 | 1061 |
|  | LVM/LVEDV (g/mL) | 0.5-0.9 | 1-1 | > 1 | 1210 | 0.5-1 | 1.1-1.2 | > 1.2 | 1043 |
|  | RVEDVi (mL/m<sup>2</sup>) | 51-99 | 100-109 | > 109 | 1208 | 57-113 | 114-123 | > 123 | 1052 |
|  | RVESVi (mL/m<sup>2</sup>) | 16-41 | 42-47 | > 47 | 1209 | 20-51 | 52-57 | > 57 | 1048 |
|  | RVEF (%) | 50-74 | 46-49 | < 46 | 1207 | 46-71 | 42-45 | < 42 | 1056 |
|  | RVSVi (mL/m<sup>2</sup>) | 30-64 | 26-29 | < 26 | 1206 | 30-70 | 24-29 | < 24 | 1055 |
|  | LAESVi (mL/m<sup>2</sup>) | 18-48 | 49-55 | > 55 | 1077 | 16-49 | 50-56 | > 56 | 889 |
|  | LAEF (%) | 54-82 | 49-53 | < 49 | 1076 | 53-83 | 48-52 | < 48 | 891 |
|  | RAESVi (mL/m<sup>2</sup>) | 21-57 | 58-66 | > 66 | 962 | 21-62 | 63-71 | > 71 | 719 |
|  | RAEF (%) | 40-75 | 35-39 | < 35 | 940 | 37-74 | 32-36 | < 32 | 694 |
| 70+ | LVEDVi (mL/m<sup>2</sup>) | 50-94 | 95-104 | > 104 | 1208 | 51-108 | 109-119 | > 119 | 1334 |
|  | LVESVi (mL/m<sup>2</sup>) | 15-39 | 40-45 | > 45 | 1207 | 18-46 | 47-53 | > 53 | 1319 |
|  | LVSVi (mL/m<sup>2</sup>) | 31-60 | 28-30 | < 28 | 1210 | 30-65 | 25-29 | < 25 | 1346 |
|  | LVEF (%) | 53-74 | 49-52 | < 49 | 1207 | 50-71 | 46-49 | < 46 | 1335 |
|  | LVMi (g/m<sup>2</sup>) | 33-59 | 60-67 | > 67 | 1210 | 39-71 | 72-78 | > 78 | 1342 |
|  | LVM/LVEDV (g/mL) | 0.5-0.9 | 1-1.1 | > 1.1 | 1208 | 0.5-1 | 1.1-1.2 | > 1.2 | 1327 |
|  | RVEDVi (mL/m<sup>2</sup>) | 50-98 | 99-109 | > 109 | 1208 | 55-113 | 114-123 | > 123 | 1335 |
|  | RVESVi (mL/m<sup>2</sup>) | 15-40 | 41-46 | > 46 | 1208 | 20-51 | 52-57 | > 57 | 1333 |
|  | RVEF (%) | 50-75 | 44-49 | < 44 | 1204 | 46-72 | 41-45 | < 41 | 1321 |
|  | RVSVi (mL/m<sup>2</sup>) | 28-64 | 24-27 | < 24 | 1205 | 28-70 | 21-27 | < 21 | 1336 |
|  | LAESVi (mL/m<sup>2</sup>) | 17-52 | 53-61 | > 61 | 1047 | 15-50 | 51-58 | > 58 | 1034 |
|  | LAEF (%) | 51-83 | 45-50 | < 45 | 1037 | 50-84 | 45-49 | < 45 | 1024 |
|  | RAESVi (mL/m<sup>2</sup>) | 20-60 | 61-71 | > 71 | 849 | 21-66 | 67-75 | > 75 | 689 |
|  | RAEF (%) | 41-77 | 36-40 | < 36 | 828 | 37-75 | 32-36 | < 32 | 676 |

**Footnote:** All measurements were derived using suiteHEART software with smooth endocardial segmentation; papillary muscles and trabeculations were excluded from left ventricular mass measurements. Volumetric and mass measurements were indexed to BSA. Reference ranges are defined as the interval between the 2.5th and 97.5th centiles derived from age-specific generalized additive models for location, scale and shape (GAMLSS) using the Box-Cox Cole and Green distribution. The 2.5th–97.5th percentiles defined reference ranges. Values between the 0.5th–2.5th or 97.5th–99.5th percentiles were classified as mild deviation, and values beyond the 0.5th or 99.5th percentiles as at least moderate deviation. Ranges are based on a non-ethnicity-stratified adult population.
Abbreviations: LV = left ventricle; RV = right ventricle; LA = left atrium; RA = right atrium; EDV = end-diastolic volume; ESV = end-systolic volume; SV = stroke volume; EF = ejection fraction; CO = cardiac output.

In linear regression models adjusted for age and sex (**Figure S3, Table S2**), South Asian and Chinese participants had consistently lower LVMi than White participants (standardized β = –0.20 and –0.11, respectively; both p < 0.001), along with smaller indexed biventricular and atrial volumes. By contrast, Black participants showed higher LVMi (β = 0.08) and a higher LVM/LVEDV ratio (β = 0.14; both p < 0.001), consistent with more concentric remodeling. Differences in the Mixed/Other group were generally nonsignificant.

### Percentile-based ranges

Our analyses yielded a comprehensive set of reference ranges and percentile-based categories for left and right ventricular volumes, myocardial mass, and functional parameters, as well as atrial volumes and function. Here, we provide age- and non-ethnicity-stratified grading for ventricular and atrial metrics using both cvi42 and suiteHEART. In **Figures 1** and **2**, the green bands indicate the reference range, while yellow and red zones denote mild and at least moderate deviations from the reference range, respectively. These plots illustrate the age-dependent trajectory of key cardiac parameters within and beyond normative bounds across the studied populations.

**Figure 1:**
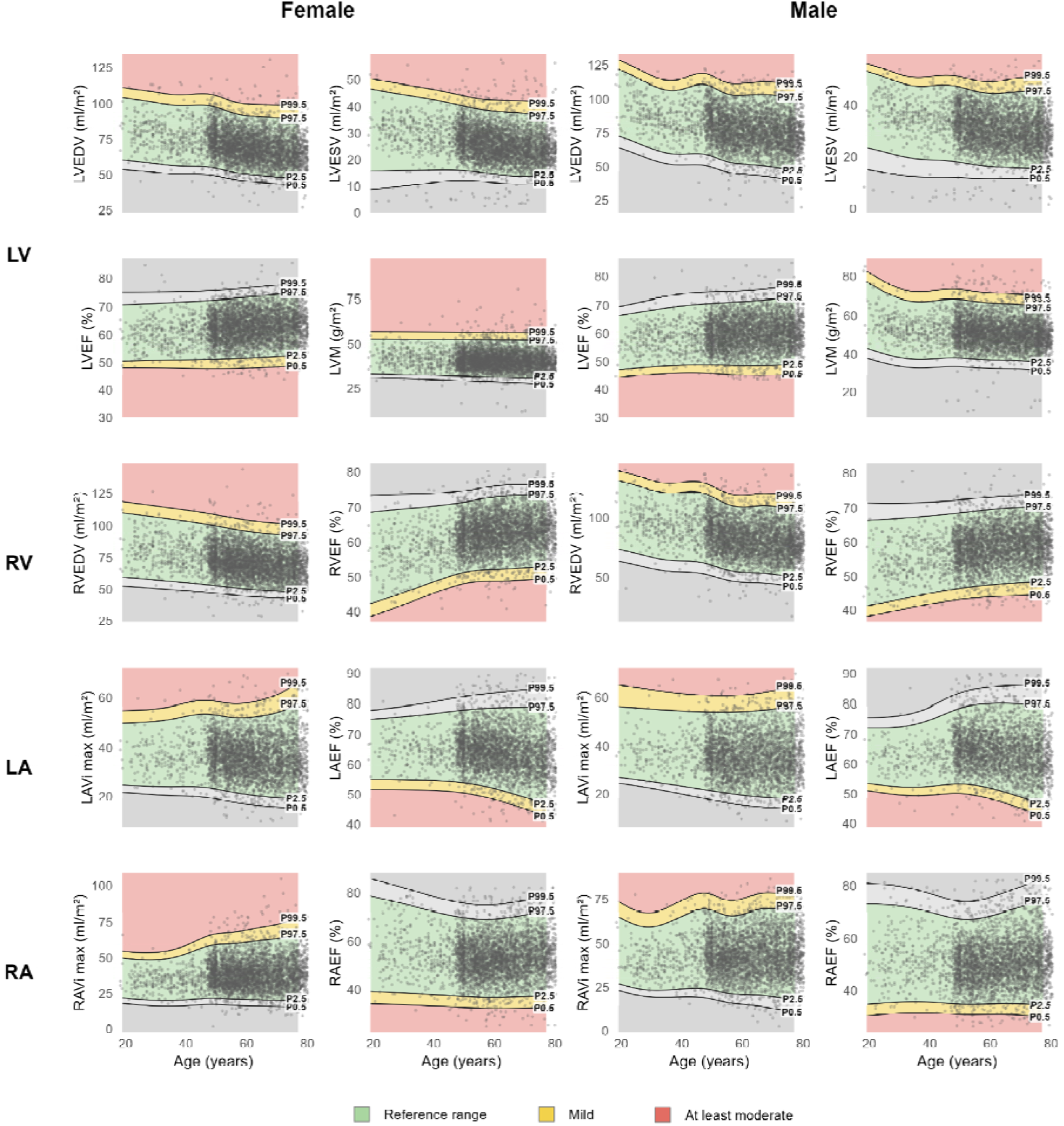
Reference ranges and centile-based categories for key cardiac chamber measurements derived using cvi42 software (smooth segmentation), indexed to BSA, in an adult population. **Footnote:** Figure shows the age- and sex-specific reference distributions for key measures of the left ventricular (LV), right ventricular (RV), left atrial (LA), and right atrial (RA) structure and function. All measurements were derived using cvi42 software with smooth endocardial segmentation; papillary muscles and trabeculations were excluded from left ventricular mass measurements. Volumetric and mass measurements were indexed to BSA. Panels show volumetric indices and ejection fractions by chamber for females and males. Dots represent individual observations; solid black lines denote the 0.5th, 2.5th, 97.5th, and 99.5th percentile reference limits. Reference ranges were defined by the 2.5th–97.5th percentiles (green). Values between the 0.5th–2.5th or 97.5th–99.5th percentiles were classified as mild deviation (yellow), and values beyond the 0.5th or 99.5th percentiles as at least moderate deviation (red).

**Figure 2:**
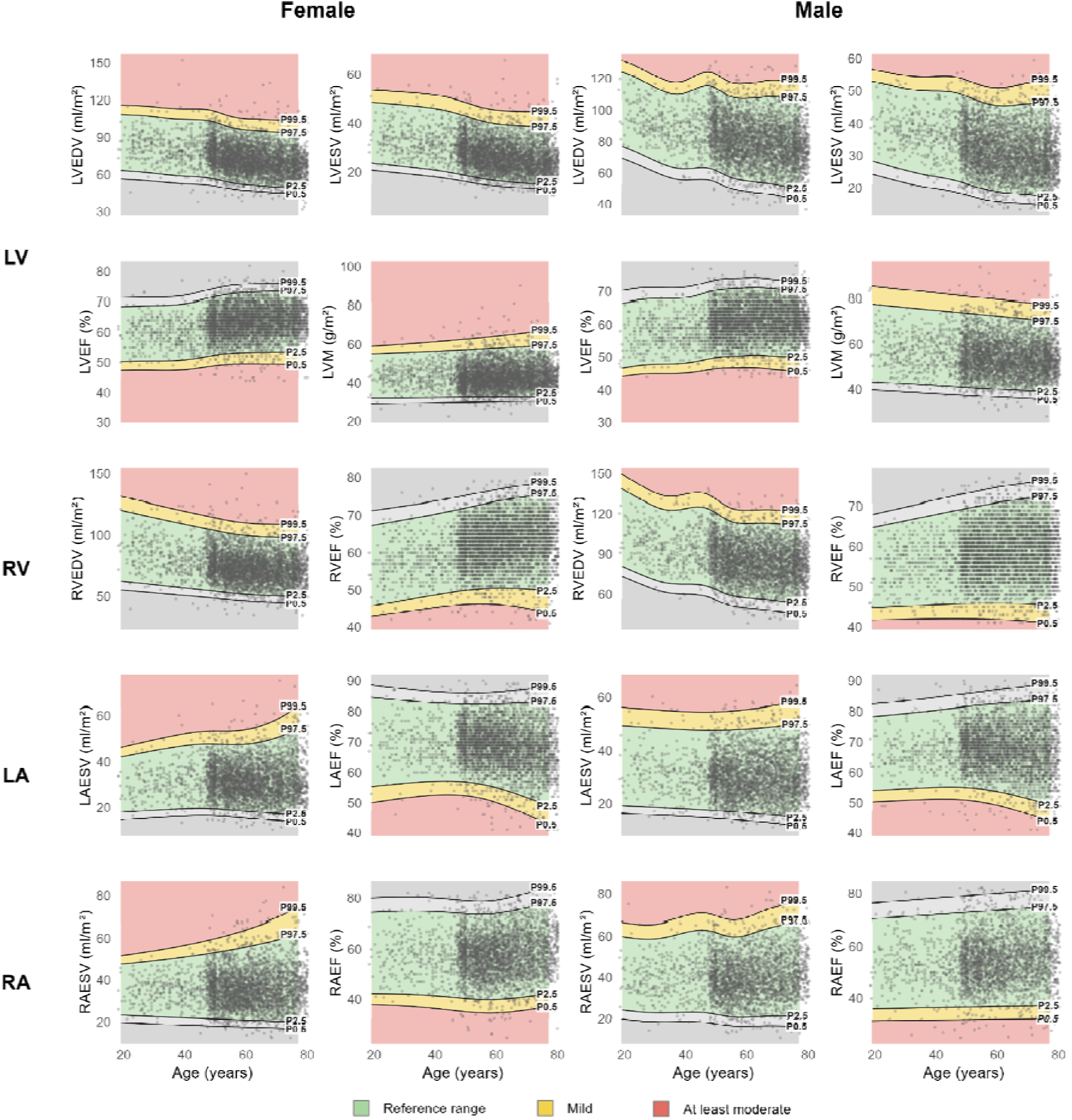
Reference ranges and centile-based categories for key cardiac chamber measurements derived using suiteHEART software (smooth segmentation), indexed to BSA, in an adult population. **Footnote:** Figure shows the age- and sex-specific reference distributions for key measures of the left ventricular (LV), right ventricular (RV), left atrial (LA), and right atrial (RA) structure and function. All measurements were derived using suiteHEART software with smooth endocardial segmentation; papillary muscles and trabeculations were excluded from left ventricular mass measurements. Volumetric and mass measurements were indexed to BSA. Panels show volumetric indices and ejection fractions by chamber for females and males. Dots represent individual observations; solid black lines denote the 0.5th, 2.5th, 97.5th, and 99.5th percentile reference limits. Reference ranges were defined by the 2.5th–97.5th percentiles (green). Values between the 0.5th–2.5th or 97.5th–99.5th percentiles were classified as mild deviation (yellow), and values beyond the 0.5th or 99.5th percentiles as at least moderate deviation (red).

Full reference ranges and severity grading across all subgroup combinations, including segmentation method, software platform, body size indexation approach, and all ethnic groups, are provided in the **Supplementary Tables.**

### Comparison of software solutions

Across both anatomical and smooth segmentation, cvi42 and suiteHEART showed close agreement for the indexed LV ventricular volumes (LVEDVi, LVESVi) and for LVEF, with small biases and limits of agreement (LoA) narrow relative to the measurement range (**Figure S4**). For LVEF, bias was 0.8% (LoA -7.0 to 8.6) under smooth and 1.9% (LoA -5.2 to 8.9) under anatomical segmentation. LVMi showed a consistent positive bias, with suiteHEART higher than cvi42 in both methods and most markedly under anatomical segmentation (+2.5 g/m² smooth, +3.8 g/m² anatomical). RV metrics agreed less well, with considerably wider LoA than the LV metrics: RVEDVi spanned -12.7 to 17.3 mL/m² (smooth) and −18.4 to 13.8 mL/m² (anatomical), and RVEF -10.3 to 10.4% (smooth) and -6.5 to 15.1% (anatomical), the latter also carrying a positive bias (+4.3%). Atrial measurements also showed generally wider LoA than the LV metrics (**Figure S5**); and suiteHEART produced slightly lower atrial volumes (LAESVi, RAESVi) and higher ejection fractions (LAEF, RAEF) than cvi42.

### Prognostic associations of CMR reference thresholds

Age- and sex-specific cvi42-derived reference ranges and percentile-based categories were applied in 59,916 UK Biobank participants with smooth-segmented CMR phenotypes and linked longitudinal outcome data. Prevalent cardiovascular disease at the time of imaging was generally uncommon, except arterial hypertension, which was present in 9,652 participants (16.1%); overall, 3,380 participants (5.6%) had prevalent major cardiovascular disease at baseline, including heart failure, ischemic heart disease, and atrial fibrillation. Over a median follow-up of 4 years, 2,192/56,536 participants (3.9%) free of major cardiovascular disease at imaging developed incident major cardiovascular disease, and 106/59,916 (0.2%) died from cardiovascular causes (**Table S3**).

Participants were classified into the within-reference-range, mild, and at least moderate categories for all ventricular metrics. Departure from the reference range was associated with progressively higher risk of incident cardiovascular outcomes (**Figure 3, Table S4**). Associations were strongest and most consistent for heart failure, with at least moderate category conferring a 6 to 9-fold higher risk for the left ventricular metrics (LVEF, LVESVi, LVEDVi, and LVMi). An at least moderate decrease in RVEF was likewise associated with a higher risk of heart failure (HR 3.95, 95% CI 3.14-4.97). Associations with any major cardiovascular disease followed the same direction but were attenuated. For LVEF, Kaplan–Meier curves showed concordant stepwise separation across severity categories, particularly for heart failure and any major cardiovascular disease (**Figure 4**). Associations with cardiovascular mortality were generally weaker, although lower (at least moderate) LVEF remained significantly associated with cardiovascular mortality (HR 2.21, 95% CI 1.44–3.39).

**Figure 3:**
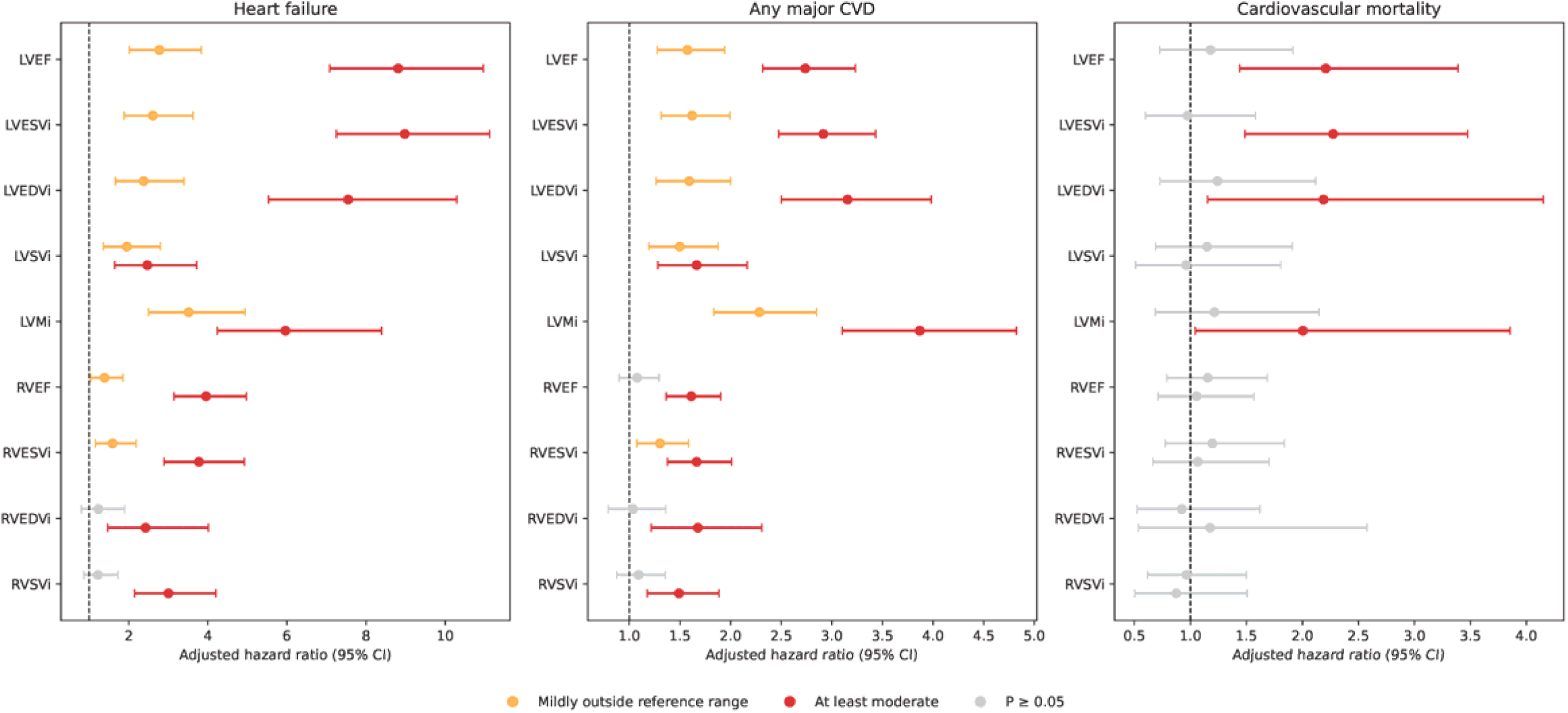
Associations of CMR grading categories with key incident outcomes. **Footnote:** Points represent adjusted hazard ratios and horizontal lines 95% confidence intervals from Cox proportional hazards models, with the within-reference-range category used as the reference. Orange markers indicate mildly outside-the-reference-range values and red markers at least moderately outside-the-reference-range values; associations not reaching statistical significance (P ≥ 0.05) are shown in grey. Models were adjusted for age, sex, and body mass index at imaging. Abbreviations: CI, confidence interval; CVD, cardiovascular disease; LVEDVi, left ventricular end-diastolic volume indexed to body surface area; LVESVi, left ventricular end-systolic volume indexed to body surface area; LVSVi, left ventricular stroke volume indexed to body surface area; LVMi, left ventricular mass indexed to body surface area; LVEF, left ventricular ejection fraction; RVEDVi, right ventricular end-diastolic volume indexed to body surface area; RVESVi, right ventricular end-systolic volume indexed to body surface area; RVSVi, right ventricular stroke volume indexed to body surface area; RVEF, right ventricular ejection fraction.

**Figure 4.**
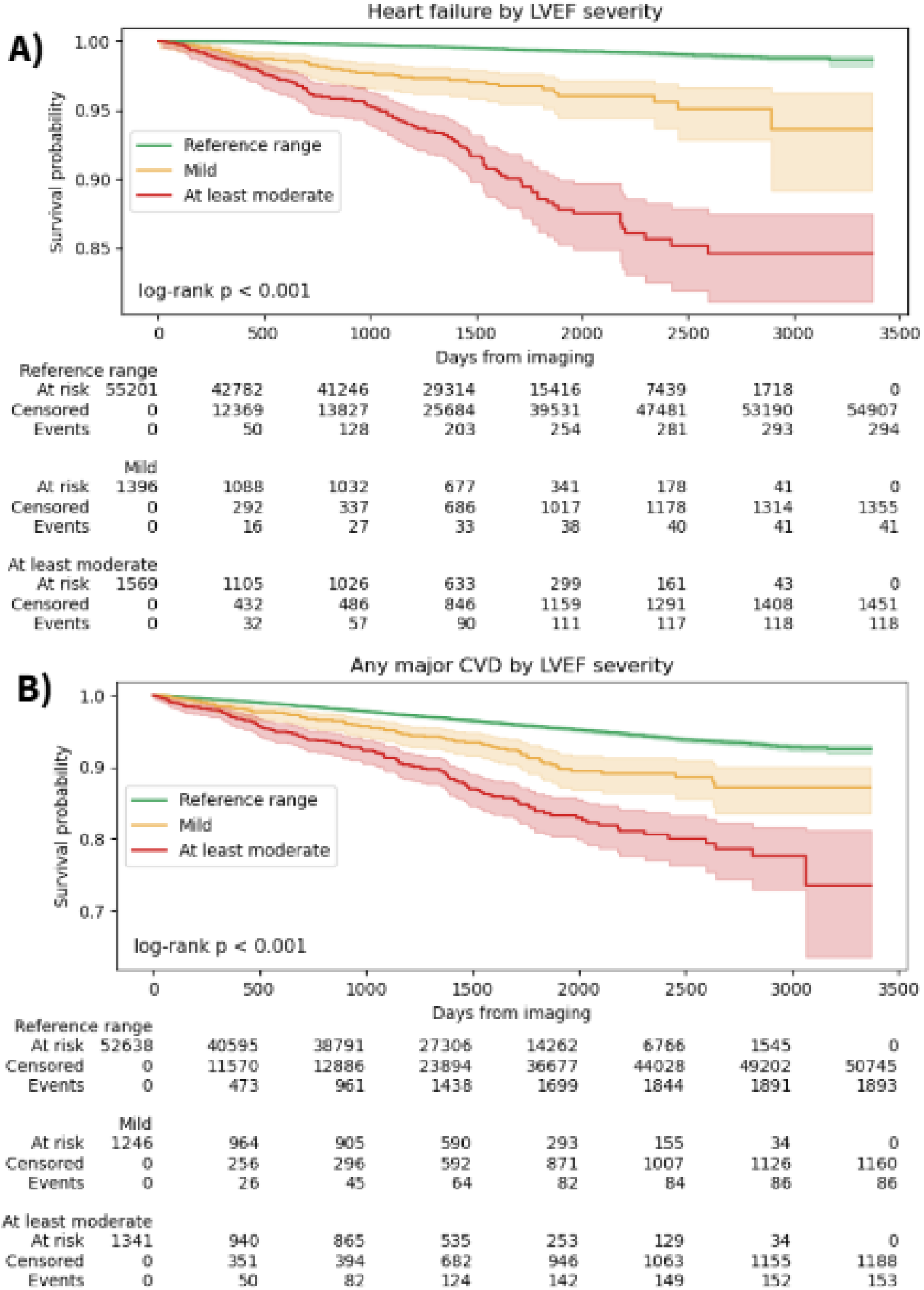
Kaplan–Meier curves for incident heart failure and any major cardiovascular disease according to LVEF category. **Footnote:** Kaplan–Meier curves showing event-free survival for incident heart failure (top) and incident major cardiovascular disease (bottom) stratified by LVEF category. Participants were classified as within reference range, mildly outside the reference range, or at least moderate category according to age- and sex-specific thresholds. Numbers at risk, censored participants, and incident events are shown below each plot. Abbreviation: LVEF, left ventricular ejection fraction.

## Discussion

This study presents population-stratified reference ranges and centile-based grading thresholds for CMR-derived ventricular and atrial parameters from a large multi-ethnic cohort of healthy adults. We tested prognostic relevance of the calculated thresholds in almost 60,000 UK Biobank Imaging Study participants, demonstrating significant associations with incident cardiovascular outcomes, most notably for heart failure. These findings support the clinical relevance of the proposed grading framework and suggest that CMR reference ranges can be translated into prognostically informative categories rather than serving only as descriptive ‘normal’ limits. By implementing the framework across two widely used post-processing platforms, cvi42 and suiteHEART, we also aimed to maximize reproducibility and facilitate uptake across both research and clinical environments.

A key strength of this study the integration of AI-based segmentation with rigorous multi-stage QC across heterogeneous cohorts. Automated segmentation is increasingly embedded in clinical CMR workflows and offers gains in efficiency and consistency[17]. Yet, human oversight remains essential to confirm contour accuracy and detect errors not apparent from automated output alone [18]. Although statistical QC alone may suffice in highly standardized large-scale epidemiological studies [19], the heterogeneity of acquisition across our cohorts and the clinical purpose of defining reference ranges necessitated additional expert review. Emerging software-driven approaches may ease this burden, either by partially automating the assessment of automated segmentations [20] or by providing easy access to structured scoring, as with the in-house Shiny-app–based quality-scoring method used in our study.

Prior studies have compared CMR measurements across post-processing platforms, but largely in small, manually or semi-automatically analysed cohorts [21, 22]. Here we assess agreement between two widely used software solutions, cvi42 and suiteHEART, applied with fully automated segmentation at scale, allowing interchangeability to be characterised across a far larger population. Agreement was close for LV volumes and ejection fraction, with small biases and narrow LoA relative to the measurement range, indicating that these metrics are broadly comparable across platforms. The two solutions were not, however, fully interchangeable: suiteHEART returned systematically higher LV mass than cvi42 (bias up to +3.8 g/m² under anatomical segmentation), and agreement was weaker for RV and atrial metrics, with substantially wider LoA. These patterns are consistent with prior inter-vendor comparisons, in which LV volumes and function agree reasonably well. At the same time, mass and right-heart measurements differ in clinically relevant, software-dependent ways [22, 23]. The practical implication is that reference ranges and centile-based categories should ideally be applied using the same platform from which they were derived; providing platform-specific ranges, as we have in this article, avoids the misclassification that a single cross-vendor cut-off would introduce. For the core LV metrics, however, volumes and ejection fraction agreed closely across platforms, indicating they are sufficiently concordant to apply pooled reference ranges across software with acceptable error.

Cardiac structure varied systematically with ethnicity, consistent with and extending prior CMR evidence [24]. On average, Black participants had the highest LVMi and the highest LVM/LVEDV ratio accounting for sex and body size, indicating more concentric left ventricular geometry. South Asian and Chinese participants had lower LVMi and smaller indexed biventricular and atrial volumes than White participants. The smaller indexed chambers in our South Asian participants are consistent with a recent UK Biobank CMR comparison of healthy South Asian and White European adults [25], and the smaller atrial volumes in Chinese participants mirror the lower indexed left atrial volumes reported for Chinese-American participants in the Multi-Ethnic Study of Atherosclerosis (MESA) [26]. Differences in the Mixed/Other group were generally not significant, likely reflecting heterogeneity and limited sample size. Ethnic variation in CMR-derived anatomy has been documented since the earliest multi-ethnic CMR reference work [27], yet it is rarely incorporated into the reference ranges used in practice.

Alongside ethnicity, the centiles captured the expected gradients with age and sex: men had higher indexed ventricular volumes and mass than women, and indexed volumes and mass were smaller in older age, while ejection fraction was relatively preserved, consistent with our previous results and the literature [1, 8, 13, 27]. Because age, sex, and ethnicity each shift the center and spread of the normal distribution, applying ranges that ignore them risks systematic misclassification [4]; for example, overcalling left ventricular hypertrophy in groups with constitutionally higher mass, supporting the jointly stratified ranges we provide.

The 2019 EACVI expert consensus was the first to propose a consensus framework for grading the severity of CMR-derived chamber size, function and aortic measurements, adapting an approach previously established in echocardiography [4]. While this was a crucial step toward standardization, it was based primarily on expert consensus, and its reference datasets were small and lacked ethnic diversity. By deriving statistically defined grading thresholds from a large, multi-ethnic cohort, our study addresses these limitations and provides an evidence-based framework for classifying cardiac abnormalities.

Our prognostic findings extend recent work using the conventional 2-SD method. Kawel-Boehm et al. applied mean ± 2 SD limits in the MESA cohort and found that several LV parameters beyond the 2-SD threshold predicted later cardiovascular events [28]. Our analysis used centile-based modeling instead and characterizes the full distribution of the data, allowing its centre, spread, and shape to vary with covariates such as age, sex, and ethnicity, an approach increasingly used for imaging reference curves and recently applied to pediatric CMR [29, 30]. There was a notable difference for the right heart: in the MESA study, several right ventricular parameters beyond the 2-SD limit were not predictive of events [28], whereas in our cohort, centile-based RVEF categories were associated with incident heart failure. Several factors could contribute to this. Our cohort was roughly ten times larger than the MESA cohort, giving greater power to detect the more modest prognostic signal typically carried by RV phenotypes for predominantly left-sided outcomes such as heart failure. RV measurements are also generally more variable than LV measurements, which reduces the precision of their disease associations. In addition, characterising the full distribution rather than applying symmetric 2-SD limits may better capture deviation at the tails, particularly where the underlying distribution is skewed. However, a head-to-head comparison in the same cohort would be needed to confirm this. More broadly, the stepwise increase of outcome hazard ratios across centile categories, strongest for heart failure, supports translating reference ranges into prognostically informative grades rather than descriptive limits alone.

Finally, this study contributes to the ongoing effort to enhance the quality and standardization of CMR services by establishing robust, population-specific reference ranges and grading [1, 4, 31]. These ranges are a benchmark rather than a rule: they should be adapted to local practice and interpreted alongside individual patient context. Accurate, consistent CMR interpretation depends on high-quality acquisition, robust post-processing and structured reporting [14, 32], and formal competency assessment for reporting clinicians remains important for the correct application of reference values in decision-making.

## Limitations

Several limitations should be acknowledged. Most importantly, the reference population had limited demographic breadth: young adults were underrepresented, and despite the ethnic diversity of the HHC resource, over 80% of CMR studies were performed in White individuals, with some ethnic groups absent altogether. Reference ranges and severity thresholds may therefore be less accurate at the younger end of the age range and unreliable for underrepresented groups, and they will require validation in more diverse and younger cohorts. The population excluded competitive and highly active endurance athletes, in whom exercise-induced remodeling alters chamber size and function, and our reference values may not apply to individuals who engage in regular, intensive physical activity. Pregnant women are also not represented in our cohort.

## Conclusions

In a large, multi-ethnic population of healthy adults, we derived reference ranges and centile-based grading thresholds for conventional ventricular and atrial CMR measures using two widely used software platforms. In a large validation cohort, stepwise increases in cardiovascular risk were observed in more extreme centile categories, with individuals in the at least moderate category having a 6- to 9-fold higher risk of incident heart failure. By pairing multi-ethnic reference values with prognostic validation, this work moves CMR interpretation toward graded categories that carry explicit clinical meaning rather than a single cut-off value.

## Additional information

### Ethics statement

This study was conducted in accordance with the principles of the Declaration of Helsinki. Analysis of the UK Biobank dataset was performed under approval from the NHS National Research Ethics Service (Ref 11/NW/0382; extension Ref 21/NW/0157), with written informed consent obtained from all participants. For all other contributing cohorts, local ethical approval and participant consent procedures were confirmed by the respective institutions.

## Supporting information

Non ethnicity statified reference ranges

Ethnicity stratified reference ranges

## Data Availability

All data produced in the present work are contained in the manuscript.

