## Supplementary material for "Cardiovascular magnetic resonance reference ranges and centile-based categories for atrial and ventricular cardiac structure and function: 2026 Update from the Healthy Hearts Consortium"

**LIST OF ABBREVIATIONS**

BSA – body surface area

i – indexed (to height or BSA as indicated in the table title)

LA – left atria

LA max – maximum left atrial volume

LAEF – left atrial ejection fraction

LAESV – left atrial end-systolic volume

LV – left ventricle

LVCO – left ventricular cardiac output

LVEDV – left ventricular end-diastolic volume

LVEF – left ventricular ejection fraction

LVESV – left ventricular end-systolic volume

LVM diast – left ventricular mass in diastole

LVM syst – left ventricular mass in systole

LVSV – left ventricular stroke volume

RA – right atria

RA max – maximum right atrial volume

RAEF – right atrial ejection fraction

RAESV – right atrial end-systolic volume

RV – right ventricle

RVCO – right ventricular cardiac output

RVEDV – right ventricular end-diastolic volume

RVEF – right ventricular ejection fraction

RVESV – right ventricular end-systolic volume

RVM diast – right ventricular mass in diastole

RVSV – right ventricular stroke volume

**Figure S1: Flowchart for the quality control for the values derived by cvi42 (Circle Cardiovascular Imaging Inc)**

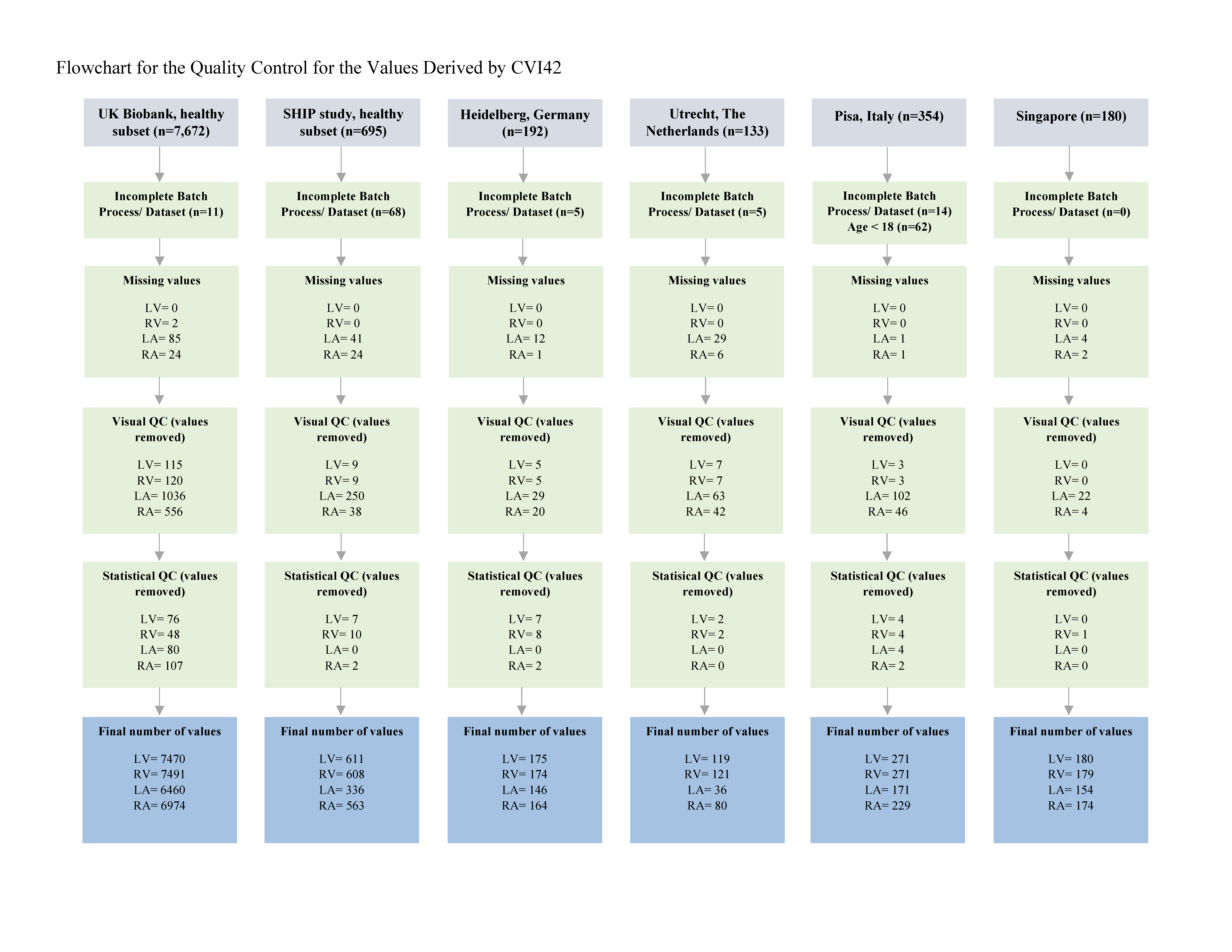

**Footnote:** Grey boxes indicate the number of DICOM files received from collaborating sites. Green boxes summarize the quality control steps applied during data cleaning. Final, individual chamber‐level values appear in the blue boxes. In the baseline characteristics table **(Table 1**), we report baseline characteristics on a per‐individual basis.

**Figure S2: Flowchart for the quality control for the values derived by suiteHEART (NeoSoft)**
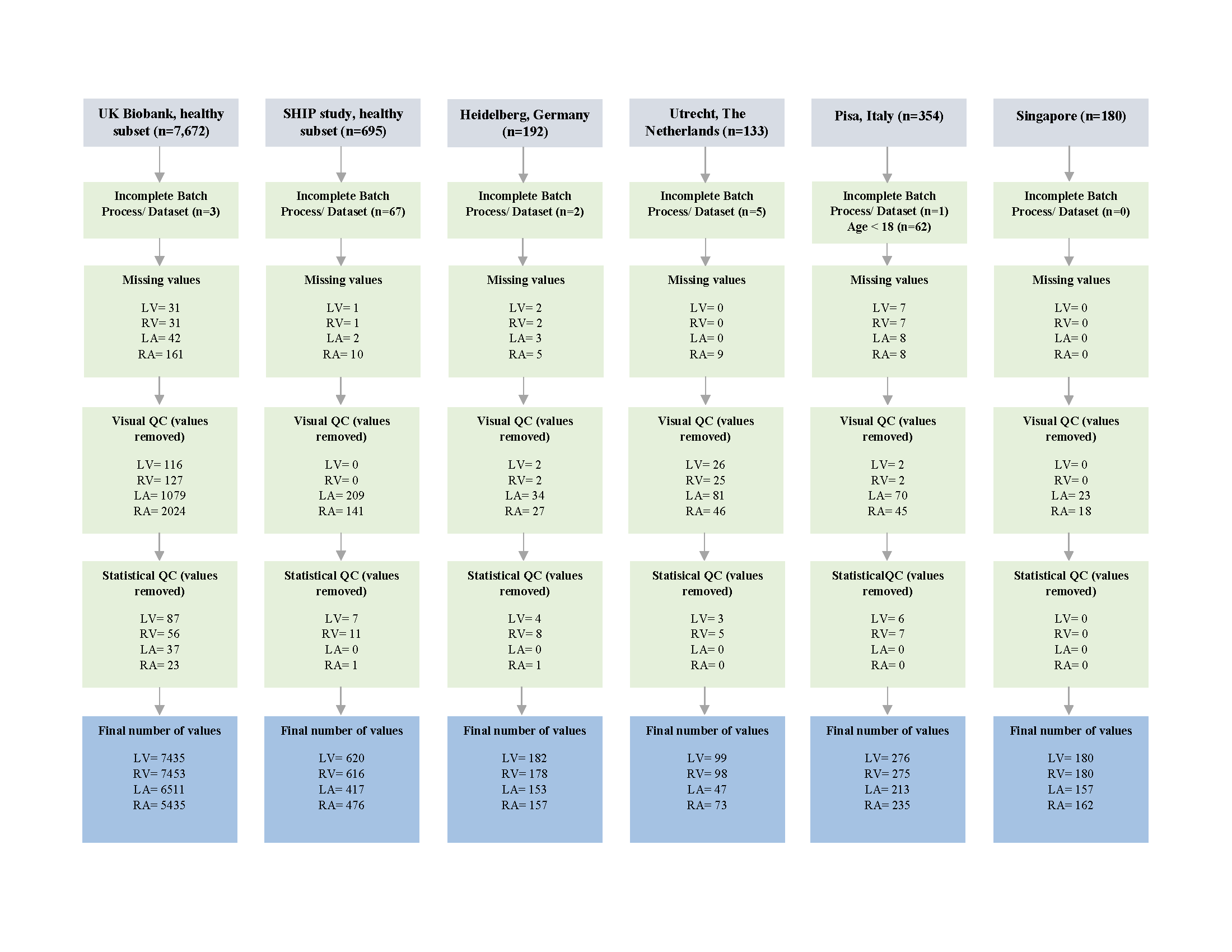

**Footnote:** Grey boxes indicate the number of DICOM files received from collaborating sites. Green boxes summarize the quality control steps applied during data cleaning. Final, individual chamber‐level values appear in the blue boxes. In the baseline characteristics table **(Table 1**), we report baseline characteristics on a per‐individual basis.

**Figure S3: Association between ethnicity and CMR metrics**

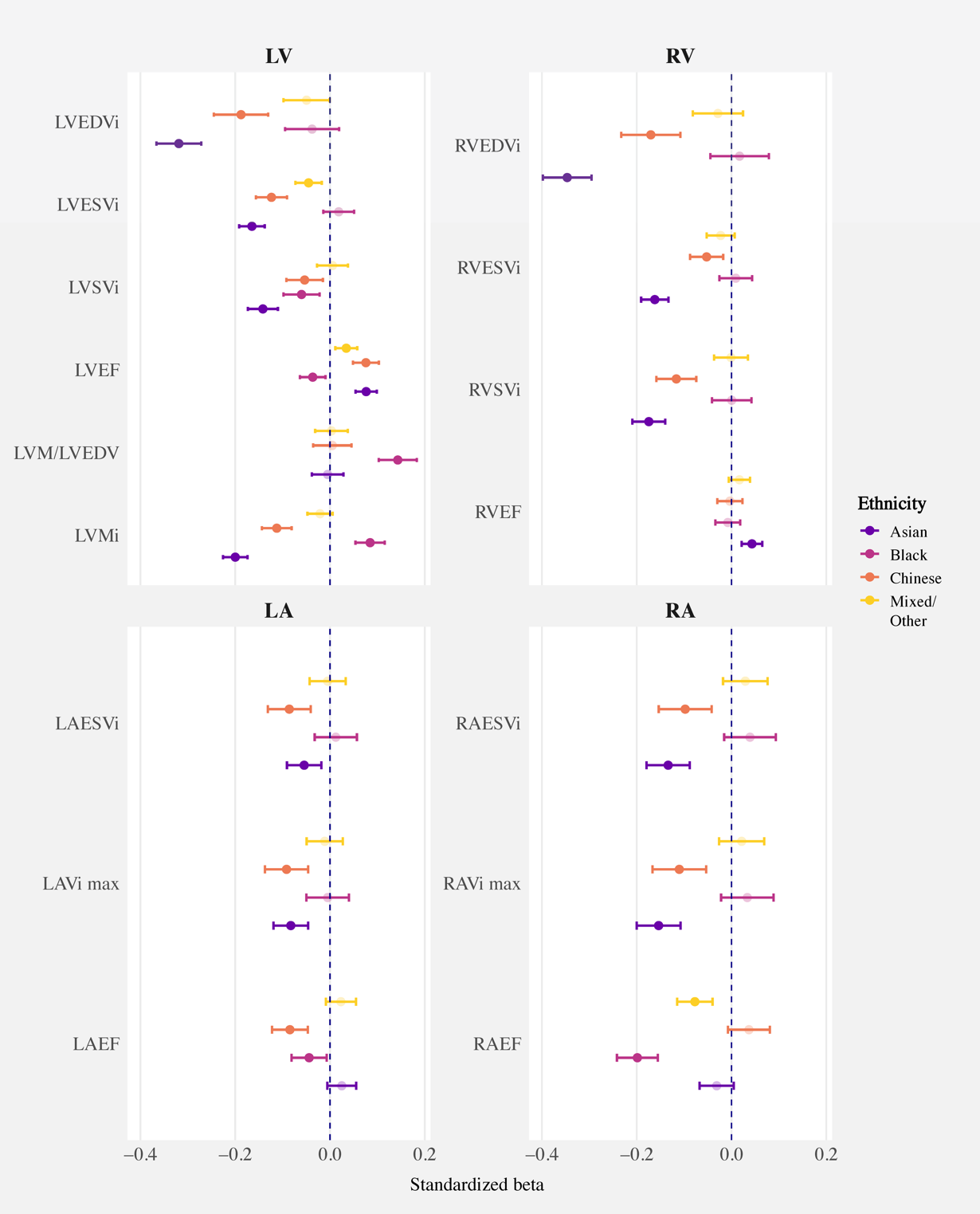

**Footnote:** Standardized β coefficients (with 95% confidence intervals) from linear regression models are shown for each ethnic group, using White participants as the reference using the smooth contour approach, indexed to BSA. Each model was adjusted for age and sex. P-values were corrected for multiple comparisons using the false discovery rate method.

**Figure S4: Comparison between suiteHEART and cvi42 batch processing algorithm output for ventricular measures**

**Footnote:** Bland-Altman plots showing the difference (suiteHEART − cvi42) were plotted against the mean of the two measurements. Ventricular metrics were stratified by segmentation method (smooth shown above, anatomical shown below). The bias was calculated as the mean of the paired differences, and the 95% limits of agreement as the bias ± 1.96·SD of the differences, assuming approximately normally distributed differences.

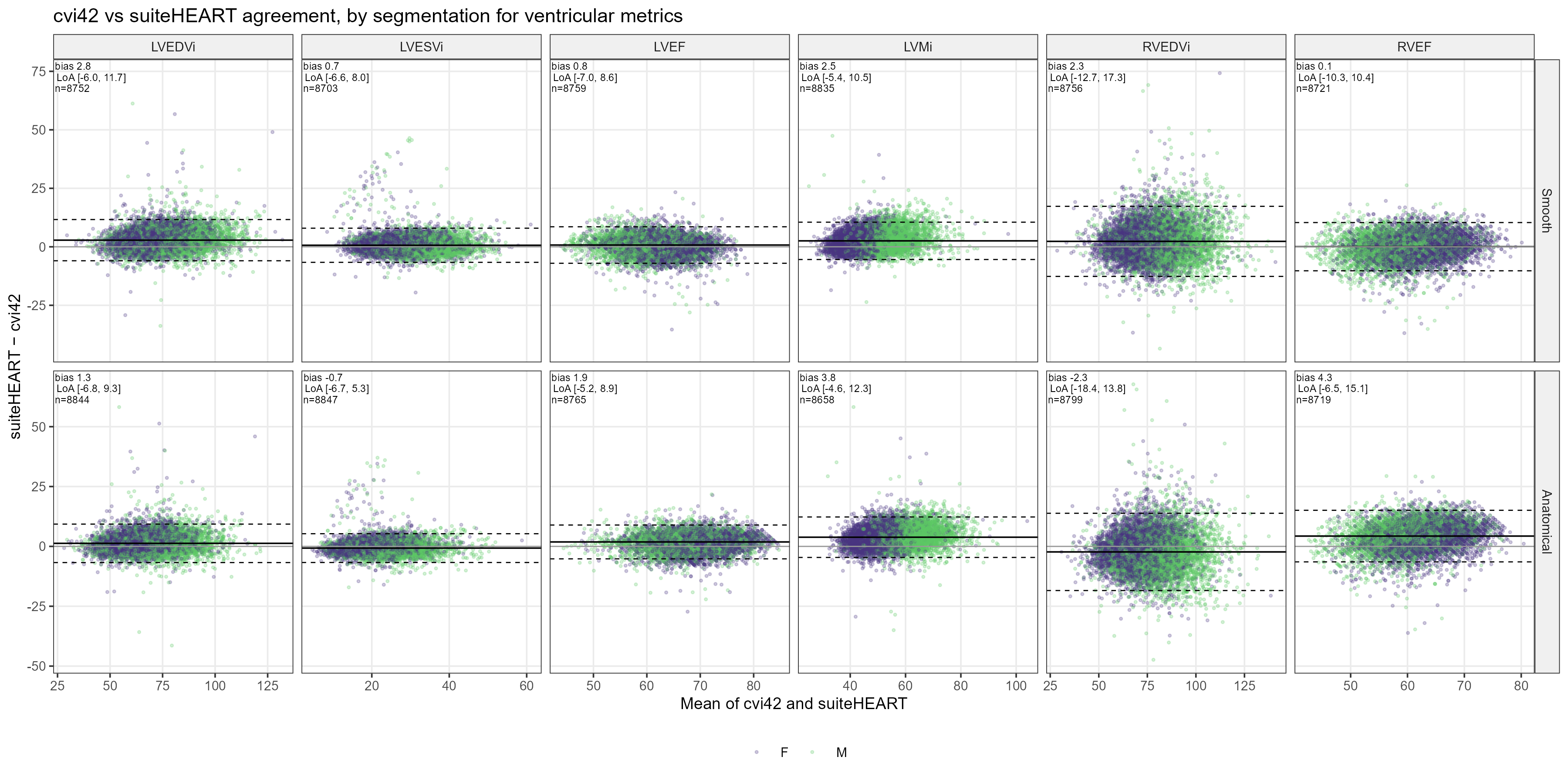

**Figure S5: Comparison between suiteHEART and cvi42 batch processing algorithm outputs for atrial measures**

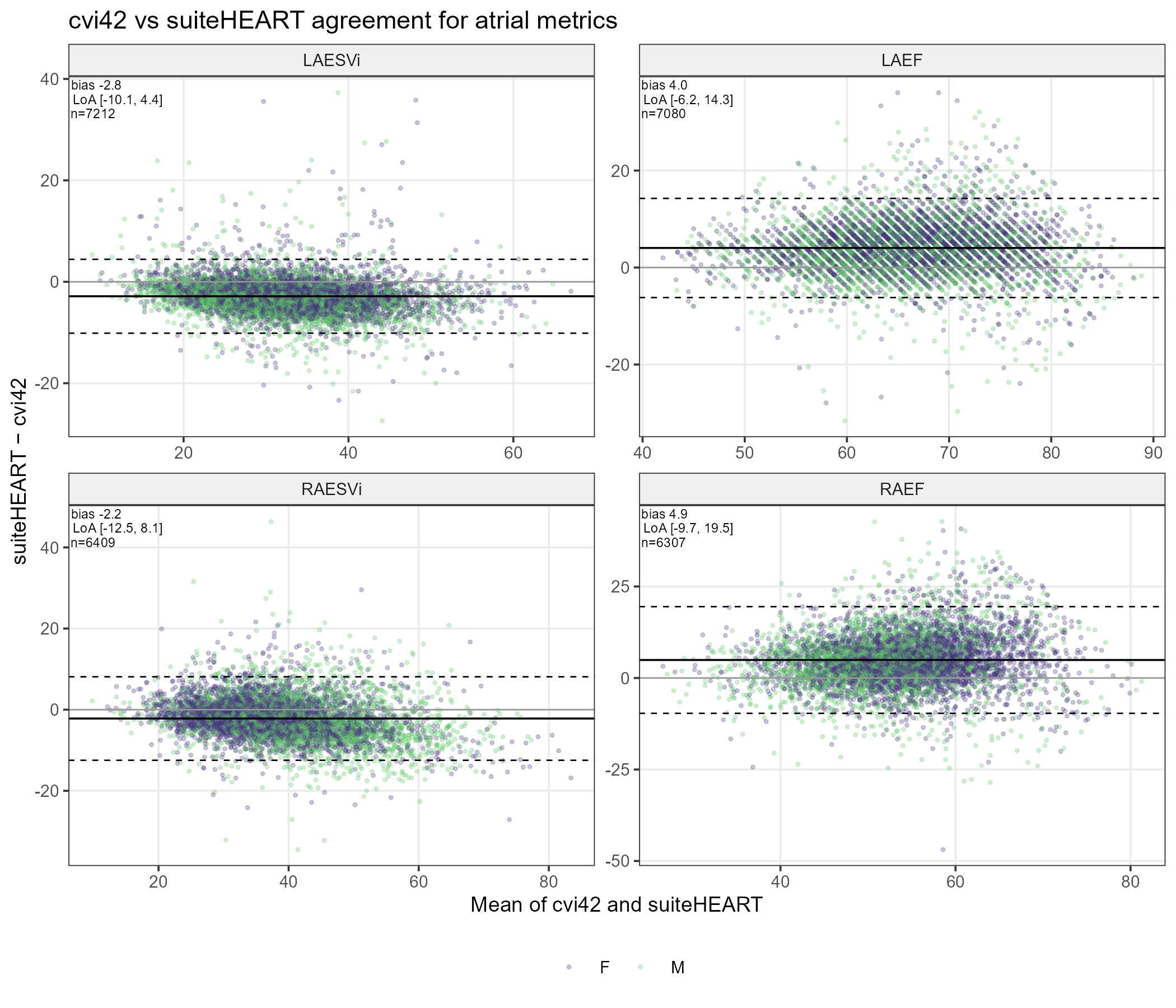

**Footnote:** Blant-Altman plots showing the difference (suiteHEART − cvi42) was plotted against the mean of the two measurements. The bias was calculated as the mean of the paired differences, and the 95% limits of agreement as the bias ± 1.96·SD of the differences, assuming approximately normally distributed differences.

**Table S1. Summary of participant selection criteria for each Healthy Hearts Consortium cohort**

| **Contributing cohort** | **Selection criteria** |
| --- | --- |
| Utrecht (The Netherlands) | - Aged between 25 and 35 years - No prior history of cardiovascular disease - No cardiac arrhythmias - No record of cardiovascular preventive medication |
| Pisa (Italy) | - Age between 15 and 80 years - No personal history of cardiac or non-cardiac disease that may affect cardiac function. - No family history of genetic disease - Normal physical examination - Normal electrocardiogram - Normal echocardiographic examination - No coronary risk factors including diabetes, hypertension (defined as diastolic blood pressure>90 mmHg, systolic blood pressure>140 mm Hg, or receiving treatment for hypertension), hyperlipidemia (abnormal lipid levels or receiving lipid-lowering therapy), obesity (defined as body mass index≥30kg/m^2^). - No history of smoking or drug use |
| Heidelberg (Germany) | - Aged over 20 years old - No signs, symptoms or history of any cardiac disease, including arterial hypertension, cardiovascular, cerebrovascular or non-cardiac diseases, any regular medication except for contraceptives or vitamins - Screening was performed with clinical history, physical examination, 12-lead ECG, and a resting blood pressure - Fasting glucose and HBA1c were measured, and those with impaired glucose tolerance were excluded - In all participants, CMR stress test (first-pass perfusion with adenosine-induced vasodilation or stress function CMR during dobutamine) was performed to exclude significant coronary artery disease |
| Singapore | - Aged 20 to 69 years old - No symptoms, clinical or family history of cardiovascular or cerebrovascular disease - No history of hypertension, hyperlipidemia, or diabetes mellitus. - Individuals with valvular heart disease or wall motion abnormalities detected on CMR were excluded |
| Study of Health in Pomerania (SHIP) | - Aged 20 to 80 years - No prior arterial hypertension (measured blood pressure), diabetes mellitus (measured HbA1c), myocardial infarction (self-report, 12-lead ECG), self-reported cardiac surgery, heart failure, stroke, chronic lung disease, peripheral artery disease. - No cardiovascular/pulmonary medication - subjects with ECG signs of atrial fibrillation or left or right bundle branch block and participants with late gadolinium enhancement in CMR imaging were excluded from the reference population |
| UK Biobank | - Age 44 to 83 years old - Exclusion of any record of cardiovascular disease according to self-report in UK Biobank assessments, or linked electronic health records – including hospital episode statistics and Office of Population Censuses and Surveys Classification of Interventions and Procedures - Matched deprivation and risk factor profile to the UK general population - For further details see [**https://doi.org/10.1016/j.jcmg.2024.01.009**](https://doi.org/10.1016/j.jcmg.2024.01.009) **or** [**https://healthy-hearts.org.uk/**](https://healthy-hearts.org.uk/) |

CMR: cardiovascular magnetic resonance; ECG: electrocardiogram; HbA1c: glycated hemoglobin

**Table S2. Associations between ethnicity and standardized CMR metrics**

| CMR metrics | Asian | Black | Chinese | Mixed/Other |
| --- | --- | --- | --- | --- |
| LVEDVi, mL/m² | -0.32 [-0.37, -0.27] p<0.001 | -0.04 [-0.09, 0.02] p=0.275 | -0.19 [-0.25, -0.13] p<0.001 | -0.05 [-0.1, 0] p=0.078 |
| LVESVi, mL/m² | -0.16 [-0.19, -0.14] p<0.001 | 0.02 [-0.01, 0.05] p=0.344 | -0.12 [-0.16, -0.09] p<0.001 | -0.05 [-0.07, -0.02] p=0.003 |
| LVSVi, mL/m² | -0.14 [-0.17, -0.11] p<0.001 | -0.06 [-0.1, -0.02] p=0.004 | -0.05 [-0.09, -0.01] p=0.012 | 0.01 [-0.03, 0.04] p=0.823 |
| LVEF, % | 0.08 [0.05, 0.1] p<0.001 | -0.04 [-0.06, -0.01] p=0.014 | 0.08 [0.05, 0.1] p<0.001 | 0.03 [0.01, 0.06] p=0.006 |
| LVM/LVEDV, ratio | 0 [-0.04, 0.03] p=0.843 | 0.14 [0.1, 0.18] p<0.001 | 0 [-0.04, 0.05] p=0.847 | 0 [-0.03, 0.04] p=0.875 |
| LVMi, g/m² | -0.2 [-0.23, -0.17] p<0.001 | 0.08 [0.05, 0.12] p<0.001 | -0.11 [-0.14, -0.08] p<0.001 | -0.02 [-0.05, 0.01] p=0.188 |
| RVEDVi, mL/m² | -0.35 [-0.4, -0.3] p<0.001 | 0.02 [-0.04, 0.08] p=0.681 | -0.17 [-0.23, -0.11] p<0.001 | -0.03 [-0.08, 0.02] p=0.369 |
| RVESVi, mL/m² | -0.16 [-0.19, -0.13] p<0.001 | 0.01 [-0.03, 0.04] p=0.698 | -0.05 [-0.09, -0.02] p=0.006 | -0.02 [-0.05, 0.01] p=0.199 |
| RVSVi, mL/m² | -0.17 [-0.21, -0.14] p<0.001 | 0 [-0.04, 0.04] p=0.982 | -0.12 [-0.16, -0.07] p<0.001 | 0 [-0.04, 0.03] p=0.964 |
| RVEF, % | 0.04 [0.02, 0.06] p<0.001 | -0.01 [-0.03, 0.02] p=0.677 | 0 [-0.03, 0.02] p=0.847 | 0.02 [-0.01, 0.04] p=0.216 |
| LAESVi, mL/m² | -0.05 [-0.09, -0.02] p=0.006 | 0.01 [-0.03, 0.06] p=0.681 | -0.09 [-0.13, -0.04] p<0.001 | 0 [-0.04, 0.03] p=0.847 |
| LAVi max, mL/m² | -0.08 [-0.12, -0.05] p<0.001 | 0 [-0.05, 0.04] p=0.861 | -0.09 [-0.14, -0.05] p<0.001 | -0.01 [-0.05, 0.03] p=0.679 |
| LAEF, % | 0.02 [-0.01, 0.06] p=0.171 | -0.04 [-0.08, -0.01] p=0.035 | -0.08 [-0.12, -0.05] p<0.001 | 0.02 [-0.01, 0.05] p=0.227 |
| RAESVi, mL/m² | -0.13 [-0.18, -0.09] p<0.001 | 0.04 [-0.02, 0.09] p=0.232 | -0.1 [-0.15, -0.04] p=0.001 | 0.03 [-0.02, 0.08] p=0.313 |
| RAVi max, L/min/m² | -0.15 [-0.2, -0.11] p<0.001 | 0.03 [-0.02, 0.09] p=0.325 | -0.11 [-0.17, -0.05] p<0.001 | 0.02 [-0.03, 0.07] p=0.474 |
| RAEF, % | -0.03 [-0.07, 0] p=0.146 | -0.2 [-0.24, -0.16] p<0.001 | 0.04 [-0.01, 0.08] p=0.164 | -0.08 [-0.11, -0.04] p<0.001 |

**Footnote:** Results from linear regression models with each CMR metric as the outcome. Values are presented as standardized β coefficients with 95% confidence intervals. P-values were adjusted for multiple comparisons using the Benjamini-Hochberg false discovery rate (FDR) method. Models were adjusted for age (standardized) and sex. The White ethnic group served as the reference category.

**Table S3:** **Baseline prevalence and incident cardiovascular outcomes in the reference range validation cohort**

| **Outcome** | **Disease-free at imaging, n (%)** | **Prevalent at imaging, n (%)** | **Incident after imaging, n (%) among disease-free** |
| --- | --- | --- | --- |
| Hypertension | 50,264 (83.9) | 9,652 (16.1) | 3,702 (7.4) |
| Valvular heart disease | 59,308 (99.0) | 608 (1.0) | 880 (1.5) |
| Myocardial infarction | 58,677 (97.9) | 1,239 (2.1) | 611 (1.0) |
| Peripheral vascular disease | 59,379 (99.1) | 537 (0.9) | 569 (1.0) |
| Ischaemic heart disease | 58,694 (98.0) | 1,222 (2.0) | 555 (0.9) |
| Heart failure | 59,553 (99.4) | 363 (0.6) | 470 (0.8) |
| Cerebrovascular disease | 59,591 (99.5) | 325 (0.5) | 454 (0.8) |
| Mitral valve disease | 59,659 (99.6) | 257 (0.4) | 281 (0.5) |
| Aortic stenosis | 59,820 (99.8) | 96 (0.2) | 212 (0.4) |
| Non-ischaemic cardiomyopathy | 59,779 (99.8) | 137 (0.2) | 116 (0.2) |
| Atrial fibrillation | 59,777 (99.8) | 139 (0.2) | 59 (0.1) |
| Any major CVD | 56,536 (94.4) | 3,380 (5.6) | 2,192 (3.9) |
| All cause mortality | 59,916 (100.0) |  | 974 (1.6) |
| CVD mortality | 59,916 (100.0) |  | 106 (0.2) |

**Footnote:** Percentages for disease-free and prevalent status were calculated using the analysis-eligible validation cohort (n = 59,916). Administrative censoring date: 31/03/2023. Incident outcome percentages were calculated among participants free of the relevant condition at the time of imaging. For mortality outcomes, prevalent disease at imaging was not applicable.

**Table 4: Associations of percentile-based CMR categories with incident cardiovascular outcomes and mortality in the validation cohort**

|  |  | Mild | | At least moderate | |  |  |
| --- | --- | --- | --- | --- | --- | --- | --- |
| Outcome | CMR metric | HR (95% CI) | P value | HR (95% CI) | P value | Model (n) | Events (n) |
| Hypertension | LVEF | 1.29 (1.07, 1.54) | 0.007 | 1.54 (1.31, 1.81) | <0.001 | 47766 | 3508 |
| Hypertension | LVESVi | 1.32 (1.10, 1.58) | 0.003 | 1.63 (1.39, 1.91) | <0.001 | 48081 | 3538 |
| Hypertension | LVEDVi | 1.02 (0.81, 1.27) | 0.89 | 1.30 (1.01, 1.68) | 0.045 | 47070 | 3477 |
| Hypertension | LVSVi | 1.07 (0.87, 1.32) | 0.51 | 0.97 (0.75, 1.25) | 0.824 | 47511 | 3502 |
| Hypertension | LVMi | 1.96 (1.63, 2.37) | <0.001 | 2.23 (1.80, 2.76) | <0.001 | 47630 | 3533 |
| Hypertension | RVEF | 0.99 (0.85, 1.15) | 0.884 | 1.05 (0.90, 1.22) | 0.565 | 48132 | 3548 |
| Hypertension | RVESVi | 0.99 (0.83, 1.17) | 0.888 | 0.93 (0.77, 1.12) | 0.424 | 48296 | 3553 |
| Hypertension | RVEDVi | 0.98 (0.79, 1.23) | 0.879 | 0.83 (0.59, 1.17) | 0.286 | 47352 | 3486 |
| Hypertension | RVSVi | 1.10 (0.93, 1.31) | 0.259 | 1.07 (0.86, 1.33) | 0.545 | 47896 | 3546 |
| VHD | LVEF | 1.60 (1.19, 2.14) | 0.002 | 2.55 (2.01, 3.23) | <0.001 | 56252 | 823 |
| VHD | LVESVi | 1.83 (1.37, 2.43) | <0.001 | 3.98 (3.25, 4.88) | <0.001 | 56609 | 832 |
| VHD | LVEDVi | 2.00 (1.48, 2.70) | <0.001 | 5.56 (4.26, 7.25) | <0.001 | 55354 | 826 |
| VHD | LVSVi | 1.19 (0.84, 1.68) | 0.321 | 1.33 (0.90, 1.98) | 0.157 | 56034 | 783 |
| VHD | LVMi | 3.21 (2.43, 4.25) | <0.001 | 6.67 (5.15, 8.64) | <0.001 | 56174 | 828 |
| VHD | RVEF | 1.20 (0.94, 1.54) | 0.137 | 1.69 (1.35, 2.13) | <0.001 | 56726 | 838 |
| VHD | RVESVi | 1.74 (1.35, 2.23) | <0.001 | 2.56 (2.02, 3.24) | <0.001 | 56905 | 835 |
| VHD | RVEDVi | 1.56 (1.12, 2.17) | 0.009 | 2.17 (1.42, 3.30) | <0.001 | 55663 | 823 |
| VHD | RVSVi | 0.92 (0.67, 1.26) | 0.595 | 1.26 (0.89, 1.78) | 0.191 | 56499 | 825 |
| MI | LVEF | 1.41 (1.00, 1.99) | 0.048 | 2.29 (1.72, 3.05) | <0.001 | 55660 | 576 |
| MI | LVESVi | 1.33 (0.94, 1.88) | 0.111 | 2.59 (1.97, 3.40) | <0.001 | 56009 | 583 |
| MI | LVEDVi | 1.24 (0.85, 1.83) | 0.267 | 1.82 (1.16, 2.85) | 0.009 | 54766 | 573 |
| MI | LVSVi | 1.13 (0.77, 1.65) | 0.542 | 1.32 (0.85, 2.06) | 0.22 | 55428 | 578 |
| MI | LVMi | 1.81 (1.24, 2.63) | 0.002 | 1.88 (1.20, 2.94) | 0.006 | 55590 | 578 |
| MI | RVEF | 1.16 (0.88, 1.54) | 0.286 | 1.32 (1.00, 1.75) | 0.051 | 56134 | 579 |
| MI | RVESVi | 1.07 (0.77, 1.47) | 0.689 | 1.23 (0.88, 1.71) | 0.222 | 56315 | 581 |
| MI | RVEDVi | 0.91 (0.59, 1.40) | 0.663 | 0.90 (0.49, 1.66) | 0.742 | 55084 | 565 |
| MI | RVSVi | 1.08 (0.77, 1.50) | 0.669 | 1.20 (0.81, 1.79) | 0.36 | 55901 | 582 |
| IHD | LVEF | 1.72 (1.23, 2.41) | 0.002 | 2.41 (1.80, 3.22) | <0.001 | 55678 | 524 |
| IHD | LVESVi | 1.34 (0.94, 1.90) | 0.106 | 2.20 (1.64, 2.95) | <0.001 | 56033 | 532 |
| IHD | LVEDVi | 1.59 (1.10, 2.30) | 0.014 | 1.73 (1.08, 2.76) | 0.022 | 54790 | 520 |
| IHD | LVSVi | 1.59 (1.11, 2.29) | 0.013 | 1.69 (1.09, 2.60) | 0.018 | 55442 | 520 |
| IHD | LVMi | 1.59 (1.06, 2.38) | 0.024 | 2.81 (1.87, 4.22) | <0.001 | 55605 | 530 |
| IHD | RVEF | 1.04 (0.78, 1.40) | 0.773 | 1.50 (1.13, 1.98) | 0.004 | 56153 | 531 |
| IHD | RVESVi | 1.20 (0.87, 1.66) | 0.261 | 1.32 (0.94, 1.85) | 0.103 | 56329 | 529 |
| IHD | RVEDVi | 1.05 (0.69, 1.61) | 0.818 | 1.16 (0.65, 2.10) | 0.616 | 55099 | 513 |
| IHD | RVSVi | 1.19 (0.85, 1.67) | 0.302 | 1.53 (1.04, 2.25) | 0.031 | 55915 | 528 |
| HF | LVEF | 2.78 (2.01, 3.84) | <0.001 | 8.81 (7.08, 10.97) | <0.001 | 56484 | 437 |
| HF | LVESVi | 2.61 (1.88, 3.62) | <0.001 | 8.98 (7.25, 11.13) | <0.001 | 56843 | 444 |
| HF | LVEDVi | 2.38 (1.67, 3.40) | <0.001 | 7.55 (5.53, 10.29) | <0.001 | 55576 | 440 |
| HF | LVSVi | 1.95 (1.36, 2.81) | <0.001 | 2.47 (1.64, 3.71) | <0.001 | 56246 | 427 |
| HF | LVMi | 3.52 (2.50, 4.95) | <0.001 | 5.96 (4.24, 8.39) | <0.001 | 56413 | 438 |
| HF | RVEF | 1.39 (1.04, 1.86) | 0.027 | 3.95 (3.14, 4.97) | <0.001 | 56961 | 447 |
| HF | RVESVi | 1.59 (1.16, 2.19) | 0.004 | 3.77 (2.89, 4.92) | <0.001 | 57142 | 445 |
| HF | RVEDVi | 1.23 (0.80, 1.90) | 0.338 | 2.43 (1.47, 4.01) | <0.001 | 55891 | 437 |
| HF | RVSVi | 1.22 (0.86, 1.73) | 0.258 | 3.01 (2.15, 4.20) | <0.001 | 56725 | 439 |
| Any major CVD | LVEF | 1.57 (1.27, 1.94) | <0.001 | 2.74 (2.32, 3.23) | <0.001 | 53639 | 2075 |
| Any major CVD | LVESVi | 1.62 (1.32, 1.99) | <0.001 | 2.92 (2.48, 3.43) | <0.001 | 53982 | 2090 |
| Any major CVD | LVEDVi | 1.59 (1.27, 2.00) | <0.001 | 3.16 (2.50, 3.98) | <0.001 | 52790 | 2054 |
| Any major CVD | LVSVi | 1.50 (1.19, 1.88) | <0.001 | 1.66 (1.28, 2.16) | <0.001 | 53445 | 2041 |
| Any major CVD | LVMi | 2.29 (1.83, 2.85) | <0.001 | 3.87 (3.10, 4.82) | <0.001 | 53587 | 2075 |
| Any major CVD | RVEF | 1.08 (0.90, 1.29) | 0.417 | 1.61 (1.36, 1.90) | <0.001 | 54108 | 2095 |
| Any major CVD | RVESVi | 1.30 (1.07, 1.58) | 0.007 | 1.66 (1.38, 2.01) | <0.001 | 54281 | 2094 |
| Any major CVD | RVEDVi | 1.04 (0.79, 1.36) | 0.792 | 1.68 (1.22, 2.31) | 0.002 | 53108 | 2045 |
| Any major CVD | RVSVi | 1.09 (0.88, 1.36) | 0.432 | 1.49 (1.18, 1.89) | <0.001 | 53873 | 2082 |
| CVD mortality | LVEF | 1.18 (0.72, 1.92) | 0.508 | 2.21 (1.44, 3.39) | <0.001 | 56828 | 96 |
| CVD mortality | LVESVi | 0.97 (0.60, 1.58) | 0.91 | 2.27 (1.49, 3.48) | <0.001 | 57190 | 96 |
| CVD mortality | LVEDVi | 1.24 (0.73, 2.12) | 0.426 | 2.19 (1.15, 4.15) | 0.017 | 55917 | 95 |
| CVD mortality | LVSVi | 1.15 (0.69, 1.91) | 0.597 | 0.96 (0.51, 1.81) | 0.902 | 56587 | 95 |
| CVD mortality | LVMi | 1.21 (0.69, 2.15) | 0.504 | 2.00 (1.04, 3.85) | 0.037 | 56750 | 96 |
| CVD mortality | RVEF | 1.15 (0.79, 1.69) | 0.457 | 1.06 (0.71, 1.57) | 0.786 | 57306 | 96 |
| CVD mortality | RVESVi | 1.20 (0.78, 1.84) | 0.416 | 1.07 (0.67, 1.70) | 0.792 | 57486 | 96 |
| CVD mortality | RVEDVi | 0.92 (0.53, 1.62) | 0.779 | 1.17 (0.54, 2.58) | 0.688 | 56227 | 93 |
| CVD mortality | RVSVi | 0.96 (0.62, 1.50) | 0.871 | 0.87 (0.50, 1.51) | 0.623 | 57072 | 95 |

**Footnote:** Hazard ratios (HRs) and 95% confidence intervals (CIs) were derived from Cox proportional hazards models adjusted for age, sex, and body mass index at imaging, using the within-reference-range category as the reference group. Mild denotes values outside the reference range but below the at least moderate threshold. Models were fitted separately for each CMR metric and outcome among participants free of the relevant outcome at baseline. N model indicates the number of participants included in each model, and n events the number of incident events.

Abbreviations: CMR, cardiac magnetic resonance; HR, hazard ratio; CI, confidence interval; LVEDVi, left ventricular end-diastolic volume indexed to body surface area; LVESVi, left ventricular end-systolic volume indexed to body surface area; LVSVi, left ventricular stroke volume indexed to body surface area; LVMi, left ventricular mass indexed to body surface area; LVEF, left ventricular ejection fraction; RVEDVi, right ventricular end-diastolic volume indexed to body surface area; RVESVi, right ventricular end-systolic volume indexed to body surface area; RVSVi, right ventricular stroke volume indexed to body surface area; RVEF, right ventricular ejection fraction; VHD, valvular heart disease; MI, myocardial infarction; IHD, ischemic heart disease; HF, heart failure; CVD, cardiovascular disease.
