## Supplementary material for "Cardiovascular magnetic resonance reference ranges and centile-based categories for atrial and ventricular cardiac structure and function: 2026 Update from the Healthy Hearts Consortium": Non ethnicity statified reference ranges

**Reference ranges and centile-based cardiac chamber measurements using the Healthy Hearts Consortium data**

Values reported in this document are not stratified by participant ethnicity.

Details about the consortium data may be found at: <https://healthy-hearts.org.uk/>

Reference ranges and centile-based measurements were calculated using the GLMSS package in R. Details may be found at <https://www.gamlss.com/> and <https://cran.r-project.org/web/packages/gamlss/index.html>.

In the tables ‘Reference range’, ‘Mild’ and ‘At least moderate’ capture the progression of deviation from the reference range for each metric. Reference ranges are defined as the interval between the 2.5th and 97.5th centiles, derived from age-specific generalized additive models for location, scale and shape (GAMLSS) using the Box–Cox Cole distribution. Values falling in the 0.5th–2.5th or 97.5th–99.5th centiles are classified as mild deviation, and those beyond the 0.5th or 99.5th centiles as at least moderate deviation. These centile-based categories describe the position of a measurement within the reference distribution rather than disease severity or diagnostic abnormality.

Please report any concerns to:

Table of content

### 1. Table: Reference ranges and centile-based categories for cardiac chamber measurements derived using cvi42 software (smooth segmentation), indexed to height, in an adult population

|  |  | **Female** | | | | | **Male** | | | |
| --- | --- | --- | --- | --- | --- | --- | --- | --- | --- | --- |
| **Age group** | **Metrics** | **Reference range** | **Mild** | **At least moderate** | **n** | **Reference range** | | **Mild** | **At least moderate** | **n** |
| 18-29 | LVEDVi (mL/m²) | 60-107 | 108-116 | > 116 | 118 | 73-128 | | 129-136 | > 136 | 118 |
|  | LVESVi (mL/m²) | 17-46 | 47-50 | > 50 | 118 | 25-58 | | 59-63 | > 63 | 113 |
|  | LVSVi (mL/m²) | 35-67 | 31-34 | < 31 | 118 | 39-77 | | 33-38 | < 33 | 118 |
|  | LVEF (%) | 51-71 | 48-50 | < 48 | 116 | 48-67 | | 45-47 | < 45 | 119 |
|  | LVCOi (L/min/m²) | 2.2-4.9 | 2.0-2.1 | < 2 | 118 | 2.4-5.2 | | 1.9-2.3 | < 1.9 | 116 |
|  | LVMi (g/m²) | 31-57 | 58-62 | > 62 | 118 | 44-80 | | 81-86 | > 86 | 124 |
|  | LVM/LVEDV (G/ML) | 0.4-0.7 |  | > 0.7 | 118 | 0.5-0.8 | |  | > 0.8 | 118 |
|  | RVEDVi (mL/m²) | 58-113 | 114-123 | > 123 | 118 | 75-138 | | 139-146 | > 146 | 117 |
|  | RVESVi (mL/m²) | 14-56 | 57-61 | > 61 | 116 | 28-70 | | 71-76 | > 76 | 102 |
|  | RVEF (%) | 44-69 | 40-43 | < 40 | 113 | 42-66 | | 39-41 | < 39 | 116 |
|  | RVCOi (L/min/m²) | 2.0-5.3 | 1.7-1.9 | < 1.7 | 117 | 2.4-5.6 | | 2.0-2.3 | < 2 | 116 |
|  | RVSVi (mL/m²) | 32-69 | 27-31 | < 27 | 118 | 39-81 | | 33-38 | < 33 | 117 |
|  | LAVi max (mL/m²) | 24-51 | 52-56 | > 56 | 63 | 27-62 | | 63-69 | > 69 | 68 |
|  | LAEF (%) | 55-75 | 52-54 | < 52 | 63 | 53-72 | | 50-52 | < 50 | 68 |
|  | RAVi max (mL/m²) | 21-51 | 52-56 | > 56 | 95 | 27-68 | | 69-78 | > 78 | 100 |
|  | RAEF (%) | 39-77 | 33-38 | < 33 | 93 | 35-73 | | 31-34 | < 31 | 100 |
| 30-39 | LVEDVi (mL/m²) | 58-106 | 107-115 | > 115 | 155 | 67-122 | | 123-130 | > 130 | 147 |
|  | LVESVi (mL/m²) | 17-46 | 47-50 | > 50 | 155 | 23-57 | | 58-62 | > 62 | 146 |
|  | LVSVi (mL/m²) | 34-66 | 30-33 | < 30 | 152 | 37-75 | | 31-36 | < 31 | 148 |
|  | LVEF (%) | 51-71 | 48-50 | < 48 | 150 | 48-69 | | 46-47 | < 46 | 151 |
|  | LVCOi (L/min/m²) | 2.2-4.9 | 2-2.1 | < 2 | 153 | 2.4-5.2 | | 1.9-2.3 | < 1.9 | 144 |
|  | LVMi (g/m²) | 31-58 | 59-63 | > 63 | 155 | 41-78 | | 79-84 | > 84 | 151 |
|  | LVM/LVEDV (G/ML) | 0.4-0.7 |  | > 0.7 | 155 | 0.5-0.8 | | 0.9-0.9 | > 0.9 | 148 |
|  | RVEDVi (mL/m²) | 57-114 | 115-124 | > 124 | 155 | 72-136 | | 137-145 | > 145 | 147 |
|  | RVESVi (mL/m²) | 15-52 | 53-58 | > 58 | 153 | 26-66 | | 67-73 | > 73 | 142 |
|  | RVEF (%) | 47-70 | 43-46 | < 43 | 145 | 44-67 | | 41-43 | < 41 | 148 |
|  | RVCOi (L/min/m²) | 2-5.2 | 1.7-1.9 | < 1.7 | 151 | 2.4-5.5 | | 1.9-2.3 | < 1.9 | 140 |
|  | RVSVi (mL/m²) | 32-70 | 28-31 | < 28 | 152 | 38-80 | | 32-37 | < 32 | 149 |
|  | LAVi max (mL/m²) | 24-54 | 55-60 | > 60 | 102 | 26-62 | | 63-70 | > 70 | 80 |
|  | LAEF (%) | 55-76 | 52-54 | < 52 | 101 | 52-73 | | 49-51 | < 49 | 80 |
|  | RAVi max (mL/m²) | 21-51 | 52-57 | > 57 | 131 | 26-69 | | 70-78 | > 78 | 135 |
|  | RAEF (%) | 38-74 | 33-37 | < 33 | 128 | 36-71 | | 32-35 | < 32 | 134 |
| 40-49 | LVEDVi (mL/m²) | 58-108 | 109-118 | > 118 | 568 | 66-125 | | 126-134 | > 134 | 483 |
|  | LVESVi (mL/m²) | 17-46 | 47-50 | > 50 | 568 | 22-55 | | 56-60 | > 60 | 478 |
|  | LVSVi (mL/m²) | 35-68 | 31-34 | < 31 | 567 | 37-77 | | 31-36 | < 31 | 489 |
|  | LVEF (%) | 51-72 | 48-50 | < 48 | 561 | 49-70 | | 46-48 | < 46 | 489 |
|  | LVCOi (L/min/m²) | 2.2-4.9 | 1.9-2.1 | < 1.9 | 565 | 2.3-5.1 | | 1.9-2.2 | < 1.9 | 480 |
|  | LVMi (g/m²) | 32-60 | 61-65 | > 65 | 568 | 41-80 | | 81-87 | > 87 | 499 |
|  | LVM/LVEDV (G/ML) | 0.4-0.7 | 0.8-0.8 | > 0.8 | 568 | 0.5-0.8 | | 0.9-0.9 | > 0.9 | 483 |
|  | RVEDVi (mL/m²) | 56-114 | 115-125 | > 125 | 567 | 70-137 | | 138-147 | > 147 | 485 |
|  | RVESVi (mL/m²) | 16-49 | 50-54 | > 54 | 567 | 25-63 | | 64-69 | > 69 | 474 |
|  | RVEF (%) | 50-71 | 46-49 | < 46 | 559 | 46-68 | | 42-45 | < 42 | 483 |
|  | RVCOi (L/min/m²) | 2.1-5 | 1.8-2 | < 1.8 | 564 | 2.3-5.4 | | 1.9-2.2 | < 1.9 | 471 |
|  | RVSVi (mL/m²) | 33-71 | 29-32 | < 29 | 566 | 38-82 | | 32-37 | < 32 | 483 |
|  | LAVi max (mL/m²) | 25-59 | 60-65 | > 65 | 487 | 25-63 | | 64-71 | > 71 | 409 |
|  | LAEF (%) | 55-77 | 51-54 | < 51 | 481 | 53-76 | | 50-52 | < 50 | 404 |
|  | RAVi max (mL/m²) | 23-59 | 60-66 | > 66 | 549 | 28-77 | | 78-87 | > 87 | 462 |
|  | RAEF (%) | 38-71 | 33-37 | < 33 | 546 | 35-69 | | 31-34 | < 31 | 464 |
| 50-59 | LVEDVi (mL/m²) | 57-109 | 110-119 | > 119 | 1289 | 63-123 | | 124-133 | > 133 | 1130 |
|  | LVESVi (mL/m²) | 16-45 | 46-49 | > 49 | 1287 | 20-54 | | 55-59 | > 59 | 1125 |
|  | LVSVi (mL/m²) | 35-69 | 31-34 | < 31 | 1288 | 36-76 | | 30-35 | < 30 | 1139 |
|  | LVEF (%) | 51-73 | 48-50 | < 48 | 1283 | 49-71 | | 46-48 | < 46 | 1148 |
|  | LVCOi (L/min/m²) | 2.1-4.7 | 1.8-2 | < 1.8 | 1282 | 2.2-5 | | 1.8-2.1 | < 1.8 | 1141 |
|  | LVMi (g/m²) | 33-62 | 63-68 | > 68 | 1289 | 41-82 | | 83-89 | > 89 | 1157 |
|  | LVM/LVEDV (G/ML) | 0.5-0.7 | 0.8-0.8 | > 0.8 | 1289 | 0.5-0.9 | | 01-1 | > 1 | 1126 |
|  | RVEDVi (mL/m²) | 55-112 | 113-123 | > 123 | 1287 | 65-132 | | 133-143 | > 143 | 1137 |
|  | RVESVi (mL/m²) | 16-46 | 47-51 | > 51 | 1287 | 23-60 | | 61-66 | > 66 | 1137 |
|  | RVEF (%) | 52-72 | 49-51 | < 49 | 1284 | 47-69 | | 44-46 | < 44 | 1154 |
|  | RVCOi (L/min/m²) | 2-4.9 | 1.7-1.9 | < 1.7 | 1282 | 2.2-5.2 | | 1.8-2.1 | < 1.8 | 1137 |
|  | RVSVi (mL/m²) | 33-71 | 29-32 | < 29 | 1286 | 36-79 | | 29-35 | < 29 | 1140 |
|  | LAVi max (mL/m²) | 24-61 | 62-68 | > 68 | 1144 | 23-64 | | 65-72 | > 72 | 962 |
|  | LAEF (%) | 53-78 | 50-52 | < 50 | 1132 | 53-80 | | 50-52 | < 50 | 958 |
|  | RAVi max (mL/m²) | 24-66 | 67-74 | > 74 | 1215 | 26-77 | | 78-86 | > 86 | 1072 |
|  | RAEF (%) | 37-69 | 32-36 | < 32 | 1200 | 35-68 | | 31-34 | < 31 | 1079 |
| 60-69 | LVEDVi (mL/m²) | 54-104 | 105-114 | > 114 | 1209 | 59-120 | | 121-130 | > 130 | 1056 |
|  | LVESVi (mL/m²) | 16-43 | 44-49 | > 49 | 1210 | 18-52 | | 53-57 | > 57 | 1045 |
|  | LVSVi (mL/m²) | 33-66 | 29-32 | < 29 | 1209 | 34-74 | | 29-33 | < 29 | 1059 |
|  | LVEF (%) | 52-74 | 48-51 | < 48 | 1203 | 49-72 | | 45-48 | < 45 | 1052 |
|  | LVCOi (L/min/m²) | 2-4.5 | 1.7-1.9 | < 1.7 | 1208 | 2.1-4.8 | | 1.8-2 | < 1.8 | 1056 |
|  | LVMi (g/m²) | 32-62 | 63-69 | > 69 | 1210 | 40-81 | | 82-88 | > 88 | 1065 |
|  | LVM/LVEDV (G/ML) | 0.5-0.8 | 0.9-0.9 | > 0.9 | 1209 | 0.5-0.9 | |  | > 1 | 1054 |
|  | RVEDVi (mL/m²) | 53-108 | 109-119 | > 119 | 1208 | 62-128 | | 129-139 | > 139 | 1058 |
|  | RVESVi (mL/m²) | 15-44 | 45-49 | > 49 | 1211 | 21-57 | | 58-62 | > 62 | 1061 |
|  | RVEF (%) | 52-73 | 49-51 | < 49 | 1205 | 48-70 | | 44-47 | < 44 | 1064 |
|  | RVCOi (L/min/m²) | 2-4.7 | 1.7-1.9 | < 1.7 | 1208 | 2.1-5 | | 1.7-2 | < 1.7 | 1059 |
|  | RVSVi (mL/m²) | 33-69 | 28-32 | < 28 | 1208 | 34-77 | | 27-33 | < 27 | 1054 |
|  | LAVi max (mL/m²) | 22-61 | 62-70 | > 70 | 1062 | 22-65 | | 66-73 | > 73 | 879 |
|  | LAEF (%) | 51-79 | 47-50 | < 47 | 1036 | 51-80 | | 47-50 | < 47 | 864 |
|  | RAVi max (mL/m²) | 23-66 | 67-75 | > 75 | 1129 | 25-78 | | 79-87 | > 87 | 992 |
|  | RAEF (%) | 37-70 | 32-36 | < 32 | 1115 | 35-70 | | 31-34 | < 31 | 1000 |
| 70+ | LVEDVi (mL/m²) | 51-100 | 101-111 | > 111 | 1209 | 55-117 | | 118-127 | > 127 | 1343 |
|  | LVESVi (mL/m²) | 15-42 | 43-48 | > 48 | 1209 | 16-51 | | 52-56 | > 56 | 1332 |
|  | LVSVi (mL/m²) | 31-64 | 28-30 | < 28 | 1210 | 32-71 | | 27-31 | < 27 | 1347 |
|  | LVEF (%) | 52-74 | 48-51 | < 48 | 1201 | 48-72 | | 45-47 | < 45 | 1333 |
|  | LVCOi (L/min/m²) | 1.9-4.4 | 1.7-1.8 | < 1.7 | 1208 | 2-4.6 | | 1.7-1.9 | < 1.7 | 1349 |
|  | LVMi (g/m²) | 32-62 | 63-68 | > 68 | 1211 | 39-78 | | 79-85 | > 85 | 1355 |
|  | LVM/LVEDV (G/ML) | 0.5-0.8 | 0.9-0.9 | > 0.9 | 1209 | 0.5-0.9 | | 1-1.1 | > 1.1 | 1342 |
|  | RVEDVi (mL/m²) | 51-103 | 104-114 | > 114 | 1209 | 60-124 | | 125-135 | > 135 | 1353 |
|  | RVESVi (mL/m²) | 15-42 | 43-46 | > 46 | 1209 | 20-54 | | 55-59 | > 59 | 1353 |
|  | RVEF (%) | 53-74 | 49-52 | < 49 | 1202 | 48-70 | | 44-47 | < 44 | 1341 |
|  | RVCOi (L/min/m²) | 1.9-4.6 | 1.7-1.8 | < 1.7 | 1208 | 2-4.8 | | 1.7-1.9 | < 1.7 | 1346 |
|  | RVSVi (mL/m²) | 32-66 | 27-31 | < 27 | 1208 | 32-75 | | 25-31 | < 25 | 1351 |
|  | LAVi max (mL/m²) | 21-63 | 64-73 | > 73 | 1041 | 21-66 | | 67-75 | > 75 | 1032 |
|  | LAEF (%) | 48-79 | 44-47 | < 44 | 1002 | 48-80 | | 44-47 | < 44 | 991 |
|  | RAVi max (mL/m²) | 22-68 | 69-77 | > 77 | 1130 | 23-78 | | 79-88 | > 88 | 1204 |
|  | RAEF (%) | 37-72 | 32-36 | < 32 | 1102 | 35-73 | | 31-34 | < 31 | 1200 |

Footnote: All measurements were derived using cvi42 software with smooth endocardial segmentation; papillary muscles and trabeculations were excluded from left ventricular mass measurements. Volumetric and mass measurements were indexed to height. Reference ranges are defined as the interval between the 2.5th and 97.5th centiles derived from age-specific generalized additive models for location, scale and shape (GAMLSS) using the Box–Cox Cole–Green distribution. Centile-based categories beyond the reference range are derived from LMS-based thresholds (P0.5, P2.5, P97.5, and P99.5) and describe the position of a measurement within the reference distribution rather than disease severity or diagnostic abnormality. Reference ranges are based on a White an adult population. Age groups are given in years.

Abbreviations: LV = left ventricle; RV = right ventricle; LA = left atrium; RA = right atrium; EDV = end-diastolic volume; ESV = end-systolic volume; SV = stroke volume; EF = ejection fraction; CO = cardiac output.

### 2. Table: Reference ranges and centile-based categories for cardiac chamber measurements derived using cvi42 software (anatomical segmentation), indexed to height, in an adult population

|  |  | **Female** | | | | **Male** | | | |
| --- | --- | --- | --- | --- | --- | --- | --- | --- | --- |
| **Age group** | **Metrics** | **Reference range** | **Mild** | **At least moderate** | **n** | **Reference range** | **Mild** | **At least moderate** | **n** |
| 18-29 | LVEDVi (mL/m²) | 53-98 | 99-105 | > 105 | 118 | 66-121 | 122-129 | > 129 | 122 |
|  | LVESVi (mL/m²) | 14-38 | 39-41 | > 41 | 118 | 18-52 | 53-58 | > 58 | 123 |
|  | LVSVi (mL/m²) | 34-65 | 29-33 | < 29 | 118 | 39-78 | 33-38 | < 33 | 120 |
|  | LVEF (%) | 54-74 | 50-53 | < 50 | 116 | 52-72 | 48-51 | < 48 | 121 |
|  | LVCOi (L/min/m²) | 2.2-4.8 | 2-2.1 | < 2 | 118 | 2.4-5.3 | 1.9-2.3 | < 1.9 | 120 |
|  | LVMi (g/m²) | 37-67 | 68-72 | > 72 | 118 | 49-91 | 92-97 | > 97 | 123 |
|  | LVM/LVEDV (G/ML) | 0.6-0.9 | 01-1 | > 1 | 118 | 0.6-1 | 1.1-1.1 | > 1.1 | 121 |
|  | RVEDVi (mL/m²) | 52-105 | 106-113 | > 113 | 118 | 64-135 | 136-144 | > 144 | 121 |
|  | RVESVi (mL/m²) | 13-48 | 49-52 | > 52 | 116 | 24-64 | 65-70 | > 70 | 112 |
|  | RVEF (%) | 47-72 | 44-46 | < 44 | 113 | 45-68 | 42-44 | < 42 | 115 |
|  | RVCOi (L/min/m²) | 1.9-5.2 | 1.6-1.8 | < 1.6 | 117 | 2.3-5.5 | 1.8-2.2 | < 1.8 | 117 |
|  | RVSVi (mL/m²) | 30-67 | 26-29 | < 26 | 118 | 37-82 | 31-36 | < 31 | 118 |
|  | LAVi max (mL/m²) | 24-51 | 52-56 | > 56 | 63 | 27-62 | 63-69 | > 69 | 68 |
|  | LAEF (%) | 55-75 | 52-54 | < 52 | 63 | 53-72 | 50-52 | < 50 | 68 |
|  | RAVi max (mL/m²) | 21-51 | 52-56 | > 56 | 95 | 27-68 | 69-78 | > 78 | 100 |
|  | RAEF (%) | 39-77 | 33-38 | < 33 | 93 | 35-73 | 31-34 | < 31 | 100 |
| 30-39 | LVEDVi (mL/m²) | 52-96 | 97-104 | > 104 | 155 | 61-115 | 116-124 | > 124 | 151 |
|  | LVESVi (mL/m²) | 13-38 | 39-41 | > 41 | 154 | 17-50 | 51-56 | > 56 | 151 |
|  | LVSVi (mL/m²) | 33-65 | 29-32 | < 29 | 153 | 37-74 | 31-36 | < 31 | 150 |
|  | LVEF (%) | 55-75 | 51-54 | < 51 | 150 | 53-74 | 50-52 | < 50 | 151 |
|  | LVCOi (L/min/m²) | 2.2-4.8 | 2-2.1 | < 2 | 154 | 2.3-5.2 | 1.9-2.2 | < 1.9 | 145 |
|  | LVMi (g/m²) | 37-68 | 69-74 | > 74 | 155 | 47-89 | 90-95 | > 95 | 151 |
|  | LVM/LVEDV (G/ML) | 0.6-0.9 | 01-1 | > 1 | 155 | 0.6-1 | 1.1-1.2 | > 1.2 | 152 |
|  | RVEDVi (mL/m²) | 52-106 | 107-114 | > 114 | 155 | 64-133 | 134-143 | > 143 | 148 |
|  | RVESVi (mL/m²) | 14-48 | 49-53 | > 53 | 153 | 24-64 | 65-70 | > 70 | 146 |
|  | RVEF (%) | 48-71 | 45-47 | < 45 | 145 | 45-68 | 42-44 | < 42 | 148 |
|  | RVCOi (L/min/m²) | 02-5 | 1.7-1.9 | < 1.7 | 152 | 2.3-5.5 | 1.8-2.2 | < 1.8 | 142 |
|  | RVSVi (mL/m²) | 31-67 | 26-30 | < 26 | 153 | 36-79 | 30-35 | < 30 | 149 |
|  | LAVi max (mL/m²) | 24-54 | 55-60 | > 60 | 102 | 26-62 | 63-70 | > 70 | 80 |
|  | LAEF (%) | 55-76 | 52-54 | < 52 | 101 | 52-73 | 49-51 | < 49 | 80 |
|  | RAVi max (mL/m²) | 21-51 | 52-57 | > 57 | 131 | 26-69 | 70-78 | > 78 | 135 |
|  | RAEF (%) | 38-74 | 33-37 | < 33 | 128 | 36-71 | 32-35 | < 32 | 134 |
| 40-49 | LVEDVi (mL/m²) | 53-99 | 100-107 | > 107 | 568 | 59-117 | 118-127 | > 127 | 494 |
|  | LVESVi (mL/m²) | 13-37 | 38-41 | > 41 | 568 | 16-48 | 49-54 | > 54 | 494 |
|  | LVSVi (mL/m²) | 34-67 | 30-33 | < 30 | 568 | 37-76 | 31-36 | < 31 | 490 |
|  | LVEF (%) | 56-77 | 52-55 | < 52 | 562 | 54-76 | 51-53 | < 51 | 489 |
|  | LVCOi (L/min/m²) | 2.2-4.8 | 1.9-2.1 | < 1.9 | 565 | 2.3-5.1 | 1.9-2.2 | < 1.9 | 484 |
|  | LVMi (g/m²) | 37-70 | 71-77 | > 77 | 568 | 47-91 | 92-97 | > 97 | 484 |
|  | LVM/LVEDV (G/ML) | 0.6-0.9 | 01-1 | > 1 | 568 | 0.6-1.1 |  | > 1.2 | 481 |
|  | RVEDVi (mL/m²) | 54-110 | 111-120 | > 120 | 567 | 65-135 | 136-145 | > 145 | 485 |
|  | RVESVi (mL/m²) | 15-49 | 50-53 | > 53 | 567 | 24-63 | 64-69 | > 69 | 478 |
|  | RVEF (%) | 50-71 | 46-49 | < 46 | 558 | 46-68 | 42-45 | < 42 | 486 |
|  | RVCOi (L/min/m²) | 2-4.9 | 1.7-1.9 | < 1.7 | 564 | 2.2-5.3 | 1.8-2.1 | < 1.8 | 480 |
|  | RVSVi (mL/m²) | 32-69 | 28-31 | < 28 | 566 | 36-80 | 30-35 | < 30 | 486 |
|  | LAVi max (mL/m²) | 25-59 | 60-65 | > 65 | 487 | 25-63 | 64-71 | > 71 | 409 |
|  | LAEF (%) | 55-77 | 51-54 | < 51 | 481 | 53-76 | 50-52 | < 50 | 404 |
|  | RAVi max (mL/m²) | 23-59 | 60-66 | > 66 | 549 | 28-77 | 78-87 | > 87 | 462 |
|  | RAEF (%) | 38-71 | 33-37 | < 33 | 546 | 35-69 | 31-34 | < 31 | 464 |
| 50-59 | LVEDVi (mL/m²) | 52-99 | 100-108 | > 108 | 1289 | 56-116 | 117-126 | > 126 | 1157 |
|  | LVESVi (mL/m²) | 12-36 | 37-40 | > 40 | 1289 | 14-46 | 47-51 | > 51 | 1161 |
|  | LVSVi (mL/m²) | 34-68 | 30-33 | < 30 | 1288 | 36-75 | 30-35 | < 30 | 1145 |
|  | LVEF (%) | 57-78 | 53-56 | < 53 | 1283 | 54-77 | 51-53 | < 51 | 1155 |
|  | LVCOi (L/min/m²) | 2.1-4.6 | 1.8-2 | < 1.8 | 1284 | 2.2-4.9 | 1.8-2.1 | < 1.8 | 1141 |
|  | LVMi (g/m²) | 38-73 | 74-80 | > 80 | 1288 | 48-93 | 94-100 | > 100 | 1131 |
|  | LVM/LVEDV (G/ML) | 0.6-1 | 1.1-1.1 | > 1.1 | 1288 | 0.7-1.1 | 1.2-1.3 | > 1.3 | 1127 |
|  | RVEDVi (mL/m²) | 55-112 | 113-123 | > 123 | 1287 | 64-132 | 133-143 | > 143 | 1142 |
|  | RVESVi (mL/m²) | 16-48 | 49-53 | > 53 | 1287 | 23-61 | 62-67 | > 67 | 1131 |
|  | RVEF (%) | 51-71 | 47-50 | < 47 | 1284 | 46-68 | 43-45 | < 43 | 1153 |
|  | RVCOi (L/min/m²) | 2-4.7 | 1.7-1.9 | < 1.7 | 1282 | 2.1-5.1 | 1.7-2 | < 1.7 | 1145 |
|  | RVSVi (mL/m²) | 33-70 | 28-32 | < 28 | 1287 | 35-79 | 28-34 | < 28 | 1153 |
|  | LAVi max (mL/m²) | 24-61 | 62-68 | > 68 | 1144 | 23-64 | 65-72 | > 72 | 962 |
|  | LAEF (%) | 53-78 | 50-52 | < 50 | 1132 | 53-80 | 50-52 | < 50 | 958 |
|  | RAVi max (mL/m²) | 24-66 | 67-74 | > 74 | 1215 | 26-77 | 78-86 | > 86 | 1072 |
|  | RAEF (%) | 37-69 | 32-36 | < 32 | 1200 | 35-68 | 31-34 | < 31 | 1079 |
| 60-69 | LVEDVi (mL/m²) | 48-94 | 95-103 | > 103 | 1209 | 52-111 | 112-121 | > 121 | 1065 |
|  | LVESVi (mL/m²) | 11-34 | 35-39 | > 39 | 1211 | 13-44 | 45-49 | > 49 | 1063 |
|  | LVSVi (mL/m²) | 33-65 | 29-32 | < 29 | 1209 | 34-73 | 29-33 | < 29 | 1061 |
|  | LVEF (%) | 58-79 | 54-57 | < 54 | 1208 | 54-78 | 49-53 | < 49 | 1064 |
|  | LVCOi (L/min/m²) | 2-4.5 | 1.8-1.9 | < 1.8 | 1209 | 2.1-4.7 | 1.8-2 | < 1.8 | 1059 |
|  | LVMi (g/m²) | 38-73 | 74-81 | > 81 | 1210 | 47-92 | 93-100 | > 100 | 1052 |
|  | LVM/LVEDV (G/ML) | 0.6-1 | 1.1-1.2 | > 1.2 | 1209 | 0.7-1.2 | 1.3-1.4 | > 1.4 | 1049 |
|  | RVEDVi (mL/m²) | 53-107 | 108-117 | > 117 | 1208 | 62-127 | 128-137 | > 137 | 1059 |
|  | RVESVi (mL/m²) | 16-45 | 46-50 | > 50 | 1211 | 22-57 | 58-63 | > 63 | 1057 |
|  | RVEF (%) | 51-72 | 48-50 | < 48 | 1203 | 47-69 | 43-46 | < 43 | 1061 |
|  | RVCOi (L/min/m²) | 2-4.6 | 1.7-1.9 | < 1.7 | 1208 | 2.1-4.9 | 1.7-2 | < 1.7 | 1061 |
|  | RVSVi (mL/m²) | 32-67 | 28-31 | < 28 | 1209 | 33-76 | 27-32 | < 27 | 1060 |
|  | LAVi max (mL/m²) | 22-61 | 62-70 | > 70 | 1062 | 22-65 | 66-73 | > 73 | 879 |
|  | LAEF (%) | 51-79 | 47-50 | < 47 | 1036 | 51-80 | 47-50 | < 47 | 864 |
|  | RAVi max (mL/m²) | 23-66 | 67-75 | > 75 | 1129 | 25-78 | 79-87 | > 87 | 992 |
|  | RAEF (%) | 37-70 | 32-36 | < 32 | 1115 | 35-70 | 31-34 | < 31 | 1000 |
| 70+ | LVEDVi (mL/m²) | 46-91 | 92-101 | > 101 | 1209 | 49-107 | 108-117 | > 117 | 1349 |
|  | LVESVi (mL/m²) | 10-33 | 34-38 | > 38 | 1211 | 11-42 | 43-47 | > 47 | 1351 |
|  | LVSVi (mL/m²) | 32-63 | 28-31 | < 28 | 1209 | 33-71 | 27-32 | < 27 | 1349 |
|  | LVEF (%) | 58-81 | 54-57 | < 54 | 1202 | 54-79 | 50-53 | < 50 | 1338 |
|  | LVCOi (L/min/m²) | 1.9-4.4 | 1.7-1.8 | < 1.7 | 1209 | 2-4.5 | 1.7-1.9 | < 1.7 | 1351 |
|  | LVMi (g/m²) | 37-72 | 73-80 | > 80 | 1210 | 45-90 | 91-98 | > 98 | 1345 |
|  | LVM/LVEDV (G/ML) | 0.6-1.1 |  | > 1.2 | 1208 | 0.7-1.3 | 1.4-1.5 | > 1.5 | 1342 |
|  | RVEDVi (mL/m²) | 52-102 | 103-113 | > 113 | 1209 | 60-123 | 124-133 | > 133 | 1352 |
|  | RVESVi (mL/m²) | 16-42 | 43-47 | > 47 | 1209 | 21-55 | 56-60 | > 60 | 1352 |
|  | RVEF (%) | 51-72 | 47-50 | < 47 | 1202 | 47-69 | 44-46 | < 44 | 1338 |
|  | RVCOi (L/min/m²) | 1.9-4.5 | 1.7-1.8 | < 1.7 | 1208 | 2-4.7 | 1.6-1.9 | < 1.6 | 1350 |
|  | RVSVi (mL/m²) | 31-65 | 27-30 | < 27 | 1208 | 32-73 | 25-31 | < 25 | 1350 |
|  | LAVi max (mL/m²) | 21-63 | 64-73 | > 73 | 1041 | 21-66 | 67-75 | > 75 | 1032 |
|  | LAEF (%) | 48-79 | 44-47 | < 44 | 1002 | 48-80 | 44-47 | < 44 | 991 |
|  | RAVi max (mL/m²) | 22-68 | 69-77 | > 77 | 1130 | 23-78 | 79-88 | > 88 | 1204 |
|  | RAEF (%) | 37-72 | 32-36 | < 32 | 1102 | 35-73 | 31-34 | < 31 | 1200 |

Footnote: All measurements were derived using cvi42 software with anatomical endocardial segmentation; papillary muscles and trabeculations were included in left ventricular mass measurements. “Reference range,” “Mild,” and “At least moderate” capture the progression of deviation from the reference range for each metric. Reference ranges are defined as the interval between the 2.5th and 97.5th centiles, derived from age-specific generalized add–Green distribution. Centile-based categories beyond the reference range are derived from LMS-based thresholds (P0.5, P2.5, P97.5, and P99.5) and describe the position of a measurement within the reference distribution rather than disease severity or diagnostic abnormality. Reference ranges are based on a White an adult population.

Abbreviations: LV = left ventricle; RV = right ventricle; LA = left atrium; RA = right atrium; EDV = end-diastolic volume; ESV = end-systolic volume; SV = stroke volume; EF = ejection fraction; CO = cardiac output.

### 3. Table: Reference ranges and centile-based categories for cardiac chamber measurements derived using cvi42 software (smooth segmentation with papillary muscles added to the LV mass segmentation), indexed to height, in an adult population

|  |  | **Female** | | | | **Male** | | | |
| --- | --- | --- | --- | --- | --- | --- | --- | --- | --- |
| **Age group** | **Metrics** | **Reference range** | **Mild** | **At least moderate** | **n** | **Reference range** | **Mild** | **At least moderate** | **n** |
| 18-29 | LVEDVi (mL/m²) | 58-105 | 106-113 | > 113 | 118 | 70-125 | 126-133 | > 133 | 119 |
|  | LVESVi (mL/m²) | 15-43 | 44-46 | > 46 | 118 | 22-56 | 57-61 | > 61 | 119 |
|  | LVSVi (mL/m²) | 35-67 | 30-34 | < 30 | 118 | 39-78 | 33-38 | < 33 | 118 |
|  | LVEF (%) | 52-72 | 50-51 | < 50 | 116 | 49-69 | 46-48 | < 46 | 121 |
|  | LVCOi (L/min/m²) | 2.3-4.9 | 2-2.2 | < 2 | 117 | 2.4-5.3 | 1.9-2.3 | < 1.9 | 116 |
|  | LVMi (g/m²) | 33-60 | 61-65 | > 65 | 118 | 46-84 | 85-90 | > 90 | 124 |
|  | LVM/LVEDV (G/ML) | 0.5-0.7 | 0.8-0.8 | > 0.8 | 118 | 0.5-0.8 | 0.9-0.9 | > 0.9 | 119 |
|  | RVEDVi (mL/m²) | 55-108 | 109-117 | > 117 | 118 | 68-137 | 138-147 | > 147 | 120 |
|  | RVESVi (mL/m²) | 14-52 | 53-56 | > 56 | 116 | 23-65 | 66-70 | > 70 | 111 |
|  | RVEF (%) | 47-70 | 43-46 | < 43 | 113 | 43-67 | 40-42 | < 40 | 117 |
|  | RVCOi (L/min/m²) | 2-5.2 | 1.7-1.9 | < 1.7 | 117 | 2.4-5.6 | 1.9-2.3 | < 1.9 | 116 |
|  | RVSVi (mL/m²) | 31-68 | 27-30 | < 27 | 118 | 38-80 | 31-37 | < 31 | 117 |
|  | LAVi max (mL/m²) | 24-51 | 52-56 | > 56 | 63 | 27-62 | 63-69 | > 69 | 68 |
|  | LAEF (%) | 55-75 | 52-54 | < 52 | 63 | 53-72 | 50-52 | < 50 | 68 |
|  | RAVi max (mL/m²) | 21-51 | 52-56 | > 56 | 95 | 27-68 | 69-78 | > 78 | 100 |
|  | RAEF (%) | 39-77 | 33-38 | < 33 | 93 | 35-73 | 31-34 | < 31 | 100 |
| 30-39 | LVEDVi (mL/m²) | 56-103 | 104-112 | > 112 | 155 | 65-120 | 121-128 | > 128 | 150 |
|  | LVESVi (mL/m²) | 15-43 | 44-46 | > 46 | 155 | 21-53 | 54-58 | > 58 | 150 |
|  | LVSVi (mL/m²) | 34-66 | 30-33 | < 30 | 152 | 37-76 | 32-36 | < 32 | 149 |
|  | LVEF (%) | 53-73 | 50-52 | < 50 | 150 | 50-71 | 47-49 | < 47 | 151 |
|  | LVCOi (L/min/m²) | 2.3-4.9 | 2-2.2 | < 2 | 154 | 2.4-5.3 | 1.9-2.3 | < 1.9 | 142 |
|  | LVMi (g/m²) | 33-61 | 62-66 | > 66 | 155 | 43-81 | 82-87 | > 87 | 151 |
|  | LVM/LVEDV (G/ML) | 0.5-0.7 | 0.8-0.8 | > 0.8 | 155 | 0.5-0.9 |  | > 0.9 | 151 |
|  | RVEDVi (mL/m²) | 55-109 | 110-119 | > 119 | 155 | 67-135 | 136-145 | > 145 | 148 |
|  | RVESVi (mL/m²) | 15-50 | 51-54 | > 54 | 153 | 23-63 | 64-68 | > 68 | 145 |
|  | RVEF (%) | 48-72 | 44-47 | < 44 | 145 | 45-68 | 41-44 | < 41 | 148 |
|  | RVCOi (L/min/m²) | 2-5.1 | 1.7-1.9 | < 1.7 | 151 | 2.3-5.5 | 1.9-2.2 | < 1.9 | 141 |
|  | RVSVi (mL/m²) | 32-69 | 27-31 | < 27 | 152 | 36-79 | 30-35 | < 30 | 149 |
|  | LAVi max (mL/m²) | 24-54 | 55-60 | > 60 | 102 | 26-62 | 63-70 | > 70 | 80 |
|  | LAEF (%) | 55-76 | 52-54 | < 52 | 101 | 52-73 | 49-51 | < 49 | 80 |
|  | RAVi max (mL/m²) | 21-51 | 52-57 | > 57 | 131 | 26-69 | 70-78 | > 78 | 135 |
|  | RAEF (%) | 38-74 | 33-37 | < 33 | 128 | 36-71 | 32-35 | < 32 | 134 |
| 40-49 | LVEDVi (mL/m²) | 57-106 | 107-115 | > 115 | 568 | 64-123 | 124-132 | > 132 | 487 |
|  | LVESVi (mL/m²) | 16-43 | 44-47 | > 47 | 568 | 20-53 | 54-58 | > 58 | 483 |
|  | LVSVi (mL/m²) | 35-69 | 31-34 | < 31 | 568 | 38-78 | 32-37 | < 32 | 487 |
|  | LVEF (%) | 53-74 | 50-52 | < 50 | 562 | 51-72 | 48-50 | < 48 | 489 |
|  | LVCOi (L/min/m²) | 2.2-4.9 | 1.9-2.1 | < 1.9 | 564 | 2.3-5.2 | 1.9-2.2 | < 1.9 | 480 |
|  | LVMi (g/m²) | 33-63 | 64-68 | > 68 | 568 | 42-84 | 85-90 | > 90 | 497 |
|  | LVM/LVEDV (G/ML) | 0.5-0.7 | 0.8-0.8 | > 0.8 | 568 | 0.5-0.9 | 01-1 | > 1 | 487 |
|  | RVEDVi (mL/m²) | 55-112 | 113-122 | > 122 | 567 | 67-136 | 137-146 | > 146 | 487 |
|  | RVESVi (mL/m²) | 15-48 | 49-52 | > 52 | 567 | 23-62 | 63-67 | > 67 | 481 |
|  | RVEF (%) | 50-72 | 46-49 | < 46 | 560 | 46-68 | 42-45 | < 42 | 483 |
|  | RVCOi (L/min/m²) | 02-5 | 1.7-1.9 | < 1.7 | 564 | 2.3-5.4 | 1.8-2.2 | < 1.8 | 475 |
|  | RVSVi (mL/m²) | 33-70 | 28-32 | < 28 | 566 | 37-81 | 30-36 | < 30 | 482 |
|  | LAVi max (mL/m²) | 25-59 | 60-65 | > 65 | 487 | 25-63 | 64-71 | > 71 | 409 |
|  | LAEF (%) | 55-77 | 51-54 | < 51 | 481 | 53-76 | 50-52 | < 50 | 404 |
|  | RAVi max (mL/m²) | 23-59 | 60-66 | > 66 | 549 | 28-77 | 78-87 | > 87 | 462 |
|  | RAEF (%) | 38-71 | 33-37 | < 33 | 546 | 35-69 | 31-34 | < 31 | 464 |
| 50-59 | LVEDVi (mL/m²) | 56-106 | 107-116 | > 116 | 1289 | 61-122 | 123-131 | > 131 | 1136 |
|  | LVESVi (mL/m²) | 15-42 | 43-47 | > 47 | 1288 | 18-52 | 53-57 | > 57 | 1141 |
|  | LVSVi (mL/m²) | 35-69 | 31-34 | < 31 | 1287 | 36-76 | 30-35 | < 30 | 1140 |
|  | LVEF (%) | 53-75 | 50-52 | < 50 | 1284 | 51-73 | 47-50 | < 47 | 1152 |
|  | LVCOi (L/min/m²) | 2.1-4.7 | 1.8-2 | < 1.8 | 1281 | 2.2-5 | 1.8-2.1 | < 1.8 | 1138 |
|  | LVMi (g/m²) | 34-64 | 65-71 | > 71 | 1289 | 42-86 | 87-93 | > 93 | 1157 |
|  | LVM/LVEDV (G/ML) | 0.5-0.8 | 0.9-0.9 | > 0.9 | 1289 | 0.6-0.9 | 01-1 | > 1 | 1132 |
|  | RVEDVi (mL/m²) | 54-111 | 112-122 | > 122 | 1287 | 64-131 | 132-142 | > 142 | 1140 |
|  | RVESVi (mL/m²) | 15-46 | 47-50 | > 50 | 1287 | 22-59 | 60-64 | > 64 | 1141 |
|  | RVEF (%) | 52-72 | 49-51 | < 49 | 1284 | 47-69 | 43-46 | < 43 | 1155 |
|  | RVCOi (L/min/m²) | 2-4.8 | 1.7-1.9 | < 1.7 | 1282 | 2.2-5.1 | 1.8-2.1 | < 1.8 | 1140 |
|  | RVSVi (mL/m²) | 33-71 | 29-32 | < 29 | 1286 | 35-79 | 28-34 | < 28 | 1143 |
|  | LAVi max (mL/m²) | 24-61 | 62-68 | > 68 | 1144 | 23-64 | 65-72 | > 72 | 962 |
|  | LAEF (%) | 53-78 | 50-52 | < 50 | 1132 | 53-79 | 50-52 | < 50 | 958 |
|  | RAVi max (mL/m²) | 24-66 | 67-74 | > 74 | 1215 | 26-77 | 78-86 | > 86 | 1072 |
|  | RAEF (%) | 37-69 | 32-36 | < 32 | 1200 | 35-68 | 31-34 | < 31 | 1079 |
| 60-69 | LVEDVi (mL/m²) | 53-101 | 102-111 | > 111 | 1209 | 58-118 | 119-128 | > 128 | 1059 |
|  | LVESVi (mL/m²) | 14-40 | 41-45 | > 45 | 1210 | 16-49 | 50-54 | > 54 | 1052 |
|  | LVSVi (mL/m²) | 33-66 | 29-32 | < 29 | 1209 | 35-75 | 29-34 | < 29 | 1060 |
|  | LVEF (%) | 54-75 | 50-53 | < 50 | 1205 | 50-73 | 46-49 | < 46 | 1057 |
|  | LVCOi (L/min/m²) | 2-4.5 | 1.8-1.9 | < 1.8 | 1208 | 2.1-4.8 | 1.8-2 | < 1.8 | 1055 |
|  | LVMi (g/m²) | 33-65 | 66-72 | > 72 | 1210 | 41-84 | 85-92 | > 92 | 1064 |
|  | LVM/LVEDV (G/ML) | 0.5-0.8 | 0.9-0.9 | > 0.9 | 1209 | 0.6-1 | 1.1-1.1 | > 1.1 | 1056 |
|  | RVEDVi (mL/m²) | 53-107 | 108-118 | > 118 | 1208 | 62-126 | 127-137 | > 137 | 1059 |
|  | RVESVi (mL/m²) | 15-44 | 45-48 | > 48 | 1211 | 21-56 | 57-61 | > 61 | 1063 |
|  | RVEF (%) | 52-73 | 49-51 | < 49 | 1204 | 48-70 | 44-47 | < 44 | 1064 |
|  | RVCOi (L/min/m²) | 2-4.7 | 1.7-1.9 | < 1.7 | 1208 | 2.1-5 | 1.7-2 | < 1.7 | 1060 |
|  | RVSVi (mL/m²) | 32-68 | 28-31 | < 28 | 1208 | 34-77 | 27-33 | < 27 | 1055 |
|  | LAVi max (mL/m²) | 22-62 | 63-70 | > 70 | 1062 | 22-65 | 66-73 | > 73 | 879 |
|  | LAEF (%) | 51-79 | 47-50 | < 47 | 1036 | 51-80 | 47-50 | < 47 | 864 |
|  | RAVi max (mL/m²) | 23-66 | 67-75 | > 75 | 1129 | 25-78 | 79-87 | > 87 | 992 |
|  | RAEF (%) | 37-70 | 32-36 | < 32 | 1115 | 35-70 | 31-34 | < 31 | 1000 |
| 70+ | LVEDVi (mL/m²) | 50-98 | 99-109 | > 109 | 1210 | 54-115 | 116-125 | > 125 | 1346 |
|  | LVESVi (mL/m²) | 13-39 | 40-44 | > 44 | 1209 | 15-48 | 49-53 | > 53 | 1341 |
|  | LVSVi (mL/m²) | 32-64 | 28-31 | < 28 | 1210 | 33-72 | 27-32 | < 27 | 1349 |
|  | LVEF (%) | 54-76 | 50-53 | < 50 | 1205 | 50-74 | 46-49 | < 46 | 1336 |
|  | LVCOi (L/min/m²) | 2-4.5 | 1.7-1.9 | < 1.7 | 1208 | 2.1-4.6 | 1.7-2 | < 1.7 | 1349 |
|  | LVMi (g/m²) | 33-64 | 65-71 | > 71 | 1211 | 40-81 | 82-88 | > 88 | 1352 |
|  | LVM/LVEDV (G/ML) | 0.5-0.9 | 1-1 | > 1 | 1210 | 0.6-1 | 1.1-1.1 | > 1.1 | 1344 |
|  | RVEDVi (mL/m²) | 51-102 | 103-113 | > 113 | 1209 | 59-123 | 124-134 | > 134 | 1353 |
|  | RVESVi (mL/m²) | 15-41 | 42-46 | > 46 | 1209 | 20-54 | 55-59 | > 59 | 1354 |
|  | RVEF (%) | 53-74 | 49-52 | < 49 | 1202 | 48-71 | 45-47 | < 45 | 1340 |
|  | RVCOi (L/min/m²) | 1.9-4.6 | 1.7-1.8 | < 1.7 | 1208 | 2-4.8 | 1.6-1.9 | < 1.6 | 1346 |
|  | RVSVi (mL/m²) | 31-66 | 27-30 | < 27 | 1208 | 32-75 | 26-31 | < 26 | 1350 |
|  | LAVi max (mL/m²) | 21-63 | 64-73 | > 73 | 1041 | 21-66 | 67-75 | > 75 | 1032 |
|  | LAEF (%) | 48-79 | 44-47 | < 44 | 1002 | 48-80 | 44-47 | < 44 | 991 |
|  | RAVi max (mL/m²) | 22-68 | 69-77 | > 77 | 1130 | 23-78 | 79-88 | > 88 | 1204 |
|  | RAEF (%) | 37-72 | 32-36 | < 32 | 1102 | 35-73 | 31-34 | < 31 | 1200 |

Footnote: All measurements were derived using CVI42 software with papillary endocardial segmentation; papillary muscles were included in left ventricular mass measurements. “Reference range,” “Mild,” and “At least moderate” capture the progression of deviation from the reference range for each metric. Reference ranges are defined as the interval between the 2.5th and 97.5th centiles, derived from age-specific generalized add–Green distribution. Centile-based categories beyond the reference range are derived from LMS-based thresholds (P0.5, P2.5, P97.5, and P99.5) and describe the position of a measurement within the reference distribution rather than disease severity or diagnostic abnormality. Reference ranges are based on a White an adult population.

Abbreviations: LV = left ventricle; RV = right ventricle; LA = left atrium; RA = right atrium; EDV = end-diastolic volume; ESV = end-systolic volume; SV = stroke volume; EF = ejection fraction; CO = cardiac output.

### 4. Table: Reference ranges and centile-based categories for cardiac chamber measurements derived using suiteHEART software (smooth segmentation), indexed to height, in an adult population

|  |  | **Female** | | | | **Male** | | | |
| --- | --- | --- | --- | --- | --- | --- | --- | --- | --- |
| **Age group** | **Metrics** | **Reference range** | **Mild** | **At least moderate** | **n** | **Reference range** | **Mild** | **At least moderate** | **n** |
| 18-29 | LVEDVi (mL/m²) | 62-112 | 113-120 | > 120 | 117 | 80-129 | 130-136 | > 136 | 114 |
|  | LVESVi (mL/m²) | 24-50 | 51-56 | > 56 | 117 | 29-57 | 58-61 | > 61 | 102 |
|  | LVSVi (mL/m²) | 35-68 | 30-34 | < 30 | 117 | 43-78 | 38-42 | < 38 | 119 |
|  | LVEF (%) | 50-69 | 48-49 | < 48 | 117 | 47-67 | 44-46 | < 44 | 123 |
|  | LVMi (g/m²) | 31-59 | 60-65 | > 65 | 117 | 45-87 | 88-96 | > 96 | 123 |
|  | LVM/LVEDV (G/ML) | 0.4-0.7 | 0.8-0.8 | > 0.8 | 117 | 0.5-0.8 | 0.9-1 | > 1 | 114 |
|  | RVEDVi (mL/m²) | 60-118 | 119-128 | > 128 | 117 | 82-140 | 141-149 | > 149 | 112 |
|  | RVESVi (mL/m²) | 24-54 | 55-60 | > 60 | 116 | 32-64 | 65-69 | > 69 | 109 |
|  | RVEF (%) | 46-68 | 43-45 | < 43 | 115 | 45-66 | 42-44 | < 42 | 121 |
|  | RVSVi (mL/m²) | 35-72 | 30-34 | < 30 | 116 | 42-81 | 37-41 | < 37 | 116 |
|  | LAESVi (mL/m²) | 19-44 | 45-48 | > 48 | 77 | 22-54 | 55-60 | > 60 | 79 |
|  | LAEF (%) | 56-84 | 51-55 | < 51 | 77 | 54-79 | 51-53 | < 51 | 77 |
|  | RAESVi (mL/m²) | 23-51 | 52-56 | > 56 | 88 | 25-65 | 66-72 | > 72 | 102 |
|  | RAEF (%) | 42-75 | 37-41 | < 37 | 83 | 36-71 | 32-35 | < 32 | 99 |
| 30-39 | LVEDVi (mL/m²) | 62-112 | 113-121 | > 121 | 149 | 73-124 | 125-132 | > 132 | 144 |
|  | LVESVi (mL/m²) | 23-49 | 50-55 | > 55 | 149 | 27-57 | 58-61 | > 61 | 140 |
|  | LVSVi (mL/m²) | 35-67 | 30-34 | < 30 | 148 | 40-75 | 35-39 | < 35 | 145 |
|  | LVEF (%) | 51-69 | 48-50 | < 48 | 149 | 48-68 | 45-47 | < 45 | 146 |
|  | LVMi (g/m²) | 31-62 | 63-69 | > 69 | 149 | 45-88 | 89-97 | > 97 | 145 |
|  | LVM/LVEDV (G/ML) | 0.4-0.7 | 0.8-0.8 | > 0.8 | 149 | 0.5-0.9 | 01-1 | > 1 | 145 |
|  | RVEDVi (mL/m²) | 60-117 | 118-128 | > 128 | 149 | 77-138 | 139-148 | > 148 | 142 |
|  | RVESVi (mL/m²) | 23-52 | 53-59 | > 59 | 149 | 30-64 | 65-69 | > 69 | 143 |
|  | RVEF (%) | 48-69 | 44-47 | < 44 | 149 | 45-67 | 42-44 | < 42 | 145 |
|  | RVSVi (mL/m²) | 34-72 | 30-33 | < 30 | 149 | 40-81 | 34-39 | < 34 | 142 |
|  | LAVi max (mL/m²) | 20-48 | 49-53 | > 53 | 111 | 21-54 | 55-62 | > 62 | 103 |
|  | LAEF (%) | 57-83 | 52-56 | < 52 | 111 | 55-80 | 51-54 | < 51 | 104 |
|  | RAVi max (mL/m²) | 24-54 | 55-60 | > 60 | 129 | 25-67 | 68-75 | > 75 | 121 |
|  | RAEF (%) | 41-75 | 37-40 | < 37 | 122 | 36-71 | 31-35 | < 31 | 120 |
| 40-49 | LVEDVi (mL/m²) | 61-113 | 114-123 | > 123 | 571 | 71-127 | 128-136 | > 136 | 478 |
|  | LVESVi (mL/m²) | 21-48 | 49-55 | > 55 | 571 | 26-56 | 57-62 | > 62 | 478 |
|  | LVSVi (mL/m²) | 35-68 | 31-34 | < 31 | 571 | 40-77 | 34-39 | < 34 | 485 |
|  | LVEF (%) | 51-70 | 48-50 | < 48 | 570 | 49-69 | 46-48 | < 46 | 499 |
|  | LVMi (g/m²) | 32-66 | 67-73 | > 73 | 571 | 44-89 | 90-99 | > 99 | 497 |
|  | LVM/LVEDV (G/ML) | 0.4-0.8 | 0.9-0.9 | > 0.9 | 571 | 0.5-0.9 | 1-1.1 | > 1.1 | 475 |
|  | RVEDVi (mL/m²) | 59-116 | 117-128 | > 128 | 571 | 75-140 | 141-151 | > 151 | 484 |
|  | RVESVi (mL/m²) | 21-50 | 51-57 | > 57 | 571 | 27-63 | 64-69 | > 69 | 478 |
|  | RVEF (%) | 49-71 | 46-48 | < 46 | 568 | 46-69 | 42-45 | < 42 | 498 |
|  | RVSVi (mL/m²) | 34-73 | 29-33 | < 29 | 569 | 39-82 | 33-38 | < 33 | 478 |
|  | LAVi max (mL/m²) | 20-52 | 53-58 | > 58 | 498 | 20-55 | 56-63 | > 63 | 425 |
|  | LAEF (%) | 57-83 | 53-56 | < 53 | 498 | 55-81 | 51-54 | < 51 | 426 |
|  | RAVi max (mL/m²) | 24-57 | 58-64 | > 64 | 498 | 26-70 | 71-78 | > 78 | 414 |
|  | RAEF (%) | 41-74 | 36-40 | < 36 | 489 | 36-72 | 31-35 | < 31 | 411 |
| 50-59 | LVEDVi (mL/m²) | 60-112 | 113-122 | > 122 | 1290 | 67-128 | 129-138 | > 138 | 1125 |
|  | LVESVi (mL/m²) | 20-46 | 47-53 | > 53 | 1289 | 24-55 | 56-61 | > 61 | 1121 |
|  | LVSVi (mL/m²) | 37-71 | 33-36 | < 33 | 1289 | 39-79 | 33-38 | < 33 | 1142 |
|  | LVEF (%) | 53-72 | 49-52 | < 49 | 1290 | 50-71 | 47-49 | < 47 | 1155 |
|  | LVMi (g/m²) | 33-69 | 70-78 | > 78 | 1290 | 44-89 | 90-99 | > 99 | 1146 |
|  | LVM/LVEDV (G/ML) | 0.4-0.8 | 0.9-1 | > 1 | 1290 | 0.5-1 | 1.1-1.1 | > 1.1 | 1112 |
|  | RVEDVi (mL/m²) | 57-115 | 116-127 | > 127 | 1288 | 69-136 | 137-147 | > 147 | 1133 |
|  | RVESVi (mL/m²) | 19-48 | 49-55 | > 55 | 1288 | 25-61 | 62-68 | > 68 | 1123 |
|  | RVEF (%) | 50-73 | 46-49 | < 46 | 1286 | 46-70 | 42-45 | < 42 | 1144 |
|  | RVSVi (mL/m²) | 33-73 | 28-32 | < 28 | 1283 | 37-81 | 30-36 | < 30 | 1140 |
|  | LAVi max (mL/m²) | 20-55 | 56-62 | > 62 | 1171 | 19-56 | 57-65 | > 65 | 986 |
|  | LAEF (%) | 56-82 | 52-55 | < 52 | 1168 | 55-82 | 50-54 | < 50 | 991 |
|  | RAVi max (mL/m²) | 24-59 | 60-67 | > 67 | 1099 | 25-69 | 70-77 | > 77 | 884 |
|  | RAEF (%) | 40-74 | 35-39 | < 35 | 1089 | 37-73 | 32-36 | < 32 | 880 |
| 60-69 | LVEDVi (mL/m²) | 56-107 | 108-118 | > 118 | 1210 | 62-125 | 126-136 | > 136 | 1051 |
|  | LVESVi (mL/m²) | 18-44 | 45-50 | > 50 | 1208 | 21-53 | 54-59 | > 59 | 1045 |
|  | LVSVi (mL/m²) | 35-68 | 31-34 | < 31 | 1210 | 36-77 | 30-35 | < 30 | 1054 |
|  | LVEF (%) | 53-73 | 49-52 | < 49 | 1207 | 50-71 | 47-49 | < 47 | 1059 |
|  | LVMi (g/m²) | 33-70 | 71-80 | > 80 | 1210 | 43-88 | 89-98 | > 98 | 1061 |
|  | LVM/LVEDV (G/ML) | 0.5-0.9 | 1-1.1 | > 1.1 | 1210 | 0.5-1 | 1.1-1.2 | > 1.2 | 1043 |
|  | RVEDVi (mL/m²) | 55-113 | 114-125 | > 125 | 1208 | 65-132 | 133-144 | > 144 | 1052 |
|  | RVESVi (mL/m²) | 18-47 | 48-53 | > 53 | 1209 | 23-60 | 61-67 | > 67 | 1048 |
|  | RVEF (%) | 50-74 | 46-49 | < 46 | 1207 | 46-71 | 42-45 | < 42 | 1056 |
|  | RVSVi (mL/m²) | 32-73 | 27-31 | < 27 | 1206 | 34-80 | 28-33 | < 28 | 1055 |
|  | LAVi max (mL/m²) | 19-56 | 57-65 | > 65 | 1077 | 18-58 | 59-67 | > 67 | 889 |
|  | LAEF (%) | 54-82 | 49-53 | < 49 | 1076 | 53-83 | 48-52 | < 48 | 891 |
|  | RAVi max (mL/m²) | 23-60 | 61-69 | > 69 | 962 | 24-69 | 70-78 | > 78 | 719 |
|  | RAEF (%) | 40-74 | 35-39 | < 35 | 940 | 37-74 | 33-36 | < 33 | 694 |
| 70+ | LVEDVi (mL/m²) | 54-104 | 105-115 | > 115 | 1208 | 59-121 | 122-133 | > 133 | 1334 |
|  | LVESVi (mL/m²) | 16-42 | 43-49 | > 49 | 1207 | 20-53 | 54-60 | > 60 | 1319 |
|  | LVSVi (mL/m²) | 34-66 | 30-33 | < 30 | 1210 | 34-74 | 28-33 | < 28 | 1346 |
|  | LVEF (%) | 53-74 | 49-52 | < 49 | 1207 | 50-71 | 46-49 | < 46 | 1335 |
|  | LVMi (g/m²) | 33-70 | 71-80 | > 80 | 1210 | 42-85 | 86-95 | > 95 | 1342 |
|  | LVM/LVEDV (G/ML) | 0.5-0.9 | 1-1.1 | > 1.1 | 1208 | 0.5-1 | 1.1-1.2 | > 1.2 | 1327 |
|  | RVEDVi (mL/m²) | 53-110 | 111-122 | > 122 | 1208 | 63-129 | 130-141 | > 141 | 1335 |
|  | RVESVi (mL/m²) | 16-45 | 46-52 | > 52 | 1208 | 23-59 | 60-67 | > 67 | 1333 |
|  | RVEF (%) | 50-75 | 45-49 | < 45 | 1204 | 46-72 | 41-45 | < 41 | 1321 |
|  | RVSVi (mL/m²) | 31-72 | 26-30 | < 26 | 1205 | 32-79 | 25-31 | < 25 | 1336 |
|  | LAVi max (mL/m²) | 18-59 | 60-70 | > 70 | 1047 | 17-59 | 60-69 | > 69 | 1034 |
|  | LAEF (%) | 50-83 | 44-49 | < 44 | 1037 | 50-83 | 45-49 | < 45 | 1024 |
|  | RAVi max (mL/m²) | 23-62 | 63-71 | > 71 | 849 | 25-72 | 73-81 | > 81 | 689 |
|  | RAEF (%) | 41-77 | 35-40 | < 35 | 828 | 37-74 | 31-36 | < 31 | 676 |

Footnote: All measurements were derived using suiteHEART software with smooth endocardial segmentation; papillary muscles and trabeculations were excluded from left ventricular mass measurements. “Reference range,” “Mild,” and “At least moderate” capture the progression of deviation from the reference range for each metric. Reference ranges are defined as the interval between the 2.5th and 97.5th centiles, derived from age-specific generalized add–Green distribution. Centile-based categories beyond the reference range are derived from LMS-based thresholds (P0.5, P2.5, P97.5, and P99.5) and describe the position of a measurement within the reference distribution rather than disease severity or diagnostic abnormality. Reference ranges are based on a White an adult population.

Abbreviations: LV = left ventricle; RV = right ventricle; LA = left atrium; RA = right atrium; EDV = end-diastolic volume; ESV = end-systolic volume; SV = stroke volume; EF = ejection fraction; CO = cardiac output.

### 5. Table: Reference ranges and centile-based categories for cardiac chamber measurements derived using suiteHEART software (anatomical segmentation), indexed to height, in an adult population

|  |  | **Female** | | | | **Male** | | | |
| --- | --- | --- | --- | --- | --- | --- | --- | --- | --- |
| **Age group** | **Metrics** | **Reference range** | **Mild** | **At least moderate** | **n** | **Reference range** | **Mild** | **At least moderate** | **n** |
| 18-29 | LVEDVi (mL/m²) | 55-102 | 103-109 | > 109 | 117 | 70-120 | 121-129 | > 129 | 120 |
|  | LVESVi (mL/m²) | 17-41 | 42-45 | > 45 | 117 | 20-49 | 50-55 | > 55 | 122 |
|  | LVSVi (mL/m²) | 34-67 | 29-33 | < 29 | 117 | 42-78 | 37-41 | < 37 | 120 |
|  | LVEF (%) | 55-75 | 52-54 | < 52 | 117 | 52-74 | 49-51 | < 49 | 123 |
|  | LVMi (g/m²) | 37-71 | 72-77 | > 77 | 117 | 53-96 | 97-103 | > 103 | 120 |
|  | LVM/LVEDV (G/ML) | 0.5-0.9 | 1-1.1 | > 1.1 | 117 | 0.6-1.1 |  | > 1.2 | 118 |
|  | RVEDVi (mL/m²) | 59-114 | 115-122 | > 122 | 117 | 78-135 | 136-144 | > 144 | 117 |
|  | RVESVi (mL/m²) | 23-52 | 53-58 | > 58 | 116 | 28-64 | 65-70 | > 70 | 117 |
|  | RVEF (%) | 47-69 | 44-46 | < 44 | 115 | 46-69 | 43-45 | < 43 | 121 |
|  | RVSVi (mL/m²) | 36-71 | 31-35 | < 31 | 116 | 42-81 | 37-41 | < 37 | 120 |
|  | LAESVi (mL/m²) | 19-44 | 45-48 | > 48 | 77 | 22-54 | 55-60 | > 60 | 79 |
|  | LAEF (%) | 56-84 | 51-55 | < 51 | 77 | 54-79 | 51-53 | < 51 | 77 |
|  | RAESVi (mL/m²) | 23-51 | 52-56 | > 56 | 88 | 25-65 | 66-72 | > 72 | 102 |
|  | RAEF (%) | 42-75 | 37-41 | < 37 | 83 | 36-71 | 32-35 | < 32 | 99 |
| 30-39 | LVEDVi (mL/m²) | 53-99 | 100-107 | > 107 | 149 | 63-113 | 114-122 | > 122 | 145 |
|  | LVESVi (mL/m²) | 16-39 | 40-44 | > 44 | 149 | 18-48 | 49-54 | > 54 | 145 |
|  | LVSVi (mL/m²) | 34-66 | 29-33 | < 29 | 148 | 39-75 | 34-38 | < 34 | 145 |
|  | LVEF (%) | 56-76 | 53-55 | < 53 | 149 | 54-75 | 50-53 | < 50 | 146 |
|  | LVMi (g/m²) | 38-74 | 75-81 | > 81 | 149 | 53-97 | 98-104 | > 104 | 142 |
|  | LVM/LVEDV (G/ML) | 0.5-1 | 1.1-1.1 | > 1.1 | 149 | 0.6-1.2 | 1.3-1.3 | > 1.3 | 143 |
|  | RVEDVi (mL/m²) | 58-112 | 113-121 | > 121 | 149 | 74-135 | 136-144 | > 144 | 144 |
|  | RVESVi (mL/m²) | 21-50 | 51-56 | > 56 | 149 | 25-62 | 63-68 | > 68 | 145 |
|  | RVEF (%) | 49-71 | 45-48 | < 45 | 149 | 47-70 | 43-46 | < 43 | 146 |
|  | RVSVi (mL/m²) | 35-71 | 30-34 | < 30 | 149 | 41-82 | 35-40 | < 35 | 143 |
|  | LAVi max (mL/m²) | 20-48 | 49-53 | > 53 | 111 | 21-54 | 55-62 | > 62 | 103 |
|  | LAEF (%) | 57-83 | 52-56 | < 52 | 111 | 55-80 | 51-54 | < 51 | 104 |
|  | RAVi max (mL/m²) | 24-54 | 55-60 | > 60 | 129 | 25-67 | 68-75 | > 75 | 121 |
|  | RAEF (%) | 41-75 | 37-40 | < 37 | 122 | 36-71 | 31-35 | < 31 | 120 |
| 40-49 | LVEDVi (mL/m²) | 53-101 | 102-110 | > 110 | 571 | 62-116 | 117-126 | > 126 | 493 |
|  | LVESVi (mL/m²) | 14-38 | 39-43 | > 43 | 571 | 16-47 | 48-53 | > 53 | 496 |
|  | LVSVi (mL/m²) | 35-68 | 30-34 | < 30 | 571 | 39-77 | 34-38 | < 34 | 488 |
|  | LVEF (%) | 57-78 | 53-56 | < 53 | 570 | 55-77 | 51-54 | < 51 | 498 |
|  | LVMi (g/m²) | 39-77 | 78-86 | > 86 | 571 | 52-97 | 98-105 | > 105 | 467 |
|  | LVM/LVEDV (G/ML) | 0.6-1.1 |  | > 1.2 | 571 | 0.6-1.2 | 1.3-1.4 | > 1.4 | 464 |
|  | RVEDVi (mL/m²) | 56-110 | 111-120 | > 120 | 571 | 70-134 | 135-145 | > 145 | 490 |
|  | RVESVi (mL/m²) | 18-47 | 48-53 | > 53 | 571 | 23-59 | 60-66 | > 66 | 497 |
|  | RVEF (%) | 51-73 | 47-50 | < 47 | 567 | 48-72 | 43-47 | < 43 | 497 |
|  | RVSVi (mL/m²) | 33-71 | 29-32 | < 29 | 569 | 39-83 | 33-38 | < 33 | 483 |
|  | LAVi max (mL/m²) | 20-52 | 53-58 | > 58 | 498 | 20-55 | 56-63 | > 63 | 425 |
|  | LAEF (%) | 57-83 | 53-56 | < 53 | 498 | 55-81 | 51-54 | < 51 | 426 |
|  | RAVi max (mL/m²) | 24-57 | 58-64 | > 64 | 498 | 26-70 | 71-78 | > 78 | 414 |
|  | RAEF (%) | 41-74 | 36-40 | < 36 | 489 | 36-72 | 31-35 | < 31 | 411 |
| 50-59 | LVEDVi (mL/m²) | 53-101 | 102-111 | > 111 | 1290 | 59-117 | 118-128 | > 128 | 1155 |
|  | LVESVi (mL/m²) | 12-36 | 37-42 | > 42 | 1290 | 15-45 | 46-52 | > 52 | 1158 |
|  | LVSVi (mL/m²) | 36-71 | 32-35 | < 32 | 1289 | 39-79 | 33-38 | < 33 | 1147 |
|  | LVEF (%) | 58-80 | 54-57 | < 54 | 1285 | 56-79 | 51-55 | < 51 | 1156 |
|  | LVMi (g/m²) | 40-81 | 82-90 | > 90 | 1289 | 52-98 | 99-106 | > 106 | 1081 |
|  | LVM/LVEDV (G/ML) | 0.6-1.1 | 1.2-1.3 | > 1.3 | 1289 | 0.7-1.3 | 1.4-1.5 | > 1.5 | 1079 |
|  | RVEDVi (mL/m²) | 53-108 | 109-118 | > 118 | 1288 | 63-128 | 129-139 | > 139 | 1149 |
|  | RVESVi (mL/m²) | 16-43 | 44-49 | > 49 | 1288 | 20-56 | 57-63 | > 63 | 1149 |
|  | RVEF (%) | 52-75 | 48-51 | < 48 | 1284 | 48-74 | 43-47 | < 43 | 1154 |
|  | RVSVi (mL/m²) | 32-71 | 28-31 | < 28 | 1285 | 36-80 | 30-35 | < 30 | 1146 |
|  | LAVi max (mL/m²) | 20-55 | 56-62 | > 62 | 1171 | 19-56 | 57-65 | > 65 | 986 |
|  | LAEF (%) | 56-82 | 52-55 | < 52 | 1168 | 55-82 | 50-54 | < 50 | 991 |
|  | RAVi max (mL/m²) | 24-59 | 60-67 | > 67 | 1099 | 25-69 | 70-77 | > 77 | 884 |
|  | RAEF (%) | 40-74 | 35-39 | < 35 | 1089 | 37-73 | 32-36 | < 32 | 880 |
| 60-69 | LVEDVi (mL/m²) | 49-96 | 97-106 | > 106 | 1210 | 54-114 | 115-126 | > 126 | 1066 |
|  | LVESVi (mL/m²) | 11-35 | 36-41 | > 41 | 1209 | 13-44 | 45-51 | > 51 | 1064 |
|  | LVSVi (mL/m²) | 35-68 | 31-34 | < 31 | 1210 | 36-76 | 30-35 | < 30 | 1055 |
|  | LVEF (%) | 59-82 | 54-58 | < 54 | 1203 | 56-80 | 51-55 | < 51 | 1066 |
|  | LVMi (g/m²) | 40-82 | 83-93 | > 93 | 1209 | 50-97 | 98-105 | > 105 | 1016 |
|  | LVM/LVEDV (G/ML) | 0.6-1.2 | 1.3-1.4 | > 1.4 | 1209 | 0.7-1.3 | 1.4-1.6 | > 1.6 | 1014 |
|  | RVEDVi (mL/m²) | 51-105 | 106-116 | > 116 | 1208 | 59-124 | 125-136 | > 136 | 1066 |
|  | RVESVi (mL/m²) | 14-41 | 42-47 | > 47 | 1209 | 18-54 | 55-62 | > 62 | 1067 |
|  | RVEF (%) | 53-77 | 47-52 | < 47 | 1203 | 48-76 | 43-47 | < 43 | 1061 |
|  | RVSVi (mL/m²) | 31-71 | 27-30 | < 27 | 1207 | 34-79 | 27-33 | < 27 | 1057 |
|  | LAVi max (mL/m²) | 19-56 | 57-65 | > 65 | 1077 | 18-58 | 59-67 | > 67 | 889 |
|  | LAEF (%) | 54-82 | 49-53 | < 49 | 1076 | 53-83 | 48-52 | < 48 | 891 |
|  | RAVi max (mL/m²) | 23-60 | 61-69 | > 69 | 962 | 24-69 | 70-78 | > 78 | 719 |
|  | RAEF (%) | 40-74 | 35-39 | < 35 | 940 | 37-74 | 33-36 | < 33 | 694 |
| 70+ | LVEDVi (mL/m²) | 47-93 | 94-104 | > 104 | 1209 | 51-110 | 111-122 | > 122 | 1347 |
|  | LVESVi (mL/m²) | 9-33 | 34-39 | > 39 | 1210 | 12-43 | 44-50 | > 50 | 1343 |
|  | LVSVi (mL/m²) | 33-66 | 30-32 | < 30 | 1210 | 34-73 | 28-33 | < 28 | 1347 |
|  | LVEF (%) | 59-83 | 54-58 | < 54 | 1194 | 56-81 | 50-55 | < 50 | 1336 |
|  | LVMi (g/m²) | 40-81 | 82-93 | > 93 | 1209 | 49-96 | 97-104 | > 104 | 1316 |
|  | LVM/LVEDV (G/ML) | 0.6-1.3 | 1.4-1.5 | > 1.5 | 1208 | 0.7-1.4 | 1.5-1.7 | > 1.7 | 1314 |
|  | RVEDVi (mL/m²) | 48-102 | 103-113 | > 113 | 1208 | 57-122 | 123-134 | > 134 | 1346 |
|  | RVESVi (mL/m²) | 13-39 | 40-46 | > 46 | 1208 | 17-52 | 53-61 | > 61 | 1347 |
|  | RVEF (%) | 52-79 | 46-51 | < 46 | 1194 | 48-77 | 42-47 | < 42 | 1330 |
|  | RVSVi (mL/m²) | 30-71 | 25-29 | < 25 | 1206 | 31-78 | 24-30 | < 24 | 1339 |
|  | LAVi max (mL/m²) | 18-59 | 60-70 | > 70 | 1047 | 17-59 | 60-69 | > 69 | 1034 |
|  | LAEF (%) | 50-83 | 44-49 | < 44 | 1037 | 50-83 | 45-49 | < 45 | 1024 |
|  | RAVi max (mL/m²) | 23-62 | 63-71 | > 71 | 849 | 25-72 | 73-81 | > 81 | 689 |
|  | RAEF (%) | 41-77 | 35-40 | < 35 | 828 | 37-74 | 31-36 | < 31 | 676 |

Footnote: All measurements were derived using suiteHEART software with anatomical endocardial segmentation; papillary muscles and trabeculations were included in left ventricular mass measurements. “Reference range,” “Mild,” and “At least moderate” capture the progression of deviation from the reference range for each metric. Reference ranges are defined as the interval between the 2.5th and 97.5th centiles, derived from age-specific generalized add–Green distribution. Centile-based categories beyond the reference range are derived from LMS-based thresholds (P0.5, P2.5, P97.5, and P99.5) and describe the position of a measurement within the reference distribution rather than disease severity or diagnostic abnormality. Reference ranges are based on a White an adult population.

Abbreviations: LV = left ventricle; RV = right ventricle; LA = left atrium; RA = right atrium; EDV = end-diastolic volume; ESV = end-systolic volume; SV = stroke volume; EF = ejection fraction; CO = cardiac output.

### 6. Table: Reference ranges and centile-based categories for cardiac chamber measurements derived using cvi42 software (smooth segmentation), indexed to bsa, in an adult population

|  |  | **Female** | | | | **Male** | | | |
| --- | --- | --- | --- | --- | --- | --- | --- | --- | --- |
| **Age group** | **Metrics** | **Reference range** | **Mild** | **At least moderate** | **n** | **Reference range** | **Mild** | **At least moderate** | **n** |
| 18-29 | LVEDVi (mL/m²) | 59-103 | 104-110 | > 110 | 118 | 69-117 | 118-124 | > 124 | 118 |
|  | LVESVi (mL/m²) | 16-46 | 47-49 | > 49 | 118 | 22-51 | 52-54 | > 54 | 113 |
|  | LVSVi (mL/m²) | 34-64 | 30-33 | < 30 | 118 | 36-71 | 31-35 | < 31 | 118 |
|  | LVEF (%) | 51-71 | 48-50 | < 48 | 116 | 48-67 | 45-47 | < 45 | 119 |
|  | LVCOi (L/min/m²) | 2.2-4.7 | 2-2.1 | < 2 | 118 | 2.3-4.7 | 1.9-2.2 | < 1.9 | 116 |
|  | LVMi (g/m²) | 33-53 | 54-57 | > 57 | 118 | 41-73 | 74-78 | > 78 | 124 |
|  | LVM/LVEDV (G/ML) | 0.4-0.7 |  | > 0.7 | 118 | 0.5-0.8 |  | > 0.8 | 118 |
|  | RVEDVi (mL/m²) | 59-109 | 110-118 | > 118 | 118 | 71-128 | 129-136 | > 136 | 117 |
|  | RVESVi (mL/m²) | 19-54 | 55-59 | > 59 | 116 | 24-61 | 62-66 | > 66 | 102 |
|  | RVEF (%) | 44-69 | 40-43 | < 40 | 113 | 42-67 | 39-41 | < 39 | 116 |
|  | RVCOi (L/min/m²) | 1.9-5.1 | 1.6-1.8 | < 1.6 | 117 | 2.3-5 | 1.9-2.2 | < 1.9 | 116 |
|  | RVSVi (mL/m²) | 32-67 | 28-31 | < 28 | 118 | 37-74 | 32-36 | < 32 | 117 |
|  | LAVi max (mL/m²) | 24-50 | 51-55 | > 55 | 63 | 26-56 | 57-64 | > 64 | 68 |
|  | LAEF (%) | 55-75 | 52-54 | < 52 | 63 | 53-72 | 50-52 | < 50 | 68 |
|  | RAVi max (mL/m²) | 21-49 | 50-54 | > 54 | 95 | 25-61 | 62-70 | > 70 | 100 |
|  | RAEF (%) | 39-77 | 34-38 | < 34 | 93 | 35-73 | 31-34 | < 31 | 100 |
| 30-39 | LVEDVi (mL/m²) | 57-100 | 101-107 | > 107 | 155 | 61-107 | 108-114 | > 114 | 147 |
|  | LVESVi (mL/m²) | 16-44 | 45-48 | > 48 | 155 | 20-47 | 48-51 | > 51 | 146 |
|  | LVSVi (mL/m²) | 33-62 | 29-32 | < 29 | 152 | 33-67 | 29-32 | < 29 | 148 |
|  | LVEF (%) | 51-71 | 48-50 | < 48 | 150 | 48-69 | 46-47 | < 46 | 151 |
|  | LVCOi (L/min/m²) | 2.2-4.7 | 2-2.1 | < 2 | 153 | 2.2-4.6 | 1.8-2.1 | < 1.8 | 144 |
|  | LVMi (g/m²) | 33-53 | 54-57 | > 57 | 155 | 37-67 | 68-72 | > 72 | 151 |
|  | LVM/LVEDV (G/ML) | 0.4-0.7 |  | > 0.7 | 155 | 0.5-0.8 | 0.9-0.9 | > 0.9 | 148 |
|  | RVEDVi (mL/m²) | 57-106 | 107-115 | > 115 | 155 | 65-121 | 122-129 | > 129 | 147 |
|  | RVESVi (mL/m²) | 18-49 | 50-54 | > 54 | 153 | 23-58 | 59-62 | > 62 | 142 |
|  | RVEF (%) | 47-70 | 43-46 | < 43 | 145 | 44-67 | 41-43 | < 41 | 148 |
|  | RVCOi (L/min/m²) | 2-4.9 | 1.7-1.9 | < 1.7 | 151 | 2.2-4.9 | 1.9-2.1 | < 1.9 | 140 |
|  | RVSVi (mL/m²) | 32-66 | 28-31 | < 28 | 152 | 34-71 | 30-33 | < 30 | 149 |
|  | LAVi max (mL/m²) | 24-51 | 52-56 | > 56 | 102 | 24-55 | 56-63 | > 63 | 80 |
|  | LAEF (%) | 55-76 | 52-54 | < 52 | 101 | 52-73 | 49-51 | < 49 | 80 |
|  | RAVi max (mL/m²) | 20-49 | 50-55 | > 55 | 131 | 23-61 | 62-69 | > 69 | 135 |
|  | RAEF (%) | 38-74 | 33-37 | < 33 | 128 | 36-72 | 32-35 | < 32 | 134 |
| 40-49 | LVEDVi (mL/m²) | 56-99 | 100-107 | > 107 | 568 | 59-110 | 111-118 | > 118 | 483 |
|  | LVESVi (mL/m²) | 16-42 | 43-46 | > 46 | 568 | 19-47 | 48-51 | > 51 | 478 |
|  | LVSVi (mL/m²) | 33-63 | 29-32 | < 29 | 567 | 33-68 | 28-32 | < 28 | 489 |
|  | LVEF (%) | 51-72 | 48-50 | < 48 | 561 | 49-70 | 46-48 | < 46 | 489 |
|  | LVCOi (L/min/m²) | 2.1-4.5 | 1.9-2 | < 1.9 | 565 | 2.1-4.5 | 1.8-2 | < 1.8 | 480 |
|  | LVMi (g/m²) | 32-53 | 54-57 | > 57 | 568 | 38-68 | 69-73 | > 73 | 499 |
|  | LVM/LVEDV (G/ML) | 0.4-0.7 | 0.8-0.8 | > 0.8 | 568 | 0.5-0.8 | 0.9-0.9 | > 0.9 | 483 |
|  | RVEDVi (mL/m²) | 54-103 | 104-111 | > 111 | 567 | 63-121 | 122-130 | > 130 | 485 |
|  | RVESVi (mL/m²) | 16-45 | 46-50 | > 50 | 567 | 21-55 | 56-59 | > 59 | 474 |
|  | RVEF (%) | 50-71 | 47-49 | < 47 | 559 | 46-68 | 42-45 | < 42 | 483 |
|  | RVCOi (L/min/m²) | 2-4.6 | 1.8-1.9 | < 1.8 | 564 | 2.1-4.8 | 1.8-2 | < 1.8 | 471 |
|  | RVSVi (mL/m²) | 32-65 | 28-31 | < 28 | 566 | 34-72 | 29-33 | < 29 | 483 |
|  | LAVi max (mL/m²) | 24-54 | 55-59 | > 59 | 487 | 22-55 | 56-61 | > 61 | 409 |
|  | LAEF (%) | 54-77 | 51-53 | < 51 | 481 | 53-76 | 50-52 | < 50 | 404 |
|  | RAVi max (mL/m²) | 21-56 | 57-62 | > 62 | 549 | 24-68 | 69-77 | > 77 | 462 |
|  | RAEF (%) | 38-71 | 33-37 | < 33 | 546 | 35-69 | 31-34 | < 31 | 464 |
| 50-59 | LVEDVi (mL/m²) | 53-95 | 96-103 | > 103 | 1289 | 55-105 | 106-113 | > 113 | 1130 |
|  | LVESVi (mL/m²) | 16-39 | 40-43 | > 43 | 1287 | 17-46 | 47-50 | > 50 | 1125 |
|  | LVSVi (mL/m²) | 31-61 | 28-30 | < 28 | 1288 | 31-64 | 26-30 | < 26 | 1139 |
|  | LVEF (%) | 51-73 | 48-50 | < 48 | 1283 | 49-71 | 46-48 | < 46 | 1148 |
|  | LVCOi (L/min/m²) | 1.9-4.1 | 1.7-1.8 | < 1.7 | 1282 | 1.9-4.2 | 1.6-1.8 | < 1.6 | 1141 |
|  | LVMi (g/m²) | 32-53 | 54-57 | > 57 | 1289 | 38-68 | 69-72 | > 72 | 1157 |
|  | LVM/LVEDV (G/ML) | 0.5-0.7 | 0.8-0.8 | > 0.8 | 1289 | 0.5-0.9 | 01-1 | > 1 | 1126 |
|  | RVEDVi (mL/m²) | 52-98 | 99-107 | > 107 | 1287 | 57-113 | 114-122 | > 122 | 1137 |
|  | RVESVi (mL/m²) | 15-41 | 42-45 | > 45 | 1287 | 20-51 | 52-56 | > 56 | 1137 |
|  | RVEF (%) | 52-72 | 49-51 | < 49 | 1284 | 47-69 | 44-46 | < 44 | 1154 |
|  | RVCOi (L/min/m²) | 1.9-4.2 | 1.7-1.8 | < 1.7 | 1282 | 1.9-4.4 | 1.6-1.8 | < 1.6 | 1137 |
|  | RVSVi (mL/m²) | 31-63 | 27-30 | < 27 | 1286 | 31-68 | 26-30 | < 26 | 1140 |
|  | LAVi max (mL/m²) | 22-53 | 54-58 | > 58 | 1144 | 21-54 | 55-61 | > 61 | 962 |
|  | LAEF (%) | 53-78 | 50-52 | < 50 | 1132 | 53-79 | 50-52 | < 50 | 958 |
|  | RAVi max (mL/m²) | 21-60 | 61-68 | > 68 | 1215 | 22-67 | 68-75 | > 75 | 1072 |
|  | RAEF (%) | 37-69 | 32-36 | < 32 | 1200 | 35-68 | 31-34 | < 31 | 1079 |
| 60-69 | LVEDVi (mL/m²) | 50-91 | 92-100 | > 100 | 1209 | 51-103 | 104-112 | > 112 | 1056 |
|  | LVESVi (mL/m²) | 14-38 | 39-42 | > 42 | 1210 | 16-45 | 46-49 | > 49 | 1045 |
|  | LVSVi (mL/m²) | 30-58 | 26-29 | < 26 | 1209 | 30-64 | 25-29 | < 25 | 1059 |
|  | LVEF (%) | 52-74 | 48-51 | < 48 | 1203 | 49-72 | 45-48 | < 45 | 1052 |
|  | LVCOi (L/min/m²) | 1.8-3.9 | 1.6-1.7 | < 1.6 | 1208 | 1.9-4.1 | 1.6-1.8 | < 1.6 | 1056 |
|  | LVMi (g/m²) | 31-52 | 53-57 | > 57 | 1210 | 37-66 | 67-71 | > 71 | 1065 |
|  | LVM/LVEDV (G/ML) | 0.5-0.8 | 0.9-0.9 | > 0.9 | 1209 | 0.5-0.9 | 01-1 | > 1 | 1054 |
|  | RVEDVi (mL/m²) | 49-95 | 96-104 | > 104 | 1208 | 54-110 | 111-119 | > 119 | 1058 |
|  | RVESVi (mL/m²) | 14-39 | 40-42 | > 42 | 1211 | 19-48 | 49-53 | > 53 | 1061 |
|  | RVEF (%) | 52-73 | 49-51 | < 49 | 1205 | 48-70 | 44-47 | < 44 | 1064 |
|  | RVCOi (L/min/m²) | 1.9-4.1 | 1.6-1.8 | < 1.6 | 1208 | 1.9-4.3 | 1.6-1.8 | < 1.6 | 1059 |
|  | RVSVi (mL/m²) | 30-61 | 26-29 | < 26 | 1208 | 30-67 | 24-29 | < 24 | 1054 |
|  | LAVi max (mL/m²) | 20-53 | 54-59 | > 59 | 1062 | 19-55 | 56-61 | > 61 | 879 |
|  | LAEF (%) | 51-79 | 47-50 | < 47 | 1036 | 51-80 | 47-50 | < 47 | 864 |
|  | RAVi max (mL/m²) | 21-62 | 63-71 | > 71 | 1129 | 21-69 | 70-77 | > 77 | 992 |
|  | RAEF (%) | 37-70 | 32-36 | < 32 | 1115 | 35-70 | 31-34 | < 31 | 1000 |
| 70+ | LVEDVi (mL/m²) | 48-90 | 91-99 | > 99 | 1209 | 49-102 | 103-112 | > 112 | 1343 |
|  | LVESVi (mL/m²) | 14-37 | 38-42 | > 42 | 1209 | 16-45 | 46-51 | > 51 | 1332 |
|  | LVSVi (mL/m²) | 29-58 | 26-28 | < 26 | 1210 | 29-62 | 24-28 | < 24 | 1347 |
|  | LVEF (%) | 52-75 | 49-51 | < 49 | 1201 | 49-72 | 45-48 | < 45 | 1333 |
|  | LVCOi (L/min/m²) | 1.8-3.9 | 1.6-1.7 | < 1.6 | 1208 | 1.8-4 | 1.5-1.7 | < 1.5 | 1349 |
|  | LVMi (g/m²) | 30-52 | 53-57 | > 57 | 1211 | 36-65 | 66-70 | > 70 | 1355 |
|  | LVM/LVEDV (G/ML) | 0.5-0.8 | 0.9-0.9 | > 0.9 | 1209 | 0.5-0.9 | 1-1.1 | > 1.1 | 1342 |
|  | RVEDVi (mL/m²) | 48-93 | 94-102 | > 102 | 1209 | 52-109 | 110-119 | > 119 | 1353 |
|  | RVESVi (mL/m²) | 13-37 | 38-41 | > 41 | 1209 | 18-48 | 49-53 | > 53 | 1353 |
|  | RVEF (%) | 53-73 | 49-52 | < 49 | 1202 | 48-70 | 44-47 | < 44 | 1341 |
|  | RVCOi (L/min/m²) | 1.9-4.1 | 1.6-1.8 | < 1.6 | 1208 | 1.8-4.2 | 1.5-1.7 | < 1.5 | 1346 |
|  | RVSVi (mL/m²) | 29-60 | 25-28 | < 25 | 1208 | 28-66 | 22-27 | < 22 | 1351 |
|  | LAVi max (mL/m²) | 19-56 | 57-63 | > 63 | 1041 | 18-56 | 57-64 | > 64 | 1032 |
|  | LAEF (%) | 48-79 | 44-47 | < 44 | 1002 | 48-80 | 44-47 | < 44 | 991 |
|  | RAVi max (mL/m²) | 20-65 | 66-75 | > 75 | 1130 | 19-70 | 71-79 | > 79 | 1204 |
|  | RAEF (%) | 37-72 | 32-36 | < 32 | 1102 | 35-73 | 30-34 | < 30 | 1200 |

Footnote: All measurements were derived using civ42 software with smooth endocardial segmentation; papillary muscles and trabeculations were excluded from left ventricular mass measurements. Volumetric and mass measurements were indexed to bsa. “Reference range,” “Mild,” and “At least moderate” capture the progression of deviation from the reference range for each metric. Reference ranges are defined as the interval between the 2.5th and 97.5th centiles, derived from age-specific generalized addreference range are derived from LMS-based thresholds (P0.5, P2.5, P97.5, and P99.5) and describe the position of a measurement within the reference distribution rather than disease severity or diagnostic abnormality. Reference ranges are based on a White an adult population.

Abbreviations: LV = left ventricle; RV = right ventricle; LA = left atrium; RA = right atrium; EDV = end-diastolic volume; ESV = end-systolic volume; SV = stroke volume; EF = ejection fraction; CO = cardiac output.

### 7. Table: Reference ranges and centile-based categories for cardiac chamber measurements derived using cvi42 software (anatomical segmentation), indexed to bsa, in an adult population

|  |  | **Female** | | | | **Male** | | | |
| --- | --- | --- | --- | --- | --- | --- | --- | --- | --- |
| **Age group** | **Metrics** | **Reference range** | **Mild** | **At least moderate** | **n** | **Reference range** | **Mild** | **At least moderate** | **n** |
| 18-29 | LVEDVi (mL/m²) | 53-95 | 96-102 | > 102 | 118 | 62-111 | 112-120 | > 120 | 122 |
|  | LVESVi (mL/m²) | 14-40 | 41-45 | > 45 | 118 | 18-48 | 49-53 | > 53 | 123 |
|  | LVSVi (mL/m²) | 33-63 | 29-32 | < 29 | 118 | 37-72 | 32-36 | < 32 | 120 |
|  | LVEF (%) | 55-75 | 52-54 | < 52 | 116 | 52-72 | 48-51 | < 48 | 121 |
|  | LVCOi (L/min/m²) | 2.2-4.6 | 2-2.1 | < 2 | 118 | 2.3-4.8 | 1.9-2.2 | < 1.9 | 120 |
|  | LVMi (g/m²) | 38-62 | 63-66 | > 66 | 118 | 48-83 | 84-88 | > 88 | 123 |
|  | LVM/LVEDV (G/ML) | 0.6-0.9 | 01-1 | > 1 | 118 | 0.6-1 | 1.1-1.1 | > 1.1 | 121 |
|  | RVEDVi (mL/m²) | 54-101 | 102-108 | > 108 | 118 | 61-124 | 125-133 | > 133 | 121 |
|  | RVESVi (mL/m²) | 17-47 | 48-52 | > 52 | 116 | 22-56 | 57-60 | > 60 | 111 |
|  | RVEF (%) | 47-72 | 44-46 | < 44 | 113 | 44-68 | 41-43 | < 41 | 115 |
|  | RVCOi (L/min/m²) | 1.9-5 | 1.6-1.8 | < 1.6 | 117 | 2.2-4.9 | 1.8-2.1 | < 1.8 | 117 |
|  | RVSVi (mL/m²) | 30-66 | 25-29 | < 25 | 118 | 35-75 | 30-34 | < 30 | 118 |
|  | LAVi max (mL/m²) | 24-50 | 51-55 | > 55 | 63 | 26-56 | 57-64 | > 64 | 68 |
|  | LAEF (%) | 55-75 | 52-54 | < 52 | 63 | 53-72 | 50-52 | < 50 | 68 |
|  | RAVi max (mL/m²) | 21-49 | 50-54 | > 54 | 95 | 25-61 | 62-70 | > 70 | 100 |
|  | RAEF (%) | 39-77 | 34-38 | < 34 | 93 | 35-73 | 31-34 | < 31 | 100 |
| 30-39 | LVEDVi (mL/m²) | 51-92 | 93-98 | > 98 | 155 | 55-102 | 103-110 | > 110 | 151 |
|  | LVESVi (mL/m²) | 13-38 | 39-42 | > 42 | 154 | 15-43 | 44-48 | > 48 | 151 |
|  | LVSVi (mL/m²) | 32-62 | 29-31 | < 29 | 153 | 33-66 | 29-32 | < 29 | 150 |
|  | LVEF (%) | 56-76 | 53-55 | < 53 | 150 | 53-74 | 50-52 | < 50 | 151 |
|  | LVCOi (L/min/m²) | 2.2-4.6 | 2-2.1 | < 2 | 154 | 2.2-4.6 | 1.8-2.1 | < 1.8 | 145 |
|  | LVMi (g/m²) | 38-62 | 63-67 | > 67 | 155 | 44-77 | 78-82 | > 82 | 151 |
|  | LVM/LVEDV (G/ML) | 0.6-0.9 | 01-1 | > 1 | 155 | 0.6-1 | 1.1-1.2 | > 1.2 | 152 |
|  | RVEDVi (mL/m²) | 53-99 | 100-107 | > 107 | 155 | 59-117 | 118-126 | > 126 | 148 |
|  | RVESVi (mL/m²) | 17-46 | 47-50 | > 50 | 153 | 21-54 | 55-58 | > 58 | 146 |
|  | RVEF (%) | 48-71 | 45-47 | < 45 | 145 | 45-68 | 42-44 | < 42 | 148 |
|  | RVCOi (L/min/m²) | 1.9-4.8 | 1.7-1.8 | < 1.7 | 152 | 2.1-4.8 | 1.7-2 | < 1.7 | 142 |
|  | RVSVi (mL/m²) | 30-65 | 26-29 | < 26 | 153 | 32-70 | 27-31 | < 27 | 149 |
|  | LAVi max (mL/m²) | 24-51 | 52-56 | > 56 | 102 | 24-55 | 56-63 | > 63 | 80 |
|  | LAEF (%) | 55-76 | 52-54 | < 52 | 101 | 52-73 | 49-51 | < 49 | 80 |
|  | RAVi max (mL/m²) | 20-49 | 50-55 | > 55 | 131 | 23-61 | 62-69 | > 69 | 135 |
|  | RAEF (%) | 38-74 | 33-37 | < 33 | 128 | 36-72 | 32-35 | < 32 | 134 |
| 40-49 | LVEDVi (mL/m²) | 50-91 | 92-98 | > 98 | 568 | 53-103 | 104-111 | > 111 | 494 |
|  | LVESVi (mL/m²) | 13-35 | 36-40 | > 40 | 568 | 14-42 | 43-47 | > 47 | 494 |
|  | LVSVi (mL/m²) | 33-62 | 29-32 | < 29 | 568 | 33-67 | 28-32 | < 28 | 490 |
|  | LVEF (%) | 56-77 | 53-55 | < 53 | 562 | 54-76 | 51-53 | < 51 | 489 |
|  | LVCOi (L/min/m²) | 2.1-4.4 | 1.9-2 | < 1.9 | 565 | 2.1-4.4 | 1.8-2 | < 1.8 | 484 |
|  | LVMi (g/m²) | 37-62 | 63-67 | > 67 | 568 | 44-78 | 79-83 | > 83 | 484 |
|  | LVM/LVEDV (G/ML) | 0.6-0.9 | 01-1 | > 1 | 568 | 0.6-1.1 |  | > 1.2 | 481 |
|  | RVEDVi (mL/m²) | 53-100 | 101-109 | > 109 | 567 | 60-119 | 120-128 | > 128 | 485 |
|  | RVESVi (mL/m²) | 16-44 | 45-49 | > 49 | 567 | 21-54 | 55-58 | > 58 | 478 |
|  | RVEF (%) | 50-71 | 46-49 | < 46 | 558 | 46-68 | 43-45 | < 43 | 486 |
|  | RVCOi (L/min/m²) | 2-4.5 | 1.7-1.9 | < 1.7 | 564 | 2.1-4.7 | 1.7-2 | < 1.7 | 480 |
|  | RVSVi (mL/m²) | 31-63 | 27-30 | < 27 | 566 | 32-71 | 27-31 | < 27 | 486 |
|  | LAVi max (mL/m²) | 24-54 | 55-59 | > 59 | 487 | 22-55 | 56-61 | > 61 | 409 |
|  | LAEF (%) | 54-77 | 51-53 | < 51 | 481 | 53-76 | 50-52 | < 50 | 404 |
|  | RAVi max (mL/m²) | 21-56 | 57-62 | > 62 | 549 | 24-68 | 69-77 | > 77 | 462 |
|  | RAEF (%) | 38-71 | 33-37 | < 33 | 546 | 35-69 | 31-34 | < 31 | 464 |
| 50-59 | LVEDVi (mL/m²) | 47-87 | 88-94 | > 94 | 1289 | 49-98 | 99-106 | > 106 | 1157 |
|  | LVESVi (mL/m²) | 11-32 | 33-36 | > 36 | 1289 | 12-39 | 40-44 | > 44 | 1161 |
|  | LVSVi (mL/m²) | 31-60 | 28-30 | < 28 | 1288 | 31-64 | 26-30 | < 26 | 1145 |
|  | LVEF (%) | 57-78 | 53-56 | < 53 | 1283 | 54-77 | 51-53 | < 51 | 1155 |
|  | LVCOi (L/min/m²) | 1.9-4 | 1.7-1.8 | < 1.7 | 1284 | 2-4.2 | 1.7-1.9 | < 1.7 | 1141 |
|  | LVMi (g/m²) | 37-62 | 63-67 | > 67 | 1288 | 43-77 | 78-82 | > 82 | 1131 |
|  | LVM/LVEDV (G/ML) | 0.6-1 | 1.1-1.1 | > 1.1 | 1288 | 0.7-1.1 | 1.2-1.3 | > 1.3 | 1127 |
|  | RVEDVi (mL/m²) | 52-98 | 99-107 | > 107 | 1287 | 56-113 | 114-122 | > 122 | 1142 |
|  | RVESVi (mL/m²) | 15-42 | 43-46 | > 46 | 1287 | 20-52 | 53-56 | > 56 | 1131 |
|  | RVEF (%) | 50-71 | 47-49 | < 47 | 1284 | 46-68 | 43-45 | < 43 | 1153 |
|  | RVCOi (L/min/m²) | 1.9-4.1 | 1.7-1.8 | < 1.7 | 1282 | 1.9-4.3 | 1.5-1.8 | < 1.5 | 1145 |
|  | RVSVi (mL/m²) | 30-61 | 26-29 | < 26 | 1287 | 30-67 | 25-29 | < 25 | 1153 |
|  | LAVi max (mL/m²) | 22-53 | 54-58 | > 58 | 1144 | 21-54 | 55-61 | > 61 | 962 |
|  | LAEF (%) | 53-78 | 50-52 | < 50 | 1132 | 53-79 | 50-52 | < 50 | 958 |
|  | RAVi max (mL/m²) | 21-60 | 61-68 | > 68 | 1215 | 22-67 | 68-75 | > 75 | 1072 |
|  | RAEF (%) | 37-69 | 32-36 | < 32 | 1200 | 35-68 | 31-34 | < 31 | 1079 |
| 60-69 | LVEDVi (mL/m²) | 44-83 | 84-90 | > 90 | 1209 | 45-95 | 96-104 | > 104 | 1065 |
|  | LVESVi (mL/m²) | 10-30 | 31-34 | > 34 | 1211 | 11-37 | 38-41 | > 41 | 1063 |
|  | LVSVi (mL/m²) | 30-57 | 27-29 | < 27 | 1209 | 30-63 | 25-29 | < 25 | 1061 |
|  | LVEF (%) | 58-79 | 54-57 | < 54 | 1208 | 54-78 | 49-53 | < 49 | 1064 |
|  | LVCOi (L/min/m²) | 1.9-3.9 | 1.6-1.8 | < 1.6 | 1209 | 1.9-4 | 1.6-1.8 | < 1.6 | 1059 |
|  | LVMi (g/m²) | 36-62 | 63-67 | > 67 | 1210 | 43-76 | 77-81 | > 81 | 1052 |
|  | LVM/LVEDV (G/ML) | 0.6-1 | 1.1-1.2 | > 1.2 | 1209 | 0.7-1.2 | 1.3-1.4 | > 1.4 | 1049 |
|  | RVEDVi (mL/m²) | 49-94 | 95-103 | > 103 | 1208 | 53-110 | 111-120 | > 120 | 1059 |
|  | RVESVi (mL/m²) | 14-40 | 41-44 | > 44 | 1211 | 19-49 | 50-54 | > 54 | 1057 |
|  | RVEF (%) | 51-72 | 48-50 | < 48 | 1203 | 47-69 | 43-46 | < 43 | 1061 |
|  | RVCOi (L/min/m²) | 1.8-4 | 1.6-1.7 | < 1.6 | 1208 | 1.8-4.2 | 1.5-1.7 | < 1.5 | 1061 |
|  | RVSVi (mL/m²) | 29-59 | 26-28 | < 26 | 1209 | 29-66 | 24-28 | < 24 | 1060 |
|  | LAVi max (mL/m²) | 20-53 | 54-59 | > 59 | 1062 | 19-55 | 56-61 | > 61 | 879 |
|  | LAEF (%) | 51-79 | 47-50 | < 47 | 1036 | 51-80 | 47-50 | < 47 | 864 |
|  | RAVi max (mL/m²) | 21-62 | 63-71 | > 71 | 1129 | 21-69 | 70-77 | > 77 | 992 |
|  | RAEF (%) | 37-70 | 32-36 | < 32 | 1115 | 35-70 | 31-34 | < 31 | 1000 |
| 70+ | LVEDVi (mL/m²) | 48-90 | 91-99 | > 99 | 1209 | 49-102 | 103-112 | > 112 | 1343 |
|  | LVESVi (mL/m²) | 14-37 | 38-42 | > 42 | 1209 | 16-45 | 46-51 | > 51 | 1332 |
|  | LVSVi (mL/m²) | 29-58 | 26-28 | < 26 | 1210 | 29-62 | 24-28 | < 24 | 1347 |
|  | LVEF (%) | 52-75 | 49-51 | < 49 | 1201 | 49-72 | 45-48 | < 45 | 1333 |
|  | LVCOi (L/min/m²) | 1.8-3.9 | 1.6-1.7 | < 1.6 | 1208 | 1.8-4 | 1.5-1.7 | < 1.5 | 1349 |
|  | LVMi (g/m²) | 30-52 | 53-57 | > 57 | 1211 | 36-65 | 66-70 | > 70 | 1355 |
|  | LVM/LVEDV (G/ML) | 0.5-0.8 | 0.9-0.9 | > 0.9 | 1209 | 0.5-0.9 | 1-1.1 | > 1.1 | 1342 |
|  | RVEDVi (mL/m²) | 48-93 | 94-102 | > 102 | 1209 | 52-109 | 110-119 | > 119 | 1353 |
|  | RVESVi (mL/m²) | 13-37 | 38-41 | > 41 | 1209 | 18-48 | 49-53 | > 53 | 1353 |
|  | RVEF (%) | 53-73 | 49-52 | < 49 | 1202 | 48-70 | 44-47 | < 44 | 1341 |
|  | RVCOi (L/min/m²) | 1.9-4.1 | 1.6-1.8 | < 1.6 | 1208 | 1.8-4.2 | 1.5-1.7 | < 1.5 | 1346 |
|  | RVSVi (mL/m²) | 29-60 | 25-28 | < 25 | 1208 | 28-66 | 22-27 | < 22 | 1351 |
|  | LAVi max (mL/m²) | 19-56 | 57-63 | > 63 | 1041 | 18-56 | 57-64 | > 64 | 1032 |
|  | LAEF (%) | 48-79 | 44-47 | < 44 | 1002 | 48-80 | 44-47 | < 44 | 991 |
|  | RAVi max (mL/m²) | 20-65 | 66-75 | > 75 | 1130 | 19-70 | 71-79 | > 79 | 1204 |
|  | RAEF (%) | 37-72 | 32-36 | < 32 | 1102 | 35-73 | 30-34 | < 30 | 1200 |

Footnote: All measurements were derived using cvi42 software with anatomical endocardial segmentation; papillary muscles and trabeculations were included in left ventricular mass measurements. Volumetric and mass measurements were indexed to bsa. “Reference range,” “Mild,” and “At least moderate” capture the progression of deviation from the reference range for each metric. Reference ranges are defined as the interval between the 2.5th and 97.5th centiles, derived from age-specific generalized addreference range are derived from LMS-based thresholds (P0.5, P2.5, P97.5, and P99.5) and describe the position of a measurement within the reference distribution rather than disease severity or diagnostic abnormality. Reference ranges are based on a White an adult population.

Abbreviations: LV = left ventricle; RV = right ventricle; LA = left atrium; RA = right atrium; EDV = end-diastolic volume; ESV = end-systolic volume; SV = stroke volume; EF = ejection fraction; CO = cardiac output.

### 8. Table: Reference ranges and centile-based categories for cardiac chamber measurements derived using cvi42 software (smooth segmentation with papillary muscles added to the LV mass segmentation), indexed to bsa, in an adult population

|  |  | **Female** | | | | **Male** | | | |
| --- | --- | --- | --- | --- | --- | --- | --- | --- | --- |
| **Age group** | **Metrics** | **Reference range** | **Mild** | **At least moderate** | **n** | **Reference range** | **Mild** | **At least moderate** | **n** |
| 18-29 | LVEDVi (mL/m²) | 57-101 | 102-108 | > 108 | 118 | 67-115 | 116-122 | > 122 | 119 |
|  | LVESVi (mL/m²) | 14-43 | 44-46 | > 46 | 118 | 20-49 | 50-52 | > 52 | 119 |
|  | LVSVi (mL/m²) | 34-64 | 30-33 | < 30 | 118 | 37-72 | 32-36 | < 32 | 118 |
|  | LVEF (%) | 52-73 | 50-51 | < 50 | 116 | 49-69 | 45-48 | < 45 | 121 |
|  | LVCOi (L/min/m²) | 2.2-4.6 | 2-2.1 | < 2 | 117 | 2.3-4.8 | 1.9-2.2 | < 1.9 | 116 |
|  | LVMi (g/m²) | 34-55 | 56-59 | > 59 | 118 | 43-76 | 77-82 | > 82 | 124 |
|  | LVM/LVEDV (G/ML) | 0.5-0.7 | 0.8-0.8 | > 0.8 | 118 | 0.5-0.8 | 0.9-0.9 | > 0.9 | 119 |
|  | RVEDVi (mL/m²) | 56-104 | 105-112 | > 112 | 118 | 66-126 | 127-135 | > 135 | 120 |
|  | RVESVi (mL/m²) | 14-49 | 50-54 | > 54 | 116 | 24-60 | 61-65 | > 65 | 111 |
|  | RVEF (%) | 46-70 | 43-45 | < 43 | 113 | 44-68 | 40-43 | < 40 | 117 |
|  | RVCOi (L/min/m²) | 1.9-4.8 | 1.6-1.8 | < 1.6 | 117 | 2.2-5 | 1.9-2.1 | < 1.9 | 116 |
|  | RVSVi (mL/m²) | 31-67 | 26-30 | < 26 | 118 | 36-75 | 31-35 | < 31 | 117 |
|  | LAVi max (mL/m²) | 24-50 | 51-55 | > 55 | 63 | 26-56 | 57-64 | > 64 | 68 |
|  | LAEF (%) | 55-75 | 52-54 | < 52 | 63 | 53-72 | 50-52 | < 50 | 68 |
|  | RAVi max (mL/m²) | 21-49 | 50-54 | > 54 | 95 | 25-61 | 62-70 | > 70 | 100 |
|  | RAEF (%) | 39-77 | 34-38 | < 34 | 93 | 35-73 | 31-34 | < 31 | 100 |
| 30-39 | LVEDVi (mL/m²) | 55-97 | 98-105 | > 105 | 155 | 59-106 | 107-113 | > 113 | 150 |
|  | LVESVi (mL/m²) | 15-41 | 42-45 | > 45 | 155 | 17-45 | 46-48 | > 48 | 150 |
|  | LVSVi (mL/m²) | 33-63 | 29-32 | < 29 | 152 | 34-67 | 29-33 | < 29 | 149 |
|  | LVEF (%) | 53-73 | 50-52 | < 50 | 150 | 50-71 | 47-49 | < 47 | 151 |
|  | LVCOi (L/min/m²) | 2.3-4.8 | 2.1-2.2 | < 2.1 | 154 | 2.2-4.7 | 1.9-2.1 | < 1.9 | 142 |
|  | LVMi (g/m²) | 34-55 | 56-59 | > 59 | 155 | 40-71 | 72-76 | > 76 | 151 |
|  | LVM/LVEDV (G/ML) | 0.5-0.7 | 0.8-0.8 | > 0.8 | 155 | 0.5-0.9 |  | > 0.9 | 151 |
|  | RVEDVi (mL/m²) | 55-102 | 103-111 | > 111 | 155 | 62-119 | 120-128 | > 128 | 148 |
|  | RVESVi (mL/m²) | 15-46 | 47-51 | > 51 | 153 | 22-57 | 58-63 | > 63 | 145 |
|  | RVEF (%) | 47-72 | 44-46 | < 44 | 145 | 45-68 | 42-44 | < 42 | 148 |
|  | RVCOi (L/min/m²) | 2-4.9 | 1.7-1.9 | < 1.7 | 151 | 2.2-4.9 | 1.8-2.1 | < 1.8 | 141 |
|  | RVSVi (mL/m²) | 31-65 | 27-30 | < 27 | 152 | 34-71 | 29-33 | < 29 | 149 |
|  | LAVi max (mL/m²) | 24-51 | 52-56 | > 56 | 102 | 24-55 | 56-63 | > 63 | 80 |
|  | LAEF (%) | 55-76 | 52-54 | < 52 | 101 | 52-73 | 49-51 | < 49 | 80 |
|  | RAVi max (mL/m²) | 20-49 | 50-55 | > 55 | 131 | 23-61 | 62-69 | > 69 | 135 |
|  | RAEF (%) | 38-74 | 33-37 | < 33 | 128 | 36-72 | 32-35 | < 32 | 134 |
| 40-49 | LVEDVi (mL/m²) | 54-97 | 98-105 | > 105 | 568 | 58-108 | 109-116 | > 116 | 487 |
|  | LVESVi (mL/m²) | 15-39 | 40-43 | > 43 | 568 | 17-45 | 46-49 | > 49 | 483 |
|  | LVSVi (mL/m²) | 33-63 | 29-32 | < 29 | 568 | 34-69 | 29-33 | < 29 | 487 |
|  | LVEF (%) | 53-74 | 50-52 | < 50 | 562 | 51-72 | 48-50 | < 48 | 489 |
|  | LVCOi (L/min/m²) | 2.2-4.6 | 2-2.1 | < 2 | 564 | 2.1-4.5 | 1.8-2 | < 1.8 | 480 |
|  | LVMi (g/m²) | 33-55 | 56-59 | > 59 | 568 | 40-71 | 72-76 | > 76 | 497 |
|  | LVM/LVEDV (G/ML) | 0.5-0.7 | 0.8-0.8 | > 0.8 | 568 | 0.5-0.9 | 01-1 | > 1 | 487 |
|  | RVEDVi (mL/m²) | 53-101 | 102-109 | > 109 | 567 | 61-120 | 121-129 | > 129 | 487 |
|  | RVESVi (mL/m²) | 15-44 | 45-48 | > 48 | 567 | 21-55 | 56-60 | > 60 | 481 |
|  | RVEF (%) | 50-72 | 46-49 | < 46 | 560 | 46-68 | 43-45 | < 43 | 483 |
|  | RVCOi (L/min/m²) | 2-4.6 | 1.8-1.9 | < 1.8 | 564 | 2.1-4.8 | 1.8-2 | < 1.8 | 475 |
|  | RVSVi (mL/m²) | 31-64 | 27-30 | < 27 | 566 | 33-71 | 28-32 | < 28 | 482 |
|  | LAVi max (mL/m²) | 24-54 | 55-59 | > 59 | 487 | 22-55 | 56-61 | > 61 | 409 |
|  | LAEF (%) | 54-77 | 51-53 | < 51 | 481 | 53-76 | 50-52 | < 50 | 404 |
|  | RAVi max (mL/m²) | 21-56 | 57-62 | > 62 | 549 | 24-68 | 69-77 | > 77 | 462 |
|  | RAEF (%) | 38-71 | 33-37 | < 33 | 546 | 35-69 | 31-34 | < 31 | 464 |
| 50-59 | LVEDVi (mL/m²) | 52-93 | 94-101 | > 101 | 1289 | 53-103 | 104-111 | > 111 | 1136 |
|  | LVESVi (mL/m²) | 14-37 | 38-41 | > 41 | 1288 | 16-43 | 44-47 | > 47 | 1141 |
|  | LVSVi (mL/m²) | 32-61 | 28-31 | < 28 | 1287 | 31-65 | 27-30 | < 27 | 1140 |
|  | LVEF (%) | 53-74 | 50-52 | < 50 | 1284 | 51-73 | 47-50 | < 47 | 1152 |
|  | LVCOi (L/min/m²) | 1.9-4.1 | 1.7-1.8 | < 1.7 | 1281 | 2-4.2 | 1.7-1.9 | < 1.7 | 1138 |
|  | LVMi (g/m²) | 33-55 | 56-59 | > 59 | 1289 | 39-70 | 71-75 | > 75 | 1157 |
|  | LVM/LVEDV (G/ML) | 0.5-0.8 | 0.9-0.9 | > 0.9 | 1289 | 0.6-0.9 | 01-1 | > 1 | 1132 |
|  | RVEDVi (mL/m²) | 51-97 | 98-106 | > 106 | 1287 | 56-112 | 113-121 | > 121 | 1140 |
|  | RVESVi (mL/m²) | 15-41 | 42-45 | > 45 | 1287 | 19-51 | 52-55 | > 55 | 1141 |
|  | RVEF (%) | 52-72 | 49-51 | < 49 | 1284 | 47-69 | 44-46 | < 44 | 1155 |
|  | RVCOi (L/min/m²) | 1.9-4.2 | 1.7-1.8 | < 1.7 | 1282 | 1.9-4.3 | 1.6-1.8 | < 1.6 | 1140 |
|  | RVSVi (mL/m²) | 31-62 | 27-30 | < 27 | 1286 | 31-68 | 25-30 | < 25 | 1143 |
|  | LAVi max (mL/m²) | 22-53 | 54-58 | > 58 | 1144 | 21-54 | 55-61 | > 61 | 962 |
|  | LAEF (%) | 53-78 | 50-52 | < 50 | 1132 | 53-79 | 50-52 | < 50 | 958 |
|  | RAVi max (mL/m²) | 21-60 | 61-68 | > 68 | 1215 | 22-67 | 68-75 | > 75 | 1072 |
|  | RAEF (%) | 37-69 | 32-36 | < 32 | 1200 | 35-68 | 31-34 | < 31 | 1079 |
| 60-69 | LVEDVi (mL/m²) | 49-89 | 90-97 | > 97 | 1209 | 50-101 | 102-110 | > 110 | 1059 |
|  | LVESVi (mL/m²) | 13-36 | 37-40 | > 40 | 1210 | 14-42 | 43-47 | > 47 | 1052 |
|  | LVSVi (mL/m²) | 30-59 | 27-29 | < 27 | 1209 | 30-64 | 25-29 | < 25 | 1060 |
|  | LVEF (%) | 54-75 | 50-53 | < 50 | 1205 | 50-73 | 46-49 | < 46 | 1057 |
|  | LVCOi (L/min/m²) | 1.9-4 | 1.7-1.8 | < 1.7 | 1208 | 1.9-4.1 | 1.6-1.8 | < 1.6 | 1055 |
|  | LVMi (g/m²) | 32-55 | 56-59 | > 59 | 1210 | 38-69 | 70-74 | > 74 | 1064 |
|  | LVM/LVEDV (G/ML) | 0.5-0.8 | 0.9-0.9 | > 0.9 | 1209 | 0.6-1 | 1.1-1.1 | > 1.1 | 1056 |
|  | RVEDVi (mL/m²) | 49-94 | 95-102 | > 102 | 1208 | 53-109 | 110-118 | > 118 | 1059 |
|  | RVESVi (mL/m²) | 14-38 | 39-42 | > 42 | 1211 | 18-48 | 49-52 | > 52 | 1063 |
|  | RVEF (%) | 52-73 | 49-51 | < 49 | 1204 | 48-70 | 44-47 | < 44 | 1064 |
|  | RVCOi (L/min/m²) | 1.8-4.1 | 1.6-1.7 | < 1.6 | 1208 | 1.9-4.3 | 1.5-1.8 | < 1.5 | 1060 |
|  | RVSVi (mL/m²) | 30-60 | 26-29 | < 26 | 1208 | 29-66 | 24-28 | < 24 | 1055 |
|  | LAVi max (mL/m²) | 20-53 | 54-59 | > 59 | 1062 | 19-55 | 56-61 | > 61 | 879 |
|  | LAEF (%) | 51-79 | 47-50 | < 47 | 1036 | 51-80 | 47-50 | < 47 | 864 |
|  | RAVi max (mL/m²) | 21-62 | 63-71 | > 71 | 1129 | 21-69 | 70-77 | > 77 | 992 |
|  | RAEF (%) | 37-70 | 32-36 | < 32 | 1115 | 35-70 | 31-34 | < 31 | 1000 |
| 70+ | LVEDVi (mL/m²) | 48-90 | 91-99 | > 99 | 1209 | 49-102 | 103-112 | > 112 | 1343 |
|  | LVESVi (mL/m²) | 14-37 | 38-42 | > 42 | 1209 | 16-45 | 46-51 | > 51 | 1332 |
|  | LVSVi (mL/m²) | 29-58 | 26-28 | < 26 | 1210 | 29-62 | 24-28 | < 24 | 1347 |
|  | LVEF (%) | 52-75 | 49-51 | < 49 | 1201 | 49-72 | 45-48 | < 45 | 1333 |
|  | LVCOi (L/min/m²) | 1.8-3.9 | 1.6-1.7 | < 1.6 | 1208 | 1.8-4 | 1.5-1.7 | < 1.5 | 1349 |
|  | LVMi (g/m²) | 30-52 | 53-57 | > 57 | 1211 | 36-65 | 66-70 | > 70 | 1355 |
|  | LVM/LVEDV (G/ML) | 0.5-0.8 | 0.9-0.9 | > 0.9 | 1209 | 0.5-0.9 | 1-1.1 | > 1.1 | 1342 |
|  | RVEDVi (mL/m²) | 48-93 | 94-102 | > 102 | 1209 | 52-109 | 110-119 | > 119 | 1353 |
|  | RVESVi (mL/m²) | 13-37 | 38-41 | > 41 | 1209 | 18-48 | 49-53 | > 53 | 1353 |
|  | RVEF (%) | 53-73 | 49-52 | < 49 | 1202 | 48-70 | 44-47 | < 44 | 1341 |
|  | RVCOi (L/min/m²) | 1.9-4.1 | 1.6-1.8 | < 1.6 | 1208 | 1.8-4.2 | 1.5-1.7 | < 1.5 | 1346 |
|  | RVSVi (mL/m²) | 29-60 | 25-28 | < 25 | 1208 | 28-66 | 22-27 | < 22 | 1351 |
|  | LAVi max (mL/m²) | 19-56 | 57-63 | > 63 | 1041 | 18-56 | 57-64 | > 64 | 1032 |
|  | LAEF (%) | 48-79 | 44-47 | < 44 | 1002 | 48-80 | 44-47 | < 44 | 991 |
|  | RAVi max (mL/m²) | 20-65 | 66-75 | > 75 | 1130 | 19-70 | 71-79 | > 79 | 1204 |
|  | RAEF (%) | 37-72 | 32-36 | < 32 | 1102 | 35-73 | 30-34 | < 30 | 1200 |

Footnote: All measurements were derived using cvi42 software with papillary endocardial segmentation; papillary muscles were included in left ventricular mass measurements. Volumetric and mass measurements were indexed to bsa. “Reference range,” “Mild,” and “At least moderate” capture the progression of deviation from the reference range for each metric. Reference ranges are defined as the interval between the 2.5th and 97.5th centiles, derived from age-specific generalized addreference range are derived from LMS-based thresholds (P0.5, P2.5, P97.5, and P99.5) and describe the position of a measurement within the reference distribution rather than disease severity or diagnostic abnormality. Reference ranges are based on a White an adult population.

Abbreviations: LV = left ventricle; RV = right ventricle; LA = left atrium; RA = right

### 9. Table: Reference ranges and centile-based categories for cardiac chamber measurements derived using suiteHEART software (smooth segmentation), indexed to bsa, in an adult population

|  |  | **Female** | | | | **Male** | | | |
| --- | --- | --- | --- | --- | --- | --- | --- | --- | --- |
| **Age group** | **Metrics** | **Reference range** | **Mild** | **At least moderate** | **n** | **Reference range** | **Mild** | **At least moderate** | **n** |
| 18-29 | LVEDVi (mL/m²) | 63-108 | 109-116 | > 116 | 117 | 73-121 | 122-128 | > 128 | 114 |
|  | LVESVi (mL/m²) | 23-48 | 49-53 | > 53 | 117 | 27-52 | 53-56 | > 56 | 102 |
|  | LVSVi (mL/m²) | 36-65 | 31-35 | < 31 | 117 | 40-73 | 36-39 | < 36 | 119 |
|  | LVEF (%) | 50-69 | 47-49 | < 47 | 117 | 47-67 | 45-46 | < 45 | 123 |
|  | LVMi (g/m²) | 32-55 | 56-60 | > 60 | 117 | 43-77 | 78-85 | > 85 | 123 |
|  | LVM/LVEDV (G/ML) | 0.4-0.7 | 0.8-0.8 | > 0.8 | 117 | 0.5-0.8 | 0.9-0.9 | > 0.9 | 114 |
|  | RVEDVi (mL/m²) | 61-118 | 119-129 | > 129 | 117 | 76-133 | 134-143 | > 143 | 112 |
|  | RVESVi (mL/m²) | 25-54 | 55-61 | > 61 | 116 | 30-59 | 60-64 | > 64 | 109 |
|  | RVEF (%) | 46-68 | 43-45 | < 43 | 115 | 45-66 | 42-44 | < 42 | 121 |
|  | RVSVi (mL/m²) | 33-71 | 30-32 | < 30 | 116 | 40-76 | 35-39 | < 35 | 116 |
|  | LAESVi (mL/m²) | 18-43 | 44-48 | > 48 | 77 | 19-49 | 50-56 | > 56 | 79 |
|  | LAEF (%) | 56-84 | 51-55 | < 51 | 77 | 54-79 | 51-53 | < 51 | 77 |
|  | RAESVi (mL/m²) | 23-48 | 49-53 | > 53 | 88 | 23-59 | 60-66 | > 66 | 102 |
|  | RAEF (%) | 42-75 | 38-41 | < 38 | 83 | 36-71 | 31-35 | < 31 | 99 |
| 30-39 | LVEDVi (mL/m²) | 60-106 | 107-114 | > 114 | 149 | 65-111 | 112-119 | > 119 | 144 |
|  | LVESVi (mL/m²) | 22-47 | 48-52 | > 52 | 149 | 25-50 | 51-54 | > 54 | 140 |
|  | LVSVi (mL/m²) | 35-63 | 30-34 | < 30 | 148 | 36-67 | 32-35 | < 32 | 145 |
|  | LVEF (%) | 50-69 | 47-49 | < 47 | 149 | 48-68 | 45-47 | < 45 | 146 |
|  | LVMi (g/m²) | 32-56 | 57-61 | > 61 | 149 | 42-76 | 77-83 | > 83 | 145 |
|  | LVM/LVEDV (G/ML) | 0.4-0.7 | 0.8-0.9 | > 0.9 | 149 | 0.5-0.9 | 01-1 | > 1 | 145 |
|  | RVEDVi (mL/m²) | 58-112 | 113-123 | > 123 | 149 | 68-123 | 124-133 | > 133 | 142 |
|  | RVESVi (mL/m²) | 23-50 | 51-57 | > 57 | 149 | 27-57 | 58-62 | > 62 | 143 |
|  | RVEF (%) | 48-69 | 45-47 | < 45 | 149 | 45-67 | 42-44 | < 42 | 145 |
|  | RVSVi (mL/m²) | 32-70 | 28-31 | < 28 | 149 | 36-72 | 31-35 | < 31 | 142 |
|  | LAVi max (mL/m²) | 19-46 | 47-51 | > 51 | 111 | 18-48 | 49-55 | > 55 | 103 |
|  | LAEF (%) | 57-83 | 52-56 | < 52 | 111 | 55-80 | 51-54 | < 51 | 104 |
|  | RAVi max (mL/m²) | 22-50 | 51-55 | > 55 | 129 | 23-60 | 61-67 | > 67 | 121 |
|  | RAEF (%) | 42-75 | 37-41 | < 37 | 122 | 36-71 | 32-35 | < 32 | 120 |
| 40-49 | LVEDVi (mL/m²) | 58-104 | 105-113 | > 113 | 571 | 63-114 | 115-122 | > 122 | 478 |
|  | LVESVi (mL/m²) | 20-45 | 46-51 | > 51 | 571 | 23-50 | 51-54 | > 54 | 478 |
|  | LVSVi (mL/m²) | 34-63 | 30-33 | < 30 | 571 | 35-69 | 31-34 | < 31 | 485 |
|  | LVEF (%) | 51-70 | 48-50 | < 48 | 570 | 49-69 | 46-48 | < 46 | 499 |
|  | LVMi (g/m²) | 32-57 | 58-62 | > 62 | 571 | 41-74 | 75-82 | > 82 | 497 |
|  | LVM/LVEDV (G/ML) | 0.4-0.8 | 0.9-0.9 | > 0.9 | 571 | 0.5-0.9 | 1-1.1 | > 1.1 | 475 |
|  | RVEDVi (mL/m²) | 56-107 | 108-117 | > 117 | 571 | 67-125 | 126-135 | > 135 | 484 |
|  | RVESVi (mL/m²) | 20-47 | 48-53 | > 53 | 571 | 25-56 | 57-61 | > 61 | 478 |
|  | RVEF (%) | 49-71 | 46-48 | < 46 | 568 | 46-69 | 42-45 | < 42 | 498 |
|  | RVSVi (mL/m²) | 32-67 | 28-31 | < 28 | 569 | 35-73 | 30-34 | < 30 | 478 |
|  | LAVi max (mL/m²) | 20-48 | 49-53 | > 53 | 498 | 18-48 | 49-55 | > 55 | 425 |
|  | LAEF (%) | 57-83 | 53-56 | < 53 | 498 | 55-81 | 51-54 | < 51 | 426 |
|  | RAVi max (mL/m²) | 22-52 | 53-58 | > 58 | 498 | 23-63 | 64-70 | > 70 | 414 |
|  | RAEF (%) | 41-74 | 36-40 | < 36 | 489 | 37-72 | 32-36 | < 32 | 411 |
| 50-59 | LVEDVi (mL/m²) | 55-99 | 100-108 | > 108 | 1290 | 58-109 | 110-118 | > 118 | 1125 |
|  | LVESVi (mL/m²) | 18-42 | 43-47 | > 47 | 1289 | 20-47 | 48-52 | > 52 | 1121 |
|  | LVSVi (mL/m²) | 33-62 | 29-32 | < 29 | 1289 | 34-68 | 29-33 | < 29 | 1142 |
|  | LVEF (%) | 53-72 | 49-52 | < 49 | 1290 | 50-71 | 47-49 | < 47 | 1155 |
|  | LVMi (g/m²) | 32-57 | 58-63 | > 63 | 1290 | 41-73 | 74-80 | > 80 | 1146 |
|  | LVM/LVEDV (G/ML) | 0.4-0.8 | 0.9-1 | > 1 | 1290 | 0.5-1 | 1.1-1.1 | > 1.1 | 1112 |
|  | RVEDVi (mL/m²) | 53-102 | 103-112 | > 112 | 1288 | 60-115 | 116-125 | > 125 | 1133 |
|  | RVESVi (mL/m²) | 18-43 | 44-49 | > 49 | 1288 | 22-52 | 53-58 | > 58 | 1123 |
|  | RVEF (%) | 50-73 | 46-49 | < 46 | 1286 | 46-70 | 42-45 | < 42 | 1144 |
|  | RVSVi (mL/m²) | 31-65 | 27-30 | < 27 | 1283 | 32-70 | 26-31 | < 26 | 1140 |
|  | LAVi max (mL/m²) | 19-48 | 49-53 | > 53 | 1171 | 17-48 | 49-55 | > 55 | 986 |
|  | LAEF (%) | 56-82 | 52-55 | < 52 | 1168 | 55-82 | 50-54 | < 50 | 991 |
|  | RAVi max (mL/m²) | 21-54 | 55-61 | > 61 | 1099 | 21-60 | 61-68 | > 68 | 884 |
|  | RAEF (%) | 40-74 | 35-39 | < 35 | 1089 | 37-73 | 32-36 | < 32 | 880 |
| 60-69 | LVEDVi (mL/m²) | 51-95 | 96-105 | > 105 | 1210 | 54-108 | 109-117 | > 117 | 1051 |
|  | LVESVi (mL/m²) | 16-39 | 40-45 | > 45 | 1208 | 18-46 | 47-51 | > 51 | 1045 |
|  | LVSVi (mL/m²) | 32-60 | 28-31 | < 28 | 1210 | 32-66 | 27-31 | < 27 | 1054 |
|  | LVEF (%) | 53-73 | 49-52 | < 49 | 1207 | 51-71 | 47-50 | < 47 | 1059 |
|  | LVMi (g/m²) | 33-58 | 59-65 | > 65 | 1210 | 40-72 | 73-79 | > 79 | 1061 |
|  | LVM/LVEDV (G/ML) | 0.5-0.9 | 01-1 | > 1 | 1210 | 0.5-1 | 1.1-1.2 | > 1.2 | 1043 |
|  | RVEDVi (mL/m²) | 51-99 | 100-109 | > 109 | 1208 | 57-113 | 114-123 | > 123 | 1052 |
|  | RVESVi (mL/m²) | 16-41 | 42-47 | > 47 | 1209 | 20-51 | 52-57 | > 57 | 1048 |
|  | RVEF (%) | 50-74 | 46-49 | < 46 | 1207 | 46-71 | 42-45 | < 42 | 1056 |
|  | RVSVi (mL/m²) | 30-64 | 26-29 | < 26 | 1206 | 30-70 | 24-29 | < 24 | 1055 |
|  | LAVi max (mL/m²) | 18-48 | 49-55 | > 55 | 1077 | 16-49 | 50-56 | > 56 | 889 |
|  | LAEF (%) | 54-82 | 49-53 | < 49 | 1076 | 53-83 | 48-52 | < 48 | 891 |
|  | RAVi max (mL/m²) | 21-57 | 58-66 | > 66 | 962 | 21-62 | 63-71 | > 71 | 719 |
|  | RAEF (%) | 40-75 | 35-39 | < 35 | 940 | 37-74 | 32-36 | < 32 | 694 |
| 70+ | LVEDVi (mL/m²) | 50-94 | 95-104 | > 104 | 1208 | 51-108 | 109-119 | > 119 | 1334 |
|  | LVESVi (mL/m²) | 15-39 | 40-45 | > 45 | 1207 | 18-46 | 47-53 | > 53 | 1319 |
|  | LVSVi (mL/m²) | 31-60 | 28-30 | < 28 | 1210 | 30-65 | 25-29 | < 25 | 1346 |
|  | LVEF (%) | 53-74 | 49-52 | < 49 | 1207 | 50-71 | 46-49 | < 46 | 1335 |
|  | LVMi (g/m²) | 33-59 | 60-67 | > 67 | 1210 | 39-71 | 72-78 | > 78 | 1342 |
|  | LVM/LVEDV (G/ML) | 0.5-0.9 | 1-1.1 | > 1.1 | 1208 | 0.5-1 | 1.1-1.2 | > 1.2 | 1327 |
|  | RVEDVi (mL/m²) | 50-98 | 99-109 | > 109 | 1208 | 55-113 | 114-123 | > 123 | 1335 |
|  | RVESVi (mL/m²) | 15-40 | 41-46 | > 46 | 1208 | 20-51 | 52-57 | > 57 | 1333 |
|  | RVEF (%) | 50-75 | 44-49 | < 44 | 1204 | 46-72 | 41-45 | < 41 | 1321 |
|  | RVSVi (mL/m²) | 28-64 | 24-27 | < 24 | 1205 | 28-70 | 21-27 | < 21 | 1336 |
|  | LAVi max (mL/m²) | 17-52 | 53-61 | > 61 | 1047 | 15-50 | 51-58 | > 58 | 1034 |
|  | LAEF (%) | 51-83 | 45-50 | < 45 | 1037 | 50-84 | 45-49 | < 45 | 1024 |
|  | RAVi max (mL/m²) | 20-60 | 61-71 | > 71 | 849 | 21-66 | 67-75 | > 75 | 689 |
|  | RAEF (%) | 41-77 | 36-40 | < 36 | 828 | 37-75 | 32-36 | < 32 | 676 |

Footnote: All measurements were derived using suiteHEART software with smooth endocardial segmentation; papillary muscles and trabeculations were excluded from left ventricular mass measurements. Volumetric and mass measurements were indexed to bsa. “Reference range,” “Mild,” and “At least moderate” capture the progression of deviation from the reference range for each metric. Reference ranges are defined as the interval between the 2.5th and 97.5th centiles, derived from age-specific generalized addreference range are derived from LMS-based thresholds (P0.5, P2.5, P97.5, and P99.5) and describe the position of a measurement within the reference distribution rather than disease severity or diagnostic abnormality. Reference ranges are based on a White an adult population.

Abbreviations: LV = left ventricle; RV = right ventricle; LA = left atrium; RA = right atrium; EDV = end-diastolic volume; ESV = end-systolic volume; SV = stroke volume; EF = ejection fraction; CO = cardiac output.

### 10. Table: Reference ranges and centile-based categories for cardiac chamber measurements derived using suiteHEART software (anatomical segmentation), indexed to bsa, in an adult population

|  |  | **Female** | | | | **Male** | | | |
| --- | --- | --- | --- | --- | --- | --- | --- | --- | --- |
| **Age group** | **Metrics** | **Reference range** | **Mild** | **At least moderate** | **n** | **Reference range** | **Mild** | **At least moderate** | **n** |
| 18-29 | LVEDVi (mL/m²) | 55-99 | 100-107 | > 107 | 117 | 64-111 | 112-119 | > 119 | 120 |
|  | LVESVi (mL/m²) | 17-39 | 40-44 | > 44 | 117 | 19-47 | 48-52 | > 52 | 122 |
|  | LVSVi (mL/m²) | 35-65 | 30-34 | < 30 | 117 | 39-73 | 35-38 | < 35 | 120 |
|  | LVEF (%) | 55-75 | 52-54 | < 52 | 117 | 52-74 | 49-51 | < 49 | 123 |
|  | LVMi (g/m²) | 39-66 | 67-71 | > 71 | 117 | 51-85 | 86-91 | > 91 | 120 |
|  | LVM/LVEDV (G/ML) | 0.5-0.9 | 1-1.1 | > 1.1 | 117 | 0.6-1.1 |  | > 1.2 | 118 |
|  | RVEDVi (mL/m²) | 61-112 | 113-120 | > 120 | 117 | 73-125 | 126-133 | > 133 | 117 |
|  | RVESVi (mL/m²) | 23-51 | 52-57 | > 57 | 116 | 27-58 | 59-63 | > 63 | 117 |
|  | RVEF (%) | 47-70 | 44-46 | < 44 | 115 | 46-68 | 43-45 | < 43 | 121 |
|  | RVSVi (mL/m²) | 35-69 | 31-34 | < 31 | 116 | 40-75 | 36-39 | < 36 | 120 |
|  | LAESVi (mL/m²) | 18-43 | 44-48 | > 48 | 77 | 19-49 | 50-56 | > 56 | 79 |
|  | LAEF (%) | 56-84 | 51-55 | < 51 | 77 | 54-79 | 51-53 | < 51 | 77 |
|  | RAESVi (mL/m²) | 23-48 | 49-53 | > 53 | 88 | 23-59 | 60-66 | > 66 | 102 |
|  | RAEF (%) | 42-75 | 38-41 | < 38 | 83 | 36-71 | 31-35 | < 31 | 99 |
| 30-39 | LVEDVi (mL/m²) | 52-95 | 96-103 | > 103 | 149 | 56-101 | 102-108 | > 108 | 145 |
|  | LVESVi (mL/m²) | 15-37 | 38-42 | > 42 | 149 | 16-41 | 42-46 | > 46 | 145 |
|  | LVSVi (mL/m²) | 33-63 | 29-32 | < 29 | 148 | 35-67 | 31-34 | < 31 | 145 |
|  | LVEF (%) | 56-76 | 52-55 | < 52 | 149 | 54-76 | 50-53 | < 50 | 146 |
|  | LVMi (g/m²) | 39-66 | 67-72 | > 72 | 149 | 50-84 | 85-90 | > 90 | 142 |
|  | LVM/LVEDV (G/ML) | 0.5-1 | 1.1-1.1 | > 1.1 | 149 | 0.6-1.2 | 1.3-1.3 | > 1.3 | 143 |
|  | RVEDVi (mL/m²) | 58-109 | 110-118 | > 118 | 149 | 66-119 | 120-128 | > 128 | 144 |
|  | RVESVi (mL/m²) | 21-49 | 50-55 | > 55 | 149 | 23-55 | 56-60 | > 60 | 145 |
|  | RVEF (%) | 49-71 | 45-48 | < 45 | 149 | 47-70 | 43-46 | < 43 | 146 |
|  | RVSVi (mL/m²) | 34-68 | 30-33 | < 30 | 149 | 37-73 | 32-36 | < 32 | 143 |
|  | LAVi max (mL/m²) | 19-46 | 47-51 | > 51 | 111 | 18-48 | 49-55 | > 55 | 103 |
|  | LAEF (%) | 57-83 | 52-56 | < 52 | 111 | 55-80 | 51-54 | < 51 | 104 |
|  | RAVi max (mL/m²) | 22-50 | 51-55 | > 55 | 129 | 23-60 | 61-67 | > 67 | 121 |
|  | RAEF (%) | 42-75 | 37-41 | < 37 | 122 | 36-71 | 32-35 | < 32 | 120 |
| 40-49 | LVEDVi (mL/m²) | 51-94 | 95-102 | > 102 | 571 | 54-103 | 104-111 | > 111 | 493 |
|  | LVESVi (mL/m²) | 13-36 | 37-41 | > 41 | 571 | 14-41 | 42-46 | > 46 | 496 |
|  | LVSVi (mL/m²) | 33-62 | 29-32 | < 29 | 571 | 35-68 | 30-34 | < 30 | 488 |
|  | LVEF (%) | 57-78 | 53-56 | < 53 | 570 | 55-77 | 51-54 | < 51 | 498 |
|  | LVMi (g/m²) | 39-67 | 68-73 | > 73 | 571 | 49-83 | 84-89 | > 89 | 467 |
|  | LVM/LVEDV (G/ML) | 0.6-1 | 1.1-1.2 | > 1.2 | 571 | 0.6-1.2 | 1.3-1.4 | > 1.4 | 464 |
|  | RVEDVi (mL/m²) | 54-104 | 105-113 | > 113 | 571 | 62-119 | 120-128 | > 128 | 490 |
|  | RVESVi (mL/m²) | 18-45 | 46-50 | > 50 | 571 | 20-52 | 53-58 | > 58 | 497 |
|  | RVEF (%) | 50-73 | 47-49 | < 47 | 567 | 48-72 | 43-47 | < 43 | 497 |
|  | RVSVi (mL/m²) | 32-66 | 28-31 | < 28 | 569 | 36-74 | 30-35 | < 30 | 483 |
|  | LAVi max (mL/m²) | 20-48 | 49-53 | > 53 | 498 | 18-48 | 49-55 | > 55 | 425 |
|  | LAEF (%) | 57-83 | 53-56 | < 53 | 498 | 55-81 | 51-54 | < 51 | 426 |
|  | RAVi max (mL/m²) | 22-52 | 53-58 | > 58 | 498 | 23-63 | 64-70 | > 70 | 414 |
|  | RAEF (%) | 41-74 | 36-40 | < 36 | 489 | 37-72 | 32-36 | < 32 | 411 |
| 50-59 | LVEDVi (mL/m²) | 48-89 | 90-98 | > 98 | 1290 | 50-100 | 101-109 | > 109 | 1155 |
|  | LVESVi (mL/m²) | 11-33 | 34-38 | > 38 | 1290 | 12-38 | 39-44 | > 44 | 1158 |
|  | LVSVi (mL/m²) | 33-62 | 29-32 | < 29 | 1289 | 33-67 | 29-32 | < 29 | 1147 |
|  | LVEF (%) | 58-80 | 54-57 | < 54 | 1285 | 56-79 | 51-55 | < 51 | 1156 |
|  | LVMi (g/m²) | 39-68 | 69-75 | > 75 | 1289 | 48-82 | 83-88 | > 88 | 1081 |
|  | LVM/LVEDV (G/ML) | 0.6-1.1 | 1.2-1.3 | > 1.3 | 1289 | 0.7-1.3 | 1.4-1.5 | > 1.5 | 1079 |
|  | RVEDVi (mL/m²) | 49-95 | 96-104 | > 104 | 1288 | 54-108 | 109-118 | > 118 | 1149 |
|  | RVESVi (mL/m²) | 15-39 | 40-44 | > 44 | 1288 | 17-48 | 49-54 | > 54 | 1149 |
|  | RVEF (%) | 52-75 | 48-51 | < 48 | 1284 | 48-74 | 43-47 | < 43 | 1154 |
|  | RVSVi (mL/m²) | 30-63 | 26-29 | < 26 | 1285 | 31-68 | 26-30 | < 26 | 1146 |
|  | LAVi max (mL/m²) | 19-48 | 49-53 | > 53 | 1171 | 17-48 | 49-55 | > 55 | 986 |
|  | LAEF (%) | 56-82 | 52-55 | < 52 | 1168 | 55-82 | 50-54 | < 50 | 991 |
|  | RAVi max (mL/m²) | 21-54 | 55-61 | > 61 | 1099 | 21-60 | 61-68 | > 68 | 884 |
|  | RAEF (%) | 40-74 | 35-39 | < 35 | 1089 | 37-73 | 32-36 | < 32 | 880 |
| 60-69 | LVEDVi (mL/m²) | 45-86 | 87-94 | > 94 | 1210 | 46-98 | 99-107 | > 107 | 1066 |
|  | LVESVi (mL/m²) | 9-31 | 32-36 | > 36 | 1209 | 11-37 | 38-43 | > 43 | 1064 |
|  | LVSVi (mL/m²) | 32-60 | 28-31 | < 28 | 1210 | 31-65 | 26-30 | < 26 | 1055 |
|  | LVEF (%) | 59-82 | 55-58 | < 55 | 1203 | 56-80 | 51-55 | < 51 | 1066 |
|  | LVMi (g/m²) | 39-69 | 70-76 | > 76 | 1209 | 47-81 | 82-87 | > 87 | 1016 |
|  | LVM/LVEDV (G/ML) | 0.6-1.2 | 1.3-1.4 | > 1.4 | 1209 | 0.7-1.3 | 1.4-1.6 | > 1.6 | 1014 |
|  | RVEDVi (mL/m²) | 47-92 | 93-101 | > 101 | 1208 | 51-107 | 108-117 | > 117 | 1066 |
|  | RVESVi (mL/m²) | 13-36 | 37-42 | > 42 | 1209 | 15-46 | 47-52 | > 52 | 1067 |
|  | RVEF (%) | 53-77 | 47-52 | < 47 | 1203 | 48-76 | 43-47 | < 43 | 1061 |
|  | RVSVi (mL/m²) | 29-62 | 25-28 | < 25 | 1207 | 29-68 | 23-28 | < 23 | 1057 |
|  | LAVi max (mL/m²) | 18-48 | 49-55 | > 55 | 1077 | 16-49 | 50-56 | > 56 | 889 |
|  | LAEF (%) | 54-82 | 49-53 | < 49 | 1076 | 53-83 | 48-52 | < 48 | 891 |
|  | RAVi max (mL/m²) | 21-57 | 58-66 | > 66 | 962 | 21-62 | 63-71 | > 71 | 719 |
|  | RAEF (%) | 40-75 | 35-39 | < 35 | 940 | 37-74 | 32-36 | < 32 | 694 |
| 70+ | LVEDVi (mL/m²) | 43-84 | 85-93 | > 93 | 1209 | 44-97 | 98-107 | > 107 | 1347 |
|  | LVESVi (mL/m²) | 8-30 | 31-36 | > 36 | 1210 | 10-38 | 39-45 | > 45 | 1343 |
|  | LVSVi (mL/m²) | 31-59 | 27-30 | < 27 | 1210 | 30-64 | 25-29 | < 25 | 1347 |
|  | LVEF (%) | 59-83 | 55-58 | < 55 | 1194 | 56-81 | 51-55 | < 51 | 1336 |
|  | LVMi (g/m²) | 39-70 | 71-78 | > 78 | 1209 | 46-80 | 81-86 | > 86 | 1316 |
|  | LVM/LVEDV (G/ML) | 0.6-1.3 | 1.4-1.5 | > 1.5 | 1208 | 0.7-1.4 | 1.5-1.7 | > 1.7 | 1314 |
|  | RVEDVi (mL/m²) | 46-92 | 93-102 | > 102 | 1208 | 50-106 | 107-117 | > 117 | 1346 |
|  | RVESVi (mL/m²) | 12-36 | 37-41 | > 41 | 1208 | 15-45 | 46-52 | > 52 | 1347 |
|  | RVEF (%) | 52-78 | 46-51 | < 46 | 1194 | 48-77 | 42-47 | < 42 | 1330 |
|  | RVSVi (mL/m²) | 28-63 | 24-27 | < 24 | 1206 | 27-69 | 21-26 | < 21 | 1339 |
|  | LAVi max (mL/m²) | 17-52 | 53-61 | > 61 | 1047 | 15-50 | 51-58 | > 58 | 1034 |
|  | LAEF (%) | 51-83 | 45-50 | < 45 | 1037 | 50-84 | 45-49 | < 45 | 1024 |
|  | RAVi max (mL/m²) | 20-60 | 61-71 | > 71 | 849 | 21-66 | 67-75 | > 75 | 689 |
|  | RAEF (%) | 41-77 | 36-40 | < 36 | 828 | 37-75 | 32-36 | < 32 | 676 |

Footnote: All measurements were derived using suiteHEART software with anatomical endocardial segmentation; papillary muscles and trabeculations were included in left ventricular mass measurements. “Reference range,” “Mild,” and “At least moderate” capture the progression of deviation from the reference range for each metric. Reference ranges are defined as the interval between the 2.5th and 97.5th centiles, derived from age-specific generalized add–Green distribution. Centile-based categories beyond the reference range are derived from LMS-based thresholds (P0.5, P2.5, P97.5, and P99.5) and describe the position of a measurement within the reference distribution rather than disease severity or diagnostic abnormality. Reference ranges are based on a White an adult population.

Abbreviations: LV = left ventricle; RV = right ventricle; LA = left atrium; RA = right atrium; EDV = end-diastolic volume; ESV = end-systolic volume; SV = stroke volume; EF = ejection fraction; CO = cardiac output.
