## Supplementary material for "Cardiovascular magnetic resonance reference ranges and centile-based categories for atrial and ventricular cardiac structure and function: 2026 Update from the Healthy Hearts Consortium": Ethnicity stratified reference ranges

**Ethnicity-stratified reference ranges and centile-based cardiac chamber measurements using the Healthy Hearts Consortium data**

Values reported in this document are stratified by participant ethnicity.

Details about the consortium data may be found at: <https://healthy-hearts.org.uk/>

Reference ranges and centile-based measurements were calculated using the GLMSS package in R. Details may be found at <https://www.gamlss.com/> and <https://cran.r-project.org/web/packages/gamlss/index.html>.

In the tables ‘Reference range’, ‘Mild’ and ‘At least moderate’ capture the progression of deviation from the reference range for each metric. Reference ranges are defined as the interval between the 2.5th and 97.5th centiles, derived from age-specific generalized additive models for location, scale and shape (GAMLSS) using the Box–Cox Cole distribution. Values falling in the 0.5th–2.5th or 97.5th–99.5th centiles are classified as mild deviation, and those beyond the 0.5th or 99.5th centiles as at least moderate deviation.

Please report any concerns to:

### 1. Table: Reference ranges and centile-based categories for cardiac chamber measurements derived using cvi42 software (smooth segmentation), indexed to height, in a White adult population

|  |  | **Female** | | | | | **Male** | | | |
| --- | --- | --- | --- | --- | --- | --- | --- | --- | --- | --- |
| **Age group** | **Metrics** | **Reference range** | **Mild** | **At least moderate** | **n** | **Reference range** | | **Mild** | **At least moderate** | **n** |
| 18-29 | LVEDVi | 60-107 | 108-114 | > 114 | 102 | 76-129 | | 130-136 | > 136 | 99 |
|  | LVESVi | 20-51 | 52-56 | > 56 | 102 | 25-57 | | 58-60 | > 60 | 94 |
|  | LVSVi | 35-66 | 31-34 | < 31 | 102 | 39-77 | | 33-38 | < 33 | 99 |
|  | LVEF | 51-71 | 49-50 | < 49 | 100 | 47-68 | | 45-46 | < 45 | 100 |
|  | LVCOi | 2.3-5 | 2.1-2.2 | < 2.1 | 102 | 2.4-5.4 | | 1.7-2.3 | < 1.7 | 97 |
|  | LVMi | 34-57 | 58-62 | > 62 | 102 | 45-80 | | 81-86 | > 86 | 105 |
|  | LVM/LVEDV | 0.4-0.7 |  | > 0.7 | 102 | 0.5-0.8 | |  | > 0.8 | 99 |
|  | RVEDVi | 57-113 | 114-121 | > 121 | 102 | 75-139 | | 140-147 | > 147 | 98 |
|  | RVESVi | 14-56 | 57-62 | > 62 | 100 | 27-72 | | 73-78 | > 78 | 89 |
|  | RVEF | 43-69 | 39-42 | < 39 | 97 | 41-66 | | 37-40 | < 37 | 97 |
|  | RVCOi | 2-5.2 | 1.6-1.9 | < 1.6 | 101 | 2.3-5.5 | | 1.8-2.2 | < 1.8 | 97 |
|  | RVSVi | 32-69 | 27-31 | < 27 | 102 | 39-80 | | 33-38 | < 33 | 98 |
|  | LAVi max | 25-51 | 52-55 | > 55 | 50 | 28-63 | | 64-70 | > 70 | 52 |
|  | LAEF | 56-75 | 54-55 | < 54 | 50 | 53-72 | | 50-52 | < 50 | 52 |
|  | RAVi max | 21-51 | 52-56 | > 56 | 80 | 28-68 | | 69-77 | > 77 | 82 |
|  | RAEF | 38-78 | 33-37 | < 33 | 78 | 35-74 | | 31-34 | < 31 | 82 |
| 30-39 | LVEDVi | 59-106 | 107-113 | > 113 | 137 | 69-121 | | 122-128 | > 128 | 130 |
|  | LVESVi | 19-49 | 50-54 | > 54 | 137 | 23-55 | | 56-59 | > 59 | 129 |
|  | LVSVi | 34-66 | 30-33 | < 30 | 134 | 37-75 | | 31-36 | < 31 | 131 |
|  | LVEF | 51-71 | 48-50 | < 48 | 132 | 48-69 | | 45-47 | < 45 | 134 |
|  | LVCOi | 2.3-5 | 2-2.2 | < 2 | 135 | 2.3-5.3 | | 1.8-2.2 | < 1.8 | 127 |
|  | LVMi | 33-58 | 59-63 | > 63 | 137 | 42-76 | | 77-82 | > 82 | 134 |
|  | LVM/LVEDV | 0.5-0.7 | 0.8-0.8 | > 0.8 | 137 | 0.5-0.8 | | 0.9-0.9 | > 0.9 | 131 |
|  | RVEDVi | 57-112 | 113-122 | > 122 | 137 | 73-138 | | 139-147 | > 147 | 130 |
|  | RVESVi | 15-53 | 54-58 | > 58 | 135 | 26-68 | | 69-74 | > 74 | 125 |
|  | RVEF | 46-70 | 43-45 | < 43 | 127 | 43-67 | | 39-42 | < 39 | 131 |
|  | RVCOi | 2.1-5.2 | 1.7-2 | < 1.7 | 133 | 2.4-5.5 | | 1.9-2.3 | < 1.9 | 123 |
|  | RVSVi | 32-70 | 28-31 | < 28 | 134 | 38-80 | | 32-37 | < 32 | 132 |
|  | LAVi max | 25-55 | 56-60 | > 60 | 84 | 27-64 | | 65-71 | > 71 | 66 |
|  | LAEF | 55-76 | 53-54 | < 53 | 83 | 53-73 | | 50-52 | < 50 | 66 |
|  | RAVi max | 21-52 | 53-57 | > 57 | 113 | 27-69 | | 70-77 | > 77 | 119 |
|  | RAEF | 38-75 | 33-37 | < 33 | 110 | 35-72 | | 31-34 | < 31 | 118 |
| 40-49 | LVEDVi | 59-108 | 109-117 | > 117 | 518 | 68-125 | | 126-133 | > 133 | 428 |
|  | LVESVi | 18-47 | 48-52 | > 52 | 518 | 22-55 | | 56-59 | > 59 | 422 |
|  | LVSVi | 35-68 | 31-34 | < 31 | 517 | 38-77 | | 31-37 | < 31 | 434 |
|  | LVEF | 51-71 | 48-50 | < 48 | 511 | 48-70 | | 45-47 | < 45 | 434 |
|  | LVCOi | 2.2-4.9 | 1.9-2.1 | < 1.9 | 515 | 2.3-5.1 | | 1.8-2.2 | < 1.8 | 425 |
|  | LVMi | 33-59 | 60-64 | > 64 | 518 | 43-79 | | 80-85 | > 85 | 443 |
|  | LVM/LVEDV | 0.5-0.7 | 0.8-0.8 | > 0.8 | 518 | 0.5-0.8 | | 0.9-0.9 | > 0.9 | 428 |
|  | RVEDVi | 57-113 | 114-123 | > 123 | 517 | 71-137 | | 138-146 | > 146 | 430 |
|  | RVESVi | 16-50 | 51-54 | > 54 | 517 | 24-64 | | 65-70 | > 70 | 418 |
|  | RVEF | 50-71 | 46-49 | < 46 | 510 | 45-68 | | 42-44 | < 42 | 429 |
|  | RVCOi | 2.1-5.1 | 1.8-2 | < 1.8 | 514 | 2.3-5.4 | | 1.9-2.2 | < 1.9 | 419 |
|  | RVSVi | 34-71 | 29-33 | < 29 | 516 | 38-82 | | 32-37 | < 32 | 430 |
|  | LAVi max | 25-59 | 60-65 | > 65 | 440 | 25-64 | | 65-72 | > 72 | 358 |
|  | LAEF | 55-78 | 52-54 | < 52 | 434 | 53-76 | | 50-52 | < 50 | 353 |
|  | RAVi max | 23-59 | 60-66 | > 66 | 500 | 28-77 | | 78-87 | > 87 | 408 |
|  | RAEF | 38-72 | 33-37 | < 33 | 497 | 35-69 | | 31-34 | < 31 | 410 |
| 50-59 | LVEDVi | 59-109 | 110-119 | > 119 | 911 | 65-125 | | 126-133 | > 133 | 809 |
|  | LVESVi | 17-45 | 46-50 | > 50 | 909 | 21-54 | | 55-58 | > 58 | 803 |
|  | LVSVi | 35-70 | 31-34 | < 31 | 910 | 37-77 | | 30-36 | < 30 | 817 |
|  | LVEF | 51-72 | 48-50 | < 48 | 906 | 49-71 | | 45-48 | < 45 | 828 |
|  | LVCOi | 2.1-4.7 | 1.8-2 | < 1.8 | 906 | 2.2-5 | | 1.8-2.1 | < 1.8 | 816 |
|  | LVMi | 34-62 | 63-67 | > 67 | 911 | 44-83 | | 84-89 | > 89 | 832 |
|  | LVM/LVEDV | 0.5-0.7 | 0.8-0.8 | > 0.8 | 911 | 0.5-0.9 | | 01-1 | > 1 | 806 |
|  | RVEDVi | 57-113 | 114-123 | > 123 | 909 | 68-134 | | 135-143 | > 143 | 815 |
|  | RVESVi | 16-47 | 48-51 | > 51 | 909 | 23-61 | | 62-66 | > 66 | 814 |
|  | RVEF | 52-72 | 49-51 | < 49 | 906 | 47-69 | | 43-46 | < 43 | 829 |
|  | RVCOi | 2.1-4.9 | 1.8-2 | < 1.8 | 906 | 2.3-5.2 | | 1.8-2.2 | < 1.8 | 812 |
|  | RVSVi | 34-72 | 30-33 | < 30 | 909 | 37-80 | | 30-36 | < 30 | 816 |
|  | LAVi max | 24-62 | 63-69 | > 69 | 797 | 24-65 | | 66-73 | > 73 | 684 |
|  | LAEF | 53-79 | 50-52 | < 50 | 791 | 53-79 | | 50-52 | < 50 | 681 |
|  | RAVi max | 24-66 | 67-74 | > 74 | 854 | 27-78 | | 79-87 | > 87 | 761 |
|  | RAEF | 38-70 | 33-37 | < 33 | 844 | 35-68 | | 31-34 | < 31 | 771 |
| 60-69 | LVEDVi | 56-105 | 106-116 | > 116 | 918 | 62-123 | | 124-132 | > 132 | 794 |
|  | LVESVi | 16-43 | 44-48 | > 48 | 919 | 20-53 | | 54-58 | > 58 | 785 |
|  | LVSVi | 33-67 | 29-32 | < 29 | 918 | 35-76 | | 29-34 | < 29 | 797 |
|  | LVEF | 52-73 | 48-51 | < 48 | 914 | 49-72 | | 46-48 | < 46 | 793 |
|  | LVCOi | 2-4.5 | 1.8-1.9 | < 1.8 | 917 | 2.2-4.8 | | 1.8-2.1 | < 1.8 | 793 |
|  | LVMi | 33-62 | 63-68 | > 68 | 919 | 42-82 | | 83-89 | > 89 | 802 |
|  | LVM/LVEDV | 0.5-0.8 | 0.9-0.9 | > 0.9 | 918 | 0.5-0.9 | | 01-1 | > 1 | 792 |
|  | RVEDVi | 54-109 | 110-119 | > 119 | 917 | 64-130 | | 131-140 | > 140 | 797 |
|  | RVESVi | 16-44 | 45-49 | > 49 | 920 | 22-57 | | 58-62 | > 62 | 799 |
|  | RVEF | 52-73 | 49-51 | < 49 | 915 | 48-70 | | 44-47 | < 44 | 803 |
|  | RVCOi | 2-4.7 | 1.8-1.9 | < 1.8 | 917 | 2.2-5.1 | | 1.8-2.1 | < 1.8 | 796 |
|  | RVSVi | 33-70 | 29-32 | < 29 | 917 | 35-79 | | 28-34 | < 28 | 791 |
|  | LAVi max | 22-63 | 64-71 | > 71 | 797 | 22-66 | | 67-74 | > 74 | 645 |
|  | LAEF | 51-79 | 47-50 | < 47 | 776 | 51-80 | | 47-50 | < 47 | 632 |
|  | RAVi max | 24-67 | 68-76 | > 76 | 855 | 25-79 | | 80-88 | > 88 | 742 |
|  | RAEF | 38-71 | 33-37 | < 33 | 841 | 35-70 | | 31-34 | < 31 | 748 |
| 70+ | LVEDVi | 53-102 | 103-114 | > 114 | 1100 | 57-118 | | 119-129 | > 129 | 1250 |
|  | LVESVi | 15-41 | 42-45 | > 45 | 1100 | 19-52 | | 53-58 | > 58 | 1240 |
|  | LVSVi | 32-65 | 28-31 | < 28 | 1102 | 33-72 | | 27-32 | < 27 | 1254 |
|  | LVEF | 53-74 | 49-52 | < 49 | 1092 | 49-72 | | 45-48 | < 45 | 1239 |
|  | LVCOi | 2-4.4 | 1.7-1.9 | < 1.7 | 1099 | 2.1-4.6 | | 1.8-2 | < 1.8 | 1254 |
|  | LVMi | 32-61 | 62-67 | > 67 | 1102 | 40-80 | | 81-87 | > 87 | 1260 |
|  | LVM/LVEDV | 0.5-0.8 | 0.9-0.9 | > 0.9 | 1100 | 0.5-0.9 | | 1-1.1 | > 1.1 | 1249 |
|  | RVEDVi | 52-105 | 106-116 | > 116 | 1100 | 61-125 | | 126-136 | > 136 | 1258 |
|  | RVESVi | 15-42 | 43-46 | > 46 | 1100 | 20-54 | | 55-59 | > 59 | 1258 |
|  | RVEF | 53-73 | 49-52 | < 49 | 1093 | 49-70 | | 45-48 | < 45 | 1247 |
|  | RVCOi | 2-4.6 | 1.8-1.9 | < 1.8 | 1099 | 2.1-4.8 | | 1.7-2 | < 1.7 | 1251 |
|  | RVSVi | 32-67 | 28-31 | < 28 | 1100 | 33-76 | | 26-32 | < 26 | 1257 |
|  | LAVi max | 21-63 | 64-73 | > 73 | 942 | 20-67 | | 68-75 | > 75 | 953 |
|  | LAEF | 48-79 | 44-47 | < 44 | 903 | 48-80 | | 44-47 | < 44 | 914 |
|  | RAVi max | 23-68 | 69-78 | > 78 | 1028 | 23-78 | | 79-87 | > 87 | 1119 |
|  | RAEF | 38-72 | 33-37 | < 33 | 1001 | 35-73 | | 30-34 | < 30 | 1115 |

Footnote: All measurements were derived using cvi42 software with smooth endocardial segmentation; papillary muscles and trabeculations were excluded from left ventricular mass measurements. Volumetric and mass measurements were indexed to height. Reference ranges are defined as the interval between the 2.5th and 97.5th centiles derived from age-specific generalized additive models for location, scale and shape (GAMLSS) using the Box–Cox Cole–Green distribution. Centile-based categories beyond the reference range are derived from LMS-based thresholds (P0.5, P2.5, P97.5, and P99.5) and describe the position of a measurement within the reference distribution rather than disease severity or diagnostic abnormality. Reference ranges are based on a White adult population.

Abbreviations: LV = left ventricle; RV = right ventricle; LA = left atrium; RA = right atrium; EDV = end-diastolic volume; ESV = end-systolic volume; SV = stroke volume; EF = ejection fraction; CO = cardiac output.

### 2. Table: Reference ranges and centile-based categories for cardiac chamber measurements derived using cvi42 software (smooth segmentation), indexed to height, in a Black adult population

|  |  | **Female** | | | | | **Male** | | | |
| --- | --- | --- | --- | --- | --- | --- | --- | --- | --- | --- |
| **Age group** | **Metrics** | **Reference range** | **Mild** | **At least moderate** | **n** | **Reference range** | | **Mild** | **At least moderate** | **n** |
| 40-49 | LVEDVi | 63-113 | 114-122 | > 122 | 10 |  | |  |  |  |
|  | LVESVi | 22-49 | 50-55 | > 55 | 10 |  | |  |  |  |
|  | LVSVi | 36-72 | 32-35 | < 32 | 10 |  | |  |  |  |
|  | LVEF | 50-74 | 48-49 | < 48 | 10 |  | |  |  |  |
|  | LVCOi | 2.1-5.9 | 1.9-2 | < 1.9 | 10 |  | |  |  |  |
|  | LVMi | 34-68 | 69-74 | > 74 | 10 |  | |  |  |  |
|  | LVM/LVEDV | 0.5-0.8 | 0.9-0.9 | > 0.9 | 10 |  | |  |  |  |
|  | RVEDVi | 61-122 | 123-135 | > 135 | 10 |  | |  |  |  |
|  | RVESVi | 21-58 | 59-70 | > 70 | 10 |  | |  |  |  |
|  | RVEF | 52-73 | 49-51 | < 49 | 10 |  | |  |  |  |
|  | RVCOi | 2.1-5.4 | 1.8-2 | < 1.8 | 10 |  | |  |  |  |
|  | RVSVi | 36-73 | 30-35 | < 30 | 10 |  | |  |  |  |
|  | LAVi max |  |  |  |  |  | |  |  |  |
|  | LAEF |  |  |  |  |  | |  |  |  |
|  | RAVi max | 28-76 | 77-91 | > 91 | 10 |  | |  |  |  |
|  | RAEF | 30-65 | 25-29 | < 25 | 10 |  | |  |  |  |
| 50-59 | LVEDVi | 59-111 | 112-120 | > 120 | 95 | 57-122 | | 123-130 | > 130 | 74 |
|  | LVESVi | 20-48 | 49-54 | > 54 | 95 | 20-55 | | 56-61 | > 61 | 74 |
|  | LVSVi | 35-70 | 31-34 | < 31 | 95 | 34-74 | | 27-33 | < 27 | 75 |
|  | LVEF | 50-73 | 48-49 | < 48 | 95 | 49-71 | | 46-48 | < 46 | 76 |
|  | LVCOi | 2.1-5.3 | 1.9-2 | < 1.9 | 95 | 2.4-5 | | 2.1-2.3 | < 2.1 | 77 |
|  | LVMi | 34-68 | 69-74 | > 74 | 95 | 38-84 | | 85-89 | > 89 | 78 |
|  | LVM/LVEDV | 0.5-0.8 | 0.9-0.9 | > 0.9 | 95 | 0.5-0.9 | |  | > 0.9 | 74 |
|  | RVEDVi | 59-118 | 119-130 | > 130 | 95 | 73-133 | | 134-146 | > 146 | 75 |
|  | RVESVi | 19-53 | 54-60 | > 60 | 95 | 21-59 | | 60-64 | > 64 | 76 |
|  | RVEF | 51-73 | 48-50 | < 48 | 95 | 47-68 | | 44-46 | < 44 | 77 |
|  | RVCOi | 2.1-5.2 | 1.9-2 | < 1.9 | 94 | 2.3-5.2 | | 2-2.2 | < 2 | 77 |
|  | RVSVi | 35-72 | 30-34 | < 30 | 95 | 33-79 | | 25-32 | < 25 | 76 |
|  | LAVi max | 25-63 | 64-71 | > 71 | 89 | 24-64 | | 65-72 | > 72 | 64 |
|  | LAEF | 54-75 | 51-53 | < 51 | 88 | 53-76 | | 50-52 | < 50 | 64 |
|  | RAVi max | 27-72 | 73-85 | > 85 | 94 | 33-74 | | 75-82 | > 82 | 73 |
|  | RAEF | 31-65 | 26-30 | < 26 | 93 | 35-63 | | 32-34 | < 32 | 71 |
| 60-69 | LVEDVi | 54-107 | 108-116 | > 116 | 66 | 61-123 | | 124-135 | > 135 | 48 |
|  | LVESVi | 17-46 | 47-52 | > 52 | 66 | 18-57 | | 58-63 | > 63 | 47 |
|  | LVSVi | 33-67 | 29-32 | < 29 | 66 | 36-74 | | 32-35 | < 32 | 48 |
|  | LVEF | 50-72 | 46-49 | < 46 | 66 | 42-71 | | 32-41 | < 32 | 48 |
|  | LVCOi | 2-4.7 | 1.8-1.9 | < 1.8 | 66 | 2.2-5.1 | | 2-2.1 | < 2 | 48 |
|  | LVMi | 34-68 | 69-73 | > 73 | 66 | 45-85 | | 86-95 | > 95 | 48 |
|  | LVM/LVEDV | 0.5-0.8 | 0.9-0.9 | > 0.9 | 66 | 0.5-0.9 | | 01-1 | > 1 | 48 |
|  | RVEDVi | 55-113 | 114-122 | > 122 | 66 | 69-132 | | 133-148 | > 148 | 48 |
|  | RVESVi | 14-47 | 48-52 | > 52 | 66 | 18-57 | | 58-62 | > 62 | 48 |
|  | RVEF | 50-73 | 46-49 | < 46 | 65 | 48-71 | | 44-47 | < 44 | 48 |
|  | RVCOi | 2.2-4.9 | 1.9-2.1 | < 1.9 | 66 | 2.4-5.2 | | 2.1-2.3 | < 2.1 | 48 |
|  | RVSVi | 34-71 | 30-33 | < 30 | 66 | 39-78 | | 35-38 | < 35 | 48 |
|  | LAVi max | 23-63 | 64-71 | > 71 | 59 | 23-65 | | 66-74 | > 74 | 42 |
|  | LAEF | 50-75 | 45-49 | < 45 | 58 | 50-75 | | 46-49 | < 46 | 43 |
|  | RAVi max | 25-67 | 68-76 | > 76 | 64 | 28-79 | | 80-88 | > 88 | 45 |
|  | RAEF | 32-67 | 27-31 | < 27 | 66 | 32-64 | | 29-31 | < 29 | 46 |
| 70+ | LVEDVi | 48-103 | 104-113 | > 113 | 18 | 58-120 | | 121-138 | > 138 | 14 |
|  | LVESVi | 14-45 | 46-51 | > 51 | 18 | 14-49 | | 50-55 | > 55 | 14 |
|  | LVSVi | 31-65 | 27-30 | < 27 | 18 | 36-76 | | 33-35 | < 33 | 14 |
|  | LVEF | 48-72 | 41-47 | < 41 | 18 | 41-71 | | 32-40 | < 32 | 14 |
|  | LVCOi | 2-4.3 | 1.7-1.9 | < 1.7 | 18 | 2.1-5.4 | | 1.9-2 | < 1.9 | 14 |
|  | LVMi | 34-67 | 68-73 | > 73 | 18 | 44-87 | | 88-117 | > 117 | 14 |
|  | LVM/LVEDV | 0.5-0.9 |  | > 0.9 | 18 | 0.5-0.9 | | 01-1 | > 1 | 14 |
|  | RVEDVi | 52-108 | 109-116 | > 116 | 18 | 66-132 | | 133-154 | > 154 | 14 |
|  | RVESVi | 13-44 | 45-47 | > 47 | 18 | 16-55 | | 56-60 | > 60 | 14 |
|  | RVEF | 47-72 | 40-46 | < 40 | 18 | 48-74 | | 44-47 | < 44 | 14 |
|  | RVCOi | 2.2-4.7 | 2-2.1 | < 2 | 18 | 2.5-5 | | 2.3-2.4 | < 2.3 | 14 |
|  | RVSVi | 34-71 | 30-33 | < 30 | 18 | 41-78 | | 39-40 | < 39 | 14 |
|  | LAVi max | 21-63 | 64-71 | > 71 | 15 |  | |  |  |  |
|  | LAEF | 44-74 | 35-43 | < 35 | 16 |  | |  |  |  |
|  | RAVi max | 23-65 | 66-72 | > 72 | 17 | 22-84 | | 85-95 | > 95 | 12 |
|  | RAEF | 33-68 | 28-32 | < 28 | 17 | 29-66 | | 25-28 | < 25 | 12 |

Footnote: All measurements were derived using cvi42 software with smooth endocardial segmentation; papillary muscles and trabeculations were excluded from left ventricular mass measurements. Volumetric and mass measurements were indexed to height. Reference ranges are defined as the interval between the 2.5th and 97.5th centiles derived from age-specific generalized additive models for location, scale and shape (GAMLSS) using the Box–Cox Cole–Green distribution. Centile-based categories beyond the reference range are derived from LMS-based thresholds (P0.5, P2.5, P97.5, and P99.5) and describe the position of a measurement within the reference distribution rather than disease severity or diagnostic abnormality. Reference ranges are based on a Black adult population.

Abbreviations: LV = left ventricle; RV = right ventricle; LA = left atrium; RA = right atrium; EDV = end-diastolic volume; ESV = end-systolic volume; SV = stroke volume; EF = ejection fraction; CO = cardiac output.

### 3. Table: Reference ranges and centile-based categories for cardiac chamber measurements derived using cvi42 software (smooth segmentation), indexed to height, in an Asian adult population

|  |  | **Female** | | | | **Male** | | | |
| --- | --- | --- | --- | --- | --- | --- | --- | --- | --- |
| **Age group** | **Metrics** | **Reference range** | **Mild** | **At least moderate** | **n** | **Reference range** | **Mild** | **At least moderate** | **n** |
| 40-49 | LVEDVi | 53-90 | 91-96 | > 96 | 10 | 62-112 | 113-122 | > 122 | 20 |
|  | LVESVi | 18-36 | 37-41 | > 41 | 10 | 20-50 | 51-56 | > 56 | 20 |
|  | LVSVi | 33-60 | 30-32 | < 30 | 10 | 40-62 | 37-39 | < 37 | 20 |
|  | LVEF | 53-72 | 49-52 | < 49 | 10 | 50-70 | 47-49 | < 47 | 20 |
|  | LVCOi | 2.1-4.2 | 1.9-2 | < 1.9 | 10 | 2.2-4.6 | 2-2.1 | < 2 | 20 |
|  | LVMi | 30-53 | 54-58 | > 58 | 10 | 38-69 | 70-75 | > 75 | 20 |
|  | LVM/LVEDV | 0.5-0.7 | 0.8-0.8 | > 0.8 | 10 | 0.5-0.8 | 0.9-0.9 | > 0.9 | 20 |
|  | RVEDVi | 52-95 | 96-105 | > 105 | 10 | 65-123 | 124-136 | > 136 | 20 |
|  | RVESVi | 16-41 | 42-46 | > 46 | 10 | 23-53 | 54-58 | > 58 | 20 |
|  | RVEF | 52-72 | 48-51 | < 48 | 10 | 49-67 | 46-48 | < 46 | 20 |
|  | RVCOi | 2-3.9 | 1.8-1.9 | < 1.8 | 10 | 2.3-4.4 | 2.1-2.2 | < 2.1 | 20 |
|  | RVSVi | 32-60 | 30-31 | < 30 | 10 | 35-73 | 31-34 | < 31 | 20 |
|  | LAVi max | 23-51 | 52-56 | > 56 | 10 | 20-57 | 58-64 | > 64 | 20 |
|  | LAEF | 57-81 | 54-56 | < 54 | 10 | 58-84 | 55-57 | < 55 | 20 |
|  | RAVi max | 22-47 | 48-51 | > 51 | 10 | 23-70 | 71-79 | > 79 | 20 |
|  | RAEF | 37-71 | 35-36 | < 35 | 10 | 32-63 | 27-31 | < 27 | 20 |
| 50-59 | LVEDVi | 52-90 | 91-97 | > 97 | 83 | 58-110 | 111-119 | > 119 | 133 |
|  | LVESVi | 17-38 | 39-43 | > 43 | 83 | 19-49 | 50-55 | > 55 | 133 |
|  | LVSVi | 32-60 | 30-31 | < 30 | 83 | 33-67 | 28-32 | < 28 | 133 |
|  | LVEF | 53-73 | 49-52 | < 49 | 82 | 50-71 | 47-49 | < 47 | 130 |
|  | LVCOi | 2-4.2 | 1.8-1.9 | < 1.8 | 83 | 2.1-4.5 | 1.9-2 | < 1.9 | 133 |
|  | LVMi | 30-54 | 55-60 | > 60 | 83 | 38-72 | 73-81 | > 81 | 133 |
|  | LVM/LVEDV | 0.5-0.7 | 0.8-0.8 | > 0.8 | 83 | 0.5-0.8 | 0.9-0.9 | > 0.9 | 133 |
|  | RVEDVi | 51-96 | 97-107 | > 107 | 83 | 60-119 | 120-131 | > 131 | 133 |
|  | RVESVi | 16-40 | 41-45 | > 45 | 83 | 21-52 | 53-58 | > 58 | 133 |
|  | RVEF | 52-73 | 49-51 | < 49 | 83 | 49-68 | 45-48 | < 45 | 133 |
|  | RVCOi | 2-4.1 | 1.8-1.9 | < 1.8 | 83 | 2.1-4.4 | 1.9-2 | < 1.9 | 133 |
|  | RVSVi | 32-61 | 29-31 | < 29 | 83 | 34-70 | 29-33 | < 29 | 133 |
|  | LAVi max | 22-52 | 53-58 | > 58 | 77 | 21-58 | 59-65 | > 65 | 121 |
|  | LAEF | 54-80 | 51-53 | < 51 | 75 | 55-82 | 52-54 | < 52 | 120 |
|  | RAVi max | 22-53 | 54-58 | > 58 | 76 | 23-69 | 70-79 | > 79 | 128 |
|  | RAEF | 37-69 | 34-36 | < 34 | 76 | 34-65 | 29-33 | < 29 | 127 |
| 60-69 | LVEDVi | 50-91 | 92-101 | > 101 | 63 | 51-106 | 107-116 | > 116 | 118 |
|  | LVESVi | 15-40 | 41-48 | > 48 | 63 | 17-47 | 48-54 | > 54 | 118 |
|  | LVSVi | 31-60 | 29-30 | < 29 | 63 | 31-68 | 27-30 | < 27 | 118 |
|  | LVEF | 53-75 | 50-52 | < 50 | 63 | 50-73 | 47-49 | < 47 | 118 |
|  | LVCOi | 1.9-4.3 | 1.7-1.8 | < 1.7 | 63 | 1.9-4.5 | 1.6-1.8 | < 1.6 | 119 |
|  | LVMi | 30-58 | 59-66 | > 66 | 63 | 37-74 | 75-86 | > 86 | 119 |
|  | LVM/LVEDV | 0.5-0.8 |  | > 0.8 | 63 | 0.5-0.9 | 01-1 | > 1 | 118 |
|  | RVEDVi | 50-98 | 99-111 | > 111 | 63 | 53-113 | 114-123 | > 123 | 119 |
|  | RVESVi | 15-38 | 39-44 | > 44 | 63 | 19-51 | 52-58 | > 58 | 119 |
|  | RVEF | 54-73 | 51-53 | < 51 | 63 | 47-70 | 42-46 | < 42 | 119 |
|  | RVCOi | 1.9-4.4 | 1.7-1.8 | < 1.7 | 63 | 1.8-4.4 | 1.5-1.7 | < 1.5 | 119 |
|  | RVSVi | 31-63 | 28-30 | < 28 | 63 | 31-67 | 25-30 | < 25 | 119 |
|  | LAVi max | 22-54 | 55-60 | > 60 | 59 | 21-59 | 60-66 | > 66 | 110 |
|  | LAEF | 50-78 | 46-49 | < 46 | 58 | 51-80 | 48-50 | < 48 | 107 |
|  | RAVi max | 21-61 | 62-68 | > 68 | 61 | 23-68 | 69-77 | > 77 | 113 |
|  | RAEF | 36-68 | 31-35 | < 31 | 59 | 35-70 | 31-34 | < 31 | 114 |
| 70+ | LVEDVi | 48-94 | 95-108 | > 108 | 28 | 44-102 | 103-111 | > 111 | 41 |
|  | LVESVi | 15-41 | 42-52 | > 52 | 28 | 15-45 | 46-53 | > 53 | 41 |
|  | LVSVi | 30-61 | 28-29 | < 28 | 28 | 32-64 | 29-31 | < 29 | 40 |
|  | LVEF | 54-77 | 51-53 | < 51 | 28 | 51-75 | 47-50 | < 47 | 41 |
|  | LVCOi | 1.8-4.4 | 1.6-1.7 | < 1.6 | 28 | 1.6-4.4 | 1.1-1.5 | < 1.1 | 41 |
|  | LVMi | 30-62 | 63-74 | > 74 | 28 | 31-68 | 69-74 | > 74 | 41 |
|  | LVM/LVEDV | 0.5-0.8 |  | > 0.8 | 28 | 0.5-1 | 1.1-1.2 | > 1.2 | 41 |
|  | RVEDVi | 49-100 | 101-115 | > 115 | 28 | 45-108 | 109-116 | > 116 | 41 |
|  | RVESVi | 15-37 | 38-42 | > 42 | 28 | 18-51 | 52-59 | > 59 | 41 |
|  | RVEF | 55-74 | 52-54 | < 52 | 28 | 46-72 | 39-45 | < 39 | 40 |
|  | RVCOi | 1.8-4.7 | 1.6-1.7 | < 1.6 | 28 | 1.4-4.4 | 0.9-1.3 | < 0.9 | 41 |
|  | RVSVi | 30-65 | 26-29 | < 26 | 28 | 28-65 | 21-27 | < 21 | 40 |
|  | LAVi max | 22-55 | 56-63 | > 63 | 28 | 21-60 | 61-67 | > 67 | 34 |
|  | LAEF | 45-76 | 40-44 | < 40 | 27 | 48-79 | 45-47 | < 45 | 34 |
|  | RAVi max | 17-65 | 66-76 | > 76 | 27 | 22-67 | 68-76 | > 76 | 38 |
|  | RAEF | 35-70 | 25-34 | < 25 | 27 | 36-77 | 32-35 | < 32 | 39 |

Footnote: All measurements were derived using cvi42 software with smooth endocardial segmentation; papillary muscles and trabeculations were excluded from left ventricular mass measurements. Volumetric and mass measurements were indexed to height. Reference ranges are defined as the interval between the 2.5th and 97.5th centiles derived from age-specific generalized additive models for location, scale and shape (GAMLSS) using the Box–Cox Cole–Green distribution. Centile-based categories beyond the reference range are derived from LMS-based thresholds (P0.5, P2.5, P97.5, and P99.5) and describe the position of a measurement within the reference distribution rather than disease severity or diagnostic abnormality. Reference ranges are based on an Asian adult population.

Abbreviations: LV = left ventricle; RV = right ventricle; LA = left atrium; RA = right atrium; EDV = end-diastolic volume; ESV = end-systolic volume; SV = stroke volume; EF = ejection fraction; CO = cardiac output.

### 4. Table: Reference ranges and centile-based categories for cardiac chamber measurements derived using cvi42 software (smooth segmentation), indexed to height, in a Chinese adult population

|  |  | **Female** | | | | **Male** | | | |
| --- | --- | --- | --- | --- | --- | --- | --- | --- | --- |
| **Age group** | **Metrics** | **Reference range** | **Mild** | **At least moderate** | **n** | **Reference range** | **Mild** | **At least moderate** | **n** |
| 18-29 | LVEDVi | 56-99 | 100-109 | > 109 | 16 | 66-109 | 110-113 | > 113 | 19 |
|  | LVESVi | 18-42 | 43-49 | > 49 | 16 | 25-48 | 49-51 | > 51 | 19 |
|  | LVSVi | 33-69 | 30-32 | < 30 | 16 | 37-66 | 29-36 | < 29 | 19 |
|  | LVEF | 52-73 | 48-51 | < 48 | 16 | 50-67 | 48-49 | < 48 | 19 |
|  | LVCOi | 2-4.8 | 1.8-1.9 | < 1.8 | 16 | 2.6-4.9 | 2.4-2.5 | < 2.4 | 19 |
|  | LVMi | 27-47 | 48-51 | > 51 | 16 | 36-69 | 70-73 | > 73 | 19 |
|  | LVM/LVEDV | 0.4-0.6 |  | > 0.6 | 16 | 0.5-0.7 | 0.8-0.8 | > 0.8 | 19 |
|  | RVEDVi | 50-101 | 102-109 | > 109 | 16 | 72-121 | 122-126 | > 126 | 19 |
|  | RVESVi | 15-44 | 45-46 | > 46 | 16 | 27-66 | 67-72 | > 72 | 19 |
|  | RVEF | 47-71 | 44-46 | < 44 | 16 | 40-62 | 35-39 | < 35 | 19 |
|  | RVCOi | 1.9-4.7 | 1.6-1.8 | < 1.6 | 16 | 2.5-4.9 | 2.2-2.4 | < 2.2 | 19 |
|  | RVSVi | 31-63 | 27-30 | < 27 | 16 | 41-68 | 37-40 | < 37 | 19 |
|  | LAVi max | 22-52 | 53-58 | > 58 | 13 | 27-52 | 53-56 | > 56 | 16 |
|  | LAEF | 54-77 | 50-53 | < 50 | 13 | 52-70 | 49-51 | < 49 | 16 |
|  | RAVi max | 19-48 | 49-57 | > 57 | 15 | 20-62 | 63-69 | > 69 | 18 |
|  | RAEF | 41-73 | 35-40 | < 35 | 15 | 38-68 | 31-37 | < 31 | 18 |
| 30-39 | LVEDVi | 55-97 | 98-107 | > 107 | 18 | 64-107 | 108-112 | > 112 | 17 |
|  | LVESVi | 18-41 | 42-48 | > 48 | 18 | 22-47 | 48-50 | > 50 | 17 |
|  | LVSVi | 34-65 | 31-33 | < 31 | 18 | 36-66 | 30-35 | < 30 | 17 |
|  | LVEF | 52-73 | 48-51 | < 48 | 18 | 50-68 | 48-49 | < 48 | 17 |
|  | LVCOi | 2-4.5 | 1.8-1.9 | < 1.8 | 18 | 2.5-4.8 | 2.3-2.4 | < 2.3 | 17 |
|  | LVMi | 28-49 | 50-54 | > 54 | 18 | 37-70 | 71-75 | > 75 | 17 |
|  | LVM/LVEDV | 0.4-0.6 |  | > 0.6 | 18 | 0.5-0.8 |  | > 0.8 | 17 |
|  | RVEDVi | 52-100 | 101-110 | > 110 | 18 | 72-119 | 120-125 | > 125 | 17 |
|  | RVESVi | 10-44 | 45-47 | > 47 | 18 | 25-63 | 64-68 | > 68 | 17 |
|  | RVEF | 48-71 | 45-47 | < 45 | 18 | 42-64 | 38-41 | < 38 | 17 |
|  | RVCOi | 1.9-4.4 | 1.7-1.8 | < 1.7 | 18 | 2.4-4.8 | 2.1-2.3 | < 2.1 | 17 |
|  | RVSVi | 31-62 | 28-30 | < 28 | 18 | 38-67 | 34-37 | < 34 | 17 |
|  | LAVi max | 22-51 | 52-57 | > 57 | 18 | 26-52 | 53-56 | > 56 | 14 |
|  | LAEF | 53-76 | 50-52 | < 50 | 18 | 51-72 | 48-50 | < 48 | 14 |
|  | RAVi max | 20-51 | 52-61 | > 61 | 18 | 20-63 | 64-71 | > 71 | 16 |
|  | RAEF | 41-72 | 35-40 | < 35 | 18 | 38-68 | 31-37 | < 31 | 16 |
| 40-49 | LVEDVi | 54-95 | 96-105 | > 105 | 24 | 62-105 | 106-111 | > 111 | 20 |
|  | LVESVi | 17-40 | 41-47 | > 47 | 24 | 20-46 | 47-49 | > 49 | 20 |
|  | LVSVi | 35-62 | 32-34 | < 32 | 24 | 36-65 | 31-35 | < 31 | 20 |
|  | LVEF | 52-74 | 48-51 | < 48 | 24 | 50-69 | 47-49 | < 47 | 19 |
|  | LVCOi | 2-4.3 | 1.8-1.9 | < 1.8 | 24 | 2.4-4.7 | 2.2-2.3 | < 2.2 | 20 |
|  | LVMi | 29-52 | 53-58 | > 58 | 24 | 38-71 | 72-76 | > 76 | 20 |
|  | LVM/LVEDV | 0.4-0.7 |  | > 0.7 | 24 | 0.5-0.8 | 0.9-0.9 | > 0.9 | 20 |
|  | RVEDVi | 52-100 | 101-110 | > 110 | 24 | 70-118 | 119-126 | > 126 | 20 |
|  | RVESVi | 14-44 | 45-50 | > 50 | 24 | 22-59 | 60-64 | > 64 | 20 |
|  | RVEF | 50-72 | 46-49 | < 46 | 23 | 44-66 | 41-43 | < 41 | 19 |
|  | RVCOi | 1.9-4.2 | 1.7-1.8 | < 1.7 | 24 | 2.2-4.7 | 2-2.1 | < 2 | 20 |
|  | RVSVi | 31-61 | 28-30 | < 28 | 24 | 36-67 | 31-35 | < 31 | 20 |
|  | LAVi max | 22-51 | 52-56 | > 56 | 22 | 24-52 | 53-56 | > 56 | 18 |
|  | LAEF | 53-75 | 49-52 | < 49 | 22 | 51-74 | 47-50 | < 47 | 18 |
|  | RAVi max | 22-54 | 55-65 | > 65 | 23 | 21-65 | 66-72 | > 72 | 19 |
|  | RAEF | 41-70 | 36-40 | < 36 | 23 | 37-67 | 30-36 | < 30 | 19 |
| 50-59 | LVEDVi | 52-93 | 94-103 | > 103 | 66 | 60-104 | 105-111 | > 111 | 49 |
|  | LVESVi | 16-39 | 40-46 | > 46 | 66 | 17-45 | 46-48 | > 48 | 49 |
|  | LVSVi | 34-59 | 32-33 | < 32 | 66 | 35-64 | 31-34 | < 31 | 49 |
|  | LVEF | 53-75 | 48-52 | < 48 | 66 | 50-70 | 47-49 | < 47 | 48 |
|  | LVCOi | 2-4.1 | 1.8-1.9 | < 1.8 | 66 | 2.3-4.6 | 2.1-2.2 | < 2.1 | 49 |
|  | LVMi | 30-55 | 56-64 | > 64 | 66 | 39-71 | 72-78 | > 78 | 49 |
|  | LVM/LVEDV | 0.5-0.7 | 0.8-0.8 | > 0.8 | 66 | 0.5-0.8 | 0.9-0.9 | > 0.9 | 49 |
|  | RVEDVi | 51-97 | 98-108 | > 108 | 66 | 67-117 | 118-128 | > 128 | 49 |
|  | RVESVi | 16-43 | 44-50 | > 50 | 66 | 20-55 | 56-60 | > 60 | 49 |
|  | RVEF | 51-73 | 47-50 | < 47 | 66 | 46-68 | 43-45 | < 43 | 49 |
|  | RVCOi | 1.9-4 | 1.7-1.8 | < 1.7 | 66 | 2.1-4.7 | 1.8-2 | < 1.8 | 49 |
|  | RVSVi | 31-60 | 28-30 | < 28 | 66 | 33-67 | 28-32 | < 28 | 49 |
|  | LAVi max | 21-50 | 51-56 | > 56 | 60 | 23-51 | 52-55 | > 55 | 40 |
|  | LAEF | 51-76 | 48-50 | < 48 | 58 | 50-77 | 46-49 | < 46 | 40 |
|  | RAVi max | 23-57 | 58-69 | > 69 | 63 | 21-66 | 67-74 | > 74 | 46 |
|  | RAEF | 40-68 | 36-39 | < 36 | 63 | 37-66 | 30-36 | < 30 | 46 |
| 60-69 | LVEDVi | 51-91 | 92-101 | > 101 | 52 | 58-103 | 104-113 | > 113 | 37 |
|  | LVESVi | 15-37 | 38-45 | > 45 | 52 | 14-43 | 44-47 | > 47 | 37 |
|  | LVSVi | 32-59 | 29-31 | < 29 | 52 | 34-64 | 31-33 | < 31 | 37 |
|  | LVEF | 53-75 | 48-52 | < 48 | 52 | 50-71 | 47-49 | < 47 | 35 |
|  | LVCOi | 2-3.9 | 1.8-1.9 | < 1.8 | 52 | 2.1-4.6 | 02-2 | < 2 | 37 |
|  | LVMi | 31-59 | 60-74 | > 74 | 52 | 39-72 | 73-81 | > 81 | 37 |
|  | LVM/LVEDV | 0.5-0.8 | 0.9-1 | > 1 | 52 | 0.5-0.9 | 01-1 | > 1 | 37 |
|  | RVEDVi | 49-92 | 93-101 | > 101 | 52 | 63-115 | 116-130 | > 130 | 36 |
|  | RVESVi | 15-38 | 39-43 | > 43 | 52 | 17-51 | 52-55 | > 55 | 37 |
|  | RVEF | 53-74 | 49-52 | < 49 | 52 | 47-71 | 45-46 | < 45 | 36 |
|  | RVCOi | 1.9-3.8 | 1.7-1.8 | < 1.7 | 52 | 1.9-4.6 | 1.7-1.8 | < 1.7 | 37 |
|  | RVSVi | 31-59 | 27-30 | < 27 | 52 | 30-67 | 24-29 | < 24 | 37 |
|  | LAVi max | 21-51 | 52-56 | > 56 | 47 | 20-49 | 50-53 | > 53 | 30 |
|  | LAEF | 49-76 | 45-48 | < 45 | 46 | 48-80 | 44-47 | < 44 | 30 |
|  | RAVi max | 24-61 | 62-73 | > 73 | 46 | 22-68 | 69-76 | > 76 | 35 |
|  | RAEF | 40-66 | 36-39 | < 36 | 46 | 38-68 | 31-37 | < 31 | 35 |
| 70+ | LVEDVi | 50-89 | 90-100 | > 100 | 12 |  |  |  |  |
|  | LVESVi | 14-36 | 37-44 | > 44 | 12 |  |  |  |  |
|  | LVSVi | 28-60 | 23-27 | < 23 | 12 |  |  |  |  |
|  | LVEF | 53-76 | 47-52 | < 47 | 12 |  |  |  |  |
|  | LVCOi | 1.9-3.8 |  | < 1.8 | 12 |  |  |  |  |
|  | LVMi | 32-64 | 65-91 | > 91 | 12 |  |  |  |  |
|  | LVM/LVEDV | 0.5-0.9 | 1-1.3 | > 1.3 | 12 |  |  |  |  |
|  | RVEDVi | 46-86 | 87-93 | > 93 | 12 |  |  |  |  |
|  | RVESVi | 13-33 | 34-36 | > 36 | 12 |  |  |  |  |
|  | RVEF | 54-74 | 50-53 | < 50 | 12 |  |  |  |  |
|  | RVCOi | 1.9-3.7 | 1.7-1.8 | < 1.7 | 12 |  |  |  |  |
|  | RVSVi | 31-58 | 27-30 | < 27 | 12 |  |  |  |  |
|  | LAVi max | 20-51 | 52-57 | > 57 | 11 |  |  |  |  |
|  | LAEF | 47-78 | 42-46 | < 42 | 12 |  |  |  |  |
|  | RAVi max | 25-64 | 65-76 | > 76 | 11 |  |  |  |  |
|  | RAEF | 39-65 | 36-38 | < 36 | 11 |  |  |  |  |

Footnote: All measurements were derived using cvi42 software with smooth endocardial segmentation; papillary muscles and trabeculations were excluded from left ventricular mass measurements. Volumetric and mass measurements were indexed to height. Reference ranges are defined as the interval between the 2.5th and 97.5th centiles derived from age-specific generalized additive models for location, scale and shape (GAMLSS) using the Box–Cox Cole–Green distribution. Centile-based categories beyond the reference range are derived from LMS-based thresholds (P0.5, P2.5, P97.5, and P99.5) and describe the position of a measurement within the reference distribution rather than disease severity or diagnostic abnormality. Reference ranges are based on a White adult population.

Abbreviations: LV = left ventricle; RV = right ventricle; LA = left atrium; RA = right atrium; EDV = end-diastolic volume; ESV = end-systolic volume; SV = stroke volume; EF = ejection fraction; CO = cardiac output.

### 5. Table: Reference ranges and centile-based categories for cardiac chamber measurements derived using cvi42 software (smooth segmentation), indexed to height, in a Mixed adult population

|  |  | **Female** | | | | **Male** | | | |
| --- | --- | --- | --- | --- | --- | --- | --- | --- | --- |
| **Age group** | **Metrics** | **Reference range** | **Mild** | **At least moderate** | **n** | **Reference range** | **Mild** | **At least moderate** | **n** |
| 50-59 | LVEDVi | 57-105 | 106-114 | > 114 | 134 | 66-117 | 118-122 | > 122 | 65 |
|  | LVESVi | 18-42 | 43-47 | > 47 | 134 | 20-53 | 54-58 | > 58 | 66 |
|  | LVSVi | 35-73 | 32-34 | < 32 | 134 | 38-75 | 32-37 | < 32 | 65 |
|  | LVEF | 53-74 | 49-52 | < 49 | 134 | 50-71 | 46-49 | < 46 | 66 |
|  | LVCOi | 2.1-5 | 1.9-2 | < 1.9 | 132 | 2.2-5.3 | 1.9-2.1 | < 1.9 | 66 |
|  | LVMi | 31-64 | 65-73 | > 73 | 134 | 41-85 | 86-94 | > 94 | 65 |
|  | LVM/LVEDV | 0.5-0.7 | 0.8-0.8 | > 0.8 | 134 | 0.5-0.8 | 0.9-0.9 | > 0.9 | 64 |
|  | RVEDVi | 55-111 | 112-122 | > 122 | 134 | 68-132 | 133-141 | > 141 | 65 |
|  | RVESVi | 17-44 | 45-49 | > 49 | 134 | 26-55 | 56-60 | > 60 | 65 |
|  | RVEF | 53-72 | 50-52 | < 50 | 134 | 51-70 | 48-50 | < 48 | 66 |
|  | RVCOi | 2-4.9 | 1.8-1.9 | < 1.8 | 133 | 2.3-5.4 | 2-2.2 | < 2 | 66 |
|  | RVSVi | 34-67 | 29-33 | < 29 | 133 | 39-81 | 32-38 | < 32 | 66 |
|  | LAVi max | 23-55 | 56-60 | > 60 | 121 | 25-70 | 71-79 | > 79 | 53 |
|  | LAEF | 55-78 | 52-54 | < 52 | 120 | 54-78 | 51-53 | < 51 | 53 |
|  | RAVi max | 24-66 | 67-75 | > 75 | 128 | 26-79 | 80-88 | > 88 | 64 |
|  | RAEF | 37-64 | 31-36 | < 31 | 124 | 35-63 | 31-34 | < 31 | 64 |
| 60-69 | LVEDVi | 53-101 | 102-111 | > 111 | 110 | 61-114 | 115-122 | > 122 | 59 |
|  | LVESVi | 14-41 | 42-46 | > 46 | 110 | 19-50 | 51-54 | > 54 | 58 |
|  | LVSVi | 33-65 | 30-32 | < 30 | 110 | 37-71 | 32-36 | < 32 | 59 |
|  | LVEF | 52-74 | 48-51 | < 48 | 108 | 49-72 | 45-48 | < 45 | 58 |
|  | LVCOi | 2-4.5 | 1.9-1.9 | < 1.9 | 110 | 2.2-4.9 | 2-2.1 | < 2 | 59 |
|  | LVMi | 31-60 | 61-65 | > 65 | 110 | 41-83 | 84-93 | > 93 | 59 |
|  | LVM/LVEDV | 0.5-0.8 |  | > 0.8 | 110 | 0.5-0.9 | 01-1 | > 1 | 59 |
|  | RVEDVi | 51-108 | 109-119 | > 119 | 110 | 63-127 | 128-138 | > 138 | 58 |
|  | RVESVi | 17-43 | 44-49 | > 49 | 110 | 20-55 | 56-60 | > 60 | 58 |
|  | RVEF | 53-72 | 49-52 | < 49 | 110 | 49-69 | 47-48 | < 47 | 58 |
|  | RVCOi | 1.9-4.5 | 1.7-1.8 | < 1.7 | 110 | 2.3-5.1 | 2-2.2 | < 2 | 59 |
|  | RVSVi | 31-69 | 27-30 | < 27 | 110 | 36-77 | 31-35 | < 31 | 59 |
|  | LAVi max | 23-58 | 59-65 | > 65 | 100 | 24-65 | 66-74 | > 74 | 52 |
|  | LAEF | 52-79 | 49-51 | < 49 | 98 | 50-79 | 47-49 | < 47 | 52 |
|  | RAVi max | 23-68 | 69-77 | > 77 | 103 | 20-78 | 79-85 | > 85 | 57 |
|  | RAEF | 35-68 | 29-34 | < 29 | 103 | 36-68 | 33-35 | < 33 | 57 |
| 70+ | LVEDVi | 52-94 | 95-102 | > 102 | 51 | 57-114 | 115-127 | > 127 | 33 |
|  | LVESVi | 10-40 | 41-44 | > 44 | 51 | 18-47 | 48-51 | > 51 | 32 |
|  | LVSVi | 31-59 | 28-30 | < 28 | 50 | 35-67 | 31-34 | < 31 | 34 |
|  | LVEF | 51-74 | 47-50 | < 47 | 51 | 48-72 | 45-47 | < 45 | 34 |
|  | LVCOi | 2-4.1 | 1.8-1.9 | < 1.8 | 51 | 2.1-4.6 | 02-2 | < 2 | 35 |
|  | LVMi | 31-56 | 57-61 | > 61 | 51 | 41-82 | 83-92 | > 92 | 35 |
|  | LVM/LVEDV | 0.5-0.7 | 0.8-0.8 | > 0.8 | 51 | 0.5-1 | 1.1-1.1 | > 1.1 | 33 |
|  | RVEDVi | 56-92 | 93-99 | > 99 | 51 | 58-123 | 124-135 | > 135 | 35 |
|  | RVESVi | 17-41 | 42-47 | > 47 | 51 | 15-55 | 56-59 | > 59 | 35 |
|  | RVEF | 52-72 | 48-51 | < 48 | 51 | 48-68 | 46-47 | < 46 | 35 |
|  | RVCOi | 1.9-4.1 | 1.7-1.8 | < 1.7 | 51 | 2.1-4.7 | 1.9-2 | < 1.9 | 35 |
|  | RVSVi | 35-60 | 32-34 | < 32 | 50 | 33-74 | 28-32 | < 28 | 35 |
|  | LAVi max | 24-62 | 63-72 | > 72 | 45 | 22-61 | 62-70 | > 70 | 32 |
|  | LAEF | 50-80 | 47-49 | < 47 | 44 | 46-81 | 42-45 | < 42 | 30 |
|  | RAVi max | 21-68 | 69-77 | > 77 | 47 | 18-76 | 77-83 | > 83 | 31 |
|  | RAEF | 33-72 | 28-32 | < 28 | 46 | 36-76 | 34-35 | < 34 | 31 |

Footnote: All measurements were derived using cvi42 software with smooth endocardial segmentation; papillary muscles and trabeculations were excluded from left ventricular mass measurements. Volumetric and mass measurements were indexed to height. Reference ranges are defined as the interval between the 2.5th and 97.5th centiles derived from age-specific generalized additive models for location, scale and shape (GAMLSS) using the Box–Cox Cole–Green distribution. Centile-based categories beyond the reference range are derived from LMS-based thresholds (P0.5, P2.5, P97.5, and P99.5) and describe the position of a measurement within the reference distribution rather than disease severity or diagnostic abnormality. Reference ranges are based on a Mixed adult population.

Abbreviations: LV = left ventricle; RV = right ventricle; LA = left atrium; RA = right atrium; EDV = end-diastolic volume; ESV = end-systolic volume; SV = stroke volume; EF = ejection fraction; CO = cardiac output.

### 6. Table: Reference ranges and centile-based categories for cardiac chamber measurements derived using cvi42 software (anatomical segmentation), indexed to height, in a White adult population

|  |  | **Female** | | | | | **Male** | | | |
| --- | --- | --- | --- | --- | --- | --- | --- | --- | --- | --- |
| **Age group** | **Metrics** | **Reference range** | **Mild** | **At least moderate** | **n** | **Reference range** | | **Mild** | **At least moderate** | **n** |
| 18-29 | LVEDVi | 54-97 | 98-104 | > 104 | 102 | 67-122 | | 123-130 | > 130 | 103 |
|  | LVESVi | 14-44 | 45-49 | > 49 | 102 | 19-53 | | 54-60 | > 60 | 104 |
|  | LVSVi | 34-65 | 29-33 | < 29 | 102 | 40-78 | | 34-39 | < 34 | 101 |
|  | LVEF | 55-74 | 52-54 | < 52 | 100 | 51-72 | | 48-50 | < 48 | 102 |
|  | LVCOi | 2.3-4.9 | 2-2.2 | < 2 | 102 | 2.3-5.4 | | 1.7-2.2 | < 1.7 | 101 |
|  | LVMi | 40-67 | 68-73 | > 73 | 102 | 52-91 | | 92-97 | > 97 | 104 |
|  | LVM/LVEDV | 0.6-0.9 | 01-1 | > 1 | 102 | 0.6-1 | | 1.1-1.1 | > 1.1 | 102 |
|  | RVEDVi | 54-105 | 106-112 | > 112 | 102 | 68-134 | | 135-142 | > 142 | 102 |
|  | RVESVi | 13-49 | 50-53 | > 53 | 100 | 25-65 | | 66-71 | > 71 | 99 |
|  | RVEF | 47-71 | 44-46 | < 44 | 97 | 45-68 | | 41-44 | < 41 | 97 |
|  | RVCOi | 1.9-5.2 | 1.6-1.8 | < 1.6 | 101 | 2.3-5.5 | | 1.7-2.2 | < 1.7 | 98 |
|  | RVSVi | 31-67 | 26-30 | < 26 | 102 | 39-81 | | 32-38 | < 32 | 99 |
|  | LAVi max | 25-51 | 52-55 | > 55 | 50 | 28-63 | | 64-70 | > 70 | 52 |
|  | LAEF | 56-75 | 54-55 | < 54 | 50 | 53-72 | | 50-52 | < 50 | 52 |
|  | RAVi max | 21-51 | 52-56 | > 56 | 80 | 28-68 | | 69-77 | > 77 | 82 |
|  | RAEF | 38-78 | 33-37 | < 33 | 78 | 35-74 | | 31-34 | < 31 | 82 |
| 30-39 | LVEDVi | 53-96 | 97-103 | > 103 | 137 | 61-115 | | 116-123 | > 123 | 134 |
|  | LVESVi | 14-42 | 43-47 | > 47 | 136 | 17-51 | | 52-57 | > 57 | 134 |
|  | LVSVi | 33-65 | 29-32 | < 29 | 135 | 37-74 | | 31-36 | < 31 | 133 |
|  | LVEF | 55-75 | 53-54 | < 53 | 132 | 52-74 | | 48-51 | < 48 | 134 |
|  | LVCOi | 2.3-4.9 | 2-2.2 | < 2 | 136 | 2.3-5.2 | | 1.8-2.2 | < 1.8 | 128 |
|  | LVMi | 39-68 | 69-74 | > 74 | 137 | 49-88 | | 89-93 | > 93 | 134 |
|  | LVM/LVEDV | 0.6-0.9 | 1-1.1 | > 1.1 | 137 | 0.6-1 | | 1.1-1.2 | > 1.2 | 135 |
|  | RVEDVi | 54-105 | 106-113 | > 113 | 137 | 68-135 | | 136-143 | > 143 | 131 |
|  | RVESVi | 15-49 | 50-53 | > 53 | 135 | 25-65 | | 66-70 | > 70 | 129 |
|  | RVEF | 48-70 | 45-47 | < 45 | 127 | 45-68 | | 42-44 | < 42 | 131 |
|  | RVCOi | 2-5.1 | 1.7-1.9 | < 1.7 | 134 | 2.3-5.5 | | 1.8-2.2 | < 1.8 | 125 |
|  | RVSVi | 31-67 | 27-30 | < 27 | 135 | 37-79 | | 31-36 | < 31 | 132 |
|  | LAVi max | 25-55 | 56-60 | > 60 | 84 | 27-64 | | 65-71 | > 71 | 66 |
|  | LAEF | 55-76 | 53-54 | < 53 | 83 | 53-73 | | 50-52 | < 50 | 66 |
|  | RAVi max | 21-52 | 53-57 | > 57 | 113 | 27-69 | | 70-77 | > 77 | 119 |
|  | RAEF | 38-75 | 33-37 | < 33 | 110 | 35-72 | | 31-34 | < 31 | 118 |
| 40-49 | LVEDVi | 53-98 | 99-106 | > 106 | 518 | 61-117 | | 118-126 | > 126 | 438 |
|  | LVESVi | 13-39 | 40-45 | > 45 | 518 | 16-48 | | 49-54 | > 54 | 438 |
|  | LVSVi | 34-67 | 30-33 | < 30 | 518 | 37-76 | | 31-36 | < 31 | 435 |
|  | LVEF | 56-76 | 53-55 | < 53 | 512 | 53-75 | | 49-52 | < 49 | 434 |
|  | LVCOi | 2.2-4.8 | 1.9-2.1 | < 1.9 | 515 | 2.3-5.1 | | 1.8-2.2 | < 1.8 | 429 |
|  | LVMi | 39-69 | 70-76 | > 76 | 518 | 50-90 | | 91-96 | > 96 | 429 |
|  | LVM/LVEDV | 0.6-0.9 | 1-1.1 | > 1.1 | 518 | 0.6-1.1 | |  | > 1.2 | 426 |
|  | RVEDVi | 56-109 | 110-118 | > 118 | 517 | 68-135 | | 136-144 | > 144 | 430 |
|  | RVESVi | 16-49 | 50-53 | > 53 | 517 | 24-64 | | 65-69 | > 69 | 422 |
|  | RVEF | 49-70 | 46-48 | < 46 | 508 | 45-68 | | 42-44 | < 42 | 431 |
|  | RVCOi | 2.1-4.9 | 1.8-2 | < 1.8 | 514 | 2.3-5.4 | | 1.8-2.2 | < 1.8 | 426 |
|  | RVSVi | 33-69 | 28-32 | < 28 | 516 | 37-80 | | 31-36 | < 31 | 432 |
|  | LAVi max | 25-59 | 60-65 | > 65 | 440 | 25-64 | | 65-72 | > 72 | 358 |
|  | LAEF | 55-78 | 52-54 | < 52 | 434 | 53-76 | | 50-52 | < 50 | 353 |
|  | RAVi max | 23-59 | 60-66 | > 66 | 500 | 28-77 | | 78-87 | > 87 | 408 |
|  | RAEF | 38-72 | 33-37 | < 33 | 497 | 35-69 | | 31-34 | < 31 | 410 |
| 50-59 | LVEDVi | 53-100 | 101-108 | > 108 | 911 | 58-117 | | 118-126 | > 126 | 831 |
|  | LVESVi | 12-37 | 38-42 | > 42 | 911 | 15-46 | | 47-52 | > 52 | 835 |
|  | LVSVi | 35-68 | 31-34 | < 31 | 911 | 37-76 | | 30-36 | < 30 | 822 |
|  | LVEF | 57-78 | 53-56 | < 53 | 906 | 54-77 | | 50-53 | < 50 | 833 |
|  | LVCOi | 2.1-4.6 | 1.8-2 | < 1.8 | 908 | 2.3-4.9 | | 1.9-2.2 | < 1.9 | 816 |
|  | LVMi | 39-72 | 73-79 | > 79 | 910 | 50-93 | | 94-100 | > 100 | 810 |
|  | LVM/LVEDV | 0.6-1 | 1.1-1.1 | > 1.1 | 910 | 0.7-1.2 | | 1.3-1.3 | > 1.3 | 806 |
|  | RVEDVi | 58-112 | 113-122 | > 122 | 909 | 67-133 | | 134-143 | > 143 | 820 |
|  | RVESVi | 17-48 | 49-52 | > 52 | 909 | 24-61 | | 62-67 | > 67 | 810 |
|  | RVEF | 51-71 | 47-50 | < 47 | 906 | 46-68 | | 43-45 | < 43 | 830 |
|  | RVCOi | 2.1-4.7 | 1.8-2 | < 1.8 | 906 | 2.2-5.1 | | 1.8-2.1 | < 1.8 | 819 |
|  | RVSVi | 34-71 | 29-33 | < 29 | 909 | 36-79 | | 29-35 | < 29 | 828 |
|  | LAVi max | 24-62 | 63-69 | > 69 | 797 | 24-65 | | 66-73 | > 73 | 684 |
|  | LAEF | 53-79 | 50-52 | < 50 | 791 | 53-79 | | 50-52 | < 50 | 681 |
|  | RAVi max | 24-66 | 67-74 | > 74 | 854 | 27-78 | | 79-87 | > 87 | 761 |
|  | RAEF | 38-70 | 33-37 | < 33 | 844 | 35-68 | | 31-34 | < 31 | 771 |
| 60-69 | LVEDVi | 50-95 | 96-105 | > 105 | 918 | 54-113 | | 114-123 | > 123 | 803 |
|  | LVESVi | 11-34 | 35-39 | > 39 | 920 | 13-44 | | 45-49 | > 49 | 801 |
|  | LVSVi | 33-65 | 29-32 | < 29 | 918 | 35-75 | | 29-34 | < 29 | 799 |
|  | LVEF | 58-79 | 54-57 | < 54 | 917 | 54-78 | | 50-53 | < 50 | 803 |
|  | LVCOi | 2-4.5 | 1.8-1.9 | < 1.8 | 918 | 2.2-4.8 | | 1.8-2.1 | < 1.8 | 796 |
|  | LVMi | 39-73 | 74-80 | > 80 | 919 | 49-93 | | 94-100 | > 100 | 789 |
|  | LVM/LVEDV | 0.6-1 | 1.1-1.2 | > 1.2 | 918 | 0.7-1.2 | | 1.3-1.4 | > 1.4 | 787 |
|  | RVEDVi | 55-107 | 108-118 | > 118 | 917 | 65-129 | | 130-140 | > 140 | 798 |
|  | RVESVi | 16-45 | 46-50 | > 50 | 920 | 22-58 | | 59-63 | > 63 | 795 |
|  | RVEF | 51-72 | 48-50 | < 48 | 914 | 47-69 | | 44-46 | < 44 | 800 |
|  | RVCOi | 2-4.6 | 1.8-1.9 | < 1.8 | 917 | 2.2-5 | | 1.7-2.1 | < 1.7 | 798 |
|  | RVSVi | 33-68 | 28-32 | < 28 | 918 | 35-78 | | 28-34 | < 28 | 797 |
|  | LAVi max | 22-63 | 64-71 | > 71 | 797 | 22-66 | | 67-74 | > 74 | 645 |
|  | LAEF | 51-79 | 47-50 | < 47 | 776 | 51-80 | | 47-50 | < 47 | 632 |
|  | RAVi max | 24-67 | 68-76 | > 76 | 855 | 25-79 | | 80-88 | > 88 | 742 |
|  | RAEF | 38-71 | 33-37 | < 33 | 841 | 35-70 | | 31-34 | < 31 | 748 |
| 70+ | LVEDVi | 47-92 | 93-103 | > 103 | 1101 | 50-108 | | 109-119 | > 119 | 1256 |
|  | LVESVi | 10-32 | 33-36 | > 36 | 1102 | 12-41 | | 42-47 | > 47 | 1257 |
|  | LVSVi | 32-64 | 29-31 | < 29 | 1101 | 33-71 | | 27-32 | < 27 | 1256 |
|  | LVEF | 58-80 | 54-57 | < 54 | 1095 | 55-79 | | 50-54 | < 50 | 1243 |
|  | LVCOi | 2-4.4 | 1.7-1.9 | < 1.7 | 1100 | 2.1-4.6 | | 1.8-2 | < 1.8 | 1256 |
|  | LVMi | 37-72 | 73-80 | > 80 | 1101 | 47-90 | | 91-98 | > 98 | 1251 |
|  | LVM/LVEDV | 0.6-1.1 |  | > 1.2 | 1100 | 0.7-1.2 | | 1.3-1.5 | > 1.5 | 1249 |
|  | RVEDVi | 53-104 | 105-115 | > 115 | 1100 | 62-124 | | 125-135 | > 135 | 1257 |
|  | RVESVi | 16-43 | 44-47 | > 47 | 1100 | 21-55 | | 56-59 | > 59 | 1257 |
|  | RVEF | 52-72 | 48-51 | < 48 | 1093 | 48-69 | | 44-47 | < 44 | 1244 |
|  | RVCOi | 2-4.5 | 1.7-1.9 | < 1.7 | 1099 | 2.1-4.7 | | 1.7-2 | < 1.7 | 1255 |
|  | RVSVi | 31-65 | 28-30 | < 28 | 1100 | 33-74 | | 26-32 | < 26 | 1256 |
|  | LAVi max | 21-63 | 64-73 | > 73 | 942 | 20-67 | | 68-75 | > 75 | 953 |
|  | LAEF | 48-79 | 44-47 | < 44 | 903 | 48-80 | | 44-47 | < 44 | 914 |
|  | RAVi max | 23-68 | 69-78 | > 78 | 1028 | 23-78 | | 79-87 | > 87 | 1119 |
|  | RAEF | 38-72 | 33-37 | < 33 | 1001 | 35-73 | | 30-34 | < 30 | 1115 |

Footnote: All measurements were derived using cvi42 software with anatomical endocardial segmentation; papillary muscles and trabeculations were included in left ventricular mass measurements. Volumetric and mass measurements were indexed to height. Reference ranges are defined as the interval between the 2.5th and 97.5th centiles derived from age-specific generalized additive models for location, scale and shape (GAMLSS) using the Box–Cox Cole–Green distribution. Centile-based categories beyond the reference range are derived from LMS-based thresholds (P0.5, P2.5, P97.5, and P99.5) and describe the position of a measurement within the reference distribution rather than disease severity or diagnostic abnormality. Reference ranges are based on a White adult population.

Abbreviations: LV = left ventricle; RV = right ventricle; LA = left atrium; RA = right atrium; EDV = end-diastolic volume; ESV = end-systolic volume; SV = stroke volume; EF = ejection fraction; CO = cardiac output.

### 7. Table: Reference ranges and centile-based categories for cardiac chamber measurements derived using cvi42 software (anatomical segmentation), indexed to height, in a Black adult population

|  |  | **Female** | | | | | **Male** | | | |
| --- | --- | --- | --- | --- | --- | --- | --- | --- | --- | --- |
| **Age group** | **Metrics** | **Reference range** | **Mild** | **At least moderate** | **n** | **Reference range** | | **Mild** | **At least moderate** | **n** |
| 40-49 | LVEDVi | 60-105 | 106-118 | > 118 | 10 |  | |  |  |  |
|  | LVESVi | 16-39 | 40-43 | > 43 | 10 |  | |  |  |  |
|  | LVSVi | 36-71 | 32-35 | < 32 | 10 |  | |  |  |  |
|  | LVEF | 57-78 | 54-56 | < 54 | 10 |  | |  |  |  |
|  | LVCOi | 2-5.1 | 1.8-1.9 | < 1.8 | 10 |  | |  |  |  |
|  | LVMi | 39-80 | 81-88 | > 88 | 10 |  | |  |  |  |
|  | LVM/LVEDV | 0.6-1 | 1.1-1.2 | > 1.2 | 10 |  | |  |  |  |
|  | RVEDVi | 64-122 | 123-136 | > 136 | 10 |  | |  |  |  |
|  | RVESVi | 22-57 | 58-66 | > 66 | 10 |  | |  |  |  |
|  | RVEF | 50-72 | 48-49 | < 48 | 10 |  | |  |  |  |
|  | RVCOi | 2.1-5.4 | 1.9-2 | < 1.9 | 10 |  | |  |  |  |
|  | RVSVi | 38-74 | 35-37 | < 35 | 10 |  | |  |  |  |
|  | LAVi max |  |  |  |  |  | |  |  |  |
|  | LAEF |  |  |  |  |  | |  |  |  |
|  | RAVi max | 28-76 | 77-91 | > 91 | 10 |  | |  |  |  |
|  | RAEF | 30-65 | 25-29 | < 25 | 10 |  | |  |  |  |
| 50-59 | LVEDVi | 56-101 | 102-111 | > 111 | 95 | 52-117 | | 118-127 | > 127 | 78 |
|  | LVESVi | 14-38 | 39-42 | > 42 | 95 | 14-48 | | 49-55 | > 55 | 78 |
|  | LVSVi | 35-69 | 31-34 | < 31 | 95 | 35-74 | | 29-34 | < 29 | 76 |
|  | LVEF | 57-79 | 54-56 | < 54 | 95 | 55-77 | | 52-54 | < 52 | 76 |
|  | LVCOi | 2-4.9 | 1.8-1.9 | < 1.8 | 94 | 2.4-5 | | 2.2-2.3 | < 2.2 | 77 |
|  | LVMi | 39-80 | 81-88 | > 88 | 95 | 47-96 | | 97-103 | > 103 | 75 |
|  | LVM/LVEDV | 0.6-1.1 |  | > 1.2 | 95 | 0.7-1.1 | |  | > 1.2 | 75 |
|  | RVEDVi | 61-117 | 118-128 | > 128 | 95 | 73-133 | | 134-147 | > 147 | 75 |
|  | RVESVi | 20-53 | 54-60 | > 60 | 95 | 28-60 | | 61-66 | > 66 | 74 |
|  | RVEF | 50-72 | 47-49 | < 47 | 95 | 46-68 | | 43-45 | < 43 | 76 |
|  | RVCOi | 2.1-5.1 | 1.9-2 | < 1.9 | 94 | 2.2-5.4 | | 1.9-2.1 | < 1.9 | 78 |
|  | RVSVi | 36-71 | 32-35 | < 32 | 95 | 33-79 | | 27-32 | < 27 | 77 |
|  | LAVi max | 25-63 | 64-71 | > 71 | 89 | 24-64 | | 65-72 | > 72 | 64 |
|  | LAEF | 54-75 | 51-53 | < 51 | 88 | 53-76 | | 50-52 | < 50 | 64 |
|  | RAVi max | 27-72 | 73-85 | > 85 | 94 | 33-74 | | 75-82 | > 82 | 73 |
|  | RAEF | 31-65 | 26-30 | < 26 | 93 | 35-63 | | 32-34 | < 32 | 71 |
| 60-69 | LVEDVi | 47-95 | 96-103 | > 103 | 66 | 52-113 | | 114-125 | > 125 | 48 |
|  | LVESVi | 11-36 | 37-41 | > 41 | 66 | 11-47 | | 48-54 | > 54 | 48 |
|  | LVSVi | 33-66 | 29-32 | < 29 | 66 | 37-74 | | 33-36 | < 33 | 48 |
|  | LVEF | 57-80 | 53-56 | < 53 | 66 | 45-78 | | 34-44 | < 34 | 48 |
|  | LVCOi | 2.1-4.6 | 1.8-2 | < 1.8 | 66 | 2.3-5 | | 2.1-2.2 | < 2.1 | 48 |
|  | LVMi | 39-80 | 81-88 | > 88 | 66 | 46-94 | | 95-101 | > 101 | 48 |
|  | LVM/LVEDV | 0.7-1.1 | 1.2-1.3 | > 1.3 | 66 | 0.7-1.2 | | 1.3-1.3 | > 1.3 | 48 |
|  | RVEDVi | 54-110 | 111-119 | > 119 | 66 | 70-130 | | 131-145 | > 145 | 48 |
|  | RVESVi | 16-49 | 50-53 | > 53 | 66 | 25-61 | | 62-69 | > 69 | 48 |
|  | RVEF | 49-71 | 45-48 | < 45 | 64 | 46-69 | | 42-45 | < 42 | 48 |
|  | RVCOi | 2.1-4.7 | 1.8-2 | < 1.8 | 66 | 2.3-5 | | 2.1-2.2 | < 2.1 | 48 |
|  | RVSVi | 33-67 | 28-32 | < 28 | 66 | 39-75 | | 35-38 | < 35 | 48 |
|  | LAVi max | 23-63 | 64-71 | > 71 | 59 | 23-65 | | 66-74 | > 74 | 42 |
|  | LAEF | 50-75 | 45-49 | < 45 | 58 | 50-75 | | 46-49 | < 46 | 43 |
|  | RAVi max | 25-67 | 68-76 | > 76 | 64 | 28-79 | | 80-88 | > 88 | 45 |
|  | RAEF | 32-67 | 27-31 | < 27 | 66 | 32-64 | | 29-31 | < 29 | 46 |
| 70+ | LVEDVi | 37-90 | 91-97 | > 97 | 18 | 51-110 | | 111-126 | > 126 | 14 |
|  | LVESVi | 9-34 | 35-40 | > 40 | 18 | 9-45 | | 46-53 | > 53 | 14 |
|  | LVSVi | 31-64 | 27-30 | < 27 | 18 | 37-75 | | 34-36 | < 34 | 14 |
|  | LVEF | 56-81 | 49-55 | < 49 | 18 | 48-79 | | 39-47 | < 39 | 14 |
|  | LVCOi | 2.1-4.4 | 1.9-2 | < 1.9 | 18 | 2.2-5.1 | | 2-2.1 | < 2 | 14 |
|  | LVMi | 40-81 | 82-89 | > 89 | 18 | 44-90 | | 91-97 | > 97 | 14 |
|  | LVM/LVEDV | 0.7-1.2 | 1.3-1.4 | > 1.4 | 18 | 0.7-1.2 | | 1.3-1.3 | > 1.3 | 14 |
|  | RVEDVi | 50-106 | 107-114 | > 114 | 18 | 66-125 | | 126-141 | > 141 | 14 |
|  | RVESVi | 13-46 | 47-49 | > 49 | 18 | 22-63 | | 64-74 | > 74 | 14 |
|  | RVEF | 46-71 | 39-45 | < 39 | 18 | 45-70 | | 41-44 | < 41 | 14 |
|  | RVCOi | 2.1-4.4 | 1.8-2 | < 1.8 | 18 | 2.4-4.7 | | 2.2-2.3 | < 2.2 | 14 |
|  | RVSVi | 31-66 | 27-30 | < 27 | 18 | 41-72 | | 38-40 | < 38 | 14 |
|  | LAVi max | 21-63 | 64-71 | > 71 | 15 |  | |  |  |  |
|  | LAEF | 44-74 | 35-43 | < 35 | 16 |  | |  |  |  |
|  | RAVi max | 23-65 | 66-72 | > 72 | 17 | 22-84 | | 85-95 | > 95 | 12 |
|  | RAEF | 33-68 | 28-32 | < 28 | 17 | 29-66 | | 25-28 | < 25 | 12 |

Footnote: All measurements were derived using cvi42 software with anatomical endocardial segmentation; papillary muscles and trabeculations were included in left ventricular mass measurements. Volumetric and mass measurements were indexed to height. Reference ranges are defined as the interval between the 2.5th and 97.5th centiles derived from age-specific generalized additive models for location, scale and shape (GAMLSS) using the Box–Cox Cole–Green distribution. Centile-based categories beyond the reference range are derived from LMS-based thresholds (P0.5, P2.5, P97.5, and P99.5) and describe the position of a measurement within the reference distribution rather than disease severity or diagnostic abnormality. Reference ranges are based on a Black adult population.

Abbreviations: LV = left ventricle; RV = right ventricle; LA = left atrium; RA = right atrium; EDV = end-diastolic volume; ESV = end-systolic volume; SV = stroke volume; EF = ejection fraction; CO = cardiac output.

### 8. Table: Reference ranges and centile-based categories for cardiac chamber measurements derived using cvi42 software (anatomical segmentation), indexed to height, in an Asian adult population

|  |  | **Female** | | | | **Male** | | | |
| --- | --- | --- | --- | --- | --- | --- | --- | --- | --- |
| **Age group** | **Metrics** | **Reference range** | **Mild** | **At least moderate** | **n** | **Reference range** | **Mild** | **At least moderate** | **n** |
| 40-49 | LVEDVi | 47-81 | 82-87 | > 87 | 10 | 56-101 | 102-111 | > 111 | 20 |
|  | LVESVi | 12-35 | 36-42 | > 42 | 10 | 15-39 | 40-44 | > 44 | 20 |
|  | LVSVi | 33-58 | 30-32 | < 30 | 10 | 39-62 | 36-38 | < 36 | 20 |
|  | LVEF | 56-78 | 51-55 | < 51 | 10 | 55-77 | 51-54 | < 51 | 20 |
|  | LVCOi | 2.1-4 | 1.9-2 | < 1.9 | 10 | 2.2-4.5 | 2-2.1 | < 2 | 20 |
|  | LVMi | 35-62 | 63-68 | > 68 | 10 | 45-81 | 82-88 | > 88 | 20 |
|  | LVM/LVEDV | 0.6-0.9 | 01-1 | > 1 | 10 | 0.7-1 | 1.1-1.1 | > 1.1 | 20 |
|  | RVEDVi | 50-93 | 94-102 | > 102 | 10 | 64-123 | 124-137 | > 137 | 20 |
|  | RVESVi | 16-41 | 42-46 | > 46 | 10 | 24-55 | 56-62 | > 62 | 20 |
|  | RVEF | 50-72 | 46-49 | < 46 | 10 | 48-67 | 45-47 | < 45 | 20 |
|  | RVCOi | 2-3.9 | 1.8-1.9 | < 1.8 | 10 | 2.2-4.3 | 2-2.1 | < 2 | 20 |
|  | RVSVi | 31-59 | 28-30 | < 28 | 10 | 35-69 | 30-34 | < 30 | 20 |
|  | LAVi max | 23-51 | 52-56 | > 56 | 10 | 20-57 | 58-64 | > 64 | 20 |
|  | LAEF | 57-81 | 54-56 | < 54 | 10 | 58-84 | 55-57 | < 55 | 20 |
|  | RAVi max | 22-47 | 48-51 | > 51 | 10 | 23-70 | 71-79 | > 79 | 20 |
|  | RAEF | 37-71 | 35-36 | < 35 | 10 | 32-63 | 27-31 | < 27 | 20 |
| 50-59 | LVEDVi | 46-81 | 82-87 | > 87 | 83 | 52-99 | 100-108 | > 108 | 133 |
|  | LVESVi | 12-33 | 34-40 | > 40 | 83 | 13-39 | 40-44 | > 44 | 133 |
|  | LVSVi | 32-58 | 29-31 | < 29 | 83 | 33-67 | 29-32 | < 29 | 133 |
|  | LVEF | 57-79 | 52-56 | < 52 | 83 | 55-78 | 50-54 | < 50 | 132 |
|  | LVCOi | 2-4.1 | 1.8-1.9 | < 1.8 | 83 | 2.1-4.4 | 1.9-2 | < 1.9 | 133 |
|  | LVMi | 35-64 | 65-71 | > 71 | 83 | 43-83 | 84-91 | > 91 | 133 |
|  | LVM/LVEDV | 0.6-1 | 1.1-1.1 | > 1.1 | 83 | 0.7-1.1 |  | > 1.2 | 133 |
|  | RVEDVi | 50-95 | 96-105 | > 105 | 83 | 60-118 | 119-130 | > 130 | 133 |
|  | RVESVi | 16-40 | 41-45 | > 45 | 83 | 22-54 | 55-60 | > 60 | 133 |
|  | RVEF | 51-72 | 48-50 | < 48 | 83 | 47-67 | 44-46 | < 44 | 132 |
|  | RVCOi | 1.9-4 | 1.7-1.8 | < 1.7 | 83 | 2-4.3 | 1.8-1.9 | < 1.8 | 133 |
|  | RVSVi | 31-60 | 28-30 | < 28 | 83 | 32-68 | 27-31 | < 27 | 133 |
|  | LAVi max | 22-52 | 53-58 | > 58 | 77 | 21-58 | 59-65 | > 65 | 121 |
|  | LAEF | 54-80 | 51-53 | < 51 | 75 | 55-82 | 52-54 | < 52 | 120 |
|  | RAVi max | 22-53 | 54-58 | > 58 | 76 | 23-69 | 70-79 | > 79 | 128 |
|  | RAEF | 37-69 | 34-36 | < 34 | 76 | 34-65 | 29-33 | < 29 | 127 |
| 60-69 | LVEDVi | 45-82 | 83-90 | > 90 | 63 | 46-95 | 96-104 | > 104 | 118 |
|  | LVESVi | 11-30 | 31-36 | > 36 | 63 | 12-38 | 39-45 | > 45 | 119 |
|  | LVSVi | 31-59 | 28-30 | < 28 | 63 | 32-67 | 28-31 | < 28 | 118 |
|  | LVEF | 58-81 | 53-57 | < 53 | 63 | 54-79 | 49-53 | < 49 | 119 |
|  | LVCOi | 1.9-4.2 | 1.7-1.8 | < 1.7 | 63 | 1.9-4.3 | 1.6-1.8 | < 1.6 | 119 |
|  | LVMi | 35-67 | 68-77 | > 77 | 63 | 39-85 | 86-95 | > 95 | 119 |
|  | LVM/LVEDV | 0.6-1 | 1.1-1.1 | > 1.1 | 63 | 0.7-1.2 | 1.3-1.4 | > 1.4 | 118 |
|  | RVEDVi | 50-98 | 99-111 | > 111 | 63 | 54-111 | 112-121 | > 121 | 119 |
|  | RVESVi | 16-39 | 40-44 | > 44 | 63 | 20-52 | 53-58 | > 58 | 119 |
|  | RVEF | 53-72 | 50-52 | < 50 | 63 | 46-68 | 42-45 | < 42 | 118 |
|  | RVCOi | 1.9-4.3 | 1.7-1.8 | < 1.7 | 63 | 1.8-4.3 | 1.4-1.7 | < 1.4 | 119 |
|  | RVSVi | 30-62 | 27-29 | < 27 | 63 | 29-65 | 24-28 | < 24 | 119 |
|  | LAVi max | 22-54 | 55-60 | > 60 | 59 | 21-59 | 60-66 | > 66 | 110 |
|  | LAEF | 50-78 | 46-49 | < 46 | 58 | 51-80 | 48-50 | < 48 | 107 |
|  | RAVi max | 21-61 | 62-68 | > 68 | 61 | 23-68 | 69-77 | > 77 | 113 |
|  | RAEF | 36-68 | 31-35 | < 31 | 59 | 35-70 | 31-34 | < 31 | 114 |
| 70+ | LVEDVi | 43-84 | 85-96 | > 96 | 28 | 40-91 | 92-99 | > 99 | 41 |
|  | LVESVi | 10-28 | 29-33 | > 33 | 28 | 10-38 | 39-47 | > 47 | 41 |
|  | LVSVi | 30-61 | 28-29 | < 28 | 28 | 33-62 | 30-32 | < 30 | 40 |
|  | LVEF | 59-83 | 54-58 | < 54 | 26 | 54-81 | 47-53 | < 47 | 41 |
|  | LVCOi | 1.8-4.4 | 1.6-1.7 | < 1.6 | 28 | 1.6-4.2 | 1.1-1.5 | < 1.1 | 41 |
|  | LVMi | 35-72 | 73-86 | > 86 | 28 | 36-89 | 90-101 | > 101 | 41 |
|  | LVM/LVEDV | 0.7-1 | 1.1-1.1 | > 1.1 | 28 | 0.7-1.3 | 1.4-1.7 | > 1.7 | 41 |
|  | RVEDVi | 48-98 | 99-114 | > 114 | 28 | 46-105 | 106-113 | > 113 | 41 |
|  | RVESVi | 16-37 | 38-43 | > 43 | 28 | 18-49 | 50-56 | > 56 | 41 |
|  | RVEF | 54-73 | 52-53 | < 52 | 28 | 45-70 | 40-44 | < 40 | 40 |
|  | RVCOi | 1.8-4.7 | 1.6-1.7 | < 1.6 | 28 | 1.4-4.2 | 0.9-1.3 | < 0.9 | 41 |
|  | RVSVi | 29-63 | 26-28 | < 26 | 28 | 29-63 | 24-28 | < 24 | 40 |
|  | LAVi max | 22-55 | 56-63 | > 63 | 28 | 21-60 | 61-67 | > 67 | 34 |
|  | LAEF | 45-76 | 40-44 | < 40 | 27 | 48-79 | 45-47 | < 45 | 34 |
|  | RAVi max | 17-65 | 66-76 | > 76 | 27 | 22-67 | 68-76 | > 76 | 38 |
|  | RAEF | 35-70 | 25-34 | < 25 | 27 | 36-77 | 32-35 | < 32 | 39 |

Footnote: All measurements were derived using cvi42 software with anatomical endocardial segmentation; papillary muscles and trabeculations were included in left ventricular mass measurements. Volumetric and mass measurements were indexed to height. Reference ranges are defined as the interval between the 2.5th and 97.5th centiles derived from age-specific generalized additive models for location, scale and shape (GAMLSS) using the Box–Cox Cole–Green distribution. Centile-based categories beyond the reference range are derived from LMS-based thresholds (P0.5, P2.5, P97.5, and P99.5) and describe the position of a measurement within the reference distribution rather than disease severity or diagnostic abnormality. Reference ranges are based on an Asian adult population.

Abbreviations: LV = left ventricle; RV = right ventricle; LA = left atrium; RA = right atrium; EDV = end-diastolic volume; ESV = end-systolic volume; SV = stroke volume; EF = ejection fraction; CO = cardiac output.

### 9. Table: Reference ranges and centile-based categories for cardiac chamber measurements derived using cvi42 software (anatomical segmentation), indexed to height, in a Chinese adult population

|  |  | **Female** | | | | **Male** | | | |
| --- | --- | --- | --- | --- | --- | --- | --- | --- | --- |
| **Age group** | **Metrics** | **Reference range** | **Mild** | **At least moderate** | **n** | **Reference range** | **Mild** | **At least moderate** | **n** |
| 18-29 | LVEDVi | 52-93 | 94-102 | > 102 | 16 | 62-102 | 103-106 | > 106 | 19 |
|  | LVESVi | 14-35 | 36-40 | > 40 | 16 | 19-40 | 41-43 | > 43 | 19 |
|  | LVSVi | 33-69 | 30-32 | < 30 | 16 | 39-70 | 35-38 | < 35 | 19 |
|  | LVEF | 56-77 | 53-55 | < 53 | 16 | 56-72 | 53-55 | < 53 | 19 |
|  | LVCOi | 2.1-4.7 | 1.9-2 | < 1.9 | 16 | 2.6-4.9 | 2.4-2.5 | < 2.4 | 19 |
|  | LVMi | 30-54 | 55-58 | > 58 | 16 | 41-79 | 80-84 | > 84 | 19 |
|  | LVM/LVEDV | 0.5-0.7 |  | > 0.7 | 16 | 0.6-0.9 | 01-1 | > 1 | 19 |
|  | RVEDVi | 46-91 | 92-99 | > 99 | 16 | 53-104 | 105-109 | > 109 | 19 |
|  | RVESVi | 14-39 | 40-44 | > 44 | 16 | 16-46 | 47-49 | > 49 | 19 |
|  | RVEF | 50-77 | 46-49 | < 46 | 16 | 48-69 | 45-47 | < 45 | 18 |
|  | RVCOi | 1.7-4.2 | 1.5-1.6 | < 1.5 | 16 | 2.1-4.8 | 1.7-2 | < 1.7 | 19 |
|  | RVSVi | 28-59 | 25-27 | < 25 | 16 | 33-71 | 28-32 | < 28 | 19 |
|  | LAVi max | 22-52 | 53-58 | > 58 | 13 | 27-52 | 53-56 | > 56 | 16 |
|  | LAEF | 54-77 | 50-53 | < 50 | 13 | 52-70 | 49-51 | < 49 | 16 |
|  | RAVi max | 19-48 | 49-57 | > 57 | 15 | 20-62 | 63-69 | > 69 | 18 |
|  | RAEF | 41-73 | 35-40 | < 35 | 15 | 38-68 | 31-37 | < 31 | 18 |
| 30-39 | LVEDVi | 51-90 | 91-99 | > 99 | 18 | 60-99 | 100-104 | > 104 | 17 |
|  | LVESVi | 13-34 | 35-39 | > 39 | 18 | 18-39 | 40-41 | > 41 | 17 |
|  | LVSVi | 34-64 | 31-33 | < 31 | 18 | 38-68 | 34-37 | < 34 | 17 |
|  | LVEF | 57-78 | 53-56 | < 53 | 18 | 56-73 | 53-55 | < 53 | 17 |
|  | LVCOi | 2-4.5 | 1.9-1.9 | < 1.9 | 18 | 2.5-4.8 | 2.3-2.4 | < 2.3 | 17 |
|  | LVMi | 32-56 | 57-62 | > 62 | 18 | 42-79 | 80-85 | > 85 | 17 |
|  | LVM/LVEDV | 0.5-0.7 | 0.8-0.8 | > 0.8 | 18 | 0.6-1 | 1.1-1.1 | > 1.1 | 17 |
|  | RVEDVi | 47-92 | 93-101 | > 101 | 18 | 53-105 | 106-109 | > 109 | 17 |
|  | RVESVi | 14-40 | 41-45 | > 45 | 18 | 15-47 | 48-50 | > 50 | 17 |
|  | RVEF | 50-76 | 46-49 | < 46 | 18 | 48-69 | 45-47 | < 45 | 17 |
|  | RVCOi | 1.7-4.1 | 1.5-1.6 | < 1.5 | 18 | 2-4.7 | 1.7-1.9 | < 1.7 | 17 |
|  | RVSVi | 29-59 | 25-28 | < 25 | 18 | 32-69 | 27-31 | < 27 | 17 |
|  | LAVi max | 22-51 | 52-57 | > 57 | 18 | 26-52 | 53-56 | > 56 | 14 |
|  | LAEF | 53-76 | 50-52 | < 50 | 18 | 51-72 | 48-50 | < 48 | 14 |
|  | RAVi max | 20-51 | 52-61 | > 61 | 18 | 20-63 | 64-71 | > 71 | 16 |
|  | RAEF | 41-72 | 35-40 | < 35 | 18 | 38-68 | 31-37 | < 31 | 16 |
| 40-49 | LVEDVi | 50-87 | 88-96 | > 96 | 24 | 58-97 | 98-103 | > 103 | 20 |
|  | LVESVi | 12-32 | 33-37 | > 37 | 24 | 16-37 | 38-40 | > 40 | 20 |
|  | LVSVi | 35-61 | 32-34 | < 32 | 24 | 37-66 | 33-36 | < 33 | 20 |
|  | LVEF | 58-79 | 54-57 | < 54 | 24 | 56-74 | 53-55 | < 53 | 19 |
|  | LVCOi | 2-4.3 | 1.8-1.9 | < 1.8 | 24 | 2.4-4.7 | 2.2-2.3 | < 2.2 | 20 |
|  | LVMi | 33-60 | 61-67 | > 67 | 24 | 43-80 | 81-87 | > 87 | 20 |
|  | LVM/LVEDV | 0.5-0.8 | 0.9-0.9 | > 0.9 | 24 | 0.6-1 | 1.1-1.1 | > 1.1 | 20 |
|  | RVEDVi | 48-93 | 94-104 | > 104 | 24 | 57-105 | 106-112 | > 112 | 20 |
|  | RVESVi | 14-40 | 41-46 | > 46 | 24 | 15-47 | 48-51 | > 51 | 20 |
|  | RVEF | 50-75 | 46-49 | < 46 | 24 | 48-70 | 45-47 | < 45 | 19 |
|  | RVCOi | 1.8-3.9 | 1.6-1.7 | < 1.6 | 24 | 2-4.6 | 1.7-1.9 | < 1.7 | 20 |
|  | RVSVi | 29-58 | 25-28 | < 25 | 24 | 31-68 | 26-30 | < 26 | 20 |
|  | LAVi max | 22-51 | 52-56 | > 56 | 22 | 24-52 | 53-56 | > 56 | 18 |
|  | LAEF | 53-75 | 49-52 | < 49 | 22 | 51-74 | 47-50 | < 47 | 18 |
|  | RAVi max | 22-54 | 55-65 | > 65 | 23 | 21-65 | 66-72 | > 72 | 19 |
|  | RAEF | 41-70 | 36-40 | < 36 | 23 | 37-67 | 30-36 | < 30 | 19 |
| 50-59 | LVEDVi | 48-85 | 86-93 | > 93 | 66 | 55-95 | 96-102 | > 102 | 49 |
|  | LVESVi | 11-30 | 31-35 | > 35 | 66 | 14-35 | 36-38 | > 38 | 49 |
|  | LVSVi | 34-58 | 32-33 | < 32 | 66 | 35-64 | 32-34 | < 32 | 49 |
|  | LVEF | 59-80 | 55-58 | < 55 | 65 | 56-76 | 53-55 | < 53 | 48 |
|  | LVCOi | 2-4.1 | 1.8-1.9 | < 1.8 | 66 | 2.3-4.6 | 2.1-2.2 | < 2.1 | 49 |
|  | LVMi | 35-64 | 65-74 | > 74 | 66 | 44-81 | 82-89 | > 89 | 49 |
|  | LVM/LVEDV | 0.6-0.9 | 01-1 | > 1 | 66 | 0.6-1.1 |  | > 1.2 | 49 |
|  | RVEDVi | 48-93 | 94-104 | > 104 | 66 | 59-107 | 108-116 | > 116 | 49 |
|  | RVESVi | 14-40 | 41-46 | > 46 | 66 | 15-48 | 49-52 | > 52 | 49 |
|  | RVEF | 50-74 | 46-49 | < 46 | 66 | 47-70 | 45-46 | < 45 | 49 |
|  | RVCOi | 1.8-3.8 | 1.6-1.7 | < 1.6 | 66 | 1.9-4.5 | 1.6-1.8 | < 1.6 | 49 |
|  | RVSVi | 29-57 | 25-28 | < 25 | 66 | 29-66 | 24-28 | < 24 | 49 |
|  | LAVi max | 21-50 | 51-56 | > 56 | 60 | 23-51 | 52-55 | > 55 | 40 |
|  | LAEF | 51-76 | 48-50 | < 48 | 58 | 50-77 | 46-49 | < 46 | 40 |
|  | RAVi max | 23-57 | 58-69 | > 69 | 63 | 21-66 | 67-74 | > 74 | 46 |
|  | RAEF | 40-68 | 36-39 | < 36 | 63 | 37-66 | 30-36 | < 30 | 46 |
| 60-69 | LVEDVi | 46-82 | 83-91 | > 91 | 52 | 53-94 | 95-103 | > 103 | 37 |
|  | LVESVi | 10-28 | 29-33 | > 33 | 52 | 12-34 | 35-37 | > 37 | 36 |
|  | LVSVi | 33-58 | 29-32 | < 29 | 52 | 34-63 | 30-33 | < 30 | 37 |
|  | LVEF | 60-81 | 55-59 | < 55 | 52 | 56-78 | 53-55 | < 53 | 36 |
|  | LVCOi | 2-3.9 | 1.8-1.9 | < 1.8 | 52 | 2.2-4.5 | 2-2.1 | < 2 | 37 |
|  | LVMi | 36-69 | 70-87 | > 87 | 52 | 45-82 | 83-93 | > 93 | 37 |
|  | LVM/LVEDV | 0.6-1.1 | 1.2-1.4 | > 1.4 | 52 | 0.6-1.1 |  | > 1.2 | 37 |
|  | RVEDVi | 47-90 | 91-99 | > 99 | 52 | 60-111 | 112-125 | > 125 | 36 |
|  | RVESVi | 14-39 | 40-44 | > 44 | 52 | 17-49 | 50-53 | > 53 | 37 |
|  | RVEF | 51-74 | 46-50 | < 46 | 52 | 47-71 | 45-46 | < 45 | 36 |
|  | RVCOi | 1.8-3.7 | 1.6-1.7 | < 1.6 | 52 | 1.8-4.4 | 1.6-1.7 | < 1.6 | 37 |
|  | RVSVi | 29-57 | 26-28 | < 26 | 52 | 28-64 | 22-27 | < 22 | 37 |
|  | LAVi max | 21-51 | 52-56 | > 56 | 47 | 20-49 | 50-53 | > 53 | 30 |
|  | LAEF | 49-76 | 45-48 | < 45 | 46 | 48-80 | 44-47 | < 44 | 30 |
|  | RAVi max | 24-61 | 62-73 | > 73 | 46 | 22-68 | 69-76 | > 76 | 35 |
|  | RAEF | 40-66 | 36-39 | < 36 | 46 | 38-68 | 31-37 | < 31 | 35 |
| 70+ | LVEDVi | 45-80 | 81-89 | > 89 | 12 |  |  |  |  |
|  | LVESVi | 9-27 | 28-31 | > 31 | 12 |  |  |  |  |
|  | LVSVi | 29-60 | 25-28 | < 25 | 12 |  |  |  |  |
|  | LVEF | 61-82 | 55-60 | < 55 | 12 |  |  |  |  |
|  | LVCOi | 2-3.7 | 1.8-1.9 | < 1.8 | 12 |  |  |  |  |
|  | LVMi | 37-75 | 76-106 | > 106 | 12 |  |  |  |  |
|  | LVM/LVEDV | 0.7-1.3 | 1.4-1.8 | > 1.8 | 12 |  |  |  |  |
|  | RVEDVi | 44-87 | 88-93 | > 93 | 12 |  |  |  |  |
|  | RVESVi | 14-37 | 38-42 | > 42 | 12 |  |  |  |  |
|  | RVEF | 51-73 | 47-50 | < 47 | 12 |  |  |  |  |
|  | RVCOi | 1.8-3.6 | 1.6-1.7 | < 1.6 | 12 |  |  |  |  |
|  | RVSVi | 29-56 | 26-28 | < 26 | 12 |  |  |  |  |
|  | LAVi max | 20-51 | 52-57 | > 57 | 11 |  |  |  |  |
|  | LAEF | 47-78 | 42-46 | < 42 | 12 |  |  |  |  |
|  | RAVi max | 25-64 | 65-76 | > 76 | 11 |  |  |  |  |
|  | RAEF | 39-65 | 36-38 | < 36 | 11 |  |  |  |  |

Footnote: All measurements were derived using cvi42 software with anatomical endocardial segmentation; papillary muscles and trabeculations were included in left ventricular mass measurements. Volumetric and mass measurements were indexed to height. Reference ranges are defined as the interval between the 2.5th and 97.5th centiles derived from age-specific generalized additive models for location, scale and shape (GAMLSS) using the Box–Cox Cole–Green distribution. Centile-based categories beyond the reference range are derived from LMS-based thresholds (P0.5, P2.5, P97.5, and P99.5) and describe the position of a measurement within the reference distribution rather than disease severity or diagnostic abnormality. Reference ranges are based on a Chinese adult population.

Abbreviations: LV = left ventricle; RV = right ventricle; LA = left atrium; RA = right atrium; EDV = end-diastolic volume; ESV = end-systolic volume; SV = stroke volume; EF = ejection fraction; CO = cardiac output.

### 10. Table: Reference ranges and centile-based categories for cardiac chamber measurements derived using cvi42 software (anatomical segmentation), indexed to height, in a Mixed adult population

|  |  | **Female** | | | | **Male** | | | |
| --- | --- | --- | --- | --- | --- | --- | --- | --- | --- |
| **Age group** | **Metrics** | **Reference range** | **Mild** | **At least moderate** | **n** | **Reference range** | **Mild** | **At least moderate** | **n** |
| 50-59 | LVEDVi | 52-95 | 96-103 | > 103 | 134 | 61-116 | 117-127 | > 127 | 66 |
|  | LVESVi | 13-34 | 35-38 | > 38 | 134 | 16-42 | 43-47 | > 47 | 66 |
|  | LVSVi | 35-69 | 32-34 | < 32 | 133 | 37-74 | 28-36 | < 28 | 65 |
|  | LVEF | 58-80 | 54-57 | < 54 | 134 | 56-79 | 52-55 | < 52 | 66 |
|  | LVCOi | 2.1-5 | 1.9-2 | < 1.9 | 133 | 2.2-5.2 | 1.9-2.1 | < 1.9 | 66 |
|  | LVMi | 36-75 | 76-84 | > 84 | 134 | 47-93 | 94-100 | > 100 | 64 |
|  | LVM/LVEDV | 0.6-0.9 | 01-1 | > 1 | 134 | 0.7-1.1 | 1.2-1.4 | > 1.4 | 64 |
|  | RVEDVi | 55-112 | 113-123 | > 123 | 134 | 66-132 | 133-142 | > 142 | 65 |
|  | RVESVi | 18-45 | 46-50 | > 50 | 134 | 23-57 | 58-62 | > 62 | 65 |
|  | RVEF | 52-70 | 48-51 | < 48 | 134 | 50-69 | 47-49 | < 47 | 66 |
|  | RVCOi | 2-4.8 | 1.7-1.9 | < 1.7 | 133 | 2.3-5.3 | 2-2.2 | < 2 | 66 |
|  | RVSVi | 32-71 | 28-31 | < 28 | 134 | 38-79 | 30-37 | < 30 | 66 |
|  | LAVi max | 23-55 | 56-60 | > 60 | 121 | 25-70 | 71-79 | > 79 | 53 |
|  | LAEF | 55-78 | 52-54 | < 52 | 120 | 54-78 | 51-53 | < 51 | 53 |
|  | RAVi max | 24-66 | 67-75 | > 75 | 128 | 26-79 | 80-88 | > 88 | 64 |
|  | RAEF | 37-64 | 31-36 | < 31 | 124 | 35-63 | 31-34 | < 31 | 64 |
| 60-69 | LVEDVi | 47-92 | 93-102 | > 102 | 110 | 54-106 | 107-115 | > 115 | 59 |
|  | LVESVi | 11-33 | 34-38 | > 38 | 110 | 13-42 | 43-48 | > 48 | 59 |
|  | LVSVi | 33-64 | 30-32 | < 30 | 110 | 36-69 | 30-35 | < 30 | 59 |
|  | LVEF | 58-81 | 54-57 | < 54 | 110 | 55-77 | 51-54 | < 51 | 58 |
|  | LVCOi | 2-4.4 | 1.8-1.9 | < 1.8 | 110 | 2.2-4.8 | 2-2.1 | < 2 | 59 |
|  | LVMi | 36-70 | 71-77 | > 77 | 110 | 47-93 | 94-102 | > 102 | 59 |
|  | LVM/LVEDV | 0.6-1 | 1.1-1.1 | > 1.1 | 110 | 0.7-1.2 | 1.3-1.5 | > 1.5 | 59 |
|  | RVEDVi | 52-106 | 107-117 | > 117 | 110 | 65-128 | 129-141 | > 141 | 58 |
|  | RVESVi | 18-44 | 45-50 | > 50 | 110 | 24-58 | 59-65 | > 65 | 58 |
|  | RVEF | 51-71 | 47-50 | < 47 | 110 | 48-67 | 46-47 | < 46 | 59 |
|  | RVCOi | 1.9-4.3 | 1.7-1.8 | < 1.7 | 110 | 2.2-5 | 2-2.1 | < 2 | 59 |
|  | RVSVi | 32-66 | 28-31 | < 28 | 110 | 36-75 | 31-35 | < 31 | 59 |
|  | LAVi max | 23-58 | 59-65 | > 65 | 100 | 24-65 | 66-74 | > 74 | 52 |
|  | LAEF | 52-79 | 49-51 | < 49 | 98 | 50-79 | 47-49 | < 47 | 52 |
|  | RAVi max | 23-68 | 69-77 | > 77 | 103 | 20-78 | 79-85 | > 85 | 57 |
|  | RAEF | 35-68 | 29-34 | < 29 | 103 | 36-68 | 33-35 | < 33 | 57 |
| 70+ | LVEDVi | 47-84 | 85-92 | > 92 | 50 | 49-98 | 99-106 | > 106 | 33 |
|  | LVESVi | 10-33 | 34-38 | > 38 | 51 | 11-42 | 43-48 | > 48 | 34 |
|  | LVSVi | 31-58 | 27-30 | < 27 | 50 | 34-66 | 30-33 | < 30 | 34 |
|  | LVEF | 59-81 | 54-58 | < 54 | 51 | 54-76 | 49-53 | < 49 | 35 |
|  | LVCOi | 02-4 | 1.8-1.9 | < 1.8 | 51 | 2.1-4.5 | 1.9-2 | < 1.9 | 35 |
|  | LVMi | 37-67 | 68-73 | > 73 | 51 | 47-93 | 94-105 | > 105 | 34 |
|  | LVM/LVEDV | 0.6-1 | 1.1-1.1 | > 1.1 | 50 | 0.7-1.3 | 1.4-1.5 | > 1.5 | 33 |
|  | RVEDVi | 55-93 | 94-100 | > 100 | 51 | 63-127 | 128-145 | > 145 | 35 |
|  | RVESVi | 18-44 | 45-51 | > 51 | 51 | 25-60 | 61-69 | > 69 | 35 |
|  | RVEF | 50-71 | 45-49 | < 45 | 51 | 47-66 | 44-46 | < 44 | 35 |
|  | RVCOi | 1.9-4 | 1.7-1.8 | < 1.7 | 51 | 2.1-4.6 | 02-2 | < 2 | 35 |
|  | RVSVi | 32-59 | 29-31 | < 29 | 50 | 34-71 | 31-33 | < 31 | 35 |
|  | LAVi max | 24-62 | 63-72 | > 72 | 45 | 22-61 | 62-70 | > 70 | 32 |
|  | LAEF | 50-80 | 47-49 | < 47 | 44 | 46-81 | 42-45 | < 42 | 30 |
|  | RAVi max | 21-68 | 69-77 | > 77 | 47 | 18-76 | 77-83 | > 83 | 31 |
|  | RAEF | 33-72 | 28-32 | < 28 | 46 | 36-76 | 34-35 | < 34 | 31 |

Footnote: All measurements were derived using cvi42 software with anatomical endocardial segmentation; papillary muscles and trabeculations were included in left ventricular mass measurements. Volumetric and mass measurements were indexed to height. Reference ranges are defined as the interval between the 2.5th and 97.5th centiles derived from age-specific generalized additive models for location, scale and shape (GAMLSS) using the Box–Cox Cole–Green distribution. Centile-based categories beyond the reference range are derived from LMS-based thresholds (P0.5, P2.5, P97.5, and P99.5) and describe the position of a measurement within the reference distribution rather than disease severity or diagnostic abnormality. Reference ranges are based on a Mixed adult population.

Abbreviations: LV = left ventricle; RV = right ventricle; LA = left atrium; RA = right atrium; EDV = end-diastolic volume; ESV = end-systolic volume; SV = stroke volume; EF = ejection fraction; CO = cardiac output.

### 11. Table: Reference ranges and centile-based categories for cardiac chamber measurements derived using cvi42 software (papillary segmentation), indexed to height, in a White adult population

|  |  | **Female** | | | | | **Male** | | | |
| --- | --- | --- | --- | --- | --- | --- | --- | --- | --- | --- |
| **Age group** | **Metrics** | **Reference range** | **Mild** | **At least moderate** | **n** | **Reference range** | | **Mild** | **At least moderate** | **n** |
| 18-29 | LVEDVi | 58-104 | 105-111 | > 111 | 102 | 73-126 | | 127-133 | > 133 | 100 |
|  | LVESVi | 17-46 | 47-52 | > 52 | 102 | 23-57 | | 58-63 | > 63 | 100 |
|  | LVSVi | 35-66 | 30-34 | < 30 | 102 | 40-78 | | 33-39 | < 33 | 99 |
|  | LVEF | 53-72 | 51-52 | < 51 | 100 | 49-69 | | 45-48 | < 45 | 102 |
|  | LVCOi | 2.3-4.9 | 2-2.2 | < 2 | 101 | 2.4-5.4 | | 1.7-2.3 | < 1.7 | 97 |
|  | LVMi | 36-60 | 61-66 | > 66 | 102 | 47-84 | | 85-90 | > 90 | 105 |
|  | LVM/LVEDV | 0.5-0.7 | 0.8-0.8 | > 0.8 | 102 | 0.5-0.8 | | 0.9-0.9 | > 0.9 | 100 |
|  | RVEDVi | 55-108 | 109-116 | > 116 | 102 | 72-138 | | 139-146 | > 146 | 101 |
|  | RVESVi | 16-52 | 53-56 | > 56 | 100 | 26-69 | | 70-75 | > 75 | 98 |
|  | RVEF | 46-69 | 42-45 | < 42 | 97 | 44-67 | | 41-43 | < 41 | 98 |
|  | RVCOi | 2-5.1 | 1.6-1.9 | < 1.6 | 101 | 2.3-5.5 | | 1.8-2.2 | < 1.8 | 97 |
|  | RVSVi | 31-68 | 26-30 | < 26 | 102 | 39-81 | | 33-38 | < 33 | 98 |
|  | LAVi max | 25-51 | 52-55 | > 55 | 50 | 28-63 | | 64-70 | > 70 | 52 |
|  | LAEF | 56-75 | 54-55 | < 54 | 50 | 53-72 | | 50-52 | < 50 | 52 |
|  | RAVi max | 21-51 | 52-56 | > 56 | 80 | 28-68 | | 69-77 | > 77 | 82 |
|  | RAEF | 38-78 | 33-37 | < 33 | 78 | 35-74 | | 31-34 | < 31 | 82 |
| 30-39 | LVEDVi | 57-103 | 104-110 | > 110 | 137 | 67-120 | | 121-127 | > 127 | 133 |
|  | LVESVi | 17-46 | 47-51 | > 51 | 137 | 22-55 | | 56-61 | > 61 | 133 |
|  | LVSVi | 34-66 | 30-33 | < 30 | 134 | 38-75 | | 31-37 | < 31 | 132 |
|  | LVEF | 53-73 | 50-52 | < 50 | 132 | 50-71 | | 47-49 | < 47 | 134 |
|  | LVCOi | 2.3-5 | 2-2.2 | < 2 | 136 | 2.4-5.3 | | 1.8-2.3 | < 1.8 | 125 |
|  | LVMi | 35-61 | 62-66 | > 66 | 137 | 44-81 | | 82-86 | > 86 | 134 |
|  | LVM/LVEDV | 0.5-0.8 |  | > 0.8 | 137 | 0.5-0.9 | | 01-1 | > 1 | 134 |
|  | RVEDVi | 55-109 | 110-117 | > 117 | 137 | 71-137 | | 138-146 | > 146 | 131 |
|  | RVESVi | 16-50 | 51-54 | > 54 | 135 | 25-66 | | 67-72 | > 72 | 128 |
|  | RVEF | 47-71 | 44-46 | < 44 | 127 | 45-68 | | 41-44 | < 41 | 131 |
|  | RVCOi | 2-5.1 | 1.7-1.9 | < 1.7 | 133 | 2.3-5.5 | | 1.8-2.2 | < 1.8 | 124 |
|  | RVSVi | 32-69 | 27-31 | < 27 | 134 | 38-81 | | 32-37 | < 32 | 132 |
|  | LAVi max | 25-55 | 56-60 | > 60 | 84 | 27-64 | | 65-71 | > 71 | 66 |
|  | LAEF | 55-76 | 53-54 | < 53 | 83 | 53-73 | | 50-52 | < 50 | 66 |
|  | RAVi max | 21-52 | 53-57 | > 57 | 113 | 27-69 | | 70-77 | > 77 | 119 |
|  | RAEF | 38-75 | 33-37 | < 33 | 110 | 35-72 | | 31-34 | < 31 | 118 |
| 40-49 | LVEDVi | 58-105 | 106-114 | > 114 | 518 | 66-123 | | 124-131 | > 131 | 432 |
|  | LVESVi | 16-45 | 46-50 | > 50 | 518 | 20-53 | | 54-59 | > 59 | 427 |
|  | LVSVi | 35-68 | 30-34 | < 30 | 518 | 38-78 | | 32-37 | < 32 | 432 |
|  | LVEF | 53-73 | 50-52 | < 50 | 512 | 50-72 | | 47-49 | < 47 | 434 |
|  | LVCOi | 2.2-4.9 | 2-2.1 | < 2 | 514 | 2.3-5.2 | | 1.8-2.2 | < 1.8 | 425 |
|  | LVMi | 35-62 | 63-68 | > 68 | 518 | 44-83 | | 84-89 | > 89 | 441 |
|  | LVM/LVEDV | 0.5-0.8 |  | > 0.8 | 518 | 0.5-0.9 | | 01-1 | > 1 | 432 |
|  | RVEDVi | 56-111 | 112-120 | > 120 | 517 | 69-136 | | 137-145 | > 145 | 432 |
|  | RVESVi | 16-48 | 49-52 | > 52 | 517 | 24-63 | | 64-69 | > 69 | 425 |
|  | RVEF | 50-71 | 47-49 | < 47 | 510 | 46-68 | | 42-45 | < 42 | 429 |
|  | RVCOi | 2.1-5 | 1.7-2 | < 1.7 | 514 | 2.3-5.4 | | 1.8-2.2 | < 1.8 | 423 |
|  | RVSVi | 33-70 | 28-32 | < 28 | 516 | 38-81 | | 31-37 | < 31 | 429 |
|  | LAVi max | 25-59 | 60-65 | > 65 | 440 | 25-64 | | 65-72 | > 72 | 358 |
|  | LAEF | 55-78 | 52-54 | < 52 | 434 | 53-76 | | 50-52 | < 50 | 353 |
|  | RAVi max | 23-59 | 60-66 | > 66 | 500 | 28-77 | | 78-87 | > 87 | 408 |
|  | RAEF | 38-72 | 33-37 | < 33 | 497 | 35-69 | | 31-34 | < 31 | 410 |
| 50-59 | LVEDVi | 58-107 | 108-117 | > 117 | 911 | 63-122 | | 123-132 | > 132 | 812 |
|  | LVESVi | 16-43 | 44-48 | > 48 | 910 | 19-52 | | 53-57 | > 57 | 818 |
|  | LVSVi | 35-70 | 31-34 | < 31 | 910 | 37-77 | | 31-36 | < 31 | 818 |
|  | LVEF | 53-74 | 49-52 | < 49 | 906 | 50-73 | | 47-49 | < 47 | 831 |
|  | LVCOi | 2.1-4.7 | 1.9-2 | < 1.9 | 906 | 2.3-5 | | 1.9-2.2 | < 1.9 | 815 |
|  | LVMi | 35-64 | 65-70 | > 70 | 911 | 45-86 | | 87-93 | > 93 | 832 |
|  | LVM/LVEDV | 0.5-0.8 | 0.9-0.9 | > 0.9 | 911 | 0.6-0.9 | | 1-1.1 | > 1.1 | 809 |
|  | RVEDVi | 57-112 | 113-122 | > 122 | 909 | 67-133 | | 134-143 | > 143 | 818 |
|  | RVESVi | 16-46 | 47-50 | > 50 | 909 | 23-60 | | 61-66 | > 66 | 818 |
|  | RVEF | 52-72 | 49-51 | < 49 | 906 | 47-69 | | 43-46 | < 43 | 830 |
|  | RVCOi | 2-4.9 | 1.8-1.9 | < 1.8 | 906 | 2.2-5.2 | | 1.8-2.1 | < 1.8 | 815 |
|  | RVSVi | 34-72 | 29-33 | < 29 | 909 | 37-80 | | 30-36 | < 30 | 820 |
|  | LAVi max | 24-62 | 63-69 | > 69 | 797 | 24-65 | | 66-73 | > 73 | 684 |
|  | LAEF | 53-79 | 50-52 | < 50 | 791 | 53-79 | | 50-52 | < 50 | 681 |
|  | RAVi max | 24-66 | 67-74 | > 74 | 854 | 27-78 | | 79-87 | > 87 | 761 |
|  | RAEF | 38-70 | 33-37 | < 33 | 844 | 35-68 | | 31-34 | < 31 | 771 |
| 60-69 | LVEDVi | 54-103 | 104-113 | > 113 | 918 | 60-120 | | 121-130 | > 130 | 797 |
|  | LVESVi | 15-40 | 41-45 | > 45 | 919 | 17-50 | | 51-55 | > 55 | 791 |
|  | LVSVi | 33-67 | 29-32 | < 29 | 918 | 36-76 | | 29-35 | < 29 | 798 |
|  | LVEF | 54-75 | 50-53 | < 50 | 916 | 50-73 | | 47-49 | < 47 | 798 |
|  | LVCOi | 2.1-4.5 | 1.8-2 | < 1.8 | 917 | 2.2-4.8 | | 1.8-2.1 | < 1.8 | 792 |
|  | LVMi | 34-65 | 66-71 | > 71 | 919 | 44-85 | | 86-92 | > 92 | 801 |
|  | LVM/LVEDV | 0.5-0.8 | 0.9-0.9 | > 0.9 | 918 | 0.6-1 | | 1.1-1.1 | > 1.1 | 794 |
|  | RVEDVi | 54-107 | 108-118 | > 118 | 917 | 64-128 | | 129-139 | > 139 | 798 |
|  | RVESVi | 15-44 | 45-48 | > 48 | 920 | 21-57 | | 58-62 | > 62 | 801 |
|  | RVEF | 52-73 | 49-51 | < 49 | 914 | 48-70 | | 44-47 | < 44 | 803 |
|  | RVCOi | 2-4.7 | 1.7-1.9 | < 1.7 | 917 | 2.2-5 | | 1.8-2.1 | < 1.8 | 797 |
|  | RVSVi | 33-69 | 29-32 | < 29 | 917 | 35-78 | | 28-34 | < 28 | 792 |
|  | LAVi max | 22-63 | 64-71 | > 71 | 797 | 22-66 | | 67-74 | > 74 | 645 |
|  | LAEF | 51-79 | 47-50 | < 47 | 776 | 51-80 | | 47-50 | < 47 | 632 |
|  | RAVi max | 24-67 | 68-76 | > 76 | 855 | 25-79 | | 80-88 | > 88 | 742 |
|  | RAEF | 38-71 | 33-37 | < 33 | 841 | 35-70 | | 31-34 | < 31 | 748 |
| 70+ | LVEDVi | 52-100 | 101-112 | > 112 | 1101 | 56-116 | | 117-127 | > 127 | 1253 |
|  | LVESVi | 14-37 | 38-42 | > 42 | 1100 | 16-48 | | 49-53 | > 53 | 1247 |
|  | LVSVi | 32-65 | 28-31 | < 28 | 1102 | 33-73 | | 27-32 | < 27 | 1256 |
|  | LVEF | 54-76 | 51-53 | < 51 | 1096 | 51-74 | | 47-50 | < 47 | 1242 |
|  | LVCOi | 2-4.5 | 1.8-1.9 | < 1.8 | 1099 | 2.1-4.7 | | 1.8-2 | < 1.8 | 1254 |
|  | LVMi | 33-64 | 65-70 | > 70 | 1102 | 42-82 | | 83-89 | > 89 | 1257 |
|  | LVM/LVEDV | 0.5-0.9 | 01-1 | > 1 | 1101 | 0.6-1 | | 1.1-1.1 | > 1.1 | 1251 |
|  | RVEDVi | 52-104 | 105-115 | > 115 | 1100 | 60-124 | | 125-134 | > 134 | 1258 |
|  | RVESVi | 15-41 | 42-46 | > 46 | 1100 | 20-53 | | 54-58 | > 58 | 1259 |
|  | RVEF | 53-74 | 50-52 | < 50 | 1093 | 49-70 | | 45-48 | < 45 | 1246 |
|  | RVCOi | 2-4.6 | 1.7-1.9 | < 1.7 | 1099 | 2.1-4.8 | | 1.7-2 | < 1.7 | 1251 |
|  | RVSVi | 32-66 | 28-31 | < 28 | 1100 | 33-76 | | 27-32 | < 27 | 1256 |
|  | LAVi max | 21-63 | 64-73 | > 73 | 942 | 20-67 | | 68-75 | > 75 | 953 |
|  | LAEF | 48-79 | 44-47 | < 44 | 903 | 48-80 | | 44-47 | < 44 | 914 |
|  | RAVi max | 23-68 | 69-78 | > 78 | 1028 | 23-78 | | 79-87 | > 87 | 1119 |
|  | RAEF | 38-72 | 33-37 | < 33 | 1001 | 35-73 | | 30-34 | < 30 | 1115 |

Footnote: All measurements were derived using cvi42 software with papillary endocardial segmentation; papillary muscles were included in left ventricular mass measurements. Volumetric and mass measurements were indexed to height. Reference ranges are defined as the interval between the 2.5th and 97.5th centiles derived from age-specific generalized additive models for location, scale and shape (GAMLSS) using the Box–Cox Cole–Green distribution. Centile-based categories beyond the reference range are derived from LMS-based thresholds (P0.5, P2.5, P97.5, and P99.5) and describe the position of a measurement within the reference distribution rather than disease severity or diagnostic abnormality. Reference ranges are based on a White adult population.

Abbreviations: LV = left ventricle; RV = right ventricle; LA = left atrium; RA = right atrium; EDV = end-diastolic volume; ESV = end-systolic volume; SV = stroke volume; EF = ejection fraction; CO = cardiac output.

### 12. Table: Reference ranges and centile-based categories for cardiac chamber measurements derived using cvi42 software (papillary segmentation), indexed to height, in a Black adult population

|  |  | **Female** | | | | | **Male** | | | |
| --- | --- | --- | --- | --- | --- | --- | --- | --- | --- | --- |
| **Age group** | **Metrics** | **Reference range** | **Mild** | **At least moderate** | **n** | **Reference range** | | **Mild** | **At least moderate** | **n** |
| 40-49 | LVEDVi | 65-115 | 116-130 | > 130 | 10 |  | |  |  |  |
|  | LVESVi | 20-46 | 47-51 | > 51 | 10 |  | |  |  |  |
|  | LVSVi | 36-72 | 33-35 | < 33 | 10 |  | |  |  |  |
|  | LVEF | 52-75 | 50-51 | < 50 | 10 |  | |  |  |  |
|  | LVCOi | 2-5.2 | 1.8-1.9 | < 1.8 | 10 |  | |  |  |  |
|  | LVMi | 35-71 | 72-77 | > 77 | 10 |  | |  |  |  |
|  | LVM/LVEDV | 0.5-0.8 | 0.9-1 | > 1 | 10 |  | |  |  |  |
|  | RVEDVi | 61-121 | 122-133 | > 133 | 10 |  | |  |  |  |
|  | RVESVi | 21-56 | 57-66 | > 66 | 10 |  | |  |  |  |
|  | RVEF | 52-73 | 49-51 | < 49 | 10 |  | |  |  |  |
|  | RVCOi | 2.1-5.5 | 1.8-2 | < 1.8 | 10 |  | |  |  |  |
|  | RVSVi | 35-72 | 30-34 | < 30 | 10 |  | |  |  |  |
|  | LAVi max |  |  |  |  |  | |  |  |  |
|  | LAEF |  |  |  |  |  | |  |  |  |
|  | RAVi max | 28-76 | 77-91 | > 91 | 10 |  | |  |  |  |
|  | RAEF | 30-65 | 25-29 | < 25 | 10 |  | |  |  |  |
| 50-59 | LVEDVi | 61-109 | 110-121 | > 121 | 95 | 56-124 | | 125-133 | > 133 | 77 |
|  | LVESVi | 18-45 | 46-50 | > 50 | 95 | 18-52 | | 53-58 | > 58 | 75 |
|  | LVSVi | 35-71 | 31-34 | < 31 | 95 | 35-75 | | 28-34 | < 28 | 75 |
|  | LVEF | 52-75 | 50-51 | < 50 | 95 | 51-73 | | 48-50 | < 48 | 76 |
|  | LVCOi | 02-5 | 1.8-1.9 | < 1.8 | 94 | 2.4-4.9 | | 2.2-2.3 | < 2.2 | 75 |
|  | LVMi | 35-71 | 72-78 | > 78 | 95 | 42-88 | | 89-95 | > 95 | 78 |
|  | LVM/LVEDV | 0.5-0.8 | 0.9-1 | > 1 | 95 | 0.5-0.9 | | 01-1 | > 1 | 77 |
|  | RVEDVi | 59-117 | 118-128 | > 128 | 95 | 72-130 | | 131-143 | > 143 | 75 |
|  | RVESVi | 19-52 | 53-59 | > 59 | 95 | 21-58 | | 59-63 | > 63 | 76 |
|  | RVEF | 51-73 | 48-50 | < 48 | 95 | 47-69 | | 44-46 | < 44 | 77 |
|  | RVCOi | 2.1-5.2 | 1.9-2 | < 1.9 | 94 | 2.3-5.2 | | 1.9-2.2 | < 1.9 | 77 |
|  | RVSVi | 35-71 | 30-34 | < 30 | 95 | 32-76 | | 23-31 | < 23 | 75 |
|  | LAVi max | 25-63 | 64-71 | > 71 | 89 | 24-64 | | 65-72 | > 72 | 64 |
|  | LAEF | 54-75 | 51-53 | < 51 | 88 | 53-76 | | 50-52 | < 50 | 64 |
|  | RAVi max | 27-72 | 73-85 | > 85 | 94 | 33-74 | | 75-82 | > 82 | 73 |
|  | RAEF | 31-65 | 26-30 | < 26 | 93 | 35-63 | | 32-34 | < 32 | 71 |
| 60-69 | LVEDVi | 52-102 | 103-110 | > 110 | 66 | 59-123 | | 124-136 | > 136 | 48 |
|  | LVESVi | 15-43 | 44-48 | > 48 | 66 | 17-53 | | 54-60 | > 60 | 48 |
|  | LVSVi | 33-68 | 29-32 | < 29 | 66 | 36-75 | | 32-35 | < 32 | 48 |
|  | LVEF | 52-75 | 48-51 | < 48 | 66 | 45-73 | | 36-44 | < 36 | 48 |
|  | LVCOi | 2.1-4.8 | 1.9-2 | < 1.9 | 66 | 2.3-5 | | 2.1-2.2 | < 2.1 | 48 |
|  | LVMi | 35-71 | 72-78 | > 78 | 66 | 41-86 | | 87-92 | > 92 | 48 |
|  | LVM/LVEDV | 0.5-0.9 | 01-1 | > 1 | 66 | 0.6-1 | | 1.1-1.1 | > 1.1 | 48 |
|  | RVEDVi | 55-111 | 112-120 | > 120 | 66 | 69-130 | | 131-146 | > 146 | 48 |
|  | RVESVi | 14-47 | 48-51 | > 51 | 66 | 18-56 | | 57-61 | > 61 | 48 |
|  | RVEF | 50-73 | 46-49 | < 46 | 65 | 48-71 | | 44-47 | < 44 | 48 |
|  | RVCOi | 2.1-4.8 | 1.9-2 | < 1.9 | 66 | 2.3-5.2 | | 2.1-2.2 | < 2.1 | 48 |
|  | RVSVi | 34-70 | 30-33 | < 30 | 66 | 39-76 | | 35-38 | < 35 | 48 |
|  | LAVi max | 23-63 | 64-71 | > 71 | 59 | 23-65 | | 66-74 | > 74 | 42 |
|  | LAEF | 50-75 | 45-49 | < 45 | 58 | 50-75 | | 46-49 | < 46 | 43 |
|  | RAVi max | 25-67 | 68-76 | > 76 | 64 | 28-79 | | 80-88 | > 88 | 45 |
|  | RAEF | 32-67 | 27-31 | < 27 | 66 | 32-64 | | 29-31 | < 29 | 46 |
| 70+ | LVEDVi | 45-99 | 100-107 | > 107 | 18 | 56-118 | | 119-137 | > 137 | 14 |
|  | LVESVi | 12-41 | 42-46 | > 46 | 18 | 13-46 | | 47-52 | > 52 | 14 |
|  | LVSVi | 31-65 | 28-30 | < 28 | 18 | 36-78 | | 33-35 | < 33 | 14 |
|  | LVEF | 51-75 | 44-50 | < 44 | 18 | 43-74 | | 33-42 | < 33 | 14 |
|  | LVCOi | 2.1-4.5 | 1.9-2 | < 1.9 | 18 | 2.2-5.3 | | 2-2.1 | < 2 | 14 |
|  | LVMi | 35-71 | 72-78 | > 78 | 18 | 38-80 | | 81-86 | > 86 | 14 |
|  | LVM/LVEDV | 0.6-0.9 | 01-1 | > 1 | 18 | 0.6-1.1 | | 1.2-1.3 | > 1.3 | 14 |
|  | RVEDVi | 51-107 | 108-115 | > 115 | 18 | 66-131 | | 132-152 | > 152 | 14 |
|  | RVESVi | 13-44 | 45-47 | > 47 | 18 | 16-54 | | 55-59 | > 59 | 14 |
|  | RVEF | 48-73 | 42-47 | < 42 | 18 | 48-74 | | 44-47 | < 44 | 14 |
|  | RVCOi | 2.2-4.6 | 1.9-2.1 | < 1.9 | 18 | 2.5-5.1 | | 2.3-2.4 | < 2.3 | 14 |
|  | RVSVi | 33-69 | 30-32 | < 30 | 18 | 41-77 | | 38-40 | < 38 | 14 |
|  | LAVi max | 21-63 | 64-71 | > 71 | 15 |  | |  |  |  |
|  | LAEF | 44-74 | 35-43 | < 35 | 16 |  | |  |  |  |
|  | RAVi max | 23-65 | 66-72 | > 72 | 17 | 22-84 | | 85-95 | > 95 | 12 |
|  | RAEF | 33-68 | 28-32 | < 28 | 17 | 29-66 | | 25-28 | < 25 | 12 |

Footnote: All measurements were derived using cvi42 software with papillary endocardial segmentation; papillary muscles were included in left ventricular mass measurements. Volumetric and mass measurements were indexed to height. Reference ranges are defined as the interval between the 2.5th and 97.5th centiles derived from age-specific generalized additive models for location, scale and shape (GAMLSS) using the Box–Cox Cole–Green distribution. Centile-based categories beyond the reference range are derived from LMS-based thresholds (P0.5, P2.5, P97.5, and P99.5) and describe the position of a measurement within the reference distribution rather than disease severity or diagnostic abnormality. Reference ranges are based on a Black adult population.

Abbreviations: LV = left ventricle; RV = right ventricle; LA = left atrium; RA = right atrium; EDV = end-diastolic volume; ESV = end-systolic volume; SV = stroke volume; EF = ejection fraction; CO = cardiac output.

### 13. Table: Reference ranges and centile-based categories for cardiac chamber measurements derived using cvi42 software (papillary segmentation), indexed to height, in an Asian adult population

|  |  | **Female** | | | | **Male** | | | |
| --- | --- | --- | --- | --- | --- | --- | --- | --- | --- |
| **Age group** | **Metrics** | **Reference range** | **Mild** | **At least moderate** | **n** | **Reference range** | **Mild** | **At least moderate** | **n** |
| 40-49 | LVEDVi | 52-88 | 89-94 | > 94 | 10 | 61-109 | 110-119 | > 119 | 20 |
|  | LVESVi | 16-40 | 41-48 | > 48 | 10 | 19-47 | 48-52 | > 52 | 20 |
|  | LVSVi | 34-60 | 31-33 | < 31 | 10 | 40-62 | 37-39 | < 37 | 20 |
|  | LVEF | 53-73 | 47-52 | < 47 | 10 | 54-77 | 52-53 | < 52 | 20 |
|  | LVCOi | 2.1-4.2 | 1.9-2 | < 1.9 | 10 | 2.2-4.6 | 2.1-2.1 | < 2.1 | 20 |
|  | LVMi | 31-55 | 56-60 | > 60 | 10 | 39-72 | 73-78 | > 78 | 20 |
|  | LVM/LVEDV | 0.5-0.8 | 0.9-0.9 | > 0.9 | 10 | 0.6-0.8 | 0.9-0.9 | > 0.9 | 20 |
|  | RVEDVi | 51-94 | 95-104 | > 104 | 10 | 64-122 | 123-136 | > 136 | 20 |
|  | RVESVi | 16-40 | 41-46 | > 46 | 10 | 22-52 | 53-57 | > 57 | 20 |
|  | RVEF | 52-72 | 48-51 | < 48 | 10 | 49-67 | 46-48 | < 46 | 20 |
|  | RVCOi | 2-3.9 | 1.8-1.9 | < 1.8 | 10 | 2.3-4.4 | 2.1-2.2 | < 2.1 | 20 |
|  | RVSVi | 32-60 | 29-31 | < 29 | 10 | 35-72 | 31-34 | < 31 | 20 |
|  | LAVi max | 23-51 | 52-56 | > 56 | 10 | 20-57 | 58-64 | > 64 | 20 |
|  | LAEF | 57-81 | 54-56 | < 54 | 10 | 58-84 | 55-57 | < 55 | 20 |
|  | RAVi max | 22-47 | 48-51 | > 51 | 10 | 23-70 | 71-79 | > 79 | 20 |
|  | RAEF | 37-71 | 35-36 | < 35 | 10 | 32-63 | 27-31 | < 27 | 20 |
| 50-59 | LVEDVi | 51-88 | 89-95 | > 95 | 83 | 57-107 | 108-117 | > 117 | 133 |
|  | LVESVi | 15-38 | 39-46 | > 46 | 83 | 17-46 | 47-52 | > 52 | 133 |
|  | LVSVi | 33-60 | 30-32 | < 30 | 83 | 34-68 | 29-33 | < 29 | 133 |
|  | LVEF | 53-74 | 48-52 | < 48 | 83 | 52-73 | 49-51 | < 49 | 131 |
|  | LVCOi | 2-4.2 | 1.8-1.9 | < 1.8 | 83 | 2.1-4.6 | 1.9-2 | < 1.9 | 133 |
|  | LVMi | 31-56 | 57-63 | > 63 | 83 | 39-75 | 76-84 | > 84 | 133 |
|  | LVM/LVEDV | 0.5-0.8 | 0.9-0.9 | > 0.9 | 83 | 0.6-0.9 | 01-1 | > 1 | 133 |
|  | RVEDVi | 50-95 | 96-106 | > 106 | 83 | 60-118 | 119-130 | > 130 | 133 |
|  | RVESVi | 16-39 | 40-45 | > 45 | 83 | 21-51 | 52-57 | > 57 | 133 |
|  | RVEF | 53-73 | 49-52 | < 49 | 83 | 48-68 | 45-47 | < 45 | 133 |
|  | RVCOi | 1.9-4 | 1.7-1.8 | < 1.7 | 83 | 2.1-4.4 | 1.9-2 | < 1.9 | 133 |
|  | RVSVi | 31-61 | 29-30 | < 29 | 83 | 33-70 | 28-32 | < 28 | 133 |
|  | LAVi max | 22-52 | 53-58 | > 58 | 77 | 21-58 | 59-65 | > 65 | 121 |
|  | LAEF | 54-80 | 51-53 | < 51 | 75 | 55-82 | 52-54 | < 52 | 120 |
|  | RAVi max | 22-53 | 54-58 | > 58 | 76 | 23-69 | 70-79 | > 79 | 128 |
|  | RAEF | 37-69 | 34-36 | < 34 | 76 | 34-65 | 29-33 | < 29 | 127 |
| 60-69 | LVEDVi | 49-89 | 90-99 | > 99 | 63 | 50-103 | 104-113 | > 113 | 118 |
|  | LVESVi | 14-36 | 37-43 | > 43 | 63 | 16-44 | 45-51 | > 51 | 118 |
|  | LVSVi | 31-61 | 29-30 | < 29 | 63 | 32-69 | 27-31 | < 27 | 118 |
|  | LVEF | 54-77 | 50-53 | < 50 | 63 | 51-74 | 46-50 | < 46 | 118 |
|  | LVCOi | 1.9-4.4 | 1.7-1.8 | < 1.7 | 63 | 1.9-4.5 | 1.6-1.8 | < 1.6 | 119 |
|  | LVMi | 31-60 | 61-68 | > 68 | 63 | 37-78 | 79-90 | > 90 | 119 |
|  | LVM/LVEDV | 0.5-0.8 | 0.9-0.9 | > 0.9 | 63 | 0.6-0.9 | 1-1.1 | > 1.1 | 118 |
|  | RVEDVi | 49-97 | 98-110 | > 110 | 63 | 53-112 | 113-122 | > 122 | 119 |
|  | RVESVi | 15-38 | 39-43 | > 43 | 63 | 19-50 | 51-57 | > 57 | 119 |
|  | RVEF | 54-73 | 51-53 | < 51 | 63 | 47-70 | 42-46 | < 42 | 119 |
|  | RVCOi | 1.8-4.3 | 1.6-1.7 | < 1.6 | 63 | 1.8-4.4 | 1.5-1.7 | < 1.5 | 119 |
|  | RVSVi | 30-62 | 27-29 | < 27 | 63 | 30-67 | 25-29 | < 25 | 119 |
|  | LAVi max | 22-54 | 55-60 | > 60 | 59 | 21-59 | 60-66 | > 66 | 110 |
|  | LAEF | 50-78 | 46-49 | < 46 | 58 | 51-80 | 48-50 | < 48 | 107 |
|  | RAVi max | 21-61 | 62-68 | > 68 | 61 | 23-68 | 69-77 | > 77 | 113 |
|  | RAEF | 36-68 | 31-35 | < 31 | 59 | 35-70 | 31-34 | < 31 | 114 |
| 70+ | LVEDVi | 47-92 | 93-105 | > 105 | 28 | 43-100 | 101-108 | > 108 | 41 |
|  | LVESVi | 13-34 | 35-41 | > 41 | 28 | 14-42 | 43-50 | > 50 | 41 |
|  | LVSVi | 30-62 | 28-29 | < 28 | 28 | 33-64 | 30-32 | < 30 | 40 |
|  | LVEF | 55-79 | 51-54 | < 51 | 28 | 52-76 | 47-51 | < 47 | 41 |
|  | LVCOi | 1.8-4.5 | 1.6-1.7 | < 1.6 | 28 | 1.6-4.4 | 1.1-1.5 | < 1.1 | 41 |
|  | LVMi | 31-64 | 65-77 | > 77 | 28 | 31-72 | 73-79 | > 79 | 41 |
|  | LVM/LVEDV | 0.5-0.8 | 0.9-0.9 | > 0.9 | 28 | 0.6-1.1 | 1.2-1.3 | > 1.3 | 41 |
|  | RVEDVi | 48-99 | 100-115 | > 115 | 28 | 45-106 | 107-115 | > 115 | 41 |
|  | RVESVi | 15-37 | 38-42 | > 42 | 28 | 17-50 | 51-58 | > 58 | 41 |
|  | RVEF | 55-74 | 52-54 | < 52 | 28 | 46-72 | 39-45 | < 39 | 40 |
|  | RVCOi | 1.8-4.7 | 1.6-1.7 | < 1.6 | 28 | 1.4-4.4 | 0.9-1.3 | < 0.9 | 41 |
|  | RVSVi | 29-64 | 26-28 | < 26 | 28 | 28-65 | 22-27 | < 22 | 40 |
|  | LAVi max | 22-55 | 56-63 | > 63 | 28 | 21-60 | 61-67 | > 67 | 34 |
|  | LAEF | 45-76 | 40-44 | < 40 | 27 | 48-79 | 45-47 | < 45 | 34 |
|  | RAVi max | 17-65 | 66-76 | > 76 | 27 | 22-67 | 68-76 | > 76 | 38 |
|  | RAEF | 35-70 | 25-34 | < 25 | 27 | 36-77 | 32-35 | < 32 | 39 |

Footnote: All measurements were derived using cvi42 software with papillary endocardial segmentation; papillary muscles were included in left ventricular mass measurements. Volumetric and mass measurements were indexed to height. Reference ranges are defined as the interval between the 2.5th and 97.5th centiles derived from age-specific generalized additive models for location, scale and shape (GAMLSS) using the Box–Cox Cole–Green distribution. Centile-based categories beyond the reference range are derived from LMS-based thresholds (P0.5, P2.5, P97.5, and P99.5) and describe the position of a measurement within the reference distribution rather than disease severity or diagnostic abnormality. Reference ranges are based on an Asian adult population.

Abbreviations: LV = left ventricle; RV = right ventricle; LA = left atrium; RA = right atrium; EDV = end-diastolic volume; ESV = end-systolic volume; SV = stroke volume; EF = ejection fraction; CO = cardiac output.

### 14. Table: Reference ranges and centile-based categories for cardiac chamber measurements derived using cvi42 software (papillary segmentation), indexed to height, in a Chinese adult population

|  |  | **Female** | | | | **Male** | | | |
| --- | --- | --- | --- | --- | --- | --- | --- | --- | --- |
| **Age group** | **Metrics** | **Reference range** | **Mild** | **At least moderate** | **n** | **Reference range** | **Mild** | **At least moderate** | **n** |
| 18-29 | LVEDVi | 55-97 | 98-106 | > 106 | 16 | 65-106 | 107-110 | > 110 | 19 |
|  | LVESVi | 16-39 | 40-45 | > 45 | 16 | 23-44 | 45-46 | > 46 | 19 |
|  | LVSVi | 33-69 | 30-32 | < 30 | 16 | 37-68 | 28-36 | < 28 | 19 |
|  | LVEF | 54-76 | 51-53 | < 51 | 16 | 53-70 | 51-52 | < 51 | 19 |
|  | LVCOi | 2.1-4.8 | 1.9-2 | < 1.9 | 16 | 2.7-5 | 2.4-2.6 | < 2.4 | 19 |
|  | LVMi | 28-49 | 50-54 | > 54 | 16 | 37-73 | 74-78 | > 78 | 19 |
|  | LVM/LVEDV | 0.4-0.6 |  | > 0.6 | 16 | 0.5-0.8 | 0.9-0.9 | > 0.9 | 19 |
|  | RVEDVi | 47-93 | 94-100 | > 100 | 16 | 60-109 | 110-113 | > 113 | 19 |
|  | RVESVi | 13-38 | 39-40 | > 40 | 16 | 19-49 | 50-51 | > 51 | 19 |
|  | RVEF | 49-78 | 46-48 | < 46 | 16 | 47-70 | 43-46 | < 43 | 19 |
|  | RVCOi | 1.8-4.4 | 1.5-1.7 | < 1.5 | 16 | 2.4-4.8 | 2-2.3 | < 2 | 19 |
|  | RVSVi | 29-62 | 25-28 | < 25 | 16 | 37-69 | 32-36 | < 32 | 19 |
|  | LAVi max | 22-52 | 53-58 | > 58 | 13 | 27-52 | 53-56 | > 56 | 16 |
|  | LAEF | 54-77 | 50-53 | < 50 | 13 | 52-70 | 49-51 | < 49 | 16 |
|  | RAVi max | 19-48 | 49-57 | > 57 | 15 | 20-62 | 63-69 | > 69 | 18 |
|  | RAEF | 41-73 | 35-40 | < 35 | 15 | 38-68 | 31-37 | < 31 | 18 |
| 30-39 | LVEDVi | 54-95 | 96-104 | > 104 | 18 | 63-104 | 105-109 | > 109 | 17 |
|  | LVESVi | 15-38 | 39-44 | > 44 | 18 | 21-43 | 44-46 | > 46 | 17 |
|  | LVSVi | 34-66 | 32-33 | < 32 | 18 | 37-66 | 30-36 | < 30 | 17 |
|  | LVEF | 55-76 | 51-54 | < 51 | 18 | 53-70 | 50-52 | < 50 | 17 |
|  | LVCOi | 2.1-4.6 | 1.9-2 | < 1.9 | 18 | 2.6-4.9 | 2.3-2.5 | < 2.3 | 17 |
|  | LVMi | 29-51 | 52-57 | > 57 | 18 | 39-74 | 75-79 | > 79 | 17 |
|  | LVM/LVEDV | 0.4-0.6 | 0.7-0.7 | > 0.7 | 18 | 0.5-0.8 | 0.9-0.9 | > 0.9 | 17 |
|  | RVEDVi | 49-94 | 95-102 | > 102 | 18 | 60-108 | 109-113 | > 113 | 17 |
|  | RVESVi | 9-39 | 40-42 | > 42 | 18 | 18-48 | 49-51 | > 51 | 17 |
|  | RVEF | 50-77 | 46-49 | < 46 | 18 | 47-70 | 44-46 | < 44 | 17 |
|  | RVCOi | 1.8-4.2 | 1.6-1.7 | < 1.6 | 18 | 2.3-4.7 | 1.9-2.2 | < 1.9 | 17 |
|  | RVSVi | 29-61 | 26-28 | < 26 | 18 | 35-68 | 30-34 | < 30 | 17 |
|  | LAVi max | 22-51 | 52-57 | > 57 | 18 | 26-52 | 53-56 | > 56 | 14 |
|  | LAEF | 53-76 | 50-52 | < 50 | 18 | 51-72 | 48-50 | < 48 | 14 |
|  | RAVi max | 20-51 | 52-61 | > 61 | 18 | 20-63 | 64-71 | > 71 | 16 |
|  | RAEF | 41-72 | 35-40 | < 35 | 18 | 38-68 | 31-37 | < 31 | 16 |
| 40-49 | LVEDVi | 53-93 | 94-102 | > 102 | 24 | 61-102 | 103-108 | > 108 | 20 |
|  | LVESVi | 15-37 | 38-43 | > 43 | 24 | 18-42 | 43-45 | > 45 | 20 |
|  | LVSVi | 35-63 | 33-34 | < 33 | 24 | 37-65 | 32-36 | < 32 | 20 |
|  | LVEF | 55-77 | 51-54 | < 51 | 24 | 53-71 | 50-52 | < 50 | 19 |
|  | LVCOi | 2.1-4.4 | 1.9-2 | < 1.9 | 24 | 2.4-4.8 | 2.2-2.3 | < 2.2 | 20 |
|  | LVMi | 30-54 | 55-61 | > 61 | 24 | 40-74 | 75-81 | > 81 | 20 |
|  | LVM/LVEDV | 0.5-0.7 |  | > 0.7 | 24 | 0.5-0.9 | 01-1 | > 1 | 20 |
|  | RVEDVi | 50-95 | 96-106 | > 106 | 24 | 61-107 | 108-113 | > 113 | 20 |
|  | RVESVi | 12-41 | 42-47 | > 47 | 24 | 17-48 | 49-51 | > 51 | 20 |
|  | RVEF | 51-75 | 47-50 | < 47 | 24 | 48-70 | 45-47 | < 45 | 19 |
|  | RVCOi | 1.8-4 | 1.6-1.7 | < 1.6 | 24 | 2.1-4.7 | 1.8-2 | < 1.8 | 20 |
|  | RVSVi | 30-60 | 26-29 | < 26 | 24 | 33-67 | 28-32 | < 28 | 20 |
|  | LAVi max | 22-51 | 52-56 | > 56 | 22 | 24-52 | 53-56 | > 56 | 18 |
|  | LAEF | 53-75 | 49-52 | < 49 | 22 | 51-74 | 47-50 | < 47 | 18 |
|  | RAVi max | 22-54 | 55-65 | > 65 | 23 | 21-65 | 66-72 | > 72 | 19 |
|  | RAEF | 41-70 | 36-40 | < 36 | 23 | 37-67 | 30-36 | < 30 | 19 |
| 50-59 | LVEDVi | 51-91 | 92-101 | > 101 | 66 | 59-101 | 102-108 | > 108 | 49 |
|  | LVESVi | 14-36 | 37-42 | > 42 | 66 | 15-41 | 42-44 | > 44 | 49 |
|  | LVSVi | 35-60 | 32-34 | < 32 | 66 | 36-65 | 32-35 | < 32 | 49 |
|  | LVEF | 55-77 | 51-54 | < 51 | 66 | 53-72 | 50-52 | < 50 | 48 |
|  | LVCOi | 2-4.2 | 1.8-1.9 | < 1.8 | 66 | 2.3-4.7 | 2.1-2.2 | < 2.1 | 49 |
|  | LVMi | 31-57 | 58-67 | > 67 | 66 | 40-74 | 75-82 | > 82 | 49 |
|  | LVM/LVEDV | 0.5-0.8 |  | > 0.8 | 66 | 0.5-0.9 | 01-1 | > 1 | 49 |
|  | RVEDVi | 50-95 | 96-107 | > 107 | 66 | 62-108 | 109-116 | > 116 | 49 |
|  | RVESVi | 15-41 | 42-49 | > 49 | 66 | 17-48 | 49-51 | > 51 | 49 |
|  | RVEF | 52-74 | 48-51 | < 48 | 66 | 48-70 | 46-47 | < 46 | 49 |
|  | RVCOi | 1.8-3.9 | 1.6-1.7 | < 1.6 | 66 | 2-4.6 | 1.7-1.9 | < 1.7 | 49 |
|  | RVSVi | 30-59 | 26-29 | < 26 | 66 | 31-66 | 26-30 | < 26 | 49 |
|  | LAVi max | 21-50 | 51-56 | > 56 | 60 | 23-51 | 52-55 | > 55 | 40 |
|  | LAEF | 51-76 | 48-50 | < 48 | 58 | 50-77 | 46-49 | < 46 | 40 |
|  | RAVi max | 23-57 | 58-69 | > 69 | 63 | 21-66 | 67-74 | > 74 | 46 |
|  | RAEF | 40-68 | 36-39 | < 36 | 63 | 37-66 | 30-36 | < 30 | 46 |
| 60-69 | LVEDVi | 50-89 | 90-99 | > 99 | 52 | 56-101 | 102-110 | > 110 | 37 |
|  | LVESVi | 13-34 | 35-40 | > 40 | 52 | 13-41 | 42-44 | > 44 | 37 |
|  | LVSVi | 33-60 | 30-32 | < 30 | 52 | 35-65 | 32-34 | < 32 | 37 |
|  | LVEF | 55-78 | 50-54 | < 50 | 52 | 53-73 | 50-52 | < 50 | 35 |
|  | LVCOi | 02-4 | 1.8-1.9 | < 1.8 | 52 | 2.2-4.6 | 2-2.1 | < 2 | 37 |
|  | LVMi | 32-61 | 62-77 | > 77 | 52 | 40-75 | 76-85 | > 85 | 37 |
|  | LVM/LVEDV | 0.5-0.8 | 0.9-1 | > 1 | 52 | 0.6-1 | 1.1-1.1 | > 1.1 | 37 |
|  | RVEDVi | 47-90 | 91-99 | > 99 | 52 | 62-111 | 112-124 | > 124 | 36 |
|  | RVESVi | 14-36 | 37-41 | > 41 | 52 | 17-48 | 49-51 | > 51 | 37 |
|  | RVEF | 53-74 | 49-52 | < 49 | 52 | 49-70 | 46-48 | < 46 | 36 |
|  | RVCOi | 1.8-3.7 | 1.6-1.7 | < 1.6 | 52 | 1.9-4.5 | 1.6-1.8 | < 1.6 | 37 |
|  | RVSVi | 30-58 | 27-29 | < 27 | 52 | 29-65 | 24-28 | < 24 | 37 |
|  | LAVi max | 21-51 | 52-56 | > 56 | 47 | 20-49 | 50-53 | > 53 | 30 |
|  | LAEF | 49-76 | 45-48 | < 45 | 46 | 48-80 | 44-47 | < 44 | 30 |
|  | RAVi max | 24-61 | 62-73 | > 73 | 46 | 22-68 | 69-76 | > 76 | 35 |
|  | RAEF | 40-66 | 36-39 | < 36 | 46 | 38-68 | 31-37 | < 31 | 35 |
| 70+ | LVEDVi | 49-87 | 88-98 | > 98 | 12 |  |  |  |  |
|  | LVESVi | 12-33 | 34-39 | > 39 | 12 |  |  |  |  |
|  | LVSVi | 29-61 | 24-28 | < 24 | 12 |  |  |  |  |
|  | LVEF | 55-79 | 50-54 | < 50 | 12 |  |  |  |  |
|  | LVCOi | 2-3.8 | 1.8-1.9 | < 1.8 | 12 |  |  |  |  |
|  | LVMi | 33-66 | 67-92 | > 92 | 12 |  |  |  |  |
|  | LVM/LVEDV | 0.5-1 | 1.1-1.3 | > 1.3 | 12 |  |  |  |  |
|  | RVEDVi | 44-86 | 87-92 | > 92 | 12 |  |  |  |  |
|  | RVESVi | 12-32 | 33-35 | > 35 | 12 |  |  |  |  |
|  | RVEF | 54-74 | 50-53 | < 50 | 12 |  |  |  |  |
|  | RVCOi | 1.9-3.6 | 1.7-1.8 | < 1.7 | 12 |  |  |  |  |
|  | RVSVi | 30-57 | 27-29 | < 27 | 12 |  |  |  |  |
|  | LAVi max | 20-51 | 52-57 | > 57 | 11 |  |  |  |  |
|  | LAEF | 47-78 | 42-46 | < 42 | 12 |  |  |  |  |
|  | RAVi max | 25-64 | 65-76 | > 76 | 11 |  |  |  |  |
|  | RAEF | 39-65 | 36-38 | < 36 | 11 |  |  |  |  |

Footnote: All measurements were derived using cvi42 software with papillary endocardial segmentation; papillary muscles were included in left ventricular mass measurements. Volumetric and mass measurements were indexed to height. Reference ranges are defined as the interval between the 2.5th and 97.5th centiles derived from age-specific generalized additive models for location, scale and shape (GAMLSS) using the Box–Cox Cole–Green distribution. Centile-based categories beyond the reference range are derived from LMS-based thresholds (P0.5, P2.5, P97.5, and P99.5) and describe the position of a measurement within the reference distribution rather than disease severity or diagnostic abnormality. Reference ranges are based on Chinese adult population.

Abbreviations: LV = left ventricle; RV = right ventricle; LA = left atrium; RA = right atrium; EDV = end-diastolic volume; ESV = end-systolic volume; SV = stroke volume; EF = ejection fraction; CO = cardiac output

### 15. Table: Reference ranges and centile-based categories for cardiac chamber measurements derived using cvi42 software (papillary segmentation), indexed to height, in a Mixed adult population

|  |  | **Female** | | | | **Male** | | | |
| --- | --- | --- | --- | --- | --- | --- | --- | --- | --- |
| **Age group** | **Metrics** | **Reference range** | **Mild** | **At least moderate** | **n** | **Reference range** | **Mild** | **At least moderate** | **n** |
| 50-59 | LVEDVi | 56-102 | 103-111 | > 111 | 134 | 64-114 | 115-120 | > 120 | 65 |
|  | LVESVi | 17-39 | 40-44 | > 44 | 134 | 17-47 | 48-50 | > 50 | 66 |
|  | LVSVi | 35-71 | 32-34 | < 32 | 133 | 38-76 | 31-37 | < 31 | 65 |
|  | LVEF | 54-76 | 51-53 | < 51 | 134 | 52-74 | 48-51 | < 48 | 66 |
|  | LVCOi | 2.1-5 | 02-2 | < 2 | 132 | 2.2-5.3 | 1.9-2.1 | < 1.9 | 66 |
|  | LVMi | 32-66 | 67-75 | > 75 | 134 | 43-89 | 90-100 | > 100 | 65 |
|  | LVM/LVEDV | 0.5-0.8 |  | > 0.8 | 134 | 0.5-0.9 | 1-1.1 | > 1.1 | 64 |
|  | RVEDVi | 54-110 | 111-121 | > 121 | 134 | 68-130 | 131-140 | > 140 | 65 |
|  | RVESVi | 17-44 | 45-48 | > 48 | 134 | 25-55 | 56-59 | > 59 | 65 |
|  | RVEF | 53-72 | 50-52 | < 50 | 134 | 51-70 | 48-50 | < 48 | 66 |
|  | RVCOi | 2-4.9 | 1.8-1.9 | < 1.8 | 133 | 2.3-5.4 | 2-2.2 | < 2 | 66 |
|  | RVSVi | 33-67 | 28-32 | < 28 | 133 | 39-81 | 32-38 | < 32 | 66 |
|  | LAVi max | 23-55 | 56-60 | > 60 | 121 | 25-70 | 71-79 | > 79 | 53 |
|  | LAEF | 55-78 | 52-54 | < 52 | 120 | 54-78 | 51-53 | < 51 | 53 |
|  | RAVi max | 24-66 | 67-75 | > 75 | 128 | 26-79 | 80-88 | > 88 | 64 |
|  | RAEF | 37-64 | 31-36 | < 31 | 124 | 35-63 | 31-34 | < 31 | 64 |
| 60-69 | LVEDVi | 51-100 | 101-110 | > 110 | 110 | 60-111 | 112-120 | > 120 | 59 |
|  | LVESVi | 13-39 | 40-43 | > 43 | 110 | 17-48 | 49-53 | > 53 | 58 |
|  | LVSVi | 34-66 | 31-33 | < 31 | 110 | 37-71 | 31-36 | < 31 | 59 |
|  | LVEF | 54-76 | 50-53 | < 50 | 108 | 51-74 | 47-50 | < 47 | 58 |
|  | LVCOi | 2.1-4.5 | 1.9-2 | < 1.9 | 110 | 2.2-4.9 | 2-2.1 | < 2 | 59 |
|  | LVMi | 32-62 | 63-68 | > 68 | 110 | 42-86 | 87-96 | > 96 | 59 |
|  | LVM/LVEDV | 0.5-0.8 | 0.9-0.9 | > 0.9 | 110 | 0.5-1 | 1.1-1.1 | > 1.1 | 59 |
|  | RVEDVi | 50-107 | 108-118 | > 118 | 110 | 62-126 | 127-136 | > 136 | 58 |
|  | RVESVi | 17-43 | 44-48 | > 48 | 110 | 20-54 | 55-59 | > 59 | 58 |
|  | RVEF | 53-72 | 49-52 | < 49 | 110 | 50-69 | 47-49 | < 47 | 58 |
|  | RVCOi | 1.9-4.5 | 1.7-1.8 | < 1.7 | 110 | 2.2-5.1 | 2-2.1 | < 2 | 59 |
|  | RVSVi | 31-68 | 27-30 | < 27 | 110 | 36-76 | 31-35 | < 31 | 59 |
|  | LAVi max | 23-58 | 59-65 | > 65 | 100 | 24-65 | 66-74 | > 74 | 52 |
|  | LAEF | 52-79 | 49-51 | < 49 | 98 | 50-79 | 47-49 | < 47 | 52 |
|  | RAVi max | 23-68 | 69-77 | > 77 | 103 | 20-78 | 79-85 | > 85 | 57 |
|  | RAEF | 35-68 | 29-34 | < 29 | 103 | 36-68 | 33-35 | < 33 | 57 |
| 70+ | LVEDVi | 51-91 | 92-99 | > 99 | 51 | 56-112 | 113-125 | > 125 | 33 |
|  | LVESVi | 9-38 | 39-42 | > 42 | 51 | 18-53 | 54-64 | > 64 | 34 |
|  | LVSVi | 32-59 | 29-31 | < 29 | 50 | 35-68 | 31-34 | < 31 | 34 |
|  | LVEF | 54-76 | 49-53 | < 49 | 51 | 50-73 | 47-49 | < 47 | 34 |
|  | LVCOi | 2-4.1 | 1.8-1.9 | < 1.8 | 51 | 2.2-4.6 | 2-2.1 | < 2 | 35 |
|  | LVMi | 32-59 | 60-63 | > 63 | 51 | 41-83 | 84-93 | > 93 | 35 |
|  | LVM/LVEDV | 0.5-0.8 | 0.9-0.9 | > 0.9 | 51 | 0.5-1 | 1.1-1.2 | > 1.2 | 33 |
|  | RVEDVi | 55-91 | 92-98 | > 98 | 51 | 58-122 | 123-134 | > 134 | 35 |
|  | RVESVi | 17-41 | 42-47 | > 47 | 51 | 15-54 | 55-59 | > 59 | 35 |
|  | RVEF | 52-72 | 48-51 | < 48 | 51 | 49-69 | 46-48 | < 46 | 35 |
|  | RVCOi | 1.9-4 | 1.7-1.8 | < 1.7 | 51 | 2.1-4.7 | 1.9-2 | < 1.9 | 35 |
|  | RVSVi | 34-60 | 31-33 | < 31 | 50 | 33-74 | 29-32 | < 29 | 35 |
|  | LAVi max | 24-62 | 63-72 | > 72 | 45 | 22-61 | 62-70 | > 70 | 32 |
|  | LAEF | 50-80 | 47-49 | < 47 | 44 | 46-81 | 42-45 | < 42 | 30 |
|  | RAVi max | 21-68 | 69-77 | > 77 | 47 | 18-76 | 77-83 | > 83 | 31 |
|  | RAEF | 33-72 | 28-32 | < 28 | 46 | 36-76 | 34-35 | < 34 | 31 |

Footnote: All measurements were derived using cvi42 software with papillary endocardial segmentation; papillary muscles were included in left ventricular mass measurements. Volumetric and mass measurements were indexed to height. Reference ranges are defined as the interval between the 2.5th and 97.5th centiles derived from age-specific generalized additive models for location, scale and shape (GAMLSS) using the Box–Cox Cole–Green distribution. Centile-based categories beyond the reference range are derived from LMS-based thresholds (P0.5, P2.5, P97.5, and P99.5) and describe the position of a measurement within the reference distribution rather than disease severity or diagnostic abnormality. Reference ranges are based on Mixed adult population.

Abbreviations: LV = left ventricle; RV = right ventricle; LA = left atrium; RA = right atrium; EDV = end-diastolic volume; ESV = end-systolic volume; SV = stroke volume; EF = ejection fraction; CO = cardiac output

### 16. Table: Reference ranges and centile-based categories for cardiac chamber measurements derived using suiteHEART software (smooth segmentation), indexed to height, in a White adult population

|  |  | **Female** | | | | **Male** | | | |
| --- | --- | --- | --- | --- | --- | --- | --- | --- | --- |
| **Age group** | **Metrics** | **Reference range** | **Mild** | **At least moderate** | **n** | **Reference range** | **Mild** | **At least moderate** | **n** |
| 18-29 | LVEDVi | 63-111 | 112-118 | > 118 | 101 | 80-130 | 131-137 | > 137 | 95 |
|  | LVESVi | 25-51 | 52-57 | > 57 | 101 | 31-58 | 59-62 | > 62 | 84 |
|  | LVSVi | 36-68 | 30-35 | < 30 | 101 | 43-78 | 37-42 | < 37 | 100 |
|  | LVEF | 50-68 | 47-49 | < 47 | 101 | 47-67 | 44-46 | < 44 | 104 |
|  | LVMi | 33-58 | 59-64 | > 64 | 101 | 46-86 | 87-93 | > 93 | 104 |
|  | LVM/LVEDV | 0.4-0.7 | 0.8-0.8 | > 0.8 | 101 | 0.5-0.8 | 0.9-1 | > 1 | 95 |
|  | RVEDVi | 61-118 | 119-126 | > 126 | 101 | 79-141 | 142-150 | > 150 | 93 |
|  | RVESVi | 25-54 | 55-61 | > 61 | 100 | 31-66 | 67-70 | > 70 | 90 |
|  | RVEF | 47-68 | 44-46 | < 44 | 99 | 45-66 | 42-44 | < 42 | 102 |
|  | RVSVi | 34-72 | 30-33 | < 30 | 100 | 41-81 | 35-40 | < 35 | 97 |
|  | LAESV | 19-43 | 44-47 | > 47 | 65 | 22-55 | 56-62 | > 62 | 64 |
|  | LAEF | 57-85 | 52-56 | < 52 | 65 | 55-79 | 51-54 | < 51 | 62 |
|  | RAESV | 24-50 | 51-55 | > 55 | 77 | 26-67 | 68-75 | > 75 | 87 |
|  | RAEF | 42-75 | 38-41 | < 38 | 72 | 35-71 | 31-34 | < 31 | 85 |
| 30-39 | LVEDVi | 63-111 | 112-119 | > 119 | 131 | 73-124 | 125-131 | > 131 | 127 |
|  | LVESVi | 24-50 | 51-55 | > 55 | 131 | 29-57 | 58-61 | > 61 | 123 |
|  | LVSVi | 35-66 | 30-34 | < 30 | 130 | 40-74 | 34-39 | < 34 | 128 |
|  | LVEF | 50-68 | 47-49 | < 47 | 131 | 48-68 | 45-47 | < 45 | 129 |
|  | LVMi | 34-63 | 64-69 | > 69 | 131 | 46-87 | 88-95 | > 95 | 128 |
|  | LVM/LVEDV | 0.4-0.8 | 0.9-0.9 | > 0.9 | 131 | 0.5-0.9 | 01-1 | > 1 | 128 |
|  | RVEDVi | 60-117 | 118-127 | > 127 | 131 | 76-139 | 140-148 | > 148 | 125 |
|  | RVESVi | 23-52 | 53-59 | > 59 | 131 | 29-64 | 65-70 | > 70 | 126 |
|  | RVEF | 48-69 | 45-47 | < 45 | 131 | 45-67 | 42-44 | < 42 | 128 |
|  | RVSVi | 34-71 | 30-33 | < 30 | 131 | 40-82 | 34-39 | < 34 | 125 |
|  | LAESV | 20-48 | 49-53 | > 53 | 95 | 21-56 | 57-63 | > 63 | 86 |
|  | LAEF | 57-84 | 53-56 | < 53 | 95 | 55-80 | 51-54 | < 51 | 87 |
|  | RAESV | 24-53 | 54-59 | > 59 | 113 | 26-68 | 69-76 | > 76 | 104 |
|  | RAEF | 41-75 | 37-40 | < 37 | 108 | 36-72 | 31-35 | < 31 | 103 |
| 40-49 | LVEDVi | 63-113 | 114-121 | > 121 | 521 | 72-127 | 128-136 | > 136 | 423 |
|  | LVESVi | 22-48 | 49-54 | > 54 | 521 | 26-56 | 57-61 | > 61 | 422 |
|  | LVSVi | 36-68 | 31-35 | < 31 | 521 | 40-77 | 34-39 | < 34 | 430 |
|  | LVEF | 51-70 | 48-50 | < 48 | 520 | 49-69 | 46-48 | < 46 | 443 |
|  | LVMi | 34-65 | 66-72 | > 72 | 521 | 47-89 | 90-98 | > 98 | 441 |
|  | LVM/LVEDV | 0.4-0.8 | 0.9-0.9 | > 0.9 | 521 | 0.5-0.9 | 1-1.1 | > 1.1 | 420 |
|  | RVEDVi | 60-117 | 118-128 | > 128 | 521 | 76-141 | 142-152 | > 152 | 428 |
|  | RVESVi | 22-51 | 52-57 | > 57 | 521 | 28-63 | 64-69 | > 69 | 422 |
|  | RVEF | 49-71 | 46-48 | < 46 | 518 | 45-69 | 42-44 | < 42 | 442 |
|  | RVSVi | 34-73 | 30-33 | < 30 | 519 | 39-82 | 32-38 | < 32 | 423 |
|  | LAESV | 20-52 | 53-58 | > 58 | 453 | 20-56 | 57-64 | > 64 | 374 |
|  | LAEF | 57-83 | 53-56 | < 53 | 453 | 55-81 | 51-54 | < 51 | 374 |
|  | RAESV | 24-57 | 58-63 | > 63 | 452 | 26-69 | 70-77 | > 77 | 362 |
|  | RAEF | 41-75 | 36-40 | < 36 | 445 | 36-73 | 31-35 | < 31 | 360 |
| 50-59 | LVEDVi | 62-113 | 114-123 | > 123 | 912 | 69-128 | 129-138 | > 138 | 805 |
|  | LVESVi | 20-46 | 47-52 | > 52 | 911 | 24-55 | 56-60 | > 60 | 801 |
|  | LVSVi | 37-72 | 33-36 | < 33 | 912 | 40-80 | 34-39 | < 34 | 820 |
|  | LVEF | 53-72 | 49-52 | < 49 | 912 | 50-71 | 47-49 | < 47 | 833 |
|  | LVMi | 34-69 | 70-78 | > 78 | 912 | 47-90 | 91-100 | > 100 | 825 |
|  | LVM/LVEDV | 0.4-0.8 | 0.9-1 | > 1 | 912 | 0.5-1 | 1.1-1.1 | > 1.1 | 795 |
|  | RVEDVi | 58-116 | 117-128 | > 128 | 910 | 72-137 | 138-148 | > 148 | 811 |
|  | RVESVi | 20-49 | 50-55 | > 55 | 910 | 26-62 | 63-68 | > 68 | 804 |
|  | RVEF | 50-72 | 46-49 | < 46 | 909 | 46-70 | 42-45 | < 42 | 823 |
|  | RVSVi | 34-74 | 30-33 | < 30 | 906 | 37-82 | 30-36 | < 30 | 818 |
|  | LAESV | 20-56 | 57-63 | > 63 | 825 | 19-57 | 58-66 | > 66 | 700 |
|  | LAEF | 56-83 | 51-55 | < 51 | 823 | 54-82 | 50-53 | < 50 | 703 |
|  | RAESV | 24-59 | 60-67 | > 67 | 769 | 25-70 | 71-79 | > 79 | 604 |
|  | RAEF | 40-75 | 35-39 | < 35 | 760 | 36-74 | 31-35 | < 31 | 604 |
| 60-69 | LVEDVi | 58-108 | 109-120 | > 120 | 920 | 65-126 | 127-136 | > 136 | 789 |
|  | LVESVi | 19-44 | 45-51 | > 51 | 918 | 22-54 | 55-60 | > 60 | 785 |
|  | LVSVi | 36-69 | 32-35 | < 32 | 920 | 38-78 | 31-37 | < 31 | 792 |
|  | LVEF | 53-73 | 49-52 | < 49 | 917 | 50-71 | 47-49 | < 47 | 796 |
|  | LVMi | 34-70 | 71-80 | > 80 | 920 | 45-89 | 90-99 | > 99 | 798 |
|  | LVM/LVEDV | 0.5-0.9 | 01-1 | > 1 | 920 | 0.5-1 | 1.1-1.2 | > 1.2 | 781 |
|  | RVEDVi | 57-114 | 115-127 | > 127 | 918 | 68-133 | 134-145 | > 145 | 791 |
|  | RVESVi | 18-47 | 48-54 | > 54 | 919 | 24-61 | 62-68 | > 68 | 788 |
|  | RVEF | 51-74 | 46-50 | < 46 | 919 | 46-71 | 42-45 | < 42 | 794 |
|  | RVSVi | 33-73 | 28-32 | < 28 | 916 | 35-81 | 28-34 | < 28 | 793 |
|  | LAESV | 19-57 | 58-66 | > 66 | 809 | 18-58 | 59-68 | > 68 | 652 |
|  | LAEF | 54-83 | 48-53 | < 48 | 807 | 53-83 | 48-52 | < 48 | 654 |
|  | RAESV | 23-61 | 62-70 | > 70 | 721 | 25-71 | 72-80 | > 80 | 521 |
|  | RAEF | 41-75 | 35-40 | < 35 | 699 | 37-74 | 32-36 | < 32 | 496 |
| 70+ | LVEDVi | 55-105 | 106-118 | > 118 | 1099 | 60-123 | 124-135 | > 135 | 1242 |
|  | LVESVi | 17-43 | 44-50 | > 50 | 1098 | 21-53 | 54-60 | > 60 | 1227 |
|  | LVSVi | 34-68 | 31-33 | < 31 | 1101 | 35-75 | 28-34 | < 28 | 1253 |
|  | LVEF | 53-73 | 49-52 | < 49 | 1098 | 50-71 | 46-49 | < 46 | 1241 |
|  | LVMi | 34-69 | 70-78 | > 78 | 1101 | 43-86 | 87-96 | > 96 | 1248 |
|  | LVM/LVEDV | 0.5-0.9 | 1-1.1 | > 1.1 | 1099 | 0.5-1 | 1.1-1.2 | > 1.2 | 1235 |
|  | RVEDVi | 54-111 | 112-125 | > 125 | 1100 | 65-130 | 131-141 | > 141 | 1242 |
|  | RVESVi | 17-45 | 46-52 | > 52 | 1100 | 23-60 | 61-67 | > 67 | 1239 |
|  | RVEF | 50-75 | 44-49 | < 44 | 1096 | 45-72 | 41-44 | < 41 | 1228 |
|  | RVSVi | 31-72 | 27-30 | < 27 | 1097 | 33-80 | 25-32 | < 25 | 1244 |
|  | LAESV | 18-59 | 60-70 | > 70 | 947 | 17-60 | 61-70 | > 70 | 953 |
|  | LAEF | 50-83 | 44-49 | < 44 | 938 | 51-84 | 45-50 | < 45 | 943 |
|  | RAESV | 23-63 | 64-73 | > 73 | 769 | 25-72 | 73-82 | > 82 | 631 |
|  | RAEF | 42-77 | 37-41 | < 37 | 753 | 37-75 | 32-36 | < 32 | 619 |

Footnote: All measurements were derived using suiteHEART software with smooth endocardial segmentation; papillary muscles and trabeculations were excluded from left ventricular mass measurements. Volumetric and mass measurements were indexed to height. Reference ranges are defined as the interval between the 2.5th and 97.5th centiles derived from age-specific generalized additive models for location, scale and shape (GAMLSS) using the Box–Cox Cole–Green distribution. Centile-based categories beyond the reference range are derived from LMS-based thresholds (P0.5, P2.5, P97.5, and P99.5) and describe the position of a measurement within the reference distribution rather than disease severity or diagnostic abnormality. Reference ranges are based on a White adult population.

Abbreviations: LV = left ventricle; RV = right ventricle; LA = left atrium; RA = right atrium; EDV = end-diastolic volume; ESV = end-systolic volume; SV = stroke volume; EF = ejection fraction; CO = cardiac output.

### 17. Table: Reference ranges and centile-based categories for cardiac chamber measurements derived using suiteHEART software (smooth segmentation), indexed to height, in a Black adult population

|  |  | **Female** | | | | **Male** | | | |
| --- | --- | --- | --- | --- | --- | --- | --- | --- | --- |
| **Age group** | **Metrics** | **Reference range** | **Mild** | **At least moderate** | **n** | **Reference range** | **Mild** | **At least moderate** | **n** |
| 40-49 | LVEDVi | 67-119 | 120-132 | > 132 | 10 |  |  |  |  |
|  | LVESVi | 23-51 | 52-57 | > 57 | 10 |  |  |  |  |
|  | LVSVi | 42-81 | 39-41 | < 39 | 10 |  |  |  |  |
|  | LVEF | 52-72 | 50-51 | < 50 | 10 |  |  |  |  |
|  | LVMi | 36-77 | 78-91 | > 91 | 10 |  |  |  |  |
|  | LVM/LVEDV | 0.4-0.8 | 0.9-1 | > 1 | 10 |  |  |  |  |
|  | RVEDVi | 59-123 | 124-137 | > 137 | 10 |  |  |  |  |
|  | RVESVi | 22-55 | 56-64 | > 64 | 10 |  |  |  |  |
|  | RVEF | 47-69 | 43-46 | < 43 | 10 |  |  |  |  |
|  | RVSVi | 33-75 | 28-32 | < 28 | 10 |  |  |  |  |
| 50-59 | LVEDVi | 64-116 | 117-128 | > 128 | 95 | 68-132 | 133-145 | > 145 | 74 |
|  | LVESVi | 21-49 | 50-55 | > 55 | 95 | 24-59 | 60-68 | > 68 | 73 |
|  | LVSVi | 39-74 | 36-38 | < 36 | 95 | 41-78 | 35-40 | < 35 | 75 |
|  | LVEF | 52-73 | 50-51 | < 50 | 95 | 50-70 | 47-49 | < 47 | 76 |
|  | LVMi | 37-77 | 78-91 | > 91 | 95 | 46-95 | 96-108 | > 108 | 77 |
|  | LVM/LVEDV | 0.4-0.9 | 01-1 | > 1 | 95 | 0.5-0.9 | 01-1 | > 1 | 74 |
|  | RVEDVi | 59-120 | 121-133 | > 133 | 95 | 75-137 | 138-154 | > 154 | 77 |
|  | RVESVi | 21-54 | 55-63 | > 63 | 95 | 28-59 | 60-67 | > 67 | 74 |
|  | RVEF | 48-71 | 43-47 | < 43 | 95 | 47-69 | 43-46 | < 43 | 75 |
|  | RVSVi | 34-74 | 29-33 | < 29 | 95 | 41-81 | 36-40 | < 36 | 76 |
|  | LAESVi | 22-54 | 55-60 | > 60 | 88 | 21-57 | 58-68 | > 68 | 67 |
|  | LAEF | 59-79 | 56-58 | < 56 | 88 | 56-83 | 53-55 | < 53 | 67 |
|  | RAESVi | 23-64 | 65-73 | > 73 | 84 | 28-66 | 67-72 | > 72 | 64 |
|  | RAEF | 38-69 | 32-37 | < 32 | 84 | 38-65 | 35-37 | < 35 | 63 |
| 60-69 | LVEDVi | 58-111 | 112-121 | > 121 | 65 | 65-130 | 131-146 | > 146 | 48 |
|  | LVESVi | 18-46 | 47-53 | > 53 | 65 | 21-58 | 59-67 | > 67 | 47 |
|  | LVSVi | 35-68 | 30-34 | < 30 | 65 | 41-78 | 38-40 | < 38 | 48 |
|  | LVEF | 53-73 | 49-52 | < 49 | 65 | 49-71 | 44-48 | < 44 | 48 |
|  | LVMi | 39-78 | 79-91 | > 91 | 65 | 46-91 | 92-105 | > 105 | 48 |
|  | LVM/LVEDV | 0.5-0.9 | 1-1.1 | > 1.1 | 65 | 0.5-1 | 1.1-1.1 | > 1.1 | 48 |
|  | RVEDVi | 60-117 | 118-129 | > 129 | 65 | 70-130 | 131-144 | > 144 | 48 |
|  | RVESVi | 18-52 | 53-60 | > 60 | 65 | 23-59 | 60-68 | > 68 | 47 |
|  | RVEF | 48-74 | 43-47 | < 43 | 64 | 46-73 | 42-45 | < 42 | 48 |
|  | RVSVi | 35-73 | 30-34 | < 30 | 65 | 38-78 | 33-37 | < 33 | 48 |
|  | LAESVi | 20-56 | 57-64 | > 64 | 60 | 19-59 | 60-70 | > 70 | 42 |
|  | LAEF | 54-81 | 49-53 | < 49 | 61 | 52-85 | 48-51 | < 48 | 42 |
|  | RAESVi | 23-61 | 62-69 | > 69 | 52 | 24-72 | 73-79 | > 79 | 40 |
|  | RAEF | 32-66 | 21-31 | < 21 | 53 | 36-71 | 32-35 | < 32 | 41 |
| 70+ | LVEDVi | 53-106 | 107-115 | > 115 | 18 | 61-129 | 130-150 | > 150 | 14 |
|  | LVESVi | 16-45 | 46-52 | > 52 | 18 | 18-57 | 58-67 | > 67 | 14 |
|  | LVSVi | 35-67 | 30-34 | < 30 | 18 | 40-82 | 38-39 | < 38 | 14 |
|  | LVEF | 52-73 | 45-51 | < 45 | 18 | 46-72 | 38-45 | < 38 | 14 |
|  | LVMi | 41-80 | 81-91 | > 91 | 18 | 46-88 | 89-102 | > 102 | 14 |
|  | LVM/LVEDV | 0.5-1 | 1.1-1.1 | > 1.1 | 18 | 0.5-1.1 | 1.2-1.3 | > 1.3 | 14 |
|  | RVEDVi | 60-117 | 118-131 | > 131 | 18 | 66-123 | 124-135 | > 135 | 14 |
|  | RVESVi | 16-51 | 52-58 | > 58 | 18 | 18-59 | 60-68 | > 68 | 14 |
|  | RVEF | 48-76 | 43-47 | < 43 | 18 | 45-79 | 42-44 | < 42 | 14 |
|  | RVSVi | 34-75 | 29-33 | < 29 | 18 | 37-74 | 32-36 | < 32 | 14 |
|  | LAESVi | 18-59 | 60-68 | > 68 | 16 |  |  |  |  |
|  | LAEF | 47-82 | 39-46 | < 39 | 17 |  |  |  |  |
|  | RAESVi | 24-58 | 59-65 | > 65 | 13 |  |  |  |  |
|  | RAEF | 35-71 | 25-34 | < 25 | 12 |  |  |  |  |

Footnote: All measurements were derived using suiteHEAR software with smooth endocardial segmentation; papillary muscles and trabeculations were excluded from left ventricular mass measurements. Volumetric and mass measurements were indexed to height. Reference ranges are defined as the interval between the 2.5th and 97.5th centiles derived from age-specific generalized additive models for location, scale and shape (GAMLSS) using the Box–Cox Cole–Green distribution. Centile-based categories beyond the reference range are derived from LMS-based thresholds (P0.5, P2.5, P97.5, and P99.5) and describe the position of a measurement within the reference distribution rather than disease severity or diagnostic abnormality. Reference ranges are based on a Black adult population.

Abbreviations: LV = left ventricle; RV = right ventricle; LA = left atrium; RA = right atrium; EDV = end-diastolic volume; ESV = end-systolic volume; SV = stroke volume; EF = ejection fraction; CO = cardiac output.

### 18. Table: Reference ranges and centile-based categories for cardiac chamber measurements derived using suiteHEART software (smooth segmentation), indexed to height, in an Asian adult population

|  |  | **Female** | | | | **Male** | | | |
| --- | --- | --- | --- | --- | --- | --- | --- | --- | --- |
| **Age group** | **Metrics** | **Reference range** | **Mild** | **At least moderate** | **n** | **Reference range** | **Mild** | **At least moderate** | **n** |
| 40-49 | LVEDVi | 54-95 | 96-102 | > 102 | 10 | 65-119 | 120-131 | > 131 | 20 |
|  | LVESVi | 19-40 | 41-48 | > 48 | 10 | 21-49 | 50-55 | > 55 | 20 |
|  | LVSVi | 33-60 | 28-32 | < 28 | 10 | 42-74 | 39-41 | < 39 | 20 |
|  | LVEF | 54-72 | 50-53 | < 50 | 10 | 53-71 | 50-52 | < 50 | 20 |
|  | LVMi | 29-60 | 61-69 | > 69 | 10 | 40-70 | 71-75 | > 75 | 20 |
|  | LVM/LVEDV | 0.4-0.8 | 0.9-1 | > 1 | 10 | 0.5-0.8 | 0.9-0.9 | > 0.9 | 20 |
|  | RVEDVi | 53-95 | 96-104 | > 104 | 10 | 65-118 | 119-126 | > 126 | 20 |
|  | RVESVi | 16-40 | 41-45 | > 45 | 10 | 22-56 | 57-61 | > 61 | 20 |
|  | RVEF | 49-76 | 45-48 | < 45 | 10 | 45-69 | 40-44 | < 40 | 20 |
|  | RVSVi | 30-60 | 27-29 | < 27 | 10 | 36-72 | 32-35 | < 32 | 20 |
|  | LAESVi | 21-46 | 47-51 | > 51 | 10 | 16-50 | 51-57 | > 57 | 19 |
|  | LAEF | 59-85 | 55-58 | < 55 | 10 | 61-81 | 57-60 | < 57 | 19 |
|  | RAESVi | 22-48 | 49-54 | > 54 | 10 | 20-58 | 59-63 | > 63 | 17 |
|  | RAEF | 39-69 | 31-38 | < 31 | 10 | 35-71 | 30-34 | < 30 | 17 |
| 50-59 | LVEDVi | 53-95 | 96-103 | > 103 | 83 | 60-114 | 115-124 | > 124 | 132 |
|  | LVESVi | 17-42 | 43-51 | > 51 | 83 | 20-49 | 50-55 | > 55 | 132 |
|  | LVSVi | 33-60 | 29-32 | < 29 | 83 | 36-70 | 31-35 | < 31 | 132 |
|  | LVEF | 53-73 | 48-52 | < 48 | 83 | 52-71 | 49-51 | < 49 | 131 |
|  | LVMi | 29-63 | 64-72 | > 72 | 83 | 37-75 | 76-83 | > 83 | 131 |
|  | LVM/LVEDV | 0.4-0.8 | 0.9-1 | > 1 | 83 | 0.5-0.9 | 01-1 | > 1 | 131 |
|  | RVEDVi | 52-97 | 98-107 | > 107 | 83 | 61-117 | 118-127 | > 127 | 131 |
|  | RVESVi | 16-39 | 40-44 | > 44 | 83 | 21-54 | 55-60 | > 60 | 131 |
|  | RVEF | 50-76 | 46-49 | < 46 | 82 | 46-70 | 41-45 | < 41 | 131 |
|  | RVSVi | 32-64 | 29-31 | < 29 | 82 | 34-72 | 29-33 | < 29 | 131 |
|  | LAESVi | 20-47 | 48-53 | > 53 | 77 | 17-51 | 52-58 | > 58 | 121 |
|  | LAEF | 58-84 | 53-57 | < 53 | 77 | 58-81 | 54-57 | < 54 | 122 |
|  | RAESVi | 22-51 | 52-59 | > 59 | 68 | 21-59 | 60-66 | > 66 | 118 |
|  | RAEF | 38-69 | 30-37 | < 30 | 68 | 38-71 | 33-37 | < 33 | 115 |
| 60-69 | LVEDVi | 52-95 | 96-105 | > 105 | 63 | 55-109 | 110-119 | > 119 | 118 |
|  | LVESVi | 16-42 | 43-51 | > 51 | 63 | 19-47 | 48-54 | > 54 | 118 |
|  | LVSVi | 33-60 | 30-32 | < 30 | 63 | 33-69 | 28-32 | < 28 | 118 |
|  | LVEF | 53-74 | 48-52 | < 48 | 63 | 51-72 | 47-50 | < 47 | 119 |
|  | LVMi | 30-67 | 68-79 | > 79 | 63 | 37-80 | 81-93 | > 93 | 119 |
|  | LVM/LVEDV | 0.5-0.9 | 01-1 | > 1 | 63 | 0.5-1 | 1.1-1.1 | > 1.1 | 118 |
|  | RVEDVi | 50-101 | 102-113 | > 113 | 63 | 56-116 | 117-127 | > 127 | 119 |
|  | RVESVi | 16-39 | 40-44 | > 44 | 63 | 19-52 | 53-58 | > 58 | 118 |
|  | RVEF | 52-75 | 48-51 | < 48 | 63 | 47-72 | 42-46 | < 42 | 119 |
|  | RVSVi | 32-71 | 29-31 | < 29 | 63 | 31-71 | 26-30 | < 26 | 119 |
|  | LAESVi | 19-50 | 51-57 | > 57 | 60 | 17-52 | 53-60 | > 60 | 111 |
|  | LAEF | 55-82 | 51-54 | < 51 | 60 | 55-82 | 51-54 | < 51 | 111 |
|  | RAESVi | 22-57 | 58-67 | > 67 | 51 | 21-62 | 63-71 | > 71 | 86 |
|  | RAEF | 37-72 | 29-36 | < 29 | 50 | 40-71 | 37-39 | < 37 | 86 |
| 70+ | LVEDVi | 51-97 | 98-110 | > 110 | 28 | 53-108 | 109-121 | > 121 | 40 |
|  | LVESVi | 15-36 | 37-42 | > 42 | 28 | 18-46 | 47-55 | > 55 | 40 |
|  | LVSVi | 33-62 | 30-32 | < 30 | 28 | 34-70 | 30-33 | < 30 | 40 |
|  | LVEF | 57-76 | 55-56 | < 55 | 28 | 50-72 | 44-49 | < 44 | 40 |
|  | LVMi | 30-71 | 72-88 | > 88 | 28 | 39-77 | 78-91 | > 91 | 40 |
|  | LVM/LVEDV | 0.5-0.9 | 1-1.1 | > 1.1 | 28 | 0.5-1.1 | 1.2-1.4 | > 1.4 | 40 |
|  | RVEDVi | 49-105 | 106-118 | > 118 | 27 | 52-115 | 116-127 | > 127 | 40 |
|  | RVESVi | 16-39 | 40-44 | > 44 | 27 | 18-49 | 50-56 | > 56 | 40 |
|  | RVEF | 53-75 | 50-52 | < 50 | 27 | 48-73 | 43-47 | < 43 | 39 |
|  | RVSVi | 29-68 | 25-28 | < 25 | 27 | 28-71 | 22-27 | < 22 | 40 |
|  | LAESVi | 18-53 | 54-60 | > 60 | 29 | 18-54 | 55-61 | > 61 | 34 |
|  | LAEF | 52-81 | 48-51 | < 48 | 28 | 52-83 | 47-51 | < 47 | 34 |
|  | RAESVi | 21-64 | 65-77 | > 77 | 21 | 21-66 | 67-77 | > 77 | 26 |
|  | RAEF | 39-79 | 30-38 | < 30 | 19 | 43-71 | 40-42 | < 40 | 25 |

Footnote: All measurements were derived using suiteHEART software with smooth endocardial segmentation; papillary muscles and trabeculations were excluded from left ventricular mass measurements. Volumetric and mass measurements were indexed to height. Reference ranges are defined as the interval between the 2.5th and 97.5th centiles derived from age-specific generalized additive models for location, scale and shape (GAMLSS) using the Box–Cox Cole–Green distribution. Centile-based categories beyond the reference range are derived from LMS-based thresholds (P0.5, P2.5, P97.5, and P99.5) and describe the position of a measurement within the reference distribution rather than disease severity or diagnostic abnormality. Reference ranges are based on an Asian adult population.

Abbreviations: LV = left ventricle; RV = right ventricle; LA = left atrium; RA = right atrium; EDV = end-diastolic volume; ESV = end-systolic volume; SV = stroke volume; EF = ejection fraction; CO = cardiac output.

### 19. Table: Reference ranges and centile-based categories for cardiac chamber measurements derived using suiteHEART software (smooth segmentation), indexed to height, in a Chinese adult population

|  |  | **Female** | | | | | **Male** | | | |
| --- | --- | --- | --- | --- | --- | --- | --- | --- | --- | --- |
| **Age group** | **Metrics** | **Reference range** | **Mild** | **At least moderate** | **n** | **Reference range** | | **Mild** | **At least moderate** | **n** |
| 18-29 | LVEDVi | 58-111 | 112-128 | > 128 | 16 | 76-115 | | 116-121 | > 121 | 19 |
|  | LVESVi | 22-48 | 49-66 | > 66 | 16 | 26-46 | | 47-49 | > 49 | 18 |
|  | LVSVi | 34-73 | 30-33 | < 30 | 16 | 47-71 | | 44-46 | < 44 | 19 |
|  | LVEF | 54-70 | 52-53 | < 52 | 16 | 51-66 | | 46-50 | < 46 | 19 |
|  | LVMi | 26-54 | 55-69 | > 69 | 16 | 39-70 | | 71-74 | > 74 | 19 |
|  | LVM/LVEDV | 0.4-0.6 |  | > 0.6 | 16 | 0.5-0.7 | | 0.8-0.8 | > 0.8 | 19 |
|  | RVEDVi | 61-110 | 111-121 | > 121 | 16 | 83-126 | | 127-131 | > 131 | 19 |
|  | RVESVi | 24-52 | 53-57 | > 57 | 16 | 33-65 | | 66-72 | > 72 | 19 |
|  | RVEF | 45-67 | 41-44 | < 41 | 16 | 48-64 | | 45-47 | < 45 | 19 |
|  | RVSVi | 32-65 | 27-31 | < 27 | 16 | 48-73 | | 45-47 | < 45 | 19 |
|  | LAESVi | 20-50 | 51-58 | > 58 | 12 | 22-47 | | 48-52 | > 52 | 15 |
|  | LAEF | 55-79 | 49-54 | < 49 | 12 | 53-77 | | 50-52 | < 50 | 15 |
|  | RAESVi | 23-47 | 48-54 | > 54 | 11 | 25-68 | | 69-82 | > 82 | 15 |
|  | RAEF | 47-69 | 45-46 | < 45 | 11 | 39-64 | | 33-38 | < 33 | 14 |
| 30-39 | LVEDVi | 57-106 | 107-120 | > 120 | 18 | 73-113 | | 114-120 | > 120 | 17 |
|  | LVESVi | 20-45 | 46-58 | > 58 | 18 | 26-46 | | 47-50 | > 50 | 17 |
|  | LVSVi | 34-69 | 31-33 | < 31 | 18 | 44-70 | | 41-43 | < 41 | 17 |
|  | LVEF | 55-71 | 52-54 | < 52 | 18 | 49-67 | | 42-48 | < 42 | 17 |
|  | LVMi | 27-56 | 57-71 | > 71 | 18 | 39-71 | | 72-76 | > 76 | 17 |
|  | LVM/LVEDV | 0.4-0.6 | 0.7-0.7 | > 0.7 | 18 | 0.5-0.8 | |  | > 0.8 | 17 |
|  | RVEDVi | 59-108 | 109-118 | > 118 | 18 | 79-123 | | 124-129 | > 129 | 17 |
|  | RVESVi | 21-49 | 50-55 | > 55 | 18 | 30-62 | | 63-69 | > 69 | 17 |
|  | RVEF | 46-69 | 42-45 | < 42 | 18 | 47-65 | | 44-46 | < 44 | 17 |
|  | RVSVi | 31-64 | 26-30 | < 26 | 18 | 45-72 | | 42-44 | < 42 | 17 |
|  | LAESVi | 20-49 | 50-55 | > 55 | 16 | 21-47 | | 48-51 | > 51 | 17 |
|  | LAEF | 55-79 | 49-54 | < 49 | 16 | 53-78 | | 50-52 | < 50 | 17 |
|  | RAESVi | 23-49 | 50-57 | > 57 | 16 | 25-66 | | 67-78 | > 78 | 17 |
|  | RAEF | 48-71 | 45-47 | < 45 | 14 | 39-66 | | 33-38 | < 33 | 17 |
| 40-49 | LVEDVi | 56-101 | 102-113 | > 113 | 24 | 70-112 | | 113-120 | > 120 | 20 |
|  | LVESVi | 19-42 | 43-52 | > 52 | 24 | 25-47 | | 48-51 | > 51 | 20 |
|  | LVSVi | 35-63 | 32-34 | < 32 | 24 | 41-70 | | 37-40 | < 37 | 20 |
|  | LVEF | 55-72 | 52-54 | < 52 | 24 | 52-70 | | 50-51 | < 50 | 20 |
|  | LVMi | 28-59 | 60-74 | > 74 | 24 | 40-72 | | 73-79 | > 79 | 20 |
|  | LVM/LVEDV | 0.4-0.7 |  | > 0.7 | 24 | 0.5-0.8 | | 0.9-0.9 | > 0.9 | 20 |
|  | RVEDVi | 56-105 | 106-115 | > 115 | 24 | 74-120 | | 121-127 | > 127 | 20 |
|  | RVESVi | 19-46 | 47-52 | > 52 | 24 | 27-59 | | 60-65 | > 65 | 20 |
|  | RVEF | 47-71 | 42-46 | < 42 | 24 | 47-67 | | 44-46 | < 44 | 20 |
|  | RVSVi | 30-63 | 26-29 | < 26 | 24 | 42-72 | | 38-41 | < 38 | 20 |
|  | LAESVi | 20-47 | 48-52 | > 52 | 20 | 20-47 | | 48-51 | > 51 | 19 |
|  | LAEF | 55-80 | 50-54 | < 50 | 20 | 53-80 | | 49-52 | < 49 | 19 |
|  | RAESVi | 23-52 | 53-60 | > 60 | 21 | 24-65 | | 66-74 | > 74 | 20 |
|  | RAEF | 48-73 | 45-47 | < 45 | 20 | 39-68 | | 33-38 | < 33 | 20 |
| 50-59 | LVEDVi | 54-96 | 97-106 | > 106 | 66 | 66-111 | | 112-121 | > 121 | 49 |
|  | LVESVi | 18-40 | 41-47 | > 47 | 66 | 24-46 | | 47-51 | > 51 | 49 |
|  | LVSVi | 35-59 | 32-34 | < 32 | 66 | 38-70 | | 34-37 | < 34 | 49 |
|  | LVEF | 55-73 | 52-54 | < 52 | 66 | 52-71 | | 49-51 | < 49 | 49 |
|  | LVMi | 30-61 | 62-76 | > 76 | 66 | 40-75 | | 76-84 | > 84 | 49 |
|  | LVM/LVEDV | 0.4-0.8 | 0.9-0.9 | > 0.9 | 66 | 0.5-0.9 | | 01-1 | > 1 | 49 |
|  | RVEDVi | 53-101 | 102-111 | > 111 | 66 | 70-119 | | 120-128 | > 128 | 49 |
|  | RVESVi | 17-44 | 45-50 | > 50 | 66 | 25-55 | | 56-61 | > 61 | 49 |
|  | RVEF | 48-72 | 43-47 | < 43 | 66 | 46-70 | | 43-45 | < 43 | 49 |
|  | RVSVi | 30-63 | 25-29 | < 25 | 66 | 38-73 | | 34-37 | < 34 | 49 |
|  | LAESVi | 20-46 | 47-52 | > 52 | 59 | 19-46 | | 47-50 | > 50 | 45 |
|  | LAEF | 55-80 | 50-54 | < 50 | 58 | 53-81 | | 48-52 | < 48 | 45 |
|  | RAESVi | 24-54 | 55-64 | > 64 | 61 | 23-63 | | 64-71 | > 71 | 44 |
|  | RAEF | 47-74 | 44-46 | < 44 | 60 | 39-70 | | 33-38 | < 33 | 44 |
| 60-69 | LVEDVi | 53-92 | 93-101 | > 101 | 52 | 63-110 | | 111-123 | > 123 | 37 |
|  | LVESVi | 16-37 | 38-43 | > 43 | 52 | 23-45 | | 46-51 | > 51 | 37 |
|  | LVSVi | 34-59 | 31-33 | < 31 | 52 | 34-71 | | 31-33 | < 31 | 37 |
|  | LVEF | 55-74 | 51-54 | < 51 | 52 | 50-71 | | 46-49 | < 46 | 37 |
|  | LVMi | 31-63 | 64-78 | > 78 | 52 | 40-78 | | 79-91 | > 91 | 37 |
|  | LVM/LVEDV | 0.5-0.9 | 01-1 | > 1 | 52 | 0.5-0.9 | | 01-1 | > 1 | 37 |
|  | RVEDVi | 50-96 | 97-105 | > 105 | 52 | 66-120 | | 121-135 | > 135 | 37 |
|  | RVESVi | 15-41 | 42-47 | > 47 | 52 | 22-51 | | 52-56 | > 56 | 37 |
|  | RVEF | 49-74 | 43-48 | < 43 | 51 | 46-73 | | 43-45 | < 43 | 37 |
|  | RVSVi | 29-62 | 24-28 | < 24 | 52 | 35-75 | | 31-34 | < 31 | 37 |
|  | LAESVi | 18-49 | 50-57 | > 57 | 48 | 17-44 | | 45-49 | > 49 | 31 |
|  | LAEF | 54-81 | 49-53 | < 49 | 48 | 52-83 | | 47-51 | < 47 | 31 |
|  | RAESVi | 24-57 | 58-69 | > 69 | 45 | 23-63 | | 64-70 | > 70 | 32 |
|  | RAEF | 46-74 | 42-45 | < 42 | 45 | 39-72 | | 33-38 | < 33 | 31 |
| 70+ | LVEDVi | 52-88 | 89-96 | > 96 | 12 |  | |  |  |  |
|  | LVESVi | 15-35 | 36-40 | > 40 | 12 |  | |  |  |  |
|  | LVSVi | 30-62 | 26-29 | < 26 | 12 |  | |  |  |  |
|  | LVEF | 54-75 | 50-53 | < 50 | 12 |  | |  |  |  |
|  | LVMi | 33-65 | 66-80 | > 80 | 12 |  | |  |  |  |
|  | LVM/LVEDV | 0.5-1 | 1.1-1.2 | > 1.2 | 12 |  | |  |  |  |
|  | RVEDVi | 46-91 | 92-99 | > 99 | 12 |  | |  |  |  |
|  | RVESVi | 14-38 | 39-45 | > 45 | 12 |  | |  |  |  |
|  | RVEF | 50-76 | 43-49 | < 43 | 12 |  | |  |  |  |
|  | RVSVi | 28-62 | 24-27 | < 24 | 12 |  | |  |  |  |
|  | LAESVi | 17-57 | 58-71 | > 71 | 12 |  | |  |  |  |
|  | LAEF | 54-82 | 48-53 | < 48 | 12 |  | |  |  |  |
|  | RAESVi | 24-60 | 61-74 | > 74 | 10 |  | |  |  |  |
|  | RAEF | 44-73 | 41-43 | < 41 | 10 |  | |  |  |  |

Footnote: All measurements were derived using suiteHEART software with smooth endocardial segmentation; papillary muscles and trabeculations were excluded from left ventricular mass measurements. Volumetric and mass measurements were indexed to height. Reference ranges are defined as the interval between the 2.5th and 97.5th centiles derived from age-specific generalized additive models for location, scale and shape (GAMLSS) using the Box–Cox Cole–Green distribution. Centile-based categories beyond the reference range are derived from LMS-based thresholds (P0.5, P2.5, P97.5, and P99.5) and describe the position of a measurement within the reference distribution rather than disease severity or diagnostic abnormality. Reference ranges are based on a Chinese adult population.

Abbreviations: LV = left ventricle; RV = right ventricle; LA = left atrium; RA = right atrium; EDV = end-diastolic volume; ESV = end-systolic volume; SV = stroke volume; EF = ejection fraction; CO = cardiac output.

### 20. Table: Reference ranges and centile-based categories for cardiac chamber measurements derived using suiteHEART software (smooth segmentation), indexed to height, in a Mixed adult population

|  |  | **Female** | | | | | **Male** | | | |
| --- | --- | --- | --- | --- | --- | --- | --- | --- | --- | --- |
| **Age group** | **Metrics** | **Reference range** | **Mild** | **At least moderate** | **n** | **Reference range** | | **Mild** | **At least moderate** | **n** |
| 50-59 | LVEDVi | 60-107 | 108-117 | > 117 | 134 | 69-123 | | 124-130 | > 130 | 65 |
|  | LVESVi | 19-42 | 43-46 | > 46 | 134 | 25-52 | | 53-57 | > 57 | 66 |
|  | LVSVi | 38-70 | 35-37 | < 35 | 133 | 40-81 | | 33-39 | < 33 | 66 |
|  | LVEF | 54-74 | 52-53 | < 52 | 134 | 52-71 | | 48-51 | < 48 | 66 |
|  | LVMi | 31-70 | 71-82 | > 82 | 134 | 41-92 | | 93-104 | > 104 | 64 |
|  | LVM/LVEDV | 0.4-0.8 | 0.9-0.9 | > 0.9 | 134 | 0.5-1 | | 1.1-1.3 | > 1.3 | 63 |
|  | RVEDVi | 55-108 | 109-116 | > 116 | 134 | 71-133 | | 134-142 | > 142 | 65 |
|  | RVESVi | 19-45 | 46-50 | > 50 | 134 | 27-59 | | 60-66 | > 66 | 65 |
|  | RVEF | 52-73 | 49-51 | < 49 | 134 | 46-71 | | 40-45 | < 40 | 66 |
|  | RVSVi | 33-70 | 27-32 | < 27 | 134 | 38-81 | | 30-37 | < 30 | 66 |
|  | LAESVi | 20-51 | 52-57 | > 57 | 122 | 21-57 | | 58-64 | > 64 | 53 |
|  | LAEF | 57-80 | 51-56 | < 51 | 122 | 56-81 | | 51-55 | < 51 | 54 |
|  | RAESVi | 23-60 | 61-69 | > 69 | 117 | 27-68 | | 69-76 | > 76 | 54 |
|  | RAEF | 41-72 | 37-40 | < 37 | 117 | 37-72 | | 32-36 | < 32 | 54 |
| 60-69 | LVEDVi | 56-105 | 106-116 | > 116 | 110 | 62-120 | | 121-130 | > 130 | 59 |
|  | LVESVi | 17-42 | 43-48 | > 48 | 110 | 22-51 | | 52-58 | > 58 | 58 |
|  | LVSVi | 35-68 | 32-34 | < 32 | 110 | 37-75 | | 30-36 | < 30 | 59 |
|  | LVEF | 54-74 | 51-53 | < 51 | 110 | 50-71 | | 45-49 | < 45 | 59 |
|  | LVMi | 32-67 | 68-75 | > 75 | 110 | 42-88 | | 89-98 | > 98 | 59 |
|  | LVM/LVEDV | 0.4-0.8 | 0.9-1 | > 1 | 110 | 0.5-1.1 | | 1.2-1.3 | > 1.3 | 59 |
|  | RVEDVi | 52-110 | 111-121 | > 121 | 110 | 65-132 | | 133-145 | > 145 | 57 |
|  | RVESVi | 18-44 | 45-49 | > 49 | 110 | 25-63 | | 64-73 | > 73 | 58 |
|  | RVEF | 52-74 | 47-51 | < 47 | 110 | 47-71 | | 43-46 | < 43 | 58 |
|  | RVSVi | 30-72 | 24-29 | < 24 | 110 | 34-79 | | 27-33 | < 27 | 58 |
|  | LAESVi | 19-53 | 54-59 | > 59 | 100 | 19-56 | | 57-65 | > 65 | 53 |
|  | LAEF | 55-81 | 50-54 | < 50 | 100 | 52-81 | | 48-51 | < 48 | 53 |
|  | RAESVi | 26-59 | 60-68 | > 68 | 93 | 25-69 | | 70-77 | > 77 | 40 |
|  | RAEF | 40-73 | 35-39 | < 35 | 93 | 37-72 | | 32-36 | < 32 | 40 |
| 70+ | LVEDVi | 53-103 | 104-114 | > 114 | 51 | 57-121 | | 122-134 | > 134 | 33 |
|  | LVESVi | 16-43 | 44-50 | > 50 | 51 | 20-53 | | 54-62 | > 62 | 33 |
|  | LVSVi | 32-66 | 28-31 | < 28 | 51 | 34-70 | | 28-33 | < 28 | 34 |
|  | LVEF | 53-75 | 49-52 | < 49 | 51 | 48-70 | | 40-47 | < 40 | 35 |
|  | LVMi | 32-66 | 67-72 | > 72 | 51 | 43-85 | | 86-94 | > 94 | 35 |
|  | LVM/LVEDV | 0.5-0.9 | 01-1 | > 1 | 51 | 0.5-1.1 | | 1.2-1.3 | > 1.3 | 33 |
|  | RVEDVi | 57-101 | 102-109 | > 109 | 51 | 60-134 | | 135-151 | > 151 | 34 |
|  | RVESVi | 17-42 | 43-49 | > 49 | 51 | 25-63 | | 64-75 | > 75 | 35 |
|  | RVEF | 50-75 | 44-49 | < 44 | 51 | 47-72 | | 44-46 | < 44 | 35 |
|  | RVSVi | 32-68 | 26-31 | < 26 | 51 | 31-79 | | 25-30 | < 25 | 33 |
|  | LAESVi | 20-59 | 60-69 | > 69 | 43 | 18-56 | | 57-67 | > 67 | 34 |
|  | LAEF | 53-83 | 49-52 | < 49 | 42 | 50-83 | | 45-49 | < 45 | 34 |
|  | RAESVi | 27-57 | 58-64 | > 64 | 36 | 24-70 | | 71-79 | > 79 | 21 |
|  | RAEF | 38-74 | 33-37 | < 33 | 34 | 35-68 | | 30-34 | < 30 | 21 |

Footnote: All measurements were derived using suiteHEART software with smooth endocardial segmentation; papillary muscles and trabeculations were excluded from left ventricular mass measurements. Volumetric and mass measurements were indexed to height. Reference ranges are defined as the interval between the 2.5th and 97.5th centiles derived from age-specific generalized additive models for location, scale and shape (GAMLSS) using the Box–Cox Cole–Green distribution. Centile-based categories beyond the reference range are derived from LMS-based thresholds (P0.5, P2.5, P97.5, and P99.5) and describe the position of a measurement within the reference distribution rather than disease severity or diagnostic abnormality. Reference ranges are based on a Mixed adult population.

Abbreviations: LV = left ventricle; RV = right ventricle; LA = left atrium; RA = right atrium; EDV = end-diastolic volume; ESV = end-systolic volume; SV = stroke volume; EF = ejection fraction; CO = cardiac output.

### 21. Table: Reference ranges and centile-based categories for cardiac chamber measurements derived using suiteHEART software (anatomical segmentation), indexed to height, in a White adult population

|  |  | **Female** | | | | **Male** | | | |
| --- | --- | --- | --- | --- | --- | --- | --- | --- | --- |
| **Age group** | **Metrics** | **Reference range** | **Mild** | **At least moderate** | **n** | **Reference range** | **Mild** | **At least moderate** | **n** |
| 18-29 | LVEDVi | 56-102 | 103-108 | > 108 | 101 | 71-121 | 122-129 | > 129 | 101 |
|  | LVESVi | 18-41 | 42-46 | > 46 | 101 | 20-50 | 51-56 | > 56 | 103 |
|  | LVSVi | 34-67 | 28-33 | < 28 | 101 | 42-78 | 37-41 | < 37 | 101 |
|  | LVEF | 55-74 | 51-54 | < 51 | 101 | 52-73 | 49-51 | < 49 | 104 |
|  | LVMi | 40-70 | 71-76 | > 76 | 101 | 55-94 | 95-99 | > 99 | 101 |
|  | LVM/LVEDV | 0.5-0.9 | 1-1.1 | > 1.1 | 101 | 0.6-1.1 |  | > 1.2 | 99 |
|  | RVEDVi | 61-115 | 116-122 | > 122 | 101 | 75-137 | 138-145 | > 145 | 98 |
|  | RVESVi | 24-52 | 53-57 | > 57 | 100 | 25-64 | 65-70 | > 70 | 99 |
|  | RVEF | 47-70 | 44-46 | < 44 | 99 | 46-68 | 43-45 | < 43 | 102 |
|  | RVSVi | 37-72 | 32-36 | < 32 | 100 | 43-81 | 36-42 | < 36 | 101 |
|  | LAESV | 19-43 | 44-47 | > 47 | 65 | 22-55 | 56-62 | > 62 | 64 |
|  | LAEF | 57-85 | 52-56 | < 52 | 65 | 55-79 | 51-54 | < 51 | 62 |
|  | RAESV | 24-50 | 51-55 | > 55 | 77 | 26-67 | 68-75 | > 75 | 87 |
|  | RAEF | 42-75 | 38-41 | < 38 | 72 | 35-71 | 31-34 | < 31 | 85 |
| 30-39 | LVEDVi | 54-99 | 100-106 | > 106 | 131 | 63-113 | 114-121 | > 121 | 128 |
|  | LVESVi | 16-40 | 41-44 | > 44 | 131 | 19-49 | 50-54 | > 54 | 128 |
|  | LVSVi | 33-65 | 28-32 | < 28 | 130 | 39-74 | 34-38 | < 34 | 128 |
|  | LVEF | 56-76 | 52-55 | < 52 | 131 | 53-75 | 50-52 | < 50 | 129 |
|  | LVMi | 40-73 | 74-80 | > 80 | 131 | 55-94 | 95-100 | > 100 | 125 |
|  | LVM/LVEDV | 0.6-1 | 1.1-1.2 | > 1.2 | 131 | 0.6-1.2 | 1.3-1.3 | > 1.3 | 126 |
|  | RVEDVi | 59-112 | 113-120 | > 120 | 131 | 73-136 | 137-145 | > 145 | 127 |
|  | RVESVi | 22-50 | 51-55 | > 55 | 131 | 23-62 | 63-68 | > 68 | 128 |
|  | RVEF | 49-71 | 45-48 | < 45 | 131 | 47-71 | 43-46 | < 43 | 129 |
|  | RVSVi | 36-72 | 31-35 | < 31 | 131 | 41-82 | 34-40 | < 34 | 126 |
|  | LAESV | 20-48 | 49-53 | > 53 | 95 | 21-56 | 57-63 | > 63 | 86 |
|  | LAEF | 57-84 | 53-56 | < 53 | 95 | 55-80 | 51-54 | < 51 | 87 |
|  | RAESV | 24-53 | 54-59 | > 59 | 113 | 26-68 | 69-76 | > 76 | 104 |
|  | RAEF | 41-75 | 37-40 | < 37 | 108 | 36-72 | 31-35 | < 31 | 103 |
| 40-49 | LVEDVi | 54-100 | 101-108 | > 108 | 521 | 62-116 | 117-126 | > 126 | 437 |
|  | LVESVi | 14-38 | 39-43 | > 43 | 521 | 17-47 | 48-53 | > 53 | 440 |
|  | LVSVi | 35-67 | 30-34 | < 30 | 521 | 39-77 | 33-38 | < 33 | 433 |
|  | LVEF | 57-78 | 53-56 | < 53 | 520 | 54-77 | 51-53 | < 51 | 442 |
|  | LVMi | 40-77 | 78-85 | > 85 | 521 | 55-96 | 97-103 | > 103 | 414 |
|  | LVM/LVEDV | 0.6-1.1 |  | > 1.2 | 521 | 0.6-1.2 | 1.3-1.4 | > 1.4 | 411 |
|  | RVEDVi | 57-110 | 111-119 | > 119 | 521 | 70-135 | 136-145 | > 145 | 434 |
|  | RVESVi | 19-47 | 48-53 | > 53 | 521 | 22-60 | 61-66 | > 66 | 441 |
|  | RVEF | 51-73 | 47-50 | < 47 | 517 | 48-72 | 43-47 | < 43 | 442 |
|  | RVSVi | 34-72 | 30-33 | < 30 | 519 | 40-83 | 33-39 | < 33 | 428 |
|  | LAESV | 20-52 | 53-58 | > 58 | 453 | 20-56 | 57-64 | > 64 | 374 |
|  | LAEF | 57-83 | 53-56 | < 53 | 453 | 55-81 | 51-54 | < 51 | 374 |
|  | RAESV | 24-57 | 58-63 | > 63 | 452 | 26-69 | 70-77 | > 77 | 362 |
|  | RAEF | 41-75 | 36-40 | < 36 | 445 | 36-73 | 31-35 | < 31 | 360 |
| 50-59 | LVEDVi | 54-102 | 103-112 | > 112 | 912 | 60-118 | 119-129 | > 129 | 831 |
|  | LVESVi | 13-37 | 38-42 | > 42 | 912 | 15-46 | 47-52 | > 52 | 835 |
|  | LVSVi | 37-71 | 32-36 | < 32 | 912 | 39-79 | 33-38 | < 33 | 825 |
|  | LVEF | 58-80 | 54-57 | < 54 | 911 | 56-79 | 51-55 | < 51 | 833 |
|  | LVMi | 41-80 | 81-89 | > 89 | 911 | 54-98 | 99-105 | > 105 | 772 |
|  | LVM/LVEDV | 0.6-1.1 | 1.2-1.3 | > 1.3 | 911 | 0.7-1.3 | 1.4-1.5 | > 1.5 | 770 |
|  | RVEDVi | 55-108 | 109-118 | > 118 | 910 | 65-129 | 130-139 | > 139 | 826 |
|  | RVESVi | 16-44 | 45-49 | > 49 | 910 | 20-57 | 58-64 | > 64 | 826 |
|  | RVEF | 52-75 | 48-51 | < 48 | 908 | 48-74 | 43-47 | < 43 | 831 |
|  | RVSVi | 33-72 | 29-32 | < 29 | 908 | 37-80 | 29-36 | < 29 | 824 |
|  | LAESV | 20-56 | 57-63 | > 63 | 825 | 19-57 | 58-66 | > 66 | 700 |
|  | LAEF | 56-83 | 51-55 | < 51 | 823 | 54-82 | 50-53 | < 50 | 703 |
|  | RAESV | 24-59 | 60-67 | > 67 | 769 | 25-70 | 71-79 | > 79 | 604 |
|  | RAEF | 40-75 | 35-39 | < 35 | 760 | 36-74 | 31-35 | < 31 | 604 |
| 60-69 | LVEDVi | 51-98 | 99-108 | > 108 | 920 | 56-116 | 117-127 | > 127 | 804 |
|  | LVESVi | 11-35 | 36-41 | > 41 | 919 | 13-44 | 45-51 | > 51 | 801 |
|  | LVSVi | 35-68 | 31-34 | < 31 | 920 | 37-77 | 31-36 | < 31 | 793 |
|  | LVEF | 59-81 | 54-58 | < 54 | 916 | 56-80 | 52-55 | < 52 | 803 |
|  | LVMi | 41-82 | 83-92 | > 92 | 919 | 53-98 | 99-106 | > 106 | 761 |
|  | LVM/LVEDV | 0.6-1.2 | 1.3-1.4 | > 1.4 | 919 | 0.7-1.3 | 1.4-1.6 | > 1.6 | 759 |
|  | RVEDVi | 52-106 | 107-117 | > 117 | 918 | 62-126 | 127-137 | > 137 | 803 |
|  | RVESVi | 14-41 | 42-47 | > 47 | 919 | 19-55 | 56-62 | > 62 | 804 |
|  | RVEF | 53-77 | 47-52 | < 47 | 915 | 48-76 | 43-47 | < 43 | 799 |
|  | RVSVi | 32-71 | 27-31 | < 27 | 917 | 35-80 | 27-34 | < 27 | 795 |
|  | LAESV | 19-57 | 58-66 | > 66 | 809 | 18-58 | 59-68 | > 68 | 652 |
|  | LAEF | 54-83 | 48-53 | < 48 | 807 | 53-83 | 48-52 | < 48 | 654 |
|  | RAESV | 23-61 | 62-70 | > 70 | 721 | 25-71 | 72-80 | > 80 | 521 |
|  | RAEF | 41-75 | 35-40 | < 35 | 699 | 37-74 | 32-36 | < 32 | 496 |
| 70+ | LVEDVi | 48-94 | 95-106 | > 106 | 1101 | 52-112 | 113-124 | > 124 | 1254 |
|  | LVESVi | 10-33 | 34-40 | > 40 | 1101 | 12-43 | 44-51 | > 51 | 1250 |
|  | LVSVi | 34-67 | 31-33 | < 31 | 1101 | 35-74 | 28-34 | < 28 | 1254 |
|  | LVEF | 59-83 | 55-58 | < 55 | 1086 | 56-80 | 51-55 | < 51 | 1242 |
|  | LVMi | 41-81 | 82-91 | > 91 | 1100 | 51-96 | 97-105 | > 105 | 1224 |
|  | LVM/LVEDV | 0.6-1.3 | 1.4-1.5 | > 1.5 | 1100 | 0.7-1.4 | 1.5-1.7 | > 1.7 | 1222 |
|  | RVEDVi | 49-104 | 105-116 | > 116 | 1100 | 59-123 | 124-135 | > 135 | 1252 |
|  | RVESVi | 13-40 | 41-46 | > 46 | 1100 | 17-53 | 54-61 | > 61 | 1253 |
|  | RVEF | 52-78 | 46-51 | < 46 | 1086 | 48-76 | 42-47 | < 42 | 1236 |
|  | RVSVi | 31-71 | 26-30 | < 26 | 1098 | 32-79 | 25-31 | < 25 | 1247 |
|  | LAESV | 18-59 | 60-70 | > 70 | 947 | 17-60 | 61-70 | > 70 | 953 |
|  | LAEF | 50-83 | 44-49 | < 44 | 938 | 51-84 | 45-50 | < 45 | 943 |
|  | RAESV | 23-63 | 64-73 | > 73 | 769 | 25-72 | 73-82 | > 82 | 631 |
|  | RAEF | 42-77 | 37-41 | < 37 | 753 | 37-75 | 32-36 | < 32 | 619 |

Footnote: All measurements were derived using suiteHEART software with anatomical endocardial segmentation; papillary muscles and trabeculations were included in left ventricular mass measurements. Volumetric and mass measurements were indexed to height. Reference ranges are defined as the interval between the 2.5th and 97.5th centiles derived from age-specific generalized additive models for location, scale and shape (GAMLSS) using the Box–Cox Cole–Green distribution. Centile-based categories beyond the reference range are derived from LMS-based thresholds (P0.5, P2.5, P97.5, and P99.5) and describe the position of a measurement within the reference distribution rather than disease severity or diagnostic abnormality. Reference ranges are based on a White adult population.

Abbreviations: LV = left ventricle; RV = right ventricle; LA = left atrium; RA = right atrium; EDV = end-diastolic volume; ESV = end-systolic volume; SV = stroke volume; EF = ejection fraction; CO = cardiac output.

### 22. Table: Reference ranges and centile-based categories for cardiac chamber measurements derived using suitheart software (anatomical segmentation), indexed to height, in a Black adult population

|  |  | **Female** | | | | **Male** | | | |
| --- | --- | --- | --- | --- | --- | --- | --- | --- | --- |
| **Age group** | **Metrics** | **Reference range** | **Mild** | **At least moderate** | **n** | **Reference range** | **Mild** | **At least moderate** | **n** |
| 40-49 | LVEDVi | 59-106 | 107-116 | > 116 | 10 |  |  |  |  |
|  | LVESVi | 14-40 | 41-45 | > 45 | 10 |  |  |  |  |
|  | LVSVi | 41-81 | 39-40 | < 39 | 10 |  |  |  |  |
|  | LVEF | 57-79 | 53-56 | < 53 | 10 |  |  |  |  |
|  | LVMi | 44-93 | 94-110 | > 110 | 10 |  |  |  |  |
|  | LVM/LVEDV | 0.6-1.2 | 1.3-1.4 | > 1.4 | 10 |  |  |  |  |
|  | RVEDVi | 55-116 | 117-129 | > 129 | 10 |  |  |  |  |
|  | RVESVi | 18-48 | 49-56 | > 56 | 10 |  |  |  |  |
|  | RVEF | 48-73 | 41-47 | < 41 | 10 |  |  |  |  |
|  | RVSVi | 33-76 | 28-32 | < 28 | 10 |  |  |  |  |
| 50-59 | LVEDVi | 56-103 | 104-113 | > 113 | 95 | 59-120 | 121-133 | > 133 | 77 |
|  | LVESVi | 12-37 | 38-42 | > 42 | 95 | 14-46 | 47-53 | > 53 | 76 |
|  | LVSVi | 39-73 | 36-38 | < 36 | 95 | 40-78 | 35-39 | < 35 | 75 |
|  | LVEF | 58-80 | 55-57 | < 55 | 92 | 56-80 | 52-55 | < 52 | 76 |
|  | LVMi | 45-93 | 94-109 | > 109 | 95 | 56-99 | 100-107 | > 107 | 68 |
|  | LVM/LVEDV | 0.6-1.2 | 1.3-1.4 | > 1.4 | 95 | 0.7-1.3 | 1.4-1.5 | > 1.5 | 68 |
|  | RVEDVi | 55-112 | 113-123 | > 123 | 95 | 69-124 | 125-137 | > 137 | 77 |
|  | RVESVi | 17-48 | 49-55 | > 55 | 95 | 22-56 | 57-67 | > 67 | 77 |
|  | RVEF | 50-75 | 45-49 | < 45 | 95 | 48-74 | 41-47 | < 41 | 77 |
|  | RVSVi | 33-72 | 28-32 | < 28 | 95 | 40-78 | 35-39 | < 35 | 76 |
|  | LAESVi | 22-54 | 55-60 | > 60 | 88 | 21-57 | 58-68 | > 68 | 67 |
|  | LAEF | 59-79 | 56-58 | < 56 | 88 | 56-83 | 53-55 | < 53 | 67 |
|  | RAESVi | 23-64 | 65-73 | > 73 | 84 | 28-66 | 67-72 | > 72 | 64 |
|  | RAEF | 38-69 | 32-37 | < 32 | 84 | 38-65 | 35-37 | < 35 | 63 |
| 60-69 | LVEDVi | 50-97 | 98-106 | > 106 | 65 | 55-115 | 116-129 | > 129 | 48 |
|  | LVESVi | 9-33 | 34-38 | > 38 | 65 | 11-46 | 47-55 | > 55 | 48 |
|  | LVSVi | 35-67 | 30-34 | < 30 | 65 | 41-77 | 38-40 | < 38 | 48 |
|  | LVEF | 60-83 | 56-59 | < 56 | 64 | 48-81 | 36-47 | < 36 | 48 |
|  | LVMi | 47-94 | 95-108 | > 108 | 65 | 56-99 | 100-108 | > 108 | 44 |
|  | LVM/LVEDV | 0.7-1.3 | 1.4-1.5 | > 1.5 | 65 | 0.7-1.3 | 1.4-1.5 | > 1.5 | 44 |
|  | RVEDVi | 55-108 | 109-119 | > 119 | 65 | 64-121 | 122-134 | > 134 | 48 |
|  | RVESVi | 14-47 | 48-54 | > 54 | 65 | 17-56 | 57-68 | > 68 | 48 |
|  | RVEF | 51-79 | 47-50 | < 47 | 64 | 48-77 | 42-47 | < 42 | 47 |
|  | RVSVi | 34-71 | 29-33 | < 29 | 65 | 37-77 | 32-36 | < 32 | 48 |
|  | LAESVi | 20-56 | 57-64 | > 64 | 60 | 19-59 | 60-70 | > 70 | 42 |
|  | LAEF | 54-81 | 49-53 | < 49 | 61 | 52-85 | 48-51 | < 48 | 42 |
|  | RAESVi | 23-61 | 62-69 | > 69 | 52 | 24-72 | 73-79 | > 79 | 40 |
|  | RAEF | 32-66 | 21-31 | < 21 | 53 | 36-71 | 32-35 | < 32 | 41 |
| 70+ | LVEDVi | 44-92 | 93-100 | > 100 | 18 | 52-110 | 111-126 | > 126 | 14 |
|  | LVESVi | 8-33 | 34-39 | > 39 | 18 | 9-47 | 48-59 | > 59 | 14 |
|  | LVSVi | 35-66 | 31-34 | < 31 | 18 | 40-79 | 38-39 | < 38 | 14 |
|  | LVEF | 60-87 | 55-59 | < 55 | 18 | 53-82 | 44-52 | < 44 | 14 |
|  | LVMi | 50-94 | 95-108 | > 108 | 18 | 54-96 | 97-106 | > 106 | 14 |
|  | LVM/LVEDV | 0.8-1.4 | 1.5-1.6 | > 1.6 | 18 | 0.7-1.4 | 1.5-1.6 | > 1.6 | 14 |
|  | RVEDVi | 55-108 | 109-121 | > 121 | 18 | 60-115 | 116-128 | > 128 | 14 |
|  | RVESVi | 12-45 | 46-54 | > 54 | 18 | 13-56 | 57-68 | > 68 | 14 |
|  | RVEF | 48-80 | 37-47 | < 37 | 18 | 48-83 | 43-47 | < 43 | 14 |
|  | RVSVi | 35-76 | 32-34 | < 32 | 18 | 37-73 | 31-36 | < 31 | 14 |
|  | LAESVi | 18-59 | 60-68 | > 68 | 16 |  |  |  |  |
|  | LAEF | 47-82 | 39-46 | < 39 | 17 |  |  |  |  |
|  | RAESVi | 24-58 | 59-65 | > 65 | 13 |  |  |  |  |
|  | RAEF | 35-71 | 25-34 | < 25 | 12 |  |  |  |  |

Footnote: All measurements were derived using suiteHEART software with anatomical endocardial segmentation; papillary muscles and trabeculations were included in left ventricular mass measurements. Volumetric and mass measurements were indexed to height. Reference ranges are defined as the interval between the 2.5th and 97.5th centiles derived from age-specific generalized additive models for location, scale and shape (GAMLSS) using the Box–Cox Cole–Green distribution. Centile-based categories beyond the reference range are derived from LMS-based thresholds (P0.5, P2.5, P97.5, and P99.5) and describe the position of a measurement within the reference distribution rather than disease severity or diagnostic abnormality. Reference ranges are based on a Black adult population.

Abbreviations: LV = left ventricle; RV = right ventricle; LA = left atrium; RA = right atrium; EDV = end-diastolic volume; ESV = end-systolic volume; SV = stroke volume; EF = ejection fraction; CO = cardiac output.

### 23. Table: Reference ranges and centile-based categories for cardiac chamber measurements derived using suiteHEART software (anatomical segmentation), indexed to height, in an Asian adult population

|  |  | **Female** | | | | **Male** | | | |
| --- | --- | --- | --- | --- | --- | --- | --- | --- | --- |
| **Age group** | **Metrics** | **Reference range** | **Mild** | **At least moderate** | **n** | **Reference range** | **Mild** | **At least moderate** | **n** |
| 40-49 | LVEDVi | 47-86 | 87-92 | > 92 | 10 | 56-106 | 107-116 | > 116 | 20 |
|  | LVESVi | 11-36 | 37-45 | > 45 | 10 | 14-38 | 39-43 | > 43 | 20 |
|  | LVSVi | 32-59 | 27-31 | < 27 | 10 | 41-73 | 38-40 | < 38 | 20 |
|  | LVEF | 59-80 | 54-58 | < 54 | 10 | 59-79 | 55-58 | < 55 | 20 |
|  | LVMi | 35-70 | 71-80 | > 80 | 10 | 46-83 | 84-88 | > 88 | 20 |
|  | LVM/LVEDV | 0.6-1.1 | 1.2-1.3 | > 1.3 | 10 | 0.6-1.1 |  | > 1.2 | 20 |
|  | RVEDVi | 49-88 | 89-96 | > 96 | 10 | 60-111 | 112-119 | > 119 | 20 |
|  | RVESVi | 14-36 | 37-41 | > 41 | 10 | 17-48 | 49-53 | > 53 | 20 |
|  | RVEF | 51-78 | 46-50 | < 46 | 10 | 48-72 | 43-47 | < 43 | 19 |
|  | RVSVi | 30-57 | 27-29 | < 27 | 10 | 36-71 | 31-35 | < 31 | 20 |
|  | LAESVi | 21-46 | 47-51 | > 51 | 10 | 16-50 | 51-57 | > 57 | 19 |
|  | LAEF | 59-85 | 55-58 | < 55 | 10 | 61-81 | 57-60 | < 57 | 19 |
|  | RAESVi | 22-48 | 49-54 | > 54 | 10 | 20-58 | 59-63 | > 63 | 17 |
|  | RAEF | 39-69 | 31-38 | < 31 | 10 | 35-71 | 30-34 | < 30 | 17 |
| 50-59 | LVEDVi | 47-85 | 86-92 | > 92 | 83 | 52-101 | 102-110 | > 110 | 132 |
|  | LVESVi | 10-34 | 35-43 | > 43 | 83 | 13-37 | 38-43 | > 43 | 132 |
|  | LVSVi | 33-59 | 28-32 | < 28 | 83 | 36-70 | 31-35 | < 31 | 132 |
|  | LVEF | 58-81 | 52-57 | < 52 | 83 | 58-80 | 54-57 | < 54 | 132 |
|  | LVMi | 35-73 | 74-84 | > 84 | 83 | 44-87 | 88-94 | > 94 | 130 |
|  | LVM/LVEDV | 0.6-1.1 | 1.2-1.3 | > 1.3 | 83 | 0.7-1.2 | 1.3-1.3 | > 1.3 | 130 |
|  | RVEDVi | 48-90 | 91-100 | > 100 | 83 | 57-109 | 110-118 | > 118 | 131 |
|  | RVESVi | 13-35 | 36-40 | > 40 | 83 | 16-47 | 48-53 | > 53 | 131 |
|  | RVEF | 52-78 | 47-51 | < 47 | 82 | 48-74 | 43-47 | < 43 | 131 |
|  | RVSVi | 31-62 | 28-30 | < 28 | 82 | 34-71 | 29-33 | < 29 | 131 |
|  | LAESVi | 20-47 | 48-53 | > 53 | 77 | 17-51 | 52-58 | > 58 | 121 |
|  | LAEF | 58-84 | 53-57 | < 53 | 77 | 58-81 | 54-57 | < 54 | 122 |
|  | RAESVi | 22-51 | 52-59 | > 59 | 68 | 21-59 | 60-66 | > 66 | 118 |
|  | RAEF | 38-69 | 30-37 | < 30 | 68 | 38-71 | 33-37 | < 33 | 115 |
| 60-69 | LVEDVi | 45-84 | 85-92 | > 92 | 63 | 48-97 | 98-106 | > 106 | 118 |
|  | LVESVi | 9-32 | 33-39 | > 39 | 63 | 11-38 | 39-45 | > 45 | 119 |
|  | LVSVi | 33-60 | 30-32 | < 30 | 63 | 33-68 | 28-32 | < 28 | 118 |
|  | LVEF | 58-83 | 53-57 | < 53 | 63 | 57-81 | 51-56 | < 51 | 119 |
|  | LVMi | 36-78 | 79-92 | > 92 | 63 | 43-90 | 91-102 | > 102 | 116 |
|  | LVM/LVEDV | 0.6-1.2 | 1.3-1.4 | > 1.4 | 63 | 0.7-1.3 | 1.4-1.5 | > 1.5 | 116 |
|  | RVEDVi | 46-94 | 95-105 | > 105 | 63 | 51-107 | 108-116 | > 116 | 119 |
|  | RVESVi | 13-34 | 35-39 | > 39 | 63 | 16-46 | 47-53 | > 53 | 119 |
|  | RVEF | 54-78 | 49-53 | < 49 | 63 | 49-76 | 43-48 | < 43 | 119 |
|  | RVSVi | 31-69 | 29-30 | < 29 | 63 | 31-70 | 25-30 | < 25 | 119 |
|  | LAESVi | 19-50 | 51-57 | > 57 | 60 | 17-52 | 53-60 | > 60 | 111 |
|  | LAEF | 55-82 | 51-54 | < 51 | 60 | 55-82 | 51-54 | < 51 | 111 |
|  | RAESVi | 22-57 | 58-67 | > 67 | 51 | 21-62 | 63-71 | > 71 | 86 |
|  | RAEF | 37-72 | 29-36 | < 29 | 50 | 40-71 | 37-39 | < 37 | 86 |
| 70+ | LVEDVi | 44-84 | 85-93 | > 93 | 28 | 46-95 | 96-105 | > 105 | 40 |
|  | LVESVi | 9-29 | 30-36 | > 36 | 28 | 10-38 | 39-48 | > 48 | 40 |
|  | LVSVi | 33-61 | 30-32 | < 30 | 28 | 33-69 | 30-32 | < 30 | 40 |
|  | LVEF | 63-83 | 60-62 | < 60 | 27 | 54-82 | 47-53 | < 47 | 40 |
|  | LVMi | 37-83 | 84-102 | > 102 | 28 | 45-88 | 89-101 | > 101 | 40 |
|  | LVM/LVEDV | 0.7-1.3 | 1.4-1.5 | > 1.5 | 28 | 0.7-1.5 | 1.6-1.8 | > 1.8 | 40 |
|  | RVEDVi | 44-98 | 99-110 | > 110 | 27 | 47-105 | 106-115 | > 115 | 40 |
|  | RVESVi | 13-33 | 34-37 | > 37 | 27 | 14-46 | 47-56 | > 56 | 40 |
|  | RVEF | 55-78 | 51-54 | < 51 | 27 | 49-78 | 43-48 | < 43 | 40 |
|  | RVSVi | 28-67 | 24-27 | < 24 | 27 | 28-69 | 22-27 | < 22 | 40 |
|  | LAESVi | 18-53 | 54-60 | > 60 | 29 | 18-54 | 55-61 | > 61 | 34 |
|  | LAEF | 52-81 | 48-51 | < 48 | 28 | 52-83 | 47-51 | < 47 | 34 |
|  | RAESVi | 21-64 | 65-77 | > 77 | 21 | 21-66 | 67-77 | > 77 | 26 |
|  | RAEF | 39-79 | 30-38 | < 30 | 19 | 43-71 | 40-42 | < 40 | 25 |

Footnote: All measurements were derived using suiteHEART software with anatomical endocardial segmentation; papillary muscles and trabeculations were included in left ventricular mass measurements. Volumetric and mass measurements were indexed to height. Reference ranges are defined as the interval between the 2.5th and 97.5th centiles derived from age-specific generalized additive models for location, scale and shape (GAMLSS) using the Box–Cox Cole–Green distribution. Centile-based categories beyond the reference range are derived from LMS-based thresholds (P0.5, P2.5, P97.5, and P99.5) and describe the position of a measurement within the reference distribution rather than disease severity or diagnostic abnormality. Reference ranges are based on an Asian adult population.

Abbreviations: LV = left ventricle; RV = right ventricle; LA = left atrium; RA = right atrium; EDV = end-diastolic volume; ESV = end-systolic volume; SV = stroke volume; EF = ejection fraction; CO = cardiac output.

### 24. Table: Reference ranges and centile-based categories for cardiac chamber measurements derived using suiteHEART software (anatomical segmentation), indexed to height, in a Chinese adult population

|  |  | **Female** | | | | | **Male** | | | |
| --- | --- | --- | --- | --- | --- | --- | --- | --- | --- | --- |
| **Age group** | **Metrics** | **Reference range** | **Mild** | **At least moderate** | **n** | **Reference range** | | **Mild** | **At least moderate** | **n** |
| 18-29 | LVEDVi | 53-101 | 102-117 | > 117 | 16 | 69-104 | | 105-110 | > 110 | 19 |
|  | LVESVi | 16-38 | 39-47 | > 47 | 16 | 21-43 | | 44-51 | > 51 | 19 |
|  | LVSVi | 33-72 | 30-32 | < 30 | 16 | 46-69 | | 43-45 | < 43 | 19 |
|  | LVEF | 58-76 | 56-57 | < 56 | 16 | 56-73 | | 51-55 | < 51 | 19 |
|  | LVMi | 30-60 | 61-68 | > 68 | 16 | 47-84 | | 85-90 | > 90 | 19 |
|  | LVM/LVEDV | 0.5-0.8 | 0.9-0.9 | > 0.9 | 16 | 0.6-1 | | 1.1-1.1 | > 1.1 | 19 |
|  | RVEDVi | 58-106 | 107-117 | > 117 | 16 | 78-119 | | 120-123 | > 123 | 19 |
|  | RVESVi | 22-48 | 49-54 | > 54 | 16 | 30-60 | | 61-66 | > 66 | 19 |
|  | RVEF | 45-68 | 41-44 | < 41 | 16 | 48-66 | | 45-47 | < 45 | 19 |
|  | RVSVi | 30-62 | 26-29 | < 26 | 16 | 46-70 | | 42-45 | < 42 | 19 |
|  | LAESVi | 20-50 | 51-58 | > 58 | 12 | 22-47 | | 48-52 | > 52 | 15 |
|  | LAEF | 55-79 | 49-54 | < 49 | 12 | 53-77 | | 50-52 | < 50 | 15 |
|  | RAESVi | 23-47 | 48-54 | > 54 | 11 | 25-68 | | 69-82 | > 82 | 15 |
|  | RAEF | 47-69 | 45-46 | < 45 | 11 | 39-64 | | 33-38 | < 33 | 14 |
| 30-39 | LVEDVi | 52-96 | 97-109 | > 109 | 18 | 66-102 | | 103-109 | > 109 | 17 |
|  | LVESVi | 15-36 | 37-44 | > 44 | 18 | 19-41 | | 42-48 | > 48 | 17 |
|  | LVSVi | 33-68 | 30-32 | < 30 | 18 | 43-69 | | 40-42 | < 40 | 17 |
|  | LVEF | 59-77 | 56-58 | < 56 | 18 | 56-75 | | 51-55 | < 51 | 17 |
|  | LVMi | 32-63 | 64-73 | > 73 | 18 | 47-85 | | 86-92 | > 92 | 17 |
|  | LVM/LVEDV | 0.5-0.8 | 0.9-0.9 | > 0.9 | 18 | 0.6-1 | | 1.1-1.2 | > 1.2 | 17 |
|  | RVEDVi | 56-103 | 104-113 | > 113 | 18 | 74-115 | | 116-121 | > 121 | 17 |
|  | RVESVi | 19-46 | 47-51 | > 51 | 18 | 27-57 | | 58-63 | > 63 | 17 |
|  | RVEF | 47-70 | 42-46 | < 42 | 18 | 48-68 | | 45-47 | < 45 | 17 |
|  | RVSVi | 30-62 | 25-29 | < 25 | 18 | 43-69 | | 40-42 | < 40 | 17 |
|  | LAESVi | 20-49 | 50-55 | > 55 | 16 | 21-47 | | 48-51 | > 51 | 17 |
|  | LAEF | 55-79 | 49-54 | < 49 | 16 | 53-78 | | 50-52 | < 50 | 17 |
|  | RAESVi | 23-49 | 50-57 | > 57 | 16 | 25-66 | | 67-78 | > 78 | 17 |
|  | RAEF | 48-71 | 45-47 | < 45 | 14 | 39-66 | | 33-38 | < 33 | 17 |
| 40-49 | LVEDVi | 50-91 | 92-102 | > 102 | 24 | 62-100 | | 101-108 | > 108 | 20 |
|  | LVESVi | 13-33 | 34-40 | > 40 | 24 | 17-39 | | 40-45 | > 45 | 20 |
|  | LVSVi | 35-62 | 32-34 | < 32 | 24 | 40-69 | | 37-39 | < 37 | 20 |
|  | LVEF | 60-79 | 57-59 | < 57 | 24 | 56-76 | | 52-55 | < 52 | 20 |
|  | LVMi | 34-68 | 69-81 | > 81 | 24 | 48-87 | | 88-96 | > 96 | 20 |
|  | LVM/LVEDV | 0.5-0.9 | 01-1 | > 1 | 24 | 0.6-1.1 | | 1.2-1.3 | > 1.3 | 20 |
|  | RVEDVi | 53-99 | 100-109 | > 109 | 24 | 69-112 | | 113-119 | > 119 | 20 |
|  | RVESVi | 17-43 | 44-49 | > 49 | 24 | 23-54 | | 55-60 | > 60 | 20 |
|  | RVEF | 48-73 | 43-47 | < 43 | 24 | 48-71 | | 45-47 | < 45 | 20 |
|  | RVSVi | 29-61 | 25-28 | < 25 | 24 | 40-69 | | 36-39 | < 36 | 20 |
|  | LAESVi | 20-47 | 48-52 | > 52 | 20 | 20-47 | | 48-51 | > 51 | 19 |
|  | LAEF | 55-80 | 50-54 | < 50 | 20 | 53-80 | | 49-52 | < 49 | 19 |
|  | RAESVi | 23-52 | 53-60 | > 60 | 21 | 24-65 | | 66-74 | > 74 | 20 |
|  | RAEF | 48-73 | 45-47 | < 45 | 20 | 39-68 | | 33-38 | < 33 | 20 |
| 50-59 | LVEDVi | 48-86 | 87-95 | > 95 | 66 | 58-99 | | 100-108 | > 108 | 49 |
|  | LVESVi | 11-31 | 32-36 | > 36 | 66 | 16-37 | | 38-43 | > 43 | 49 |
|  | LVSVi | 35-59 | 32-34 | < 32 | 66 | 37-69 | | 34-36 | < 34 | 49 |
|  | LVEF | 61-81 | 57-60 | < 57 | 66 | 56-78 | | 52-55 | < 52 | 49 |
|  | LVMi | 37-72 | 73-90 | > 90 | 66 | 48-89 | | 90-100 | > 100 | 49 |
|  | LVM/LVEDV | 0.6-1 | 1.1-1.2 | > 1.2 | 66 | 0.6-1.2 | | 1.3-1.3 | > 1.3 | 49 |
|  | RVEDVi | 49-94 | 95-104 | > 104 | 66 | 65-110 | | 111-119 | > 119 | 49 |
|  | RVESVi | 14-40 | 41-46 | > 46 | 66 | 20-50 | | 51-56 | > 56 | 49 |
|  | RVEF | 50-75 | 44-49 | < 44 | 66 | 48-74 | | 45-47 | < 45 | 49 |
|  | RVSVi | 29-61 | 24-28 | < 24 | 66 | 37-70 | | 33-36 | < 33 | 49 |
|  | LAESVi | 20-46 | 47-52 | > 52 | 59 | 19-46 | | 47-50 | > 50 | 45 |
|  | LAEF | 55-80 | 50-54 | < 50 | 58 | 53-81 | | 48-52 | < 48 | 45 |
|  | RAESVi | 24-54 | 55-64 | > 64 | 61 | 23-63 | | 64-71 | > 71 | 44 |
|  | RAEF | 47-74 | 44-46 | < 44 | 60 | 39-70 | | 33-38 | < 33 | 44 |
| 60-69 | LVEDVi | 46-82 | 83-90 | > 90 | 52 | 55-98 | | 99-108 | > 108 | 37 |
|  | LVESVi | 9-28 | 29-33 | > 33 | 52 | 14-35 | | 36-40 | > 40 | 37 |
|  | LVSVi | 34-59 | 30-33 | < 30 | 52 | 34-70 | | 30-33 | < 30 | 37 |
|  | LVEF | 61-83 | 57-60 | < 57 | 51 | 57-81 | | 53-56 | < 53 | 37 |
|  | LVMi | 39-77 | 78-103 | > 103 | 52 | 48-92 | | 93-108 | > 108 | 37 |
|  | LVM/LVEDV | 0.6-1.2 | 1.3-1.5 | > 1.5 | 52 | 0.6-1.3 | |  | > 1.4 | 37 |
|  | RVEDVi | 46-90 | 91-98 | > 98 | 52 | 61-110 | | 111-123 | > 123 | 37 |
|  | RVESVi | 12-37 | 38-43 | > 43 | 52 | 19-44 | | 45-49 | > 49 | 37 |
|  | RVEF | 52-78 | 45-51 | < 45 | 51 | 48-77 | | 45-47 | < 45 | 37 |
|  | RVSVi | 28-60 | 24-27 | < 24 | 52 | 33-73 | | 30-32 | < 30 | 37 |
|  | LAESVi | 18-49 | 50-57 | > 57 | 48 | 17-44 | | 45-49 | > 49 | 31 |
|  | LAEF | 54-81 | 49-53 | < 49 | 48 | 52-83 | | 47-51 | < 47 | 31 |
|  | RAESVi | 24-57 | 58-69 | > 69 | 45 | 23-63 | | 64-70 | > 70 | 32 |
|  | RAEF | 46-74 | 42-45 | < 42 | 45 | 39-72 | | 33-38 | < 33 | 31 |
| 70+ | LVEDVi | 44-78 | 79-85 | > 85 | 12 |  | |  |  |  |
|  | LVESVi | 7-25 | 26-29 | > 29 | 12 |  | |  |  |  |
|  | LVSVi | 30-62 | 26-29 | < 26 | 12 |  | |  |  |  |
|  | LVEF | 61-85 | 55-60 | < 55 | 12 |  | |  |  |  |
|  | LVMi | 41-82 | 83-116 | > 116 | 12 |  | |  |  |  |
|  | LVM/LVEDV | 0.7-1.4 | 1.5-1.7 | > 1.7 | 12 |  | |  |  |  |
|  | RVEDVi | 42-85 | 86-93 | > 93 | 12 |  | |  |  |  |
|  | RVESVi | 10-34 | 35-40 | > 40 | 12 |  | |  |  |  |
|  | RVEF | 53-80 | 46-52 | < 46 | 12 |  | |  |  |  |
|  | RVSVi | 28-60 | 24-27 | < 24 | 12 |  | |  |  |  |
|  | LAESVi | 17-57 | 58-71 | > 71 | 12 |  | |  |  |  |
|  | LAEF | 54-82 | 48-53 | < 48 | 12 |  | |  |  |  |
|  | RAESVi | 24-60 | 61-74 | > 74 | 10 |  | |  |  |  |
|  | RAEF | 44-73 | 41-43 | < 41 | 10 |  | |  |  |  |

Footnote: All measurements were derived using suiteHEART software with anatomical endocardial segmentation; papillary muscles and trabeculations were included in left ventricular mass measurements. Volumetric and mass measurements were indexed to height. Reference ranges are defined as the interval between the 2.5th and 97.5th centiles derived from age-specific generalized additive models for location, scale and shape (GAMLSS) using the Box–Cox Cole–Green distribution. Centile-based categories beyond the reference range are derived from LMS-based thresholds (P0.5, P2.5, P97.5, and P99.5) and describe the position of a measurement within the reference distribution rather than disease severity or diagnostic abnormality. Reference ranges are based on a Chinese adult population.

Abbreviations: LV = left ventricle; RV = right ventricle; LA = left atrium; RA = right atrium; EDV = end-diastolic volume; ESV = end-systolic volume; SV = stroke volume; EF = ejection fraction; CO = cardiac output

### 25. Table: Reference ranges and centile-based categories for cardiac chamber measurements derived using suitheart software (anatomical segmentation), indexed to height, in a Mixed adult population

|  |  | **Female** | | | | | **Male** | | | |
| --- | --- | --- | --- | --- | --- | --- | --- | --- | --- | --- |
| **Age group** | **Metrics** | **Reference range** | **Mild** | **At least moderate** | **n** | **Reference range** | | **Mild** | **At least moderate** | **n** |
| 50-59 | LVEDVi | 53-95 | 96-103 | > 103 | 134 | 62-116 | | 117-126 | > 126 | 66 |
|  | LVESVi | 11-32 | 33-36 | > 36 | 134 | 15-42 | | 43-48 | > 48 | 66 |
|  | LVSVi | 38-69 | 35-37 | < 35 | 133 | 39-80 | | 32-38 | < 32 | 66 |
|  | LVEF | 60-81 | 56-59 | < 56 | 133 | 58-82 | | 55-57 | < 55 | 66 |
|  | LVMi | 38-82 | 83-96 | > 96 | 134 | 47-98 | | 99-106 | > 106 | 62 |
|  | LVM/LVEDV | 0.6-1.1 |  | > 1.2 | 134 | 0.6-1.3 | | 1.4-1.9 | > 1.9 | 62 |
|  | RVEDVi | 51-101 | 102-109 | > 109 | 134 | 66-128 | | 129-140 | > 140 | 66 |
|  | RVESVi | 15-40 | 41-45 | > 45 | 134 | 20-52 | | 53-58 | > 58 | 66 |
|  | RVEF | 54-76 | 51-53 | < 51 | 133 | 47-76 | | 40-46 | < 40 | 66 |
|  | RVSVi | 32-69 | 26-31 | < 26 | 134 | 37-79 | | 29-36 | < 29 | 66 |
|  | LAESVi | 20-51 | 52-57 | > 57 | 122 | 21-57 | | 58-64 | > 64 | 53 |
|  | LAEF | 57-80 | 51-56 | < 51 | 122 | 56-81 | | 51-55 | < 51 | 54 |
|  | RAESVi | 23-60 | 61-69 | > 69 | 117 | 27-68 | | 69-76 | > 76 | 54 |
|  | RAEF | 41-72 | 37-40 | < 37 | 117 | 37-72 | | 32-36 | < 32 | 54 |
| 60-69 | LVEDVi | 49-94 | 95-103 | > 103 | 110 | 54-111 | | 112-121 | > 121 | 59 |
|  | LVESVi | 10-32 | 33-37 | > 37 | 110 | 13-42 | | 43-49 | > 49 | 59 |
|  | LVSVi | 35-67 | 32-34 | < 32 | 110 | 36-74 | | 30-35 | < 30 | 59 |
|  | LVEF | 60-83 | 55-59 | < 55 | 109 | 56-80 | | 51-55 | < 51 | 59 |
|  | LVMi | 38-79 | 80-87 | > 87 | 110 | 50-98 | | 99-107 | > 107 | 58 |
|  | LVM/LVEDV | 0.6-1.1 | 1.2-1.3 | > 1.3 | 110 | 0.7-1.4 | | 1.5-1.8 | > 1.8 | 58 |
|  | RVEDVi | 48-104 | 105-114 | > 114 | 110 | 59-129 | | 130-143 | > 143 | 59 |
|  | RVESVi | 14-38 | 39-43 | > 43 | 110 | 19-56 | | 57-66 | > 66 | 59 |
|  | RVEF | 54-77 | 50-53 | < 50 | 110 | 49-75 | | 45-48 | < 45 | 59 |
|  | RVSVi | 29-70 | 23-28 | < 23 | 110 | 33-78 | | 27-32 | < 27 | 58 |
|  | LAESVi | 19-53 | 54-59 | > 59 | 100 | 19-56 | | 57-65 | > 65 | 53 |
|  | LAEF | 55-81 | 50-54 | < 50 | 100 | 52-81 | | 48-51 | < 48 | 53 |
|  | RAESVi | 26-59 | 60-68 | > 68 | 93 | 25-69 | | 70-77 | > 77 | 40 |
|  | RAEF | 40-73 | 35-39 | < 35 | 93 | 37-72 | | 32-36 | < 32 | 40 |
| 70+ | LVEDVi | 46-92 | 93-103 | > 103 | 50 | 49-108 | | 109-121 | > 121 | 34 |
|  | LVESVi | 9-33 | 34-39 | > 39 | 51 | 11-42 | | 43-50 | > 50 | 34 |
|  | LVSVi | 31-64 | 26-30 | < 26 | 51 | 34-69 | | 28-33 | < 28 | 34 |
|  | LVEF | 59-84 | 54-58 | < 54 | 51 | 53-79 | | 43-52 | < 43 | 35 |
|  | LVMi | 38-77 | 78-83 | > 83 | 51 | 51-98 | | 99-108 | > 108 | 33 |
|  | LVM/LVEDV | 0.6-1.2 | 1.3-1.3 | > 1.3 | 50 | 0.6-1.5 | | 1.6-1.8 | > 1.8 | 33 |
|  | RVEDVi | 53-94 | 95-102 | > 102 | 51 | 53-131 | | 132-149 | > 149 | 35 |
|  | RVESVi | 14-36 | 37-42 | > 42 | 51 | 20-56 | | 57-69 | > 69 | 35 |
|  | RVEF | 54-79 | 48-53 | < 48 | 51 | 50-74 | | 47-49 | < 47 | 35 |
|  | RVSVi | 31-67 | 25-30 | < 25 | 51 | 30-78 | | 25-29 | < 25 | 33 |
|  | LAESVi | 20-59 | 60-69 | > 69 | 43 | 18-56 | | 57-67 | > 67 | 34 |
|  | LAEF | 53-83 | 49-52 | < 49 | 42 | 50-83 | | 45-49 | < 45 | 34 |
|  | RAESVi | 27-57 | 58-64 | > 64 | 36 | 24-70 | | 71-79 | > 79 | 21 |
|  | RAEF | 38-74 | 33-37 | < 33 | 34 | 35-68 | | 30-34 | < 30 | 21 |

Footnote: All measurements were derived using suiteHEART software with anatomical endocardial segmentation; papillary muscles and trabeculations were included in left ventricular mass measurements. Volumetric and mass measurements were indexed to height. Reference ranges are defined as the interval between the 2.5th and 97.5th centiles derived from age-specific generalized additive models for location, scale and shape (GAMLSS) using the Box–Cox Cole–Green distribution. Centile-based categories beyond the reference range are derived from LMS-based thresholds (P0.5, P2.5, P97.5, and P99.5) and describe the position of a measurement within the reference distribution rather than disease severity or diagnostic abnormality. Reference ranges are based on a Mixed adult population.

Abbreviations: LV = left ventricle; RV = right ventricle; LA = left atrium; RA = right atrium; EDV = end-diastolic volume; ESV = end-systolic volume; SV = stroke volume; EF = ejection fraction; CO = cardiac output

### 26. Table: Reference ranges and centile-based categories for cardiac chamber measurements derived using cvi42 software (smooth segmentation), indexed to BSA, in a White adult population

|  |  | **Female** | | | | | **Male** | | | |
| --- | --- | --- | --- | --- | --- | --- | --- | --- | --- | --- |
| **Age group** | **Metrics** | **Reference range** | **Mild** | **At least moderate** | **n** | **Reference range** | | **Mild** | **At least moderate** | **n** |
| 18-29 | LVEDVi | 58-102 | 103-109 | > 109 | 102 | 67-116 | | 117-123 | > 123 | 100 |
|  | LVESVi | 15-43 | 44-46 | > 46 | 102 | 19-48 | | 49-52 | > 52 | 100 |
|  | LVSVi | 34-65 | 30-33 | < 30 | 102 | 37-73 | | 32-36 | < 32 | 99 |
|  | LVEF | 53-72 | 51-52 | < 51 | 100 | 49-69 | | 45-48 | < 45 | 102 |
|  | LVCOi | 2.3-4.6 | 2-2.2 | < 2 | 101 | 2.2-4.8 | | 1.7-2.1 | < 1.7 | 97 |
|  | LVMi | 36-55 | 56-59 | > 59 | 102 | 44-78 | | 79-83 | > 83 | 105 |
|  | LVM/LVEDV | 0.5-0.7 | 0.8-0.8 | > 0.8 | 102 | 0.5-0.8 | | 0.9-0.9 | > 0.9 | 100 |
|  | RVEDVi | 57-104 | 105-111 | > 111 | 102 | 67-127 | | 128-136 | > 136 | 101 |
|  | RVESVi | 18-50 | 51-55 | > 55 | 100 | 23-59 | | 60-63 | > 63 | 92 |
|  | RVEF | 46-69 | 42-45 | < 42 | 97 | 44-67 | | 41-43 | < 41 | 98 |
|  | RVCOi | 1.9-5 | 1.6-1.8 | < 1.6 | 101 | 2.2-5 | | 1.7-2.1 | < 1.7 | 97 |
|  | RVSVi | 30-67 | 26-29 | < 26 | 102 | 36-75 | | 31-35 | < 31 | 98 |
|  | LAVi max | 25-50 | 51-54 | > 54 | 50 | 27-58 | | 59-67 | > 67 | 52 |
|  | LAEF | 56-75 | 54-55 | < 54 | 50 | 53-72 | | 50-52 | < 50 | 52 |
|  | RAVi max | 21-49 | 50-54 | > 54 | 80 | 25-62 | | 63-70 | > 70 | 82 |
|  | RAEF | 38-78 | 33-37 | < 33 | 78 | 35-74 | | 31-34 | < 31 | 82 |
| 30-39 | LVEDVi | 55-98 | 99-105 | > 105 | 137 | 59-106 | | 107-112 | > 112 | 133 |
|  | LVESVi | 15-41 | 42-45 | > 45 | 137 | 18-47 | | 48-50 | > 50 | 133 |
|  | LVSVi | 33-63 | 29-32 | < 29 | 134 | 34-67 | | 29-33 | < 29 | 132 |
|  | LVEF | 53-73 | 50-52 | < 50 | 132 | 50-71 | | 47-49 | < 47 | 134 |
|  | LVCOi | 2.3-4.8 | 2.1-2.2 | < 2.1 | 136 | 2.2-4.7 | | 1.8-2.1 | < 1.8 | 125 |
|  | LVMi | 35-55 | 56-59 | > 59 | 137 | 40-71 | | 72-76 | > 76 | 134 |
|  | LVM/LVEDV | 0.5-0.8 |  | > 0.8 | 137 | 0.5-0.9 | | 01-1 | > 1 | 134 |
|  | RVEDVi | 55-103 | 104-110 | > 110 | 137 | 62-120 | | 121-128 | > 128 | 131 |
|  | RVESVi | 17-47 | 48-52 | > 52 | 135 | 22-56 | | 57-60 | > 60 | 128 |
|  | RVEF | 47-71 | 44-46 | < 44 | 127 | 45-68 | | 41-44 | < 41 | 131 |
|  | RVCOi | 2-4.9 | 1.7-1.9 | < 1.7 | 133 | 2.1-4.9 | | 1.7-2 | < 1.7 | 124 |
|  | RVSVi | 31-66 | 26-30 | < 26 | 134 | 34-72 | | 29-33 | < 29 | 132 |
|  | LAVi max | 25-51 | 52-56 | > 56 | 84 | 25-56 | | 57-64 | > 64 | 66 |
|  | LAEF | 55-76 | 53-54 | < 53 | 83 | 53-73 | | 50-52 | < 50 | 66 |
|  | RAVi max | 20-50 | 51-55 | > 55 | 113 | 23-61 | | 62-69 | > 69 | 119 |
|  | RAEF | 38-75 | 33-37 | < 33 | 110 | 35-72 | | 31-34 | < 31 | 118 |
| 40-49 | LVEDVi | 55-97 | 98-105 | > 105 | 518 | 58-109 | | 110-116 | > 116 | 432 |
|  | LVESVi | 15-39 | 40-43 | > 43 | 518 | 17-45 | | 46-49 | > 49 | 427 |
|  | LVSVi | 33-63 | 29-32 | < 29 | 518 | 34-69 | | 28-33 | < 28 | 432 |
|  | LVEF | 53-73 | 50-52 | < 50 | 512 | 50-72 | | 47-49 | < 47 | 434 |
|  | LVCOi | 2.2-4.6 | 2-2.1 | < 2 | 514 | 2.1-4.5 | | 1.7-2 | < 1.7 | 425 |
|  | LVMi | 34-55 | 56-59 | > 59 | 518 | 41-71 | | 72-76 | > 76 | 441 |
|  | LVM/LVEDV | 0.5-0.8 |  | > 0.8 | 518 | 0.5-0.9 | | 01-1 | > 1 | 432 |
|  | RVEDVi | 54-101 | 102-109 | > 109 | 517 | 62-121 | | 122-129 | > 129 | 432 |
|  | RVESVi | 16-44 | 45-49 | > 49 | 517 | 21-55 | | 56-59 | > 59 | 425 |
|  | RVEF | 50-71 | 47-49 | < 47 | 510 | 46-68 | | 42-45 | < 42 | 429 |
|  | RVCOi | 2-4.6 | 1.7-1.9 | < 1.7 | 514 | 2.1-4.7 | | 1.7-2 | < 1.7 | 423 |
|  | RVSVi | 31-64 | 27-30 | < 27 | 516 | 33-72 | | 28-32 | < 28 | 429 |
|  | LAVi max | 24-54 | 55-59 | > 59 | 440 | 23-55 | | 56-62 | > 62 | 358 |
|  | LAEF | 55-78 | 52-54 | < 52 | 434 | 53-76 | | 50-52 | < 50 | 353 |
|  | RAVi max | 21-56 | 57-62 | > 62 | 500 | 24-69 | | 70-78 | > 78 | 408 |
|  | RAEF | 38-72 | 33-37 | < 33 | 497 | 35-69 | | 31-34 | < 31 | 410 |
| 50-59 | LVEDVi | 52-94 | 95-101 | > 101 | 911 | 53-104 | | 105-111 | > 111 | 812 |
|  | LVESVi | 14-37 | 38-41 | > 41 | 910 | 16-44 | | 45-48 | > 48 | 818 |
|  | LVSVi | 32-61 | 28-31 | < 28 | 910 | 31-65 | | 26-30 | < 26 | 818 |
|  | LVEF | 53-74 | 49-52 | < 49 | 906 | 50-73 | | 47-49 | < 47 | 831 |
|  | LVCOi | 2-4.1 | 1.7-1.9 | < 1.7 | 906 | 2-4.3 | | 1.7-1.9 | < 1.7 | 815 |
|  | LVMi | 33-54 | 55-59 | > 59 | 911 | 40-70 | | 71-75 | > 75 | 832 |
|  | LVM/LVEDV | 0.5-0.8 | 0.9-0.9 | > 0.9 | 911 | 0.6-0.9 | | 1-1.1 | > 1.1 | 809 |
|  | RVEDVi | 52-98 | 99-106 | > 106 | 909 | 56-113 | | 114-121 | > 121 | 818 |
|  | RVESVi | 15-41 | 42-45 | > 45 | 909 | 20-51 | | 52-55 | > 55 | 818 |
|  | RVEF | 52-72 | 49-51 | < 49 | 906 | 47-69 | | 43-46 | < 43 | 830 |
|  | RVCOi | 1.9-4.2 | 1.7-1.8 | < 1.7 | 906 | 1.9-4.4 | | 1.6-1.8 | < 1.6 | 815 |
|  | RVSVi | 31-62 | 27-30 | < 27 | 909 | 31-68 | | 25-30 | < 25 | 820 |
|  | LAVi max | 22-53 | 54-59 | > 59 | 797 | 20-55 | | 56-61 | > 61 | 684 |
|  | LAEF | 53-79 | 50-52 | < 50 | 791 | 53-79 | | 50-52 | < 50 | 681 |
|  | RAVi max | 21-60 | 61-68 | > 68 | 854 | 22-67 | | 68-75 | > 75 | 761 |
|  | RAEF | 38-70 | 33-37 | < 33 | 844 | 35-68 | | 31-34 | < 31 | 771 |
| 60-69 | LVEDVi | 49-90 | 91-98 | > 98 | 918 | 51-102 | | 103-111 | > 111 | 797 |
|  | LVESVi | 13-36 | 37-40 | > 40 | 919 | 15-43 | | 44-47 | > 47 | 791 |
|  | LVSVi | 30-59 | 27-29 | < 27 | 918 | 30-65 | | 25-29 | < 25 | 798 |
|  | LVEF | 54-75 | 50-53 | < 50 | 916 | 50-73 | | 47-49 | < 47 | 798 |
|  | LVCOi | 1.9-4 | 1.7-1.8 | < 1.7 | 917 | 1.9-4.1 | | 1.6-1.8 | < 1.6 | 792 |
|  | LVMi | 32-54 | 55-59 | > 59 | 919 | 39-69 | | 70-74 | > 74 | 801 |
|  | LVM/LVEDV | 0.5-0.8 | 0.9-0.9 | > 0.9 | 918 | 0.6-1 | | 1.1-1.1 | > 1.1 | 794 |
|  | RVEDVi | 49-94 | 95-102 | > 102 | 917 | 54-110 | | 111-119 | > 119 | 798 |
|  | RVESVi | 14-38 | 39-42 | > 42 | 920 | 19-48 | | 49-53 | > 53 | 801 |
|  | RVEF | 52-73 | 49-51 | < 49 | 914 | 48-70 | | 44-47 | < 44 | 803 |
|  | RVCOi | 1.8-4.1 | 1.6-1.7 | < 1.6 | 917 | 1.9-4.3 | | 1.6-1.8 | < 1.6 | 797 |
|  | RVSVi | 30-60 | 26-29 | < 26 | 917 | 30-67 | | 24-29 | < 24 | 792 |
|  | LAVi max | 20-53 | 54-60 | > 60 | 797 | 19-55 | | 56-62 | > 62 | 645 |
|  | LAEF | 51-79 | 47-50 | < 47 | 776 | 51-80 | | 47-50 | < 47 | 632 |
|  | RAVi max | 21-62 | 63-71 | > 71 | 855 | 21-69 | | 70-78 | > 78 | 742 |
|  | RAEF | 38-71 | 33-37 | < 33 | 841 | 35-70 | | 31-34 | < 31 | 748 |
| 70+ | LVEDVi | 47-89 | 90-98 | > 98 | 1101 | 48-101 | | 102-110 | > 110 | 1253 |
|  | LVESVi | 13-35 | 36-39 | > 39 | 1100 | 14-42 | | 43-47 | > 47 | 1247 |
|  | LVSVi | 30-58 | 26-29 | < 26 | 1102 | 29-63 | | 24-28 | < 24 | 1256 |
|  | LVEF | 54-76 | 51-53 | < 51 | 1096 | 51-74 | | 47-50 | < 47 | 1242 |
|  | LVCOi | 1.9-4 | 1.7-1.8 | < 1.7 | 1099 | 1.9-4 | | 1.6-1.8 | < 1.6 | 1254 |
|  | LVMi | 32-55 | 56-59 | > 59 | 1102 | 38-67 | | 68-72 | > 72 | 1257 |
|  | LVM/LVEDV | 0.5-0.9 | 01-1 | > 1 | 1101 | 0.6-1 | | 1.1-1.1 | > 1.1 | 1251 |
|  | RVEDVi | 48-92 | 93-101 | > 101 | 1100 | 52-108 | | 109-117 | > 117 | 1258 |
|  | RVESVi | 13-37 | 38-40 | > 40 | 1100 | 18-47 | | 48-53 | > 53 | 1259 |
|  | RVEF | 53-74 | 50-52 | < 50 | 1093 | 49-70 | | 45-48 | < 45 | 1246 |
|  | RVCOi | 1.8-4.1 | 1.6-1.7 | < 1.6 | 1099 | 1.8-4.2 | | 1.5-1.7 | < 1.5 | 1251 |
|  | RVSVi | 29-59 | 25-28 | < 25 | 1100 | 28-66 | | 23-27 | < 23 | 1256 |
|  | LAVi max | 19-56 | 57-63 | > 63 | 942 | 18-56 | | 57-64 | > 64 | 953 |
|  | LAEF | 48-79 | 44-47 | < 44 | 903 | 48-80 | | 44-47 | < 44 | 914 |
|  | RAVi max | 20-65 | 66-76 | > 76 | 1028 | 19-70 | | 71-79 | > 79 | 1119 |
|  | RAEF | 38-72 | 33-37 | < 33 | 1001 | 35-73 | | 30-34 | < 30 | 1115 |

Footnote: All measurements were derived using cvi42 software with smooth endocardial segmentation; papillary muscles and trabeculations were excluded from left ventricular mass measurements. Volumetric and mass measurements were indexed to bsa. Reference ranges are defined as the interval between the 2.5th and 97.5th centiles derived from age-specific generalized additive models for location, scale and shape (GAMLSS) using the Box–Cox Cole–Green distribution. Centile-based categories beyond the reference range are derived from LMS-based thresholds (P0.5, P2.5, P97.5, and P99.5) and describe the position of a measurement within the reference distribution rather than disease severity or diagnostic abnormality. Reference ranges are based on a White adult population.

Abbreviations: LV = left ventricle; RV = right ventricle; LA = left atrium; RA = right atrium; EDV = end-diastolic volume; ESV = end-systolic volume; SV = stroke volume; EF = ejection fraction; CO = cardiac output.

### 27. Table: Reference ranges and centile-based categories for cardiac chamber measurements derived using cvi42 software (smooth segmentation), indexed to BSA, in a Black adult population

|  |  | **Female** | | | | | **Male** | | | |
| --- | --- | --- | --- | --- | --- | --- | --- | --- | --- | --- |
| **Age group** | **Metrics** | **Reference range** | **Mild** | **At least moderate** | **n** | **Reference range** | | **Mild** | **At least moderate** | **n** |
| 40-49 | LVEDVi | 54-91 | 92-96 | > 96 | 10 |  | |  |  |  |
|  | LVESVi | 16-39 | 40-42 | > 42 | 10 |  | |  |  |  |
|  | LVSVi | 33-59 | 29-32 | < 29 | 10 |  | |  |  |  |
|  | LVEF | 52-75 | 50-51 | < 50 | 10 |  | |  |  |  |
|  | LVCOi | 1.9-4.1 | 1.7-1.8 | < 1.7 | 10 |  | |  |  |  |
|  | LVMi | 36-60 | 61-69 | > 69 | 10 |  | |  |  |  |
|  | LVM/LVEDV | 0.5-0.8 | 0.9-1 | > 1 | 10 |  | |  |  |  |
|  | RVEDVi | 53-99 | 100-106 | > 106 | 10 |  | |  |  |  |
|  | RVESVi | 17-44 | 45-49 | > 49 | 10 |  | |  |  |  |
|  | RVEF | 52-73 | 49-51 | < 49 | 10 |  | |  |  |  |
|  | RVCOi | 2-4.2 | 1.8-1.9 | < 1.8 | 10 |  | |  |  |  |
|  | RVSVi | 33-60 | 28-32 | < 28 | 10 |  | |  |  |  |
|  | LAVi max |  |  |  |  |  | |  |  |  |
|  | LAEF |  |  |  |  |  | |  |  |  |
|  | RAVi max | 23-67 | 68-80 | > 80 | 10 |  | |  |  |  |
|  | RAEF | 30-65 | 25-29 | < 25 | 10 |  | |  |  |  |
| 50-59 | LVEDVi | 51-90 | 91-95 | > 95 | 95 | 49-104 | | 105-112 | > 112 | 77 |
|  | LVESVi | 15-38 | 39-41 | > 41 | 95 | 15-42 | | 43-46 | > 46 | 75 |
|  | LVSVi | 31-58 | 28-30 | < 28 | 95 | 30-63 | | 25-29 | < 25 | 75 |
|  | LVEF | 52-75 | 50-51 | < 50 | 95 | 51-73 | | 48-50 | < 48 | 76 |
|  | LVCOi | 1.9-4 | 1.7-1.8 | < 1.7 | 94 | 2.1-4.1 | | 1.9-2 | < 1.9 | 75 |
|  | LVMi | 35-59 | 60-65 | > 65 | 95 | 37-71 | | 72-75 | > 75 | 78 |
|  | LVM/LVEDV | 0.5-0.8 | 0.9-1 | > 1 | 95 | 0.5-0.9 | | 01-1 | > 1 | 77 |
|  | RVEDVi | 52-98 | 99-106 | > 106 | 95 | 64-109 | | 110-121 | > 121 | 75 |
|  | RVESVi | 16-43 | 44-48 | > 48 | 95 | 18-49 | | 50-53 | > 53 | 76 |
|  | RVEF | 51-73 | 48-50 | < 48 | 95 | 47-69 | | 44-46 | < 44 | 77 |
|  | RVCOi | 2-4.2 | 1.8-1.9 | < 1.8 | 94 | 1.9-4.3 | | 1.7-1.8 | < 1.7 | 77 |
|  | RVSVi | 32-61 | 28-31 | < 28 | 95 | 29-64 | | 22-28 | < 22 | 75 |
|  | LAVi max | 22-51 | 52-55 | > 55 | 89 | 20-53 | | 54-60 | > 60 | 64 |
|  | LAEF | 54-75 | 51-53 | < 51 | 88 | 53-76 | | 50-52 | < 50 | 64 |
|  | RAVi max | 22-63 | 64-74 | > 74 | 94 | 28-64 | | 65-70 | > 70 | 73 |
|  | RAEF | 31-65 | 26-30 | < 26 | 93 | 35-63 | | 32-34 | < 32 | 71 |
| 60-69 | LVEDVi | 47-88 | 89-95 | > 95 | 66 | 53-107 | | 108-117 | > 117 | 48 |
|  | LVESVi | 14-37 | 38-41 | > 41 | 66 | 17-47 | | 48-54 | > 54 | 48 |
|  | LVSVi | 29-58 | 25-28 | < 25 | 66 | 32-65 | | 28-31 | < 28 | 48 |
|  | LVEF | 52-75 | 48-51 | < 48 | 66 | 45-73 | | 36-44 | < 36 | 48 |
|  | LVCOi | 1.8-3.9 | 1.6-1.7 | < 1.6 | 66 | 2-4.3 | | 1.8-1.9 | < 1.8 | 48 |
|  | LVMi | 32-58 | 59-62 | > 62 | 66 | 42-73 | | 74-80 | > 80 | 48 |
|  | LVM/LVEDV | 0.5-0.9 | 01-1 | > 1 | 66 | 0.6-1 | | 1.1-1.1 | > 1.1 | 48 |
|  | RVEDVi | 50-97 | 98-106 | > 106 | 66 | 62-113 | | 114-125 | > 125 | 48 |
|  | RVESVi | 14-41 | 42-46 | > 46 | 66 | 16-49 | | 50-53 | > 53 | 48 |
|  | RVEF | 50-73 | 46-49 | < 46 | 65 | 48-71 | | 44-47 | < 44 | 48 |
|  | RVCOi | 2-4.1 | 1.8-1.9 | < 1.8 | 66 | 2.1-4.4 | | 1.9-2 | < 1.9 | 48 |
|  | RVSVi | 31-62 | 28-30 | < 28 | 66 | 34-67 | | 30-33 | < 30 | 48 |
|  | LAVi max | 20-55 | 56-61 | > 61 | 59 | 20-56 | | 57-64 | > 64 | 42 |
|  | LAEF | 50-75 | 45-49 | < 45 | 58 | 50-75 | | 46-49 | < 46 | 43 |
|  | RAVi max | 21-59 | 60-67 | > 67 | 64 | 24-69 | | 70-76 | > 76 | 45 |
|  | RAEF | 32-67 | 27-31 | < 27 | 66 | 32-64 | | 29-31 | < 29 | 46 |
| 70+ | LVEDVi | 43-87 | 88-96 | > 96 | 18 | 49-98 | | 99-109 | > 109 | 14 |
|  | LVESVi | 13-36 | 37-42 | > 42 | 18 | 15-47 | | 48-59 | > 59 | 14 |
|  | LVSVi | 26-57 | 23-25 | < 23 | 18 | 31-63 | | 28-30 | < 28 | 14 |
|  | LVEF | 51-75 | 44-50 | < 44 | 18 | 43-74 | | 33-42 | < 33 | 14 |
|  | LVCOi | 1.8-3.8 | 1.6-1.7 | < 1.6 | 18 | 1.9-4.6 | |  | < 1.8 | 14 |
|  | LVMi | 26-58 | 59-61 | > 61 | 18 | 41-72 | | 73-85 | > 85 | 14 |
|  | LVM/LVEDV | 0.6-0.9 | 01-1 | > 1 | 18 | 0.6-1.1 | | 1.2-1.3 | > 1.3 | 14 |
|  | RVEDVi | 49-97 | 98-106 | > 106 | 18 | 56-108 | | 109-120 | > 120 | 14 |
|  | RVESVi | 13-40 | 41-44 | > 44 | 18 | 14-49 | | 50-53 | > 53 | 14 |
|  | RVEF | 48-73 | 42-47 | < 42 | 18 | 48-74 | | 44-47 | < 44 | 14 |
|  | RVCOi | 2-4.1 | 1.8-1.9 | < 1.8 | 18 | 2.2-4.5 | | 2-2.1 | < 2 | 14 |
|  | RVSVi | 30-63 | 27-29 | < 27 | 18 | 35-64 | | 32-34 | < 32 | 14 |
|  | LAVi max | 19-61 | 62-73 | > 73 | 15 |  | |  |  |  |
|  | LAEF | 44-74 | 35-43 | < 35 | 16 |  | |  |  |  |
|  | RAVi max | 20-57 | 58-63 | > 63 | 17 | 19-74 | | 75-82 | > 82 | 12 |
|  | RAEF | 33-68 | 28-32 | < 28 | 17 | 29-66 | | 25-28 | < 25 | 12 |

Footnote: All measurements were derived using cvi42 software with smooth endocardial segmentation; papillary muscles and trabeculations were excluded from left ventricular mass measurements. Volumetric and mass measurements were indexed to bsa. Reference ranges are defined as the interval between the 2.5th and 97.5th centiles derived from age-specific generalized additive models for location, scale and shape (GAMLSS) using the Box–Cox Cole–Green distribution. Centile-based categories beyond the reference range are derived from LMS-based thresholds (P0.5, P2.5, P97.5, and P99.5) and describe the position of a measurement within the reference distribution rather than disease severity or diagnostic abnormality. Reference ranges are based on a Black adult population.

Abbreviations: LV = left ventricle; RV = right ventricle; LA = left atrium; RA = right atrium; EDV = end-diastolic volume; ESV = end-systolic volume; SV = stroke volume; EF = ejection fraction; CO = cardiac output.

### 28. Table: Reference ranges and centile-based categories for cardiac chamber measurements derived using cvi42 software (smooth segmentation), indexed to BSA, in an Asian adult population

|  |  | **Female** | | | | **Male** | | | |
| --- | --- | --- | --- | --- | --- | --- | --- | --- | --- |
| **Age group** | **Metrics** | **Reference range** | **Mild** | **At least moderate** | **n** | **Reference range** | **Mild** | **At least moderate** | **n** |
| 40-49 | LVEDVi | 53-83 | 84-90 | > 90 | 10 | 58-97 | 98-106 | > 106 | 20 |
|  | LVESVi | 18-32 | 33-36 | > 36 | 10 | 19-43 | 44-48 | > 48 | 20 |
|  | LVSVi | 31-53 | 28-30 | < 28 | 10 | 35-56 | 32-34 | < 32 | 20 |
|  | LVEF | 53-72 | 49-52 | < 49 | 10 | 50-70 | 47-49 | < 47 | 20 |
|  | LVCOi | 2-3.7 | 1.8-1.9 | < 1.8 | 10 | 2-4.1 | 1.8-1.9 | < 1.8 | 20 |
|  | LVMi | 31-45 | 46-49 | > 49 | 10 | 37-58 | 59-62 | > 62 | 20 |
|  | LVM/LVEDV | 0.5-0.7 | 0.8-0.8 | > 0.8 | 10 | 0.5-0.8 | 0.9-0.9 | > 0.9 | 20 |
|  | RVEDVi | 51-83 | 84-91 | > 91 | 10 | 60-106 | 107-118 | > 118 | 20 |
|  | RVESVi | 16-35 | 36-40 | > 40 | 10 | 21-46 | 47-50 | > 50 | 20 |
|  | RVEF | 52-72 | 48-51 | < 48 | 10 | 49-67 | 46-48 | < 46 | 20 |
|  | RVCOi | 2-3.5 | 1.9-1.9 | < 1.9 | 10 | 2-3.9 | 1.8-1.9 | < 1.8 | 20 |
|  | RVSVi | 30-53 | 28-29 | < 28 | 10 | 34-65 | 31-33 | < 31 | 20 |
|  | LAVi max | 21-45 | 46-49 | > 49 | 10 | 18-48 | 49-53 | > 53 | 20 |
|  | LAEF | 57-81 | 54-56 | < 54 | 10 | 58-84 | 55-57 | < 55 | 20 |
|  | RAVi max | 21-44 | 45-48 | > 48 | 10 | 21-62 | 63-71 | > 71 | 20 |
|  | RAEF | 37-71 | 35-36 | < 35 | 10 | 32-63 | 27-31 | < 27 | 20 |
| 50-59 | LVEDVi | 51-84 | 85-92 | > 92 | 83 | 55-96 | 97-106 | > 106 | 133 |
|  | LVESVi | 16-35 | 36-41 | > 41 | 83 | 17-43 | 44-48 | > 48 | 133 |
|  | LVSVi | 30-54 | 27-29 | < 27 | 83 | 31-60 | 27-30 | < 27 | 133 |
|  | LVEF | 53-73 | 49-52 | < 49 | 82 | 50-71 | 47-49 | < 47 | 130 |
|  | LVCOi | 1.9-3.7 | 1.7-1.8 | < 1.7 | 83 | 1.9-4 | 1.7-1.8 | < 1.7 | 133 |
|  | LVMi | 30-47 | 48-52 | > 52 | 83 | 35-60 | 61-64 | > 64 | 133 |
|  | LVM/LVEDV | 0.5-0.7 | 0.8-0.8 | > 0.8 | 83 | 0.5-0.8 | 0.9-0.9 | > 0.9 | 133 |
|  | RVEDVi | 50-86 | 87-95 | > 95 | 83 | 56-104 | 105-115 | > 115 | 133 |
|  | RVESVi | 16-36 | 37-41 | > 41 | 83 | 20-46 | 47-51 | > 51 | 133 |
|  | RVEF | 52-73 | 49-51 | < 49 | 83 | 49-68 | 45-48 | < 45 | 133 |
|  | RVCOi | 1.9-3.7 |  | < 1.8 | 83 | 1.9-3.9 | 1.7-1.8 | < 1.7 | 133 |
|  | RVSVi | 30-55 | 27-29 | < 27 | 83 | 31-62 | 28-30 | < 28 | 133 |
|  | LAVi max | 21-47 | 48-51 | > 51 | 77 | 19-50 | 51-55 | > 55 | 121 |
|  | LAEF | 54-80 | 51-53 | < 51 | 75 | 55-82 | 52-54 | < 52 | 120 |
|  | RAVi max | 21-50 | 51-55 | > 55 | 76 | 21-62 | 63-71 | > 71 | 128 |
|  | RAEF | 37-69 | 34-36 | < 34 | 76 | 34-65 | 29-33 | < 29 | 127 |
| 60-69 | LVEDVi | 48-86 | 87-97 | > 97 | 63 | 48-95 | 96-104 | > 104 | 118 |
|  | LVESVi | 15-38 | 39-46 | > 46 | 63 | 16-42 | 43-48 | > 48 | 118 |
|  | LVSVi | 29-56 | 27-28 | < 27 | 63 | 30-59 | 26-29 | < 26 | 118 |
|  | LVEF | 53-75 | 50-52 | < 50 | 63 | 50-73 | 47-49 | < 47 | 118 |
|  | LVCOi | 1.8-3.8 | 1.6-1.7 | < 1.6 | 63 | 1.8-3.9 | 1.5-1.7 | < 1.5 | 119 |
|  | LVMi | 30-52 | 53-58 | > 58 | 63 | 33-62 | 63-68 | > 68 | 119 |
|  | LVM/LVEDV | 0.5-0.8 |  | > 0.8 | 63 | 0.5-0.9 | 01-1 | > 1 | 118 |
|  | RVEDVi | 48-92 | 93-104 | > 104 | 63 | 50-101 | 102-110 | > 110 | 119 |
|  | RVESVi | 15-37 | 38-42 | > 42 | 63 | 18-46 | 47-51 | > 51 | 119 |
|  | RVEF | 54-73 | 51-53 | < 51 | 63 | 47-70 | 42-46 | < 42 | 119 |
|  | RVCOi | 1.8-3.9 | 1.6-1.7 | < 1.6 | 63 | 1.7-3.9 | 1.4-1.6 | < 1.4 | 119 |
|  | RVSVi | 29-59 | 26-28 | < 26 | 63 | 28-60 | 24-27 | < 24 | 119 |
|  | LAVi max | 21-50 | 51-55 | > 55 | 59 | 20-52 | 53-57 | > 57 | 110 |
|  | LAEF | 50-78 | 46-49 | < 46 | 58 | 51-80 | 48-50 | < 48 | 107 |
|  | RAVi max | 19-60 | 61-67 | > 67 | 61 | 21-62 | 63-71 | > 71 | 113 |
|  | RAEF | 36-68 | 31-35 | < 31 | 59 | 35-70 | 31-34 | < 31 | 114 |
| 70+ | LVEDVi | 45-90 | 91-103 | > 103 | 28 | 40-93 | 94-102 | > 102 | 41 |
|  | LVESVi | 14-36 | 37-43 | > 43 | 28 | 14-42 | 43-49 | > 49 | 41 |
|  | LVSVi | 29-61 | 26-28 | < 26 | 28 | 28-59 | 24-27 | < 24 | 40 |
|  | LVEF | 54-77 | 51-53 | < 51 | 28 | 51-75 | 47-50 | < 47 | 41 |
|  | LVCOi | 1.6-4 | 1.4-1.5 | < 1.4 | 28 | 1.4-4 | 1-1.3 | < 1 | 41 |
|  | LVMi | 29-58 | 59-68 | > 68 | 28 | 30-65 | 66-73 | > 73 | 41 |
|  | LVM/LVEDV | 0.5-0.8 |  | > 0.8 | 28 | 0.5-1 | 1.1-1.2 | > 1.2 | 41 |
|  | RVEDVi | 46-99 | 100-115 | > 115 | 28 | 42-98 | 99-106 | > 106 | 41 |
|  | RVESVi | 14-38 | 39-44 | > 44 | 28 | 16-46 | 47-53 | > 53 | 41 |
|  | RVEF | 55-74 | 52-54 | < 52 | 28 | 46-72 | 39-45 | < 39 | 40 |
|  | RVCOi | 1.6-4.1 | 1.4-1.5 | < 1.4 | 28 | 1.3-4 | 0.7-1.2 | < 0.7 | 41 |
|  | RVSVi | 28-64 | 26-27 | < 26 | 28 | 26-60 | 19-25 | < 19 | 40 |
|  | LAVi max | 21-54 | 55-61 | > 61 | 28 | 21-53 | 54-59 | > 59 | 34 |
|  | LAEF | 45-76 | 40-44 | < 40 | 27 | 48-79 | 45-47 | < 45 | 34 |
|  | RAVi max | 15-66 | 67-77 | > 77 | 27 | 21-62 | 63-71 | > 71 | 38 |
|  | RAEF | 35-70 | 25-34 | < 25 | 27 | 36-77 | 32-35 | < 32 | 39 |

Footnote: All measurements were derived using cvi42 software with smooth endocardial segmentation; papillary muscles and trabeculations were excluded from left ventricular mass measurements. Volumetric and mass measurements were indexed to bsa. Reference ranges are defined as the interval between the 2.5th and 97.5th centiles derived from age-specific generalized additive models for location, scale and shape (GAMLSS) using the Box–Cox Cole–Green distribution. Centile-based categories beyond the reference range are derived from LMS-based thresholds (P0.5, P2.5, P97.5, and P99.5) and describe the position of a measurement within the reference distribution rather than disease severity or diagnostic abnormality. Reference ranges are based on an Asian adult population.

Abbreviations: LV = left ventricle; RV = right ventricle; LA = left atrium; RA = right atrium; EDV = end-diastolic volume; ESV = end-systolic volume; SV = stroke volume; EF = ejection fraction; CO = cardiac output.

### 29. Table: Reference ranges and centile-based categories for cardiac chamber measurements derived using cvi42 software (smooth segmentation), indexed to BSA, in a Chinese adult population

|  |  | **Female** | | | | **Male** | | | |
| --- | --- | --- | --- | --- | --- | --- | --- | --- | --- |
| **Age group** | **Metrics** | **Reference range** | **Mild** | **At least moderate** | **n** | **Reference range** | **Mild** | **At least moderate** | **n** |
| 18-29 | LVEDVi | 60-100 | 101-110 | > 110 | 16 | 67-100 | 101-105 | > 105 | 19 |
|  | LVESVi | 20-44 | 45-53 | > 53 | 16 | 24-44 | 45-47 | > 47 | 19 |
|  | LVSVi | 36-68 | 34-35 | < 34 | 16 | 36-60 | 30-35 | < 30 | 19 |
|  | LVEF | 52-73 | 48-51 | < 48 | 16 | 50-67 | 48-49 | < 48 | 19 |
|  | LVCOi | 2.2-4.6 | 2-2.1 | < 2 | 16 | 2.7-4.4 | 2.6-2.6 | < 2.6 | 19 |
|  | LVMi | 29-48 | 49-54 | > 54 | 16 | 38-59 | 60-62 | > 62 | 19 |
|  | LVM/LVEDV | 0.4-0.6 |  | > 0.6 | 16 | 0.5-0.7 | 0.8-0.8 | > 0.8 | 19 |
|  | RVEDVi | 58-105 | 106-115 | > 115 | 16 | 75-115 | 116-123 | > 123 | 19 |
|  | RVESVi | 18-49 | 50-54 | > 54 | 16 | 27-61 | 62-67 | > 67 | 19 |
|  | RVEF | 47-71 | 44-46 | < 44 | 16 | 40-62 | 35-39 | < 35 | 19 |
|  | RVCOi | 1.9-4.5 | 1.7-1.8 | < 1.7 | 16 | 2.5-4.5 | 2.3-2.4 | < 2.3 | 19 |
|  | RVSVi | 32-63 | 29-31 | < 29 | 16 | 39-62 | 36-38 | < 36 | 19 |
|  | LAVi max | 23-53 | 54-59 | > 59 | 13 | 25-47 | 48-51 | > 51 | 16 |
|  | LAEF | 54-77 | 50-53 | < 50 | 13 | 52-70 | 49-51 | < 49 | 16 |
|  | RAVi max | 21-45 | 46-53 | > 53 | 15 | 18-58 | 59-65 | > 65 | 18 |
|  | RAEF | 41-73 | 35-40 | < 35 | 15 | 36-68 | 30-35 | < 30 | 18 |
| 30-39 | LVEDVi | 58-97 | 98-106 | > 106 | 18 | 64-99 | 100-105 | > 105 | 17 |
|  | LVESVi | 19-42 | 43-50 | > 50 | 18 | 21-43 | 44-46 | > 46 | 17 |
|  | LVSVi | 36-64 | 33-35 | < 33 | 18 | 35-60 | 30-34 | < 30 | 17 |
|  | LVEF | 52-73 | 48-51 | < 48 | 18 | 50-68 | 48-49 | < 48 | 17 |
|  | LVCOi | 2.1-4.5 | 02-2 | < 2 | 18 | 2.5-4.4 | 2.4-2.4 | < 2.4 | 17 |
|  | LVMi | 30-49 | 50-55 | > 55 | 18 | 38-60 | 61-63 | > 63 | 17 |
|  | LVM/LVEDV | 0.4-0.6 |  | > 0.6 | 18 | 0.5-0.8 |  | > 0.8 | 17 |
|  | RVEDVi | 56-101 | 102-111 | > 111 | 18 | 71-113 | 114-122 | > 122 | 17 |
|  | RVESVi | 17-46 | 47-51 | > 51 | 18 | 24-58 | 59-63 | > 63 | 17 |
|  | RVEF | 48-71 | 45-47 | < 45 | 18 | 42-64 | 38-41 | < 38 | 17 |
|  | RVCOi | 2-4.3 | 1.8-1.9 | < 1.8 | 18 | 2.3-4.4 | 2.1-2.2 | < 2.1 | 17 |
|  | RVSVi | 32-62 | 29-31 | < 29 | 18 | 36-62 | 33-35 | < 33 | 17 |
|  | LAVi max | 23-52 | 53-58 | > 58 | 18 | 24-47 | 48-51 | > 51 | 14 |
|  | LAEF | 53-76 | 50-52 | < 50 | 18 | 51-72 | 48-50 | < 48 | 14 |
|  | RAVi max | 22-49 | 50-57 | > 57 | 18 | 18-59 | 60-67 | > 67 | 16 |
|  | RAEF | 41-72 | 35-40 | < 35 | 18 | 36-69 | 31-35 | < 31 | 16 |
| 40-49 | LVEDVi | 56-93 | 94-102 | > 102 | 24 | 60-98 | 99-104 | > 104 | 20 |
|  | LVESVi | 18-39 | 40-46 | > 46 | 24 | 19-42 | 43-45 | > 45 | 20 |
|  | LVSVi | 35-61 | 32-34 | < 32 | 24 | 33-59 | 29-32 | < 29 | 20 |
|  | LVEF | 52-74 | 48-51 | < 48 | 24 | 50-69 | 47-49 | < 47 | 19 |
|  | LVCOi | 2.1-4.4 | 1.9-2 | < 1.9 | 24 | 2.3-4.3 |  | < 2.2 | 20 |
|  | LVMi | 31-51 | 52-56 | > 56 | 24 | 38-62 | 63-66 | > 66 | 20 |
|  | LVM/LVEDV | 0.4-0.7 |  | > 0.7 | 24 | 0.5-0.8 | 0.9-0.9 | > 0.9 | 20 |
|  | RVEDVi | 54-97 | 98-106 | > 106 | 24 | 67-110 | 111-120 | > 120 | 20 |
|  | RVESVi | 16-43 | 44-48 | > 48 | 24 | 22-54 | 55-59 | > 59 | 20 |
|  | RVEF | 50-72 | 46-49 | < 46 | 23 | 44-66 | 41-43 | < 41 | 19 |
|  | RVCOi | 2-4.1 | 1.8-1.9 | < 1.8 | 24 | 2.1-4.4 | 1.9-2 | < 1.9 | 20 |
|  | RVSVi | 32-60 | 29-31 | < 29 | 24 | 34-62 | 30-33 | < 30 | 20 |
|  | LAVi max | 22-51 | 52-57 | > 57 | 22 | 22-47 | 48-51 | > 51 | 18 |
|  | LAEF | 53-75 | 49-52 | < 49 | 22 | 51-74 | 47-50 | < 47 | 18 |
|  | RAVi max | 22-53 | 54-63 | > 63 | 23 | 19-61 | 62-69 | > 69 | 19 |
|  | RAEF | 41-70 | 36-40 | < 36 | 23 | 35-69 | 31-34 | < 31 | 19 |
| 50-59 | LVEDVi | 54-90 | 91-99 | > 99 | 66 | 56-97 | 98-104 | > 104 | 49 |
|  | LVESVi | 16-37 | 38-43 | > 43 | 66 | 16-42 | 43-45 | > 45 | 49 |
|  | LVSVi | 34-59 | 31-33 | < 31 | 66 | 32-60 | 28-31 | < 28 | 49 |
|  | LVEF | 53-75 | 48-52 | < 48 | 66 | 50-70 | 47-49 | < 47 | 48 |
|  | LVCOi | 2.1-4 | 1.9-2 | < 1.9 | 66 | 2.1-4.3 | 1.9-2 | < 1.9 | 49 |
|  | LVMi | 32-52 | 53-58 | > 58 | 66 | 38-64 | 65-70 | > 70 | 49 |
|  | LVM/LVEDV | 0.5-0.7 | 0.8-0.8 | > 0.8 | 66 | 0.5-0.8 | 0.9-0.9 | > 0.9 | 49 |
|  | RVEDVi | 52-93 | 94-102 | > 102 | 66 | 63-108 | 109-118 | > 118 | 49 |
|  | RVESVi | 16-40 | 41-45 | > 45 | 66 | 19-51 | 52-55 | > 55 | 49 |
|  | RVEF | 51-73 | 47-50 | < 47 | 66 | 46-68 | 43-45 | < 43 | 49 |
|  | RVCOi | 2-3.9 | 1.8-1.9 | < 1.8 | 66 | 2-4.3 | 1.7-1.9 | < 1.7 | 49 |
|  | RVSVi | 32-59 | 29-31 | < 29 | 66 | 31-62 | 26-30 | < 26 | 49 |
|  | LAVi max | 22-50 | 51-56 | > 56 | 60 | 21-46 | 47-50 | > 50 | 40 |
|  | LAEF | 51-76 | 48-50 | < 48 | 58 | 50-77 | 46-49 | < 46 | 40 |
|  | RAVi max | 23-58 | 59-69 | > 69 | 63 | 20-63 | 64-71 | > 71 | 46 |
|  | RAEF | 40-68 | 36-39 | < 36 | 63 | 35-71 | 31-34 | < 31 | 46 |
| 60-69 | LVEDVi | 52-87 | 88-95 | > 95 | 52 | 53-97 | 98-105 | > 105 | 37 |
|  | LVESVi | 15-35 | 36-40 | > 40 | 52 | 13-41 | 42-44 | > 44 | 37 |
|  | LVSVi | 32-58 | 29-31 | < 29 | 52 | 31-60 | 27-30 | < 27 | 37 |
|  | LVEF | 53-75 | 48-52 | < 48 | 52 | 50-71 | 47-49 | < 47 | 35 |
|  | LVCOi | 2.1-3.7 | 02-2 | < 2 | 52 | 1.9-4.2 | 1.6-1.8 | < 1.6 | 37 |
|  | LVMi | 33-53 | 54-60 | > 60 | 52 | 37-67 | 68-78 | > 78 | 37 |
|  | LVM/LVEDV | 0.5-0.8 | 0.9-1 | > 1 | 52 | 0.5-0.9 | 01-1 | > 1 | 37 |
|  | RVEDVi | 50-89 | 90-97 | > 97 | 52 | 58-105 | 106-116 | > 116 | 36 |
|  | RVESVi | 15-37 | 38-41 | > 41 | 52 | 16-47 | 48-51 | > 51 | 37 |
|  | RVEF | 53-74 | 49-52 | < 49 | 52 | 47-71 | 45-46 | < 45 | 36 |
|  | RVCOi | 2-3.8 | 1.8-1.9 | < 1.8 | 52 | 1.8-4.3 | 1.5-1.7 | < 1.5 | 37 |
|  | RVSVi | 32-58 | 29-31 | < 29 | 52 | 27-63 | 22-26 | < 22 | 37 |
|  | LAVi max | 21-49 | 50-55 | > 55 | 47 | 19-46 | 47-50 | > 50 | 30 |
|  | LAEF | 49-76 | 45-48 | < 45 | 46 | 48-80 | 44-47 | < 44 | 30 |
|  | RAVi max | 23-63 | 64-76 | > 76 | 46 | 20-65 | 66-73 | > 73 | 35 |
|  | RAEF | 40-66 | 36-39 | < 36 | 46 | 35-72 | 32-34 | < 32 | 35 |
| 70+ | LVEDVi | 51-84 | 85-91 | > 91 | 12 |  |  |  |  |
|  | LVESVi | 14-33 | 34-38 | > 38 | 12 |  |  |  |  |
|  | LVSVi | 29-58 | 25-28 | < 25 | 12 |  |  |  |  |
|  | LVEF | 53-76 | 47-52 | < 47 | 12 |  |  |  |  |
|  | LVCOi | 2-3.8 | 1.8-1.9 | < 1.8 | 12 |  |  |  |  |
|  | LVMi | 34-54 | 55-61 | > 61 | 12 |  |  |  |  |
|  | LVM/LVEDV | 0.5-0.9 | 1-1.3 | > 1.3 | 12 |  |  |  |  |
|  | RVEDVi | 48-86 | 87-93 | > 93 | 12 |  |  |  |  |
|  | RVESVi | 14-34 | 35-38 | > 38 | 12 |  |  |  |  |
|  | RVEF | 54-74 | 50-53 | < 50 | 12 |  |  |  |  |
|  | RVCOi | 2-3.6 | 1.8-1.9 | < 1.8 | 12 |  |  |  |  |
|  | RVSVi | 31-58 | 28-30 | < 28 | 12 |  |  |  |  |
|  | LAVi max | 21-48 | 49-53 | > 53 | 11 |  |  |  |  |
|  | LAEF | 47-78 | 42-46 | < 42 | 12 |  |  |  |  |
|  | RAVi max | 23-68 | 69-82 | > 82 | 11 |  |  |  |  |
|  | RAEF | 39-65 | 36-38 | < 36 | 11 |  |  |  |  |

Footnote: All measurements were derived using cvi42 software with smooth endocardial segmentation; papillary muscles and trabeculations were excluded from left ventricular mass measurements. Volumetric and mass measurements were indexed to bsa. Reference ranges are defined as the interval between the 2.5th and 97.5th centiles derived from age-specific generalized additive models for location, scale and shape (GAMLSS) using the Box–Cox Cole–Green distribution. Centile-based categories beyond the reference range are derived from LMS-based thresholds (P0.5, P2.5, P97.5, and P99.5) and describe the position of a measurement within the reference distribution rather than disease severity or diagnostic abnormality. Reference ranges are based on a Chinese adult population.

Abbreviations: LV = left ventricle; RV = right ventricle; LA = left atrium; RA = right atrium; EDV = end-diastolic volume; ESV = end-systolic volume; SV = stroke volume; EF = ejection fraction; CO = cardiac output.

### 30. Table: Reference ranges and centile-based categories for cardiac chamber measurements derived using cvi42 software (smooth segmentation), indexed to BSA, in a Mixed adult population

|  |  | **Female** | | | | **Male** | | | |
| --- | --- | --- | --- | --- | --- | --- | --- | --- | --- |
| **Age group** | **Metrics** | **Reference range** | **Mild** | **At least moderate** | **n** | **Reference range** | **Mild** | **At least moderate** | **n** |
| 50-59 | LVEDVi | 54-94 | 95-103 | > 103 | 134 | 61-106 | 107-113 | > 113 | 65 |
|  | LVESVi | 16-38 | 39-42 | > 42 | 134 | 20-45 | 46-48 | > 48 | 66 |
|  | LVSVi | 32-63 | 29-31 | < 29 | 134 | 36-69 | 32-35 | < 32 | 65 |
|  | LVEF | 53-74 | 49-52 | < 49 | 134 | 50-71 | 46-49 | < 46 | 66 |
|  | LVCOi | 2-4.1 | 1.8-1.9 | < 1.8 | 132 | 2.1-4.6 | 1.9-2 | < 1.9 | 66 |
|  | LVMi | 31-53 | 54-58 | > 58 | 134 | 39-70 | 71-76 | > 76 | 65 |
|  | LVM/LVEDV | 0.5-0.7 | 0.8-0.8 | > 0.8 | 134 | 0.5-0.8 | 0.9-0.9 | > 0.9 | 64 |
|  | RVEDVi | 53-98 | 99-108 | > 108 | 134 | 62-121 | 122-134 | > 134 | 65 |
|  | RVESVi | 17-40 | 41-44 | > 44 | 134 | 22-49 | 50-53 | > 53 | 65 |
|  | RVEF | 53-72 | 50-52 | < 50 | 134 | 51-70 | 48-50 | < 48 | 66 |
|  | RVCOi | 1.9-4.1 | 1.7-1.8 | < 1.7 | 133 | 2.2-4.6 | 2-2.1 | < 2 | 66 |
|  | RVSVi | 32-61 | 28-31 | < 28 | 133 | 36-74 | 32-35 | < 32 | 66 |
|  | LAVi max | 21-48 | 49-51 | > 51 | 121 | 24-57 | 58-64 | > 64 | 53 |
|  | LAEF | 55-78 | 52-54 | < 52 | 120 | 54-78 | 51-53 | < 51 | 53 |
|  | RAVi max | 21-63 | 64-73 | > 73 | 128 | 21-68 | 69-76 | > 76 | 64 |
|  | RAEF | 37-64 | 31-36 | < 31 | 124 | 35-63 | 31-34 | < 31 | 64 |
| 60-69 | LVEDVi | 48-91 | 92-99 | > 99 | 110 | 53-101 | 102-108 | > 108 | 59 |
|  | LVESVi | 13-37 | 38-40 | > 40 | 110 | 15-45 | 46-50 | > 50 | 58 |
|  | LVSVi | 31-58 | 27-30 | < 27 | 110 | 33-63 | 29-32 | < 29 | 59 |
|  | LVEF | 52-74 | 48-51 | < 48 | 108 | 49-72 | 45-48 | < 45 | 58 |
|  | LVCOi | 1.9-4 |  | < 1.8 | 110 | 2-4.2 | 1.8-1.9 | < 1.8 | 59 |
|  | LVMi | 30-52 | 53-56 | > 56 | 110 | 38-69 | 70-75 | > 75 | 59 |
|  | LVM/LVEDV | 0.5-0.8 |  | > 0.8 | 110 | 0.5-0.9 | 01-1 | > 1 | 59 |
|  | RVEDVi | 47-94 | 95-101 | > 101 | 110 | 56-114 | 115-125 | > 125 | 58 |
|  | RVESVi | 16-38 | 39-43 | > 43 | 110 | 18-48 | 49-53 | > 53 | 58 |
|  | RVEF | 53-72 | 49-52 | < 49 | 110 | 49-69 | 47-48 | < 47 | 58 |
|  | RVCOi | 1.9-4.1 | 1.7-1.8 | < 1.7 | 110 | 2-4.4 | 1.8-1.9 | < 1.8 | 59 |
|  | RVSVi | 29-60 | 24-28 | < 24 | 110 | 32-69 | 27-31 | < 27 | 59 |
|  | LAVi max | 22-51 | 52-56 | > 56 | 100 | 21-56 | 57-63 | > 63 | 52 |
|  | LAEF | 52-79 | 49-51 | < 49 | 98 | 50-79 | 47-49 | < 47 | 52 |
|  | RAVi max | 20-62 | 63-70 | > 70 | 103 | 18-71 | 72-79 | > 79 | 57 |
|  | RAEF | 35-68 | 29-34 | < 29 | 103 | 36-68 | 33-35 | < 33 | 57 |
| 70+ | LVEDVi | 48-83 | 84-88 | > 88 | 51 | 47-96 | 97-103 | > 103 | 33 |
|  | LVESVi | 10-36 | 37-39 | > 39 | 51 | 15-43 | 44-47 | > 47 | 32 |
|  | LVSVi | 28-53 | 23-27 | < 23 | 50 | 30-58 | 26-29 | < 26 | 34 |
|  | LVEF | 51-74 | 47-50 | < 47 | 51 | 48-72 | 45-47 | < 45 | 34 |
|  | LVCOi | 1.9-3.9 | 1.7-1.8 | < 1.7 | 51 | 1.9-3.9 | 1.7-1.8 | < 1.7 | 35 |
|  | LVMi | 30-50 | 51-53 | > 53 | 51 | 37-67 | 68-74 | > 74 | 35 |
|  | LVM/LVEDV | 0.5-0.7 | 0.8-0.8 | > 0.8 | 51 | 0.5-1 | 1.1-1.1 | > 1.1 | 33 |
|  | RVEDVi | 52-81 | 82-85 | > 85 | 51 | 50-108 | 109-117 | > 117 | 35 |
|  | RVESVi | 17-35 | 36-39 | > 39 | 51 | 14-48 | 49-52 | > 52 | 35 |
|  | RVEF | 52-72 | 48-51 | < 48 | 51 | 48-68 | 46-47 | < 46 | 35 |
|  | RVCOi | 1.9-3.9 | 1.7-1.8 | < 1.7 | 51 | 1.8-4.2 | 1.6-1.7 | < 1.6 | 35 |
|  | RVSVi | 31-55 | 27-30 | < 27 | 50 | 28-64 | 21-27 | < 21 | 35 |
|  | LAVi max | 22-55 | 56-61 | > 61 | 45 | 18-55 | 56-63 | > 63 | 32 |
|  | LAEF | 50-80 | 47-49 | < 47 | 44 | 46-81 | 42-45 | < 42 | 30 |
|  | RAVi max | 19-61 | 62-68 | > 68 | 47 | 17-72 | 73-81 | > 81 | 31 |
|  | RAEF | 33-72 | 28-32 | < 28 | 46 | 36-76 | 34-35 | < 34 | 31 |

Footnote: All measurements were derived using cvi42 software with smooth endocardial segmentation; papillary muscles and trabeculations were excluded from left ventricular mass measurements. Volumetric and mass measurements were indexed to bsa. Reference ranges are defined as the interval between the 2.5th and 97.5th centiles derived from age-specific generalized additive models for location, scale and shape (GAMLSS) using the Box–Cox Cole–Green distribution. Centile-based categories beyond the reference range are derived from LMS-based thresholds (P0.5, P2.5, P97.5, and P99.5) and describe the position of a measurement within the reference distribution rather than disease severity or diagnostic abnormality. Reference ranges are based on a Mixed adult population.

Abbreviations: LV = left ventricle; RV = right ventricle; LA = left atrium; RA = right atrium; EDV = end-diastolic volume; ESV = end-systolic volume; SV = stroke volume; EF = ejection fraction; CO = cardiac output.

### 31. Table: Reference ranges and centile-based categories for cardiac chamber measurements derived using cvi42 software (anatomical segmentation), indexed to BSA, in a White adult population

|  |  | **Female** | | | | | **Male** | | | |
| --- | --- | --- | --- | --- | --- | --- | --- | --- | --- | --- |
| **Age group** | **Metrics** | **Reference range** | **Mild** | **At least moderate** | **n** | **Reference range** | | **Mild** | **At least moderate** | **n** |
| 18-29 | LVEDVi | 53-96 | 97-102 | > 102 | 102 | 62-112 | | 113-120 | > 120 | 103 |
|  | LVESVi | 14-38 | 39-41 | > 41 | 102 | 15-46 | | 47-49 | > 49 | 104 |
|  | LVSVi | 33-63 | 29-32 | < 29 | 102 | 37-73 | | 32-36 | < 32 | 101 |
|  | LVEF | 55-74 | 52-54 | < 52 | 100 | 51-72 | | 48-50 | < 48 | 102 |
|  | LVCOi | 2.3-4.7 | 2-2.2 | < 2 | 102 | 2.2-4.8 | | 1.7-2.1 | < 1.7 | 101 |
|  | LVMi | 40-62 | 63-66 | > 66 | 102 | 50-84 | | 85-89 | > 89 | 104 |
|  | LVM/LVEDV | 0.6-0.9 | 01-1 | > 1 | 102 | 0.6-1 | | 1.1-1.1 | > 1.1 | 102 |
|  | RVEDVi | 55-101 | 102-107 | > 107 | 102 | 63-125 | | 126-133 | > 133 | 102 |
|  | RVESVi | 13-51 | 52-57 | > 57 | 100 | 25-59 | | 60-64 | > 64 | 92 |
|  | RVEF | 47-71 | 44-46 | < 44 | 97 | 45-68 | | 41-44 | < 41 | 97 |
|  | RVCOi | 1.9-5 | 1.5-1.8 | < 1.5 | 101 | 2.2-4.9 | | 1.7-2.1 | < 1.7 | 98 |
|  | RVSVi | 29-67 | 24-28 | < 24 | 102 | 36-75 | | 31-35 | < 31 | 99 |
|  | LAVi max | 25-50 | 51-54 | > 54 | 50 | 27-58 | | 59-67 | > 67 | 52 |
|  | LAEF | 56-75 | 54-55 | < 54 | 50 | 53-72 | | 50-52 | < 50 | 52 |
|  | RAVi max | 21-49 | 50-54 | > 54 | 80 | 25-62 | | 63-70 | > 70 | 82 |
|  | RAEF | 38-78 | 33-37 | < 33 | 78 | 35-74 | | 31-34 | < 31 | 82 |
| 30-39 | LVEDVi | 51-92 | 93-99 | > 99 | 137 | 54-102 | | 103-109 | > 109 | 134 |
|  | LVESVi | 13-36 | 37-40 | > 40 | 136 | 13-41 | | 42-44 | > 44 | 134 |
|  | LVSVi | 32-61 | 28-31 | < 28 | 135 | 33-66 | | 28-32 | < 28 | 133 |
|  | LVEF | 55-75 | 53-54 | < 53 | 132 | 52-74 | | 48-51 | < 48 | 134 |
|  | LVCOi | 2.2-4.6 | 2-2.1 | < 2 | 136 | 2.1-4.6 | | 1.7-2 | < 1.7 | 128 |
|  | LVMi | 39-61 | 62-66 | > 66 | 137 | 45-77 | | 78-81 | > 81 | 134 |
|  | LVM/LVEDV | 0.6-0.9 | 1-1.1 | > 1.1 | 137 | 0.6-1 | | 1.1-1.2 | > 1.2 | 135 |
|  | RVEDVi | 54-99 | 100-107 | > 107 | 137 | 61-117 | | 118-125 | > 125 | 131 |
|  | RVESVi | 14-48 | 49-54 | > 54 | 135 | 24-56 | | 57-61 | > 61 | 129 |
|  | RVEF | 48-70 | 45-47 | < 45 | 127 | 45-68 | | 42-44 | < 42 | 131 |
|  | RVCOi | 2-4.8 | 1.7-1.9 | < 1.7 | 134 | 2.1-4.8 | | 1.7-2 | < 1.7 | 125 |
|  | RVSVi | 30-65 | 25-29 | < 25 | 135 | 33-71 | | 28-32 | < 28 | 132 |
|  | LAVi max | 25-51 | 52-56 | > 56 | 84 | 25-56 | | 57-64 | > 64 | 66 |
|  | LAEF | 55-76 | 53-54 | < 53 | 83 | 53-73 | | 50-52 | < 50 | 66 |
|  | RAVi max | 20-50 | 51-55 | > 55 | 113 | 23-61 | | 62-69 | > 69 | 119 |
|  | RAEF | 38-75 | 33-37 | < 33 | 110 | 35-72 | | 31-34 | < 31 | 118 |
| 40-49 | LVEDVi | 50-91 | 92-98 | > 98 | 518 | 53-103 | | 104-111 | > 111 | 438 |
|  | LVESVi | 12-34 | 35-38 | > 38 | 518 | 13-41 | | 42-45 | > 45 | 438 |
|  | LVSVi | 32-62 | 28-31 | < 28 | 518 | 33-68 | | 28-32 | < 28 | 435 |
|  | LVEF | 56-76 | 53-55 | < 53 | 512 | 53-75 | | 49-52 | < 49 | 434 |
|  | LVCOi | 2.1-4.4 | 1.9-2 | < 1.9 | 515 | 2.1-4.5 | | 1.7-2 | < 1.7 | 429 |
|  | LVMi | 38-61 | 62-66 | > 66 | 518 | 46-78 | | 79-83 | > 83 | 429 |
|  | LVM/LVEDV | 0.6-0.9 | 1-1.1 | > 1.1 | 518 | 0.6-1.1 | |  | > 1.2 | 426 |
|  | RVEDVi | 54-100 | 101-108 | > 108 | 517 | 61-119 | | 120-127 | > 127 | 430 |
|  | RVESVi | 16-46 | 47-50 | > 50 | 517 | 23-56 | | 57-61 | > 61 | 422 |
|  | RVEF | 49-70 | 46-48 | < 46 | 508 | 45-68 | | 42-44 | < 42 | 431 |
|  | RVCOi | 2-4.5 | 1.7-1.9 | < 1.7 | 514 | 2.1-4.8 | | 1.7-2 | < 1.7 | 426 |
|  | RVSVi | 31-64 | 26-30 | < 26 | 516 | 33-71 | | 27-32 | < 27 | 432 |
|  | LAVi max | 24-54 | 55-59 | > 59 | 440 | 23-55 | | 56-62 | > 62 | 358 |
|  | LAEF | 55-78 | 52-54 | < 52 | 434 | 53-76 | | 50-52 | < 50 | 353 |
|  | RAVi max | 21-56 | 57-62 | > 62 | 500 | 24-69 | | 70-78 | > 78 | 408 |
|  | RAEF | 38-72 | 33-37 | < 33 | 497 | 35-69 | | 31-34 | < 31 | 410 |
| 50-59 | LVEDVi | 48-87 | 88-95 | > 95 | 911 | 49-98 | | 99-107 | > 107 | 831 |
|  | LVESVi | 11-32 | 33-36 | > 36 | 911 | 13-39 | | 40-44 | > 44 | 835 |
|  | LVSVi | 31-60 | 28-30 | < 28 | 911 | 31-65 | | 26-30 | < 26 | 822 |
|  | LVEF | 57-78 | 53-56 | < 53 | 906 | 54-77 | | 50-53 | < 50 | 833 |
|  | LVCOi | 1.9-4.1 | 1.7-1.8 | < 1.7 | 908 | 2-4.2 | | 1.7-1.9 | < 1.7 | 816 |
|  | LVMi | 37-61 | 62-66 | > 66 | 910 | 45-77 | | 78-82 | > 82 | 810 |
|  | LVM/LVEDV | 0.6-1 | 1.1-1.1 | > 1.1 | 910 | 0.7-1.2 | | 1.3-1.3 | > 1.3 | 806 |
|  | RVEDVi | 53-98 | 99-107 | > 107 | 909 | 57-114 | | 115-123 | > 123 | 820 |
|  | RVESVi | 16-42 | 43-46 | > 46 | 909 | 21-53 | | 54-57 | > 57 | 810 |
|  | RVEF | 51-71 | 47-50 | < 47 | 906 | 46-68 | | 43-45 | < 43 | 830 |
|  | RVCOi | 1.9-4.2 | 1.7-1.8 | < 1.7 | 906 | 1.9-4.3 | | 1.5-1.8 | < 1.5 | 819 |
|  | RVSVi | 31-61 | 27-30 | < 27 | 909 | 31-68 | | 25-30 | < 25 | 828 |
|  | LAVi max | 22-53 | 54-59 | > 59 | 797 | 20-55 | | 56-61 | > 61 | 684 |
|  | LAEF | 53-79 | 50-52 | < 50 | 791 | 53-79 | | 50-52 | < 50 | 681 |
|  | RAVi max | 21-60 | 61-68 | > 68 | 854 | 22-67 | | 68-75 | > 75 | 761 |
|  | RAEF | 38-70 | 33-37 | < 33 | 844 | 35-68 | | 31-34 | < 31 | 771 |
| 60-69 | LVEDVi | 44-83 | 84-91 | > 91 | 918 | 46-96 | | 97-105 | > 105 | 803 |
|  | LVESVi | 10-31 | 32-35 | > 35 | 920 | 12-38 | | 39-43 | > 43 | 801 |
|  | LVSVi | 30-57 | 26-29 | < 26 | 918 | 30-64 | | 25-29 | < 25 | 799 |
|  | LVEF | 58-79 | 54-57 | < 54 | 917 | 54-78 | | 50-53 | < 50 | 803 |
|  | LVCOi | 1.9-3.9 | 1.6-1.8 | < 1.6 | 918 | 1.9-4.1 | | 1.6-1.8 | < 1.6 | 796 |
|  | LVMi | 36-61 | 62-66 | > 66 | 919 | 44-76 | | 77-81 | > 81 | 789 |
|  | LVM/LVEDV | 0.6-1 | 1.1-1.2 | > 1.2 | 918 | 0.7-1.2 | | 1.3-1.4 | > 1.4 | 787 |
|  | RVEDVi | 49-94 | 95-102 | > 102 | 917 | 54-111 | | 112-121 | > 121 | 798 |
|  | RVESVi | 15-39 | 40-43 | > 43 | 920 | 19-49 | | 50-54 | > 54 | 795 |
|  | RVEF | 51-72 | 48-50 | < 48 | 914 | 47-69 | | 44-46 | < 44 | 800 |
|  | RVCOi | 1.8-4 | 1.6-1.7 | < 1.6 | 917 | 1.9-4.2 | | 1.5-1.8 | < 1.5 | 798 |
|  | RVSVi | 29-59 | 26-28 | < 26 | 918 | 29-66 | | 24-28 | < 24 | 797 |
|  | LAVi max | 20-53 | 54-60 | > 60 | 797 | 19-55 | | 56-62 | > 62 | 645 |
|  | LAEF | 51-79 | 47-50 | < 47 | 776 | 51-80 | | 47-50 | < 47 | 632 |
|  | RAVi max | 21-62 | 63-71 | > 71 | 855 | 21-69 | | 70-78 | > 78 | 742 |
|  | RAEF | 38-71 | 33-37 | < 33 | 841 | 35-70 | | 31-34 | < 31 | 748 |
| 70+ | LVEDVi | 43-82 | 83-91 | > 91 | 1101 | 43-94 | | 95-103 | > 103 | 1256 |
|  | LVESVi | 9-30 | 31-34 | > 34 | 1102 | 11-37 | | 38-42 | > 42 | 1257 |
|  | LVSVi | 30-57 | 26-29 | < 26 | 1101 | 28-62 | | 23-27 | < 23 | 1256 |
|  | LVEF | 58-80 | 54-57 | < 54 | 1095 | 55-79 | | 50-54 | < 50 | 1243 |
|  | LVCOi | 1.8-3.9 | 1.6-1.7 | < 1.6 | 1100 | 1.9-4 | | 1.6-1.8 | < 1.6 | 1256 |
|  | LVMi | 36-62 | 63-67 | > 67 | 1101 | 43-75 | | 76-80 | > 80 | 1251 |
|  | LVM/LVEDV | 0.6-1.1 |  | > 1.2 | 1100 | 0.7-1.2 | | 1.3-1.5 | > 1.5 | 1249 |
|  | RVEDVi | 49-93 | 94-102 | > 102 | 1100 | 53-108 | | 109-118 | > 118 | 1257 |
|  | RVESVi | 14-38 | 39-42 | > 42 | 1100 | 18-48 | | 49-52 | > 52 | 1257 |
|  | RVEF | 52-72 | 48-51 | < 48 | 1093 | 48-69 | | 44-47 | < 44 | 1244 |
|  | RVCOi | 1.8-4 | 1.6-1.7 | < 1.6 | 1099 | 1.8-4.1 | | 1.5-1.7 | < 1.5 | 1255 |
|  | RVSVi | 28-59 | 25-27 | < 25 | 1100 | 28-65 | | 22-27 | < 22 | 1256 |
|  | LAVi max | 19-56 | 57-63 | > 63 | 942 | 18-56 | | 57-64 | > 64 | 953 |
|  | LAEF | 48-79 | 44-47 | < 44 | 903 | 48-80 | | 44-47 | < 44 | 914 |
|  | RAVi max | 20-65 | 66-76 | > 76 | 1028 | 19-70 | | 71-79 | > 79 | 1119 |
|  | RAEF | 38-72 | 33-37 | < 33 | 1001 | 35-73 | | 30-34 | < 30 | 1115 |

Footnote: All measurements were derived using cvi42 software with anatomical endocardial segmentation; papillary muscles and trabeculations were included in left ventricular mass measurements. Volumetric and mass measurements were indexed to bsa. Reference ranges are defined as the interval between the 2.5th and 97.5th centiles derived from age-specific generalized additive models for location, scale and shape (GAMLSS) using the Box–Cox Cole–Green distribution. Centile-based categories beyond the reference range are derived from LMS-based thresholds (P0.5, P2.5, P97.5, and P99.5) and describe the position of a measurement within the reference distribution rather than disease severity or diagnostic abnormality. Reference ranges are based on a White adult population.

Abbreviations: LV = left ventricle; RV = right ventricle; LA = left atrium; RA = right atrium; EDV = end-diastolic volume; ESV = end-systolic volume; SV = stroke volume; EF = ejection fraction; CO = cardiac output.

### 32. Table: Reference ranges and centile-based categories for cardiac chamber measurements derived using cvi42 software (anatomical segmentation), indexed to height, in a Black adult population

|  |  | **Female** | | | | | **Male** | | | |
| --- | --- | --- | --- | --- | --- | --- | --- | --- | --- | --- |
| **Age group** | **Metrics** | **Reference range** | **Mild** | **At least moderate** | **n** | **Reference range** | | **Mild** | **At least moderate** | **n** |
| 40-49 | LVEDVi | 50-85 | 86-89 | > 89 | 10 |  | |  |  |  |
|  | LVESVi | 12-33 | 34-36 | > 36 | 10 |  | |  |  |  |
|  | LVSVi | 33-57 | 29-32 | < 29 | 10 |  | |  |  |  |
|  | LVEF | 57-78 | 54-56 | < 54 | 10 |  | |  |  |  |
|  | LVCOi | 1.9-4 |  | < 1.8 | 10 |  | |  |  |  |
|  | LVMi | 41-66 | 67-75 | > 75 | 10 |  | |  |  |  |
|  | LVM/LVEDV | 0.6-1 | 1.1-1.2 | > 1.2 | 10 |  | |  |  |  |
|  | RVEDVi | 55-102 | 103-110 | > 110 | 10 |  | |  |  |  |
|  | RVESVi | 18-48 | 49-54 | > 54 | 10 |  | |  |  |  |
|  | RVEF | 50-72 | 48-49 | < 48 | 10 |  | |  |  |  |
|  | RVCOi | 2-3.8 | 1.8-1.9 | < 1.8 | 10 |  | |  |  |  |
|  | RVSVi | 33-59 | 28-32 | < 28 | 10 |  | |  |  |  |
|  | LAVi max |  |  |  |  |  | |  |  |  |
|  | LAEF |  |  |  |  |  | |  |  |  |
|  | RAVi max | 23-67 | 68-80 | > 80 | 10 |  | |  |  |  |
|  | RAEF | 30-65 | 25-29 | < 25 | 10 |  | |  |  |  |
| 50-59 | LVEDVi | 47-84 | 85-89 | > 89 | 95 | 44-98 | | 99-106 | > 106 | 78 |
|  | LVESVi | 12-32 | 33-35 | > 35 | 95 | 12-40 | | 41-45 | > 45 | 78 |
|  | LVSVi | 32-57 | 28-31 | < 28 | 95 | 30-62 | | 25-29 | < 25 | 76 |
|  | LVEF | 57-79 | 54-56 | < 54 | 95 | 55-77 | | 52-54 | < 52 | 76 |
|  | LVCOi | 1.9-3.9 | 1.7-1.8 | < 1.7 | 94 | 2.1-4.1 | | 1.9-2 | < 1.9 | 77 |
|  | LVMi | 40-66 | 67-72 | > 72 | 95 | 42-79 | | 80-84 | > 84 | 75 |
|  | LVM/LVEDV | 0.6-1.1 |  | > 1.2 | 95 | 0.7-1.1 | |  | > 1.2 | 75 |
|  | RVEDVi | 53-100 | 101-108 | > 108 | 95 | 65-111 | | 112-124 | > 124 | 75 |
|  | RVESVi | 17-46 | 47-51 | > 51 | 95 | 25-50 | | 51-55 | > 55 | 74 |
|  | RVEF | 50-72 | 47-49 | < 47 | 95 | 46-68 | | 43-45 | < 43 | 76 |
|  | RVCOi | 02-4 | 1.8-1.9 | < 1.8 | 94 | 1.9-4.5 | | 1.7-1.8 | < 1.7 | 78 |
|  | RVSVi | 32-59 | 28-31 | < 28 | 95 | 29-67 | | 24-28 | < 24 | 77 |
|  | LAVi max | 22-51 | 52-55 | > 55 | 89 | 20-53 | | 54-60 | > 60 | 64 |
|  | LAEF | 54-75 | 51-53 | < 51 | 88 | 53-76 | | 50-52 | < 50 | 64 |
|  | RAVi max | 22-63 | 64-74 | > 74 | 94 | 28-64 | | 65-70 | > 70 | 73 |
|  | RAEF | 31-65 | 26-30 | < 26 | 93 | 35-63 | | 32-34 | < 32 | 71 |
| 60-69 | LVEDVi | 42-81 | 82-88 | > 88 | 66 | 48-99 | | 100-108 | > 108 | 48 |
|  | LVESVi | 10-31 | 32-35 | > 35 | 66 | 10-41 | | 42-47 | > 47 | 48 |
|  | LVSVi | 29-56 | 25-28 | < 25 | 66 | 33-65 | | 30-32 | < 30 | 48 |
|  | LVEF | 57-80 | 53-56 | < 53 | 66 | 45-78 | | 34-44 | < 34 | 48 |
|  | LVCOi | 1.8-3.8 | 1.6-1.7 | < 1.6 | 66 | 2-4.2 | | 1.9-1.9 | < 1.9 | 48 |
|  | LVMi | 36-66 | 67-70 | > 70 | 66 | 43-80 | | 81-85 | > 85 | 48 |
|  | LVM/LVEDV | 0.7-1.1 | 1.2-1.3 | > 1.3 | 66 | 0.7-1.2 | | 1.3-1.3 | > 1.3 | 48 |
|  | RVEDVi | 50-97 | 98-106 | > 106 | 66 | 61-112 | | 113-125 | > 125 | 48 |
|  | RVESVi | 15-43 | 44-48 | > 48 | 66 | 22-53 | | 54-61 | > 61 | 48 |
|  | RVEF | 49-71 | 45-48 | < 45 | 64 | 46-69 | | 42-45 | < 42 | 48 |
|  | RVCOi | 1.9-4.1 | 1.7-1.8 | < 1.7 | 66 | 2.1-4.3 | | 1.9-2 | < 1.9 | 48 |
|  | RVSVi | 30-60 | 26-29 | < 26 | 66 | 33-66 | | 30-32 | < 30 | 48 |
|  | LAVi max | 20-55 | 56-61 | > 61 | 59 | 20-56 | | 57-64 | > 64 | 42 |
|  | LAEF | 50-75 | 45-49 | < 45 | 58 | 50-75 | | 46-49 | < 46 | 43 |
|  | RAVi max | 21-59 | 60-67 | > 67 | 64 | 24-69 | | 70-76 | > 76 | 45 |
|  | RAEF | 32-67 | 27-31 | < 27 | 66 | 32-64 | | 29-31 | < 29 | 46 |
| 70+ | LVEDVi | 38-80 | 81-88 | > 88 | 18 | 45-90 | | 91-99 | > 99 | 14 |
|  | LVESVi | 9-30 | 31-36 | > 36 | 18 | 7-37 | | 38-43 | > 43 | 14 |
|  | LVSVi | 26-56 | 22-25 | < 22 | 18 | 32-62 | | 30-31 | < 30 | 14 |
|  | LVEF | 56-81 | 49-55 | < 49 | 18 | 48-79 | | 39-47 | < 39 | 14 |
|  | LVCOi | 1.8-3.7 | 1.5-1.7 | < 1.5 | 18 | 2-4.5 | | 1.8-1.9 | < 1.8 | 14 |
|  | LVMi | 28-66 | 67-70 | > 70 | 18 | 41-76 | | 77-81 | > 81 | 14 |
|  | LVM/LVEDV | 0.7-1.2 | 1.3-1.4 | > 1.4 | 18 | 0.7-1.2 | | 1.3-1.3 | > 1.3 | 14 |
|  | RVEDVi | 47-95 | 96-103 | > 103 | 18 | 57-112 | | 113-125 | > 125 | 14 |
|  | RVESVi | 13-41 | 42-45 | > 45 | 18 | 20-58 | | 59-70 | > 70 | 14 |
|  | RVEF | 46-71 | 39-45 | < 39 | 18 | 45-70 | | 41-44 | < 41 | 14 |
|  | RVCOi | 2-3.9 | 1.8-1.9 | < 1.8 | 18 | 2.2-4.2 | | 2-2.1 | < 2 | 14 |
|  | RVSVi | 28-60 | 25-27 | < 25 | 18 | 35-65 | | 33-34 | < 33 | 14 |
|  | LAVi max | 19-61 | 62-73 | > 73 | 15 |  | |  |  |  |
|  | LAEF | 44-74 | 35-43 | < 35 | 16 |  | |  |  |  |
|  | RAVi max | 20-57 | 58-63 | > 63 | 17 | 19-74 | | 75-82 | > 82 | 12 |
|  | RAEF | 33-68 | 28-32 | < 28 | 17 | 29-66 | | 25-28 | < 25 | 12 |

Footnote: All measurements were derived using cvi42 software with anatomical endocardial segmentation; papillary muscles and trabeculations were included in left ventricular mass measurements. Volumetric and mass measurements were indexed to bsa. Reference ranges are defined as the interval between the 2.5th and 97.5th centiles derived from age-specific generalized additive models for location, scale and shape (GAMLSS) using the Box–Cox Cole–Green distribution. Centile-based categories beyond the reference range are derived from LMS-based thresholds (P0.5, P2.5, P97.5, and P99.5) and describe the position of a measurement within the reference distribution rather than disease severity or diagnostic abnormality. Reference ranges are based on a Black adult population.

Abbreviations: LV = left ventricle; RV = right ventricle; LA = left atrium; RA = right atrium; EDV = end-diastolic volume; ESV = end-systolic volume; SV = stroke volume; EF = ejection fraction; CO = cardiac output.

### 33. Table: Reference ranges and centile-based categories for cardiac chamber measurements derived using cvi42 software (anatomical segmentation), indexed to BSA, in an Asian adult population

|  |  | **Female** | | | | **Male** | | | |
| --- | --- | --- | --- | --- | --- | --- | --- | --- | --- |
| **Age group** | **Metrics** | **Reference range** | **Mild** | **At least moderate** | **n** | **Reference range** | **Mild** | **At least moderate** | **n** |
| 40-49 | LVEDVi | 47-76 | 77-82 | > 82 | 10 | 53-88 | 89-97 | > 97 | 20 |
|  | LVESVi | 14-26 | 27-30 | > 30 | 10 | 14-34 | 35-38 | > 38 | 20 |
|  | LVSVi | 31-51 | 27-30 | < 27 | 10 | 32-59 | 28-31 | < 28 | 20 |
|  | LVEF | 56-78 | 51-55 | < 51 | 10 | 55-77 | 51-54 | < 51 | 20 |
|  | LVCOi | 2-3.5 | 1.8-1.9 | < 1.8 | 10 | 02-4 | 1.8-1.9 | < 1.8 | 20 |
|  | LVMi | 36-53 | 54-58 | > 58 | 10 | 43-68 | 69-73 | > 73 | 20 |
|  | LVM/LVEDV | 0.6-0.9 | 01-1 | > 1 | 10 | 0.7-1 | 1.1-1.1 | > 1.1 | 20 |
|  | RVEDVi | 50-82 | 83-89 | > 89 | 10 | 60-106 | 107-118 | > 118 | 20 |
|  | RVESVi | 16-35 | 36-39 | > 39 | 10 | 23-49 | 50-54 | > 54 | 20 |
|  | RVEF | 50-72 | 46-49 | < 46 | 10 | 48-67 | 45-47 | < 45 | 20 |
|  | RVCOi | 1.9-3.5 |  | < 1.8 | 10 | 2-3.8 | 1.8-1.9 | < 1.8 | 20 |
|  | RVSVi | 30-53 | 27-29 | < 27 | 10 | 31-62 | 27-30 | < 27 | 20 |
|  | LAVi max | 21-45 | 46-49 | > 49 | 10 | 18-48 | 49-53 | > 53 | 20 |
|  | LAEF | 57-81 | 54-56 | < 54 | 10 | 58-84 | 55-57 | < 55 | 20 |
|  | RAVi max | 21-44 | 45-48 | > 48 | 10 | 21-62 | 63-71 | > 71 | 20 |
|  | RAEF | 37-71 | 35-36 | < 35 | 10 | 32-63 | 27-31 | < 27 | 20 |
| 50-59 | LVEDVi | 46-76 | 77-83 | > 83 | 83 | 49-87 | 88-96 | > 96 | 133 |
|  | LVESVi | 12-29 | 30-35 | > 35 | 83 | 12-34 | 35-39 | > 39 | 133 |
|  | LVSVi | 30-52 | 27-29 | < 27 | 83 | 31-59 | 27-30 | < 27 | 133 |
|  | LVEF | 57-79 | 52-56 | < 52 | 83 | 55-78 | 50-54 | < 50 | 132 |
|  | LVCOi | 1.9-3.6 |  | < 1.8 | 83 | 1.9-3.9 | 1.7-1.8 | < 1.7 | 133 |
|  | LVMi | 36-56 | 57-61 | > 61 | 83 | 41-70 | 71-76 | > 76 | 133 |
|  | LVM/LVEDV | 0.6-1 | 1.1-1.1 | > 1.1 | 83 | 0.7-1.1 |  | > 1.2 | 133 |
|  | RVEDVi | 49-86 | 87-94 | > 94 | 83 | 56-103 | 104-114 | > 114 | 133 |
|  | RVESVi | 16-37 | 38-41 | > 41 | 83 | 21-48 | 49-53 | > 53 | 133 |
|  | RVEF | 51-72 | 48-50 | < 48 | 83 | 47-67 | 44-46 | < 44 | 132 |
|  | RVCOi | 1.9-3.6 | 1.7-1.8 | < 1.7 | 83 | 1.8-3.8 | 1.6-1.7 | < 1.6 | 133 |
|  | RVSVi | 29-54 | 26-28 | < 26 | 83 | 29-60 | 25-28 | < 25 | 133 |
|  | LAVi max | 21-47 | 48-51 | > 51 | 77 | 19-50 | 51-55 | > 55 | 121 |
|  | LAEF | 54-80 | 51-53 | < 51 | 75 | 55-82 | 52-54 | < 52 | 120 |
|  | RAVi max | 21-50 | 51-55 | > 55 | 76 | 21-62 | 63-71 | > 71 | 128 |
|  | RAEF | 37-69 | 34-36 | < 34 | 76 | 34-65 | 29-33 | < 29 | 127 |
| 60-69 | LVEDVi | 43-78 | 79-87 | > 87 | 63 | 43-85 | 86-93 | > 93 | 118 |
|  | LVESVi | 9-30 | 31-36 | > 36 | 63 | 11-34 | 35-41 | > 41 | 119 |
|  | LVSVi | 29-55 | 27-28 | < 27 | 63 | 29-58 | 26-28 | < 26 | 118 |
|  | LVEF | 58-81 | 53-57 | < 53 | 63 | 54-79 | 49-53 | < 49 | 119 |
|  | LVCOi | 1.8-3.8 | 1.6-1.7 | < 1.6 | 63 | 1.8-3.8 | 1.5-1.7 | < 1.5 | 119 |
|  | LVMi | 35-61 | 62-69 | > 69 | 63 | 38-73 | 74-81 | > 81 | 119 |
|  | LVM/LVEDV | 0.6-1 | 1.1-1.1 | > 1.1 | 63 | 0.7-1.2 | 1.3-1.4 | > 1.4 | 118 |
|  | RVEDVi | 48-91 | 92-103 | > 103 | 63 | 50-99 | 100-108 | > 108 | 119 |
|  | RVESVi | 16-38 | 39-43 | > 43 | 63 | 19-46 | 47-52 | > 52 | 119 |
|  | RVEF | 53-72 | 50-52 | < 50 | 63 | 46-68 | 42-45 | < 42 | 118 |
|  | RVCOi | 1.8-3.8 | 1.6-1.7 | < 1.6 | 63 | 1.6-3.8 | 1.3-1.5 | < 1.3 | 119 |
|  | RVSVi | 28-58 | 25-27 | < 25 | 63 | 27-58 | 23-26 | < 23 | 119 |
|  | LAVi max | 21-50 | 51-55 | > 55 | 59 | 20-52 | 53-57 | > 57 | 110 |
|  | LAEF | 50-78 | 46-49 | < 46 | 58 | 51-80 | 48-50 | < 48 | 107 |
|  | RAVi max | 19-60 | 61-67 | > 67 | 61 | 21-62 | 63-71 | > 71 | 113 |
|  | RAEF | 36-68 | 31-35 | < 31 | 59 | 35-70 | 31-34 | < 31 | 114 |
| 70+ | LVEDVi | 40-80 | 81-91 | > 91 | 28 | 35-83 | 84-90 | > 90 | 41 |
|  | LVESVi | 9-25 | 26-28 | > 28 | 28 | 9-35 | 36-44 | > 44 | 41 |
|  | LVSVi | 29-61 | 26-28 | < 26 | 28 | 28-58 | 25-27 | < 25 | 40 |
|  | LVEF | 59-83 | 54-58 | < 54 | 26 | 54-81 | 47-53 | < 47 | 41 |
|  | LVCOi | 1.7-4 | 1.5-1.6 | < 1.5 | 28 | 1.4-3.8 | 1-1.3 | < 1 | 41 |
|  | LVMi | 34-67 | 68-80 | > 80 | 28 | 35-78 | 79-87 | > 87 | 41 |
|  | LVM/LVEDV | 0.7-1 | 1.1-1.1 | > 1.1 | 28 | 0.7-1.3 | 1.4-1.7 | > 1.7 | 41 |
|  | RVEDVi | 45-96 | 97-110 | > 110 | 28 | 43-96 | 97-103 | > 103 | 41 |
|  | RVESVi | 15-37 | 38-43 | > 43 | 28 | 17-45 | 46-51 | > 51 | 41 |
|  | RVEF | 54-73 | 52-53 | < 52 | 28 | 45-70 | 40-44 | < 40 | 40 |
|  | RVCOi | 1.7-4.1 | 1.5-1.6 | < 1.5 | 28 | 1.3-3.8 | 0.8-1.2 | < 0.8 | 41 |
|  | RVSVi | 27-62 | 24-26 | < 24 | 28 | 26-58 | 21-25 | < 21 | 40 |
|  | LAVi max | 21-54 | 55-61 | > 61 | 28 | 21-53 | 54-59 | > 59 | 34 |
|  | LAEF | 45-76 | 40-44 | < 40 | 27 | 48-79 | 45-47 | < 45 | 34 |
|  | RAVi max | 15-66 | 67-77 | > 77 | 27 | 21-62 | 63-71 | > 71 | 38 |
|  | RAEF | 35-70 | 25-34 | < 25 | 27 | 36-77 | 32-35 | < 32 | 39 |

Footnote: All measurements were derived using cvi42 software with anatomical endocardial segmentation; papillary muscles and trabeculations were included in left ventricular mass measurements. Volumetric and mass measurements were indexed to bsa. Reference ranges are defined as the interval between the 2.5th and 97.5th centiles derived from age-specific generalized additive models for location, scale and shape (GAMLSS) using the Box–Cox Cole–Green distribution. Centile-based categories beyond the reference range are derived from LMS-based thresholds (P0.5, P2.5, P97.5, and P99.5) and describe the position of a measurement within the reference distribution rather than disease severity or diagnostic abnormality. Reference ranges are based on an Asian adult population.

Abbreviations: LV = left ventricle; RV = right ventricle; LA = left atrium; RA = right atrium; EDV = end-diastolic volume; ESV = end-systolic volume; SV = stroke volume; EF = ejection fraction; CO = cardiac output.

### 34. Table: Reference ranges and centile-based categories for cardiac chamber measurements derived using cvi42 software (anatomical segmentation), indexed to BSA, in a Chinese adult population

|  |  | **Female** | | | | **Male** | | | |
| --- | --- | --- | --- | --- | --- | --- | --- | --- | --- |
| **Age group** | **Metrics** | **Reference range** | **Mild** | **At least moderate** | **n** | **Reference range** | **Mild** | **At least moderate** | **n** |
| 18-29 | LVEDVi | 56-93 | 94-103 | > 103 | 16 | 62-94 | 95-99 | > 99 | 19 |
|  | LVESVi | 15-37 | 38-43 | > 43 | 16 | 18-38 | 39-40 | > 40 | 19 |
|  | LVSVi | 36-67 | 33-35 | < 33 | 16 | 36-60 | 29-35 | < 29 | 19 |
|  | LVEF | 56-77 | 53-55 | < 53 | 16 | 56-72 | 53-55 | < 53 | 19 |
|  | LVCOi | 2.2-4.6 | 2-2.1 | < 2 | 16 | 2.7-4.4 | 2.6-2.6 | < 2.6 | 19 |
|  | LVMi | 33-55 | 56-61 | > 61 | 16 | 42-68 | 69-71 | > 71 | 19 |
|  | LVM/LVEDV | 0.5-0.7 |  | > 0.7 | 16 | 0.6-0.9 | 01-1 | > 1 | 19 |
|  | RVEDVi | 51-92 | 93-100 | > 100 | 16 | 54-101 | 102-108 | > 108 | 19 |
|  | RVESVi | 15-42 | 43-48 | > 48 | 16 | 15-43 | 44-45 | > 45 | 19 |
|  | RVEF | 50-77 | 46-49 | < 46 | 16 | 48-69 | 45-47 | < 45 | 18 |
|  | RVCOi | 1.7-4.1 | 1.5-1.6 | < 1.5 | 16 | 2-4.3 | 1.6-1.9 | < 1.6 | 19 |
|  | RVSVi | 31-60 | 29-30 | < 29 | 16 | 30-65 | 26-29 | < 26 | 19 |
|  | LAVi max | 23-53 | 54-59 | > 59 | 13 | 25-47 | 48-51 | > 51 | 16 |
|  | LAEF | 54-77 | 50-53 | < 50 | 13 | 52-70 | 49-51 | < 49 | 16 |
|  | RAVi max | 21-45 | 46-53 | > 53 | 15 | 18-58 | 59-65 | > 65 | 18 |
|  | RAEF | 41-73 | 35-40 | < 35 | 15 | 36-68 | 30-35 | < 30 | 18 |
| 30-39 | LVEDVi | 54-90 | 91-99 | > 99 | 18 | 58-92 | 93-98 | > 98 | 17 |
|  | LVESVi | 14-34 | 35-40 | > 40 | 18 | 16-35 | 36-38 | > 38 | 17 |
|  | LVSVi | 36-63 | 33-35 | < 33 | 18 | 35-59 | 29-34 | < 29 | 17 |
|  | LVEF | 57-78 | 53-56 | < 53 | 18 | 56-73 | 53-55 | < 53 | 17 |
|  | LVCOi | 2.2-4.5 | 2-2.1 | < 2 | 18 | 2.6-4.4 | 2.4-2.5 | < 2.4 | 17 |
|  | LVMi | 34-57 | 58-63 | > 63 | 18 | 43-69 | 70-73 | > 73 | 17 |
|  | LVM/LVEDV | 0.5-0.7 | 0.8-0.8 | > 0.8 | 18 | 0.6-1 | 1.1-1.1 | > 1.1 | 17 |
|  | RVEDVi | 51-91 | 92-100 | > 100 | 18 | 54-101 | 102-108 | > 108 | 17 |
|  | RVESVi | 14-41 | 42-46 | > 46 | 18 | 15-43 | 44-46 | > 46 | 17 |
|  | RVEF | 50-76 | 46-49 | < 46 | 18 | 48-69 | 45-47 | < 45 | 17 |
|  | RVCOi | 1.8-3.9 | 1.5-1.7 | < 1.5 | 18 | 1.9-4.3 | 1.6-1.8 | < 1.6 | 17 |
|  | RVSVi | 31-58 | 28-30 | < 28 | 18 | 29-64 | 24-28 | < 24 | 17 |
|  | LAVi max | 23-52 | 53-58 | > 58 | 18 | 24-47 | 48-51 | > 51 | 14 |
|  | LAEF | 53-76 | 50-52 | < 50 | 18 | 51-72 | 48-50 | < 48 | 14 |
|  | RAVi max | 22-49 | 50-57 | > 57 | 18 | 18-59 | 60-67 | > 67 | 16 |
|  | RAEF | 41-72 | 35-40 | < 35 | 18 | 36-69 | 31-35 | < 31 | 16 |
| 40-49 | LVEDVi | 52-86 | 87-95 | > 95 | 24 | 55-91 | 92-97 | > 97 | 20 |
|  | LVESVi | 13-32 | 33-37 | > 37 | 24 | 14-34 | 35-37 | > 37 | 20 |
|  | LVSVi | 35-60 | 32-34 | < 32 | 24 | 34-59 | 29-33 | < 29 | 20 |
|  | LVEF | 58-79 | 54-57 | < 54 | 24 | 56-74 | 53-55 | < 53 | 19 |
|  | LVCOi | 2.1-4.4 | 1.9-2 | < 1.9 | 24 | 2.4-4.3 | 2.2-2.3 | < 2.2 | 20 |
|  | LVMi | 35-58 | 59-65 | > 65 | 24 | 43-70 | 71-75 | > 75 | 20 |
|  | LVM/LVEDV | 0.5-0.8 | 0.9-0.9 | > 0.9 | 24 | 0.6-1 | 1.1-1.1 | > 1.1 | 20 |
|  | RVEDVi | 50-91 | 92-100 | > 100 | 24 | 53-100 | 101-108 | > 108 | 20 |
|  | RVESVi | 14-40 | 41-45 | > 45 | 24 | 15-44 | 45-47 | > 47 | 20 |
|  | RVEF | 50-75 | 46-49 | < 46 | 24 | 48-70 | 45-47 | < 45 | 19 |
|  | RVCOi | 1.8-3.8 | 1.6-1.7 | < 1.6 | 24 | 1.8-4.2 | 1.5-1.7 | < 1.5 | 20 |
|  | RVSVi | 30-57 | 27-29 | < 27 | 24 | 28-63 | 23-27 | < 23 | 20 |
|  | LAVi max | 22-51 | 52-57 | > 57 | 22 | 22-47 | 48-51 | > 51 | 18 |
|  | LAEF | 53-75 | 49-52 | < 49 | 22 | 51-74 | 47-50 | < 47 | 18 |
|  | RAVi max | 22-53 | 54-63 | > 63 | 23 | 19-61 | 62-69 | > 69 | 19 |
|  | RAEF | 41-70 | 36-40 | < 36 | 23 | 35-69 | 31-34 | < 31 | 19 |
| 50-59 | LVEDVi | 50-83 | 84-90 | > 90 | 66 | 51-89 | 90-97 | > 97 | 49 |
|  | LVESVi | 11-29 | 30-34 | > 34 | 66 | 13-34 | 35-37 | > 37 | 49 |
|  | LVSVi | 34-58 | 31-33 | < 31 | 66 | 33-59 | 29-32 | < 29 | 49 |
|  | LVEF | 59-80 | 55-58 | < 55 | 65 | 56-76 | 53-55 | < 53 | 48 |
|  | LVCOi | 2.1-4 | 02-2 | < 2 | 66 | 2.2-4.2 | 2-2.1 | < 2 | 49 |
|  | LVMi | 37-60 | 61-67 | > 67 | 66 | 44-73 | 74-81 | > 81 | 49 |
|  | LVM/LVEDV | 0.6-0.9 | 01-1 | > 1 | 66 | 0.6-1.1 |  | > 1.2 | 49 |
|  | RVEDVi | 49-90 | 91-99 | > 99 | 66 | 53-100 | 101-108 | > 108 | 49 |
|  | RVESVi | 14-39 | 40-43 | > 43 | 66 | 15-45 | 46-48 | > 48 | 49 |
|  | RVEF | 50-74 | 46-49 | < 46 | 66 | 47-70 | 45-46 | < 45 | 49 |
|  | RVCOi | 1.9-3.7 | 1.7-1.8 | < 1.7 | 66 | 1.8-4.2 | 1.5-1.7 | < 1.5 | 49 |
|  | RVSVi | 30-57 | 27-29 | < 27 | 66 | 27-62 | 22-26 | < 22 | 49 |
|  | LAVi max | 22-50 | 51-56 | > 56 | 60 | 21-46 | 47-50 | > 50 | 40 |
|  | LAEF | 51-76 | 48-50 | < 48 | 58 | 50-77 | 46-49 | < 46 | 40 |
|  | RAVi max | 23-58 | 59-69 | > 69 | 63 | 20-63 | 64-71 | > 71 | 46 |
|  | RAEF | 40-68 | 36-39 | < 36 | 63 | 35-71 | 31-34 | < 31 | 46 |
| 60-69 | LVEDVi | 47-79 | 80-86 | > 86 | 52 | 47-88 | 89-97 | > 97 | 37 |
|  | LVESVi | 10-27 | 28-31 | > 31 | 52 | 11-32 | 33-35 | > 35 | 36 |
|  | LVSVi | 33-58 | 29-32 | < 29 | 52 | 32-61 | 28-31 | < 28 | 37 |
|  | LVEF | 60-81 | 55-59 | < 55 | 52 | 56-78 | 53-55 | < 53 | 36 |
|  | LVCOi | 2.1-3.7 | 02-2 | < 2 | 52 | 1.9-4.1 | 1.7-1.8 | < 1.7 | 37 |
|  | LVMi | 38-61 | 62-70 | > 70 | 52 | 44-78 | 79-92 | > 92 | 37 |
|  | LVM/LVEDV | 0.6-1.1 | 1.2-1.4 | > 1.4 | 52 | 0.6-1.1 |  | > 1.2 | 37 |
|  | RVEDVi | 48-88 | 89-97 | > 97 | 52 | 53-100 | 101-107 | > 107 | 36 |
|  | RVESVi | 14-37 | 38-42 | > 42 | 52 | 17-46 | 47-50 | > 50 | 37 |
|  | RVEF | 51-74 | 46-50 | < 46 | 52 | 47-71 | 45-46 | < 45 | 36 |
|  | RVCOi | 1.9-3.6 |  | < 1.8 | 52 | 1.7-4.2 | 1.4-1.6 | < 1.4 | 37 |
|  | RVSVi | 30-56 | 27-29 | < 27 | 52 | 26-61 | 20-25 | < 20 | 37 |
|  | LAVi max | 21-49 | 50-55 | > 55 | 47 | 19-46 | 47-50 | > 50 | 30 |
|  | LAEF | 49-76 | 45-48 | < 45 | 46 | 48-80 | 44-47 | < 44 | 30 |
|  | RAVi max | 23-63 | 64-76 | > 76 | 46 | 20-65 | 66-73 | > 73 | 35 |
|  | RAEF | 40-66 | 36-39 | < 36 | 46 | 35-72 | 32-34 | < 32 | 35 |
| 70+ | LVEDVi | 45-76 | 77-82 | > 82 | 12 |  |  |  |  |
|  | LVESVi | 9-25 | 26-28 | > 28 | 12 |  |  |  |  |
|  | LVSVi | 30-58 | 27-29 | < 27 | 12 |  |  |  |  |
|  | LVEF | 61-82 | 55-60 | < 55 | 12 |  |  |  |  |
|  | LVCOi | 2-3.8 | 1.9-1.9 | < 1.9 | 12 |  |  |  |  |
|  | LVMi | 39-63 | 64-72 | > 72 | 12 |  |  |  |  |
|  | LVM/LVEDV | 0.7-1.3 | 1.4-1.8 | > 1.8 | 12 |  |  |  |  |
|  | RVEDVi | 47-86 | 87-94 | > 94 | 12 |  |  |  |  |
|  | RVESVi | 14-36 | 37-40 | > 40 | 12 |  |  |  |  |
|  | RVEF | 51-73 | 47-50 | < 47 | 12 |  |  |  |  |
|  | RVCOi | 1.9-3.7 |  | < 1.8 | 12 |  |  |  |  |
|  | RVSVi | 30-57 | 27-29 | < 27 | 12 |  |  |  |  |
|  | LAVi max | 21-48 | 49-53 | > 53 | 11 |  |  |  |  |
|  | LAEF | 47-78 | 42-46 | < 42 | 12 |  |  |  |  |
|  | RAVi max | 23-68 | 69-82 | > 82 | 11 |  |  |  |  |
|  | RAEF | 39-65 | 36-38 | < 36 | 11 |  |  |  |  |

Footnote: All measurements were derived using cvi42 software with anatomical endocardial segmentation; papillary muscles and trabeculations were included in left ventricular mass measurements. Volumetric and mass measurements were indexed to bsa. Reference ranges are defined as the interval between the 2.5th and 97.5th centiles derived from age-specific generalized additive models for location, scale and shape (GAMLSS) using the Box–Cox Cole–Green distribution. Centile-based categories beyond the reference range are derived from LMS-based thresholds (P0.5, P2.5, P97.5, and P99.5) and describe the position of a measurement within the reference distribution rather than disease severity or diagnostic abnormality. Reference ranges are based on a Chinese adult population.

Abbreviations: LV = left ventricle; RV = right ventricle; LA = left atrium; RA = right atrium; EDV = end-diastolic volume; ESV = end-systolic volume; SV = stroke volume; EF = ejection fraction; CO = cardiac output.

### 35. Table: Reference ranges and centile-based categories for cardiac chamber measurements derived using cvi42 software (anatomical segmentation), indexed to BSA, in a Mixed adult population

|  |  | **Female** | | | | **Male** | | | |
| --- | --- | --- | --- | --- | --- | --- | --- | --- | --- |
| **Age group** | **Metrics** | **Reference range** | **Mild** | **At least moderate** | **n** | **Reference range** | **Mild** | **At least moderate** | **n** |
| 50-59 | LVEDVi | 49-86 | 87-94 | > 94 | 134 | 54-103 | 104-114 | > 114 | 66 |
|  | LVESVi | 11-31 | 32-35 | > 35 | 134 | 13-37 | 38-41 | > 41 | 66 |
|  | LVSVi | 33-62 | 30-32 | < 30 | 133 | 35-68 | 32-34 | < 32 | 65 |
|  | LVEF | 58-80 | 54-57 | < 54 | 134 | 56-79 | 52-55 | < 52 | 66 |
|  | LVCOi | 2-4.1 | 1.8-1.9 | < 1.8 | 133 | 2.1-4.6 | 1.9-2 | < 1.9 | 66 |
|  | LVMi | 36-63 | 64-69 | > 69 | 134 | 45-75 | 76-79 | > 79 | 64 |
|  | LVM/LVEDV | 0.6-0.9 | 01-1 | > 1 | 134 | 0.7-1.1 | 1.2-1.4 | > 1.4 | 64 |
|  | RVEDVi | 53-102 | 103-114 | > 114 | 134 | 60-121 | 122-132 | > 132 | 65 |
|  | RVESVi | 17-42 | 43-47 | > 47 | 134 | 23-52 | 53-57 | > 57 | 65 |
|  | RVEF | 52-70 | 48-51 | < 48 | 134 | 50-69 | 47-49 | < 47 | 66 |
|  | RVCOi | 1.9-4 | 1.6-1.8 | < 1.6 | 133 | 2.2-4.5 | 1.9-2.1 | < 1.9 | 66 |
|  | RVSVi | 31-61 | 27-30 | < 27 | 134 | 35-72 | 31-34 | < 31 | 66 |
|  | LAVi max | 21-48 | 49-51 | > 51 | 121 | 24-57 | 58-64 | > 64 | 53 |
|  | LAEF | 55-78 | 52-54 | < 52 | 120 | 54-78 | 51-53 | < 51 | 53 |
|  | RAVi max | 21-63 | 64-73 | > 73 | 128 | 21-68 | 69-76 | > 76 | 64 |
|  | RAEF | 37-64 | 31-36 | < 31 | 124 | 35-63 | 31-34 | < 31 | 64 |
| 60-69 | LVEDVi | 43-83 | 84-89 | > 89 | 110 | 47-93 | 94-101 | > 101 | 59 |
|  | LVESVi | 10-30 | 31-34 | > 34 | 110 | 11-37 | 38-41 | > 41 | 59 |
|  | LVSVi | 31-57 | 27-30 | < 27 | 110 | 32-62 | 27-31 | < 27 | 59 |
|  | LVEF | 58-81 | 54-57 | < 54 | 110 | 55-77 | 51-54 | < 51 | 58 |
|  | LVCOi | 1.9-3.9 |  | < 1.8 | 110 | 2-4.2 | 1.8-1.9 | < 1.8 | 59 |
|  | LVMi | 35-61 | 62-66 | > 66 | 110 | 44-76 | 77-81 | > 81 | 59 |
|  | LVM/LVEDV | 0.6-1 | 1.1-1.1 | > 1.1 | 110 | 0.7-1.2 | 1.3-1.5 | > 1.5 | 59 |
|  | RVEDVi | 49-93 | 94-100 | > 100 | 110 | 57-115 | 116-128 | > 128 | 58 |
|  | RVESVi | 17-39 | 40-44 | > 44 | 110 | 20-52 | 53-58 | > 58 | 58 |
|  | RVEF | 51-71 | 47-50 | < 47 | 110 | 48-67 | 46-47 | < 46 | 59 |
|  | RVCOi | 1.9-3.9 | 1.7-1.8 | < 1.7 | 110 | 2-4.3 | 1.8-1.9 | < 1.8 | 59 |
|  | RVSVi | 29-58 | 24-28 | < 24 | 110 | 32-67 | 28-31 | < 28 | 59 |
|  | LAVi max | 22-51 | 52-56 | > 56 | 100 | 21-56 | 57-63 | > 63 | 52 |
|  | LAEF | 52-79 | 49-51 | < 49 | 98 | 50-79 | 47-49 | < 47 | 52 |
|  | RAVi max | 20-62 | 63-70 | > 70 | 103 | 18-71 | 72-79 | > 79 | 57 |
|  | RAEF | 35-68 | 29-34 | < 29 | 103 | 36-68 | 33-35 | < 33 | 57 |
| 70+ | LVEDVi | 41-76 | 77-81 | > 81 | 50 | 39-85 | 86-91 | > 91 | 33 |
|  | LVESVi | 9-28 | 29-32 | > 32 | 51 | 9-36 | 37-41 | > 41 | 34 |
|  | LVSVi | 27-52 | 22-26 | < 22 | 50 | 29-57 | 24-28 | < 24 | 34 |
|  | LVEF | 59-81 | 54-58 | < 54 | 51 | 54-76 | 49-53 | < 49 | 35 |
|  | LVCOi | 1.9-3.8 | 1.7-1.8 | < 1.7 | 51 | 1.8-3.8 | 1.7-1.7 | < 1.7 | 35 |
|  | LVMi | 35-60 | 61-64 | > 64 | 51 | 43-78 | 79-85 | > 85 | 34 |
|  | LVM/LVEDV | 0.6-1 | 1.1-1.1 | > 1.1 | 50 | 0.7-1.3 | 1.4-1.5 | > 1.5 | 33 |
|  | RVEDVi | 51-82 | 83-86 | > 86 | 51 | 54-111 | 112-125 | > 125 | 35 |
|  | RVESVi | 17-37 | 38-41 | > 41 | 51 | 21-49 | 50-54 | > 54 | 35 |
|  | RVEF | 50-71 | 45-49 | < 45 | 51 | 47-66 | 44-46 | < 44 | 35 |
|  | RVCOi | 1.8-3.9 | 1.7-1.7 | < 1.7 | 51 | 1.8-4.1 | 1.7-1.7 | < 1.7 | 35 |
|  | RVSVi | 28-54 | 24-27 | < 24 | 50 | 29-62 | 25-28 | < 25 | 35 |
|  | LAVi max | 22-55 | 56-61 | > 61 | 45 | 18-55 | 56-63 | > 63 | 32 |
|  | LAEF | 50-80 | 47-49 | < 47 | 44 | 46-81 | 42-45 | < 42 | 30 |
|  | RAVi max | 19-61 | 62-68 | > 68 | 47 | 17-72 | 73-81 | > 81 | 31 |
|  | RAEF | 33-72 | 28-32 | < 28 | 46 | 36-76 | 34-35 | < 34 | 31 |

Footnote: All measurements were derived using cvi42 software with anatomical endocardial segmentation; papillary muscles and trabeculations were included in left ventricular mass measurements. Volumetric and mass measurements were indexed to bsa. Reference ranges are defined as the interval between the 2.5th and 97.5th centiles derived from age-specific generalized additive models for location, scale and shape (GAMLSS) using the Box–Cox Cole–Green distribution. Centile-based categories beyond the reference range are derived from LMS-based thresholds (P0.5, P2.5, P97.5, and P99.5) and describe the position of a measurement within the reference distribution rather than disease severity or diagnostic abnormality. Reference ranges are based on a Mixed adult population.

Abbreviations: LV = left ventricle; RV = right ventricle; LA = left atrium; RA = right atrium; EDV = end-diastolic volume; ESV = end-systolic volume; SV = stroke volume; EF = ejection fraction; CO = cardiac output.

### 36. Table: Reference ranges and centile-based categories for cardiac chamber measurements derived using cvi42 software (papillary segmentation), indexed to BSA, in a White adult population

|  |  | **Female** | | | | | **Male** | | | |
| --- | --- | --- | --- | --- | --- | --- | --- | --- | --- | --- |
| **Age group** | **Metrics** | **Reference range** | **Mild** | **At least moderate** | **n** | **Reference range** | | **Mild** | **At least moderate** | **n** |
| 18-29 | LVEDVi | 53-96 | 97-102 | > 102 | 102 | 62-112 | | 113-120 | > 120 | 103 |
|  | LVESVi | 14-38 | 39-41 | > 41 | 102 | 15-46 | | 47-49 | > 49 | 104 |
|  | LVSVi | 33-63 | 29-32 | < 29 | 102 | 37-73 | | 32-36 | < 32 | 101 |
|  | LVEF | 55-74 | 52-54 | < 52 | 100 | 51-72 | | 48-50 | < 48 | 102 |
|  | LVCOi | 2.3-4.7 | 2-2.2 | < 2 | 102 | 2.2-4.8 | | 1.7-2.1 | < 1.7 | 101 |
|  | LVMi | 40-62 | 63-66 | > 66 | 102 | 50-84 | | 85-89 | > 89 | 104 |
|  | LVM/LVEDV | 0.6-0.9 | 01-1 | > 1 | 102 | 0.6-1 | | 1.1-1.1 | > 1.1 | 102 |
|  | RVEDVi | 55-101 | 102-107 | > 107 | 102 | 63-125 | | 126-133 | > 133 | 102 |
|  | RVESVi | 13-51 | 52-57 | > 57 | 100 | 25-59 | | 60-64 | > 64 | 92 |
|  | RVEF | 47-71 | 44-46 | < 44 | 97 | 45-68 | | 41-44 | < 41 | 97 |
|  | RVCOi | 1.9-5 | 1.5-1.8 | < 1.5 | 101 | 2.2-4.9 | | 1.7-2.1 | < 1.7 | 98 |
|  | RVSVi | 29-67 | 24-28 | < 24 | 102 | 36-75 | | 31-35 | < 31 | 99 |
|  | LAVi max | 25-50 | 51-54 | > 54 | 50 | 27-58 | | 59-67 | > 67 | 52 |
|  | LAEF | 56-75 | 54-55 | < 54 | 50 | 53-72 | | 50-52 | < 50 | 52 |
|  | RAVi max | 21-49 | 50-54 | > 54 | 80 | 25-62 | | 63-70 | > 70 | 82 |
|  | RAEF | 38-78 | 33-37 | < 33 | 78 | 35-74 | | 31-34 | < 31 | 82 |
| 30-39 | LVEDVi | 51-92 | 93-99 | > 99 | 137 | 54-102 | | 103-109 | > 109 | 134 |
|  | LVESVi | 13-36 | 37-40 | > 40 | 136 | 13-41 | | 42-44 | > 44 | 134 |
|  | LVSVi | 32-61 | 28-31 | < 28 | 135 | 33-66 | | 28-32 | < 28 | 133 |
|  | LVEF | 55-75 | 53-54 | < 53 | 132 | 52-74 | | 48-51 | < 48 | 134 |
|  | LVCOi | 2.2-4.6 | 2-2.1 | < 2 | 136 | 2.1-4.6 | | 1.7-2 | < 1.7 | 128 |
|  | LVMi | 39-61 | 62-66 | > 66 | 137 | 45-77 | | 78-81 | > 81 | 134 |
|  | LVM/LVEDV | 0.6-0.9 | 1-1.1 | > 1.1 | 137 | 0.6-1 | | 1.1-1.2 | > 1.2 | 135 |
|  | RVEDVi | 54-99 | 100-107 | > 107 | 137 | 61-117 | | 118-125 | > 125 | 131 |
|  | RVESVi | 14-48 | 49-54 | > 54 | 135 | 24-56 | | 57-61 | > 61 | 129 |
|  | RVEF | 48-70 | 45-47 | < 45 | 127 | 45-68 | | 42-44 | < 42 | 131 |
|  | RVCOi | 2-4.8 | 1.7-1.9 | < 1.7 | 134 | 2.1-4.8 | | 1.7-2 | < 1.7 | 125 |
|  | RVSVi | 30-65 | 25-29 | < 25 | 135 | 33-71 | | 28-32 | < 28 | 132 |
|  | LAVi max | 25-51 | 52-56 | > 56 | 84 | 25-56 | | 57-64 | > 64 | 66 |
|  | LAEF | 55-76 | 53-54 | < 53 | 83 | 53-73 | | 50-52 | < 50 | 66 |
|  | RAVi max | 20-50 | 51-55 | > 55 | 113 | 23-61 | | 62-69 | > 69 | 119 |
|  | RAEF | 38-75 | 33-37 | < 33 | 110 | 35-72 | | 31-34 | < 31 | 118 |
| 40-49 | LVEDVi | 50-91 | 92-98 | > 98 | 518 | 53-103 | | 104-111 | > 111 | 438 |
|  | LVESVi | 12-34 | 35-38 | > 38 | 518 | 13-41 | | 42-45 | > 45 | 438 |
|  | LVSVi | 32-62 | 28-31 | < 28 | 518 | 33-68 | | 28-32 | < 28 | 435 |
|  | LVEF | 56-76 | 53-55 | < 53 | 512 | 53-75 | | 49-52 | < 49 | 434 |
|  | LVCOi | 2.1-4.4 | 1.9-2 | < 1.9 | 515 | 2.1-4.5 | | 1.7-2 | < 1.7 | 429 |
|  | LVMi | 38-61 | 62-66 | > 66 | 518 | 46-78 | | 79-83 | > 83 | 429 |
|  | LVM/LVEDV | 0.6-0.9 | 1-1.1 | > 1.1 | 518 | 0.6-1.1 | |  | > 1.2 | 426 |
|  | RVEDVi | 54-100 | 101-108 | > 108 | 517 | 61-119 | | 120-127 | > 127 | 430 |
|  | RVESVi | 16-46 | 47-50 | > 50 | 517 | 23-56 | | 57-61 | > 61 | 422 |
|  | RVEF | 49-70 | 46-48 | < 46 | 508 | 45-68 | | 42-44 | < 42 | 431 |
|  | RVCOi | 2-4.5 | 1.7-1.9 | < 1.7 | 514 | 2.1-4.8 | | 1.7-2 | < 1.7 | 426 |
|  | RVSVi | 31-64 | 26-30 | < 26 | 516 | 33-71 | | 27-32 | < 27 | 432 |
|  | LAVi max | 24-54 | 55-59 | > 59 | 440 | 23-55 | | 56-62 | > 62 | 358 |
|  | LAEF | 55-78 | 52-54 | < 52 | 434 | 53-76 | | 50-52 | < 50 | 353 |
|  | RAVi max | 21-56 | 57-62 | > 62 | 500 | 24-69 | | 70-78 | > 78 | 408 |
|  | RAEF | 38-72 | 33-37 | < 33 | 497 | 35-69 | | 31-34 | < 31 | 410 |
| 50-59 | LVEDVi | 48-87 | 88-95 | > 95 | 911 | 49-98 | | 99-107 | > 107 | 831 |
|  | LVESVi | 11-32 | 33-36 | > 36 | 911 | 13-39 | | 40-44 | > 44 | 835 |
|  | LVSVi | 31-60 | 28-30 | < 28 | 911 | 31-65 | | 26-30 | < 26 | 822 |
|  | LVEF | 57-78 | 53-56 | < 53 | 906 | 54-77 | | 50-53 | < 50 | 833 |
|  | LVCOi | 1.9-4.1 | 1.7-1.8 | < 1.7 | 908 | 2-4.2 | | 1.7-1.9 | < 1.7 | 816 |
|  | LVMi | 37-61 | 62-66 | > 66 | 910 | 45-77 | | 78-82 | > 82 | 810 |
|  | LVM/LVEDV | 0.6-1 | 1.1-1.1 | > 1.1 | 910 | 0.7-1.2 | | 1.3-1.3 | > 1.3 | 806 |
|  | RVEDVi | 53-98 | 99-107 | > 107 | 909 | 57-114 | | 115-123 | > 123 | 820 |
|  | RVESVi | 16-42 | 43-46 | > 46 | 909 | 21-53 | | 54-57 | > 57 | 810 |
|  | RVEF | 51-71 | 47-50 | < 47 | 906 | 46-68 | | 43-45 | < 43 | 830 |
|  | RVCOi | 1.9-4.2 | 1.7-1.8 | < 1.7 | 906 | 1.9-4.3 | | 1.5-1.8 | < 1.5 | 819 |
|  | RVSVi | 31-61 | 27-30 | < 27 | 909 | 31-68 | | 25-30 | < 25 | 828 |
|  | LAVi max | 22-53 | 54-59 | > 59 | 797 | 20-55 | | 56-61 | > 61 | 684 |
|  | LAEF | 53-79 | 50-52 | < 50 | 791 | 53-79 | | 50-52 | < 50 | 681 |
|  | RAVi max | 21-60 | 61-68 | > 68 | 854 | 22-67 | | 68-75 | > 75 | 761 |
|  | RAEF | 38-70 | 33-37 | < 33 | 844 | 35-68 | | 31-34 | < 31 | 771 |
| 60-69 | LVEDVi | 44-83 | 84-91 | > 91 | 918 | 46-96 | | 97-105 | > 105 | 803 |
|  | LVESVi | 10-31 | 32-35 | > 35 | 920 | 12-38 | | 39-43 | > 43 | 801 |
|  | LVSVi | 30-57 | 26-29 | < 26 | 918 | 30-64 | | 25-29 | < 25 | 799 |
|  | LVEF | 58-79 | 54-57 | < 54 | 917 | 54-78 | | 50-53 | < 50 | 803 |
|  | LVCOi | 1.9-3.9 | 1.6-1.8 | < 1.6 | 918 | 1.9-4.1 | | 1.6-1.8 | < 1.6 | 796 |
|  | LVMi | 36-61 | 62-66 | > 66 | 919 | 44-76 | | 77-81 | > 81 | 789 |
|  | LVM/LVEDV | 0.6-1 | 1.1-1.2 | > 1.2 | 918 | 0.7-1.2 | | 1.3-1.4 | > 1.4 | 787 |
|  | RVEDVi | 49-94 | 95-102 | > 102 | 917 | 54-111 | | 112-121 | > 121 | 798 |
|  | RVESVi | 15-39 | 40-43 | > 43 | 920 | 19-49 | | 50-54 | > 54 | 795 |
|  | RVEF | 51-72 | 48-50 | < 48 | 914 | 47-69 | | 44-46 | < 44 | 800 |
|  | RVCOi | 1.8-4 | 1.6-1.7 | < 1.6 | 917 | 1.9-4.2 | | 1.5-1.8 | < 1.5 | 798 |
|  | RVSVi | 29-59 | 26-28 | < 26 | 918 | 29-66 | | 24-28 | < 24 | 797 |
|  | LAVi max | 20-53 | 54-60 | > 60 | 797 | 19-55 | | 56-62 | > 62 | 645 |
|  | LAEF | 51-79 | 47-50 | < 47 | 776 | 51-80 | | 47-50 | < 47 | 632 |
|  | RAVi max | 21-62 | 63-71 | > 71 | 855 | 21-69 | | 70-78 | > 78 | 742 |
|  | RAEF | 38-71 | 33-37 | < 33 | 841 | 35-70 | | 31-34 | < 31 | 748 |
| 70+ | LVEDVi | 43-82 | 83-91 | > 91 | 1101 | 43-94 | | 95-103 | > 103 | 1256 |
|  | LVESVi | 9-30 | 31-34 | > 34 | 1102 | 11-37 | | 38-42 | > 42 | 1257 |
|  | LVSVi | 30-57 | 26-29 | < 26 | 1101 | 28-62 | | 23-27 | < 23 | 1256 |
|  | LVEF | 58-80 | 54-57 | < 54 | 1095 | 55-79 | | 50-54 | < 50 | 1243 |
|  | LVCOi | 1.8-3.9 | 1.6-1.7 | < 1.6 | 1100 | 1.9-4 | | 1.6-1.8 | < 1.6 | 1256 |
|  | LVMi | 36-62 | 63-67 | > 67 | 1101 | 43-75 | | 76-80 | > 80 | 1251 |
|  | LVM/LVEDV | 0.6-1.1 |  | > 1.2 | 1100 | 0.7-1.2 | | 1.3-1.5 | > 1.5 | 1249 |
|  | RVEDVi | 49-93 | 94-102 | > 102 | 1100 | 53-108 | | 109-118 | > 118 | 1257 |
|  | RVESVi | 14-38 | 39-42 | > 42 | 1100 | 18-48 | | 49-52 | > 52 | 1257 |
|  | RVEF | 52-72 | 48-51 | < 48 | 1093 | 48-69 | | 44-47 | < 44 | 1244 |
|  | RVCOi | 1.8-4 | 1.6-1.7 | < 1.6 | 1099 | 1.8-4.1 | | 1.5-1.7 | < 1.5 | 1255 |
|  | RVSVi | 28-59 | 25-27 | < 25 | 1100 | 28-65 | | 22-27 | < 22 | 1256 |
|  | LAVi max | 19-56 | 57-63 | > 63 | 942 | 18-56 | | 57-64 | > 64 | 953 |
|  | LAEF | 48-79 | 44-47 | < 44 | 903 | 48-80 | | 44-47 | < 44 | 914 |
|  | RAVi max | 20-65 | 66-76 | > 76 | 1028 | 19-70 | | 71-79 | > 79 | 1119 |
|  | RAEF | 38-72 | 33-37 | < 33 | 1001 | 35-73 | | 30-34 | < 30 | 1115 |

### 37. Table: Reference ranges and centile-based categories for cardiac chamber measurements derived using cvi42 software (papillary segmentation), indexed to BSA, in a Black adult population

|  |  | **Female** | | | | | **Male** | | | |
| --- | --- | --- | --- | --- | --- | --- | --- | --- | --- | --- |
| **Age group** | **Metrics** | **Reference range** | **Mild** | **At least moderate** | **n** | **Reference range** | | **Mild** | **At least moderate** | **n** |
| 40-49 | LVEDVi | 50-85 | 86-89 | > 89 | 10 |  | |  |  |  |
|  | LVESVi | 12-33 | 34-36 | > 36 | 10 |  | |  |  |  |
|  | LVSVi | 33-57 | 29-32 | < 29 | 10 |  | |  |  |  |
|  | LVEF | 57-78 | 54-56 | < 54 | 10 |  | |  |  |  |
|  | LVCOi | 1.9-4 |  | < 1.8 | 10 |  | |  |  |  |
|  | LVMi | 41-66 | 67-75 | > 75 | 10 |  | |  |  |  |
|  | LVM/LVEDV | 0.6-1 | 1.1-1.2 | > 1.2 | 10 |  | |  |  |  |
|  | RVEDVi | 55-102 | 103-110 | > 110 | 10 |  | |  |  |  |
|  | RVESVi | 18-48 | 49-54 | > 54 | 10 |  | |  |  |  |
|  | RVEF | 50-72 | 48-49 | < 48 | 10 |  | |  |  |  |
|  | RVCOi | 2-3.8 | 1.8-1.9 | < 1.8 | 10 |  | |  |  |  |
|  | RVSVi | 33-59 | 28-32 | < 28 | 10 |  | |  |  |  |
|  | LAVi max |  |  |  |  |  | |  |  |  |
|  | LAEF |  |  |  |  |  | |  |  |  |
|  | RAVi max | 23-67 | 68-80 | > 80 | 10 |  | |  |  |  |
|  | RAEF | 30-65 | 25-29 | < 25 | 10 |  | |  |  |  |
| 50-59 | LVEDVi | 47-84 | 85-89 | > 89 | 95 | 44-98 | | 99-106 | > 106 | 78 |
|  | LVESVi | 12-32 | 33-35 | > 35 | 95 | 12-40 | | 41-45 | > 45 | 78 |
|  | LVSVi | 32-57 | 28-31 | < 28 | 95 | 30-62 | | 25-29 | < 25 | 76 |
|  | LVEF | 57-79 | 54-56 | < 54 | 95 | 55-77 | | 52-54 | < 52 | 76 |
|  | LVCOi | 1.9-3.9 | 1.7-1.8 | < 1.7 | 94 | 2.1-4.1 | | 1.9-2 | < 1.9 | 77 |
|  | LVMi | 40-66 | 67-72 | > 72 | 95 | 42-79 | | 80-84 | > 84 | 75 |
|  | LVM/LVEDV | 0.6-1.1 |  | > 1.2 | 95 | 0.7-1.1 | |  | > 1.2 | 75 |
|  | RVEDVi | 53-100 | 101-108 | > 108 | 95 | 65-111 | | 112-124 | > 124 | 75 |
|  | RVESVi | 17-46 | 47-51 | > 51 | 95 | 25-50 | | 51-55 | > 55 | 74 |
|  | RVEF | 50-72 | 47-49 | < 47 | 95 | 46-68 | | 43-45 | < 43 | 76 |
|  | RVCOi | 02-4 | 1.8-1.9 | < 1.8 | 94 | 1.9-4.5 | | 1.7-1.8 | < 1.7 | 78 |
|  | RVSVi | 32-59 | 28-31 | < 28 | 95 | 29-67 | | 24-28 | < 24 | 77 |
|  | LAVi max | 22-51 | 52-55 | > 55 | 89 | 20-53 | | 54-60 | > 60 | 64 |
|  | LAEF | 54-75 | 51-53 | < 51 | 88 | 53-76 | | 50-52 | < 50 | 64 |
|  | RAVi max | 22-63 | 64-74 | > 74 | 94 | 28-64 | | 65-70 | > 70 | 73 |
|  | RAEF | 31-65 | 26-30 | < 26 | 93 | 35-63 | | 32-34 | < 32 | 71 |
| 60-69 | LVEDVi | 42-81 | 82-88 | > 88 | 66 | 48-99 | | 100-108 | > 108 | 48 |
|  | LVESVi | 10-31 | 32-35 | > 35 | 66 | 10-41 | | 42-47 | > 47 | 48 |
|  | LVSVi | 29-56 | 25-28 | < 25 | 66 | 33-65 | | 30-32 | < 30 | 48 |
|  | LVEF | 57-80 | 53-56 | < 53 | 66 | 45-78 | | 34-44 | < 34 | 48 |
|  | LVCOi | 1.8-3.8 | 1.6-1.7 | < 1.6 | 66 | 2-4.2 | | 1.9-1.9 | < 1.9 | 48 |
|  | LVMi | 36-66 | 67-70 | > 70 | 66 | 43-80 | | 81-85 | > 85 | 48 |
|  | LVM/LVEDV | 0.7-1.1 | 1.2-1.3 | > 1.3 | 66 | 0.7-1.2 | | 1.3-1.3 | > 1.3 | 48 |
|  | RVEDVi | 50-97 | 98-106 | > 106 | 66 | 61-112 | | 113-125 | > 125 | 48 |
|  | RVESVi | 15-43 | 44-48 | > 48 | 66 | 22-53 | | 54-61 | > 61 | 48 |
|  | RVEF | 49-71 | 45-48 | < 45 | 64 | 46-69 | | 42-45 | < 42 | 48 |
|  | RVCOi | 1.9-4.1 | 1.7-1.8 | < 1.7 | 66 | 2.1-4.3 | | 1.9-2 | < 1.9 | 48 |
|  | RVSVi | 30-60 | 26-29 | < 26 | 66 | 33-66 | | 30-32 | < 30 | 48 |
|  | LAVi max | 20-55 | 56-61 | > 61 | 59 | 20-56 | | 57-64 | > 64 | 42 |
|  | LAEF | 50-75 | 45-49 | < 45 | 58 | 50-75 | | 46-49 | < 46 | 43 |
|  | RAVi max | 21-59 | 60-67 | > 67 | 64 | 24-69 | | 70-76 | > 76 | 45 |
|  | RAEF | 32-67 | 27-31 | < 27 | 66 | 32-64 | | 29-31 | < 29 | 46 |
| 70+ | LVEDVi | 38-80 | 81-88 | > 88 | 18 | 45-90 | | 91-99 | > 99 | 14 |
|  | LVESVi | 9-30 | 31-36 | > 36 | 18 | 7-37 | | 38-43 | > 43 | 14 |
|  | LVSVi | 26-56 | 22-25 | < 22 | 18 | 32-62 | | 30-31 | < 30 | 14 |
|  | LVEF | 56-81 | 49-55 | < 49 | 18 | 48-79 | | 39-47 | < 39 | 14 |
|  | LVCOi | 1.8-3.7 | 1.5-1.7 | < 1.5 | 18 | 2-4.5 | | 1.8-1.9 | < 1.8 | 14 |
|  | LVMi | 28-66 | 67-70 | > 70 | 18 | 41-76 | | 77-81 | > 81 | 14 |
|  | LVM/LVEDV | 0.7-1.2 | 1.3-1.4 | > 1.4 | 18 | 0.7-1.2 | | 1.3-1.3 | > 1.3 | 14 |
|  | RVEDVi | 47-95 | 96-103 | > 103 | 18 | 57-112 | | 113-125 | > 125 | 14 |
|  | RVESVi | 13-41 | 42-45 | > 45 | 18 | 20-58 | | 59-70 | > 70 | 14 |
|  | RVEF | 46-71 | 39-45 | < 39 | 18 | 45-70 | | 41-44 | < 41 | 14 |
|  | RVCOi | 2-3.9 | 1.8-1.9 | < 1.8 | 18 | 2.2-4.2 | | 2-2.1 | < 2 | 14 |
|  | RVSVi | 28-60 | 25-27 | < 25 | 18 | 35-65 | | 33-34 | < 33 | 14 |
|  | LAVi max | 19-61 | 62-73 | > 73 | 15 |  | |  |  |  |
|  | LAEF | 44-74 | 35-43 | < 35 | 16 |  | |  |  |  |
|  | RAVi max | 20-57 | 58-63 | > 63 | 17 | 19-74 | | 75-82 | > 82 | 12 |
|  | RAEF | 33-68 | 28-32 | < 28 | 17 | 29-66 | | 25-28 | < 25 | 12 |

### 38. Table: Reference ranges and centile-based categories for cardiac chamber measurements derived using cvi42 software (papillary segmentation), indexed to BSA, in an Asian adult population

|  |  | **Female** | | | | **Male** | | | |
| --- | --- | --- | --- | --- | --- | --- | --- | --- | --- |
| **Age group** | **Metrics** | **Reference range** | **Mild** | **At least moderate** | **n** | **Reference range** | **Mild** | **At least moderate** | **n** |
| 40-49 | LVEDVi | 52-81 | 82-88 | > 88 | 10 | 57-95 | 96-104 | > 104 | 20 |
|  | LVESVi | 17-31 | 32-35 | > 35 | 10 | 17-41 | 42-45 | > 45 | 20 |
|  | LVSVi | 31-53 | 28-30 | < 28 | 10 | 36-56 | 33-35 | < 33 | 20 |
|  | LVEF | 53-73 | 47-52 | < 47 | 10 | 54-77 | 52-53 | < 52 | 20 |
|  | LVCOi | 1.9-3.7 | 1.7-1.8 | < 1.7 | 10 | 2-4.1 | 1.8-1.9 | < 1.8 | 20 |
|  | LVMi | 32-47 | 48-51 | > 51 | 10 | 38-61 | 62-66 | > 66 | 20 |
|  | LVM/LVEDV | 0.5-0.8 | 0.9-0.9 | > 0.9 | 10 | 0.6-0.8 | 0.9-0.9 | > 0.9 | 20 |
|  | RVEDVi | 51-82 | 83-90 | > 90 | 10 | 59-105 | 106-117 | > 117 | 20 |
|  | RVESVi | 16-35 | 36-39 | > 39 | 10 | 21-46 | 47-50 | > 50 | 20 |
|  | RVEF | 52-72 | 48-51 | < 48 | 10 | 49-67 | 46-48 | < 46 | 20 |
|  | RVCOi | 2-3.5 | 1.8-1.9 | < 1.8 | 10 | 2-3.9 | 1.8-1.9 | < 1.8 | 20 |
|  | RVSVi | 30-52 | 28-29 | < 28 | 10 | 33-65 | 30-32 | < 30 | 20 |
|  | LAVi max | 21-45 | 46-49 | > 49 | 10 | 18-48 | 49-53 | > 53 | 20 |
|  | LAEF | 57-81 | 54-56 | < 54 | 10 | 58-84 | 55-57 | < 55 | 20 |
|  | RAVi max | 21-44 | 45-48 | > 48 | 10 | 21-62 | 63-71 | > 71 | 20 |
|  | RAEF | 37-71 | 35-36 | < 35 | 10 | 32-63 | 27-31 | < 27 | 20 |
| 50-59 | LVEDVi | 50-82 | 83-90 | > 90 | 83 | 54-94 | 95-103 | > 103 | 133 |
|  | LVESVi | 15-34 | 35-40 | > 40 | 83 | 16-40 | 41-45 | > 45 | 133 |
|  | LVSVi | 31-54 | 27-30 | < 27 | 83 | 31-61 | 28-30 | < 28 | 133 |
|  | LVEF | 53-74 | 48-52 | < 48 | 83 | 52-73 | 49-51 | < 49 | 131 |
|  | LVCOi | 1.9-3.7 | 1.7-1.8 | < 1.7 | 83 | 1.9-4 | 1.7-1.8 | < 1.7 | 133 |
|  | LVMi | 31-49 | 50-54 | > 54 | 83 | 37-64 | 65-71 | > 71 | 133 |
|  | LVM/LVEDV | 0.5-0.8 | 0.9-0.9 | > 0.9 | 83 | 0.6-0.9 | 01-1 | > 1 | 133 |
|  | RVEDVi | 49-85 | 86-94 | > 94 | 83 | 56-103 | 104-114 | > 114 | 133 |
|  | RVESVi | 16-36 | 37-40 | > 40 | 83 | 20-45 | 46-50 | > 50 | 133 |
|  | RVEF | 53-73 | 49-52 | < 49 | 83 | 48-68 | 45-47 | < 45 | 133 |
|  | RVCOi | 1.9-3.6 | 1.7-1.8 | < 1.7 | 83 | 1.9-3.9 | 1.7-1.8 | < 1.7 | 133 |
|  | RVSVi | 30-55 | 27-29 | < 27 | 83 | 31-62 | 27-30 | < 27 | 133 |
|  | LAVi max | 21-47 | 48-51 | > 51 | 77 | 19-50 | 51-55 | > 55 | 121 |
|  | LAEF | 54-80 | 51-53 | < 51 | 75 | 55-82 | 52-54 | < 52 | 120 |
|  | RAVi max | 21-50 | 51-55 | > 55 | 76 | 21-62 | 63-71 | > 71 | 128 |
|  | RAEF | 37-69 | 34-36 | < 34 | 76 | 34-65 | 29-33 | < 29 | 127 |
| 60-69 | LVEDVi | 47-85 | 86-95 | > 95 | 63 | 47-93 | 94-101 | > 101 | 118 |
|  | LVESVi | 13-36 | 37-44 | > 44 | 63 | 14-40 | 41-46 | > 46 | 118 |
|  | LVSVi | 30-56 | 27-29 | < 27 | 63 | 30-60 | 26-29 | < 26 | 118 |
|  | LVEF | 54-77 | 50-53 | < 50 | 63 | 51-74 | 46-50 | < 46 | 118 |
|  | LVCOi | 1.8-3.8 | 1.6-1.7 | < 1.6 | 63 | 1.8-4 | 1.5-1.7 | < 1.5 | 119 |
|  | LVMi | 31-54 | 55-61 | > 61 | 63 | 36-67 | 68-77 | > 77 | 119 |
|  | LVM/LVEDV | 0.5-0.8 | 0.9-0.9 | > 0.9 | 63 | 0.6-0.9 | 1-1.1 | > 1.1 | 118 |
|  | RVEDVi | 47-91 | 92-103 | > 103 | 63 | 49-100 | 101-109 | > 109 | 119 |
|  | RVESVi | 15-36 | 37-42 | > 42 | 63 | 18-45 | 46-51 | > 51 | 119 |
|  | RVEF | 54-73 | 51-53 | < 51 | 63 | 47-70 | 42-46 | < 42 | 119 |
|  | RVCOi | 1.8-3.9 | 1.6-1.7 | < 1.6 | 63 | 1.7-3.8 | 1.4-1.6 | < 1.4 | 119 |
|  | RVSVi | 29-58 | 26-28 | < 26 | 63 | 28-60 | 24-27 | < 24 | 119 |
|  | LAVi max | 21-50 | 51-55 | > 55 | 59 | 20-52 | 53-57 | > 57 | 110 |
|  | LAEF | 50-78 | 46-49 | < 46 | 58 | 51-80 | 48-50 | < 48 | 107 |
|  | RAVi max | 19-60 | 61-67 | > 67 | 61 | 21-62 | 63-71 | > 71 | 113 |
|  | RAEF | 36-68 | 31-35 | < 31 | 59 | 35-70 | 31-34 | < 31 | 114 |
| 70+ | LVEDVi | 44-88 | 89-100 | > 100 | 28 | 39-91 | 92-99 | > 99 | 41 |
|  | LVESVi | 13-32 | 33-37 | > 37 | 28 | 13-40 | 41-47 | > 47 | 41 |
|  | LVSVi | 29-62 | 26-28 | < 26 | 28 | 29-59 | 25-28 | < 25 | 40 |
|  | LVEF | 55-79 | 51-54 | < 51 | 28 | 52-76 | 47-51 | < 47 | 41 |
|  | LVCOi | 1.7-4.2 | 1.5-1.6 | < 1.5 | 28 | 1.5-4 | 1.1-1.4 | < 1.1 | 41 |
|  | LVMi | 30-59 | 60-70 | > 70 | 28 | 31-63 | 64-68 | > 68 | 41 |
|  | LVM/LVEDV | 0.5-0.8 | 0.9-0.9 | > 0.9 | 28 | 0.6-1.1 | 1.2-1.3 | > 1.3 | 41 |
|  | RVEDVi | 45-98 | 99-114 | > 114 | 28 | 41-97 | 98-104 | > 104 | 41 |
|  | RVESVi | 14-38 | 39-44 | > 44 | 28 | 16-45 | 46-52 | > 52 | 41 |
|  | RVEF | 55-74 | 52-54 | < 52 | 28 | 46-72 | 39-45 | < 39 | 40 |
|  | RVCOi | 1.6-4.1 | 1.4-1.5 | < 1.4 | 28 | 1.2-4 | 0.7-1.1 | < 0.7 | 41 |
|  | RVSVi | 28-63 | 25-27 | < 25 | 28 | 26-60 | 19-25 | < 19 | 40 |
|  | LAVi max | 21-54 | 55-61 | > 61 | 28 | 21-53 | 54-59 | > 59 | 34 |
|  | LAEF | 45-76 | 40-44 | < 40 | 27 | 48-79 | 45-47 | < 45 | 34 |
|  | RAVi max | 15-66 | 67-77 | > 77 | 27 | 21-62 | 63-71 | > 71 | 38 |
|  | RAEF | 35-70 | 25-34 | < 25 | 27 | 36-77 | 32-35 | < 32 | 39 |

### 39. Table: Reference ranges and centile-based categories for cardiac chamber measurements derived using cvi42 software (papillary segmentation), indexed to BSA, in a Chinese adult population

|  |  | **Female** | | | | **Male** | | | |
| --- | --- | --- | --- | --- | --- | --- | --- | --- | --- |
| **Age group** | **Metrics** | **Reference range** | **Mild** | **At least moderate** | **n** | **Reference range** | **Mild** | **At least moderate** | **n** |
| 18-29 | LVEDVi | 59-98 | 99-107 | > 107 | 16 | 65-98 | 99-103 | > 103 | 19 |
|  | LVESVi | 17-41 | 42-48 | > 48 | 16 | 22-41 | 42-44 | > 44 | 19 |
|  | LVSVi | 36-70 | 34-35 | < 34 | 16 | 36-61 | 29-35 | < 29 | 19 |
|  | LVEF | 54-76 | 51-53 | < 51 | 16 | 53-70 | 51-52 | < 51 | 19 |
|  | LVCOi | 2.3-4.7 | 2.1-2.2 | < 2.1 | 16 | 2.8-4.5 | 2.6-2.7 | < 2.6 | 19 |
|  | LVMi | 30-51 | 52-56 | > 56 | 16 | 40-63 | 64-66 | > 66 | 19 |
|  | LVM/LVEDV | 0.4-0.6 |  | > 0.6 | 16 | 0.5-0.8 | 0.9-0.9 | > 0.9 | 19 |
|  | RVEDVi | 54-95 | 96-103 | > 103 | 16 | 61-105 | 106-112 | > 112 | 19 |
|  | RVESVi | 16-44 | 45-50 | > 50 | 16 | 18-46 | 47-48 | > 48 | 19 |
|  | RVEF | 49-78 | 46-48 | < 46 | 16 | 47-70 | 43-46 | < 43 | 19 |
|  | RVCOi | 1.8-4.2 | 1.5-1.7 | < 1.5 | 16 | 2.3-4.4 | 2-2.2 | < 2 | 19 |
|  | RVSVi | 32-61 | 28-31 | < 28 | 16 | 34-63 | 28-33 | < 28 | 19 |
|  | LAVi max | 23-53 | 54-59 | > 59 | 13 | 25-47 | 48-51 | > 51 | 16 |
|  | LAEF | 54-77 | 50-53 | < 50 | 13 | 52-70 | 49-51 | < 49 | 16 |
|  | RAVi max | 21-45 | 46-53 | > 53 | 15 | 18-58 | 59-65 | > 65 | 18 |
|  | RAEF | 41-73 | 35-40 | < 35 | 15 | 36-68 | 30-35 | < 30 | 18 |
| 30-39 | LVEDVi | 57-95 | 96-104 | > 104 | 18 | 62-97 | 98-103 | > 103 | 17 |
|  | LVESVi | 16-39 | 40-45 | > 45 | 18 | 19-39 | 40-42 | > 42 | 17 |
|  | LVSVi | 36-65 | 33-35 | < 33 | 18 | 35-60 | 29-34 | < 29 | 17 |
|  | LVEF | 55-76 | 51-54 | < 51 | 18 | 53-70 | 50-52 | < 50 | 17 |
|  | LVCOi | 2.2-4.6 | 2-2.1 | < 2 | 18 | 2.6-4.4 | 2.4-2.5 | < 2.4 | 17 |
|  | LVMi | 31-52 | 53-57 | > 57 | 18 | 40-64 | 65-68 | > 68 | 17 |
|  | LVM/LVEDV | 0.4-0.6 | 0.7-0.7 | > 0.7 | 18 | 0.5-0.8 | 0.9-0.9 | > 0.9 | 17 |
|  | RVEDVi | 53-94 | 95-102 | > 102 | 18 | 59-103 | 104-111 | > 111 | 17 |
|  | RVESVi | 15-42 | 43-48 | > 48 | 18 | 18-45 | 46-48 | > 48 | 17 |
|  | RVEF | 50-77 | 46-49 | < 46 | 18 | 47-70 | 44-46 | < 44 | 17 |
|  | RVCOi | 1.9-4.1 | 1.6-1.8 | < 1.6 | 18 | 2.1-4.4 | 1.9-2 | < 1.9 | 17 |
|  | RVSVi | 32-60 | 28-31 | < 28 | 18 | 32-63 | 27-31 | < 27 | 17 |
|  | LAVi max | 23-52 | 53-58 | > 58 | 18 | 24-47 | 48-51 | > 51 | 14 |
|  | LAEF | 53-76 | 50-52 | < 50 | 18 | 51-72 | 48-50 | < 48 | 14 |
|  | RAVi max | 22-49 | 50-57 | > 57 | 18 | 18-59 | 60-67 | > 67 | 16 |
|  | RAEF | 41-72 | 35-40 | < 35 | 18 | 36-69 | 31-35 | < 31 | 16 |
| 40-49 | LVEDVi | 55-92 | 93-100 | > 100 | 24 | 58-96 | 97-102 | > 102 | 20 |
|  | LVESVi | 15-37 | 38-42 | > 42 | 24 | 17-39 | 40-41 | > 41 | 20 |
|  | LVSVi | 35-62 | 33-34 | < 33 | 24 | 34-60 | 29-33 | < 29 | 20 |
|  | LVEF | 55-77 | 51-54 | < 51 | 24 | 53-71 | 50-52 | < 50 | 19 |
|  | LVCOi | 2.1-4.5 | 1.9-2 | < 1.9 | 24 | 2.4-4.4 | 2.2-2.3 | < 2.2 | 20 |
|  | LVMi | 32-53 | 54-59 | > 59 | 24 | 40-65 | 66-70 | > 70 | 20 |
|  | LVM/LVEDV | 0.5-0.7 |  | > 0.7 | 24 | 0.5-0.9 | 01-1 | > 1 | 20 |
|  | RVEDVi | 52-92 | 93-101 | > 101 | 24 | 58-102 | 103-109 | > 109 | 20 |
|  | RVESVi | 15-40 | 41-45 | > 45 | 24 | 17-45 | 46-48 | > 48 | 20 |
|  | RVEF | 51-75 | 47-50 | < 47 | 24 | 48-70 | 45-47 | < 45 | 19 |
|  | RVCOi | 1.9-3.9 | 1.7-1.8 | < 1.7 | 24 | 2-4.3 | 1.7-1.9 | < 1.7 | 20 |
|  | RVSVi | 31-59 | 28-30 | < 28 | 24 | 30-62 | 25-29 | < 25 | 20 |
|  | LAVi max | 22-51 | 52-57 | > 57 | 22 | 22-47 | 48-51 | > 51 | 18 |
|  | LAEF | 53-75 | 49-52 | < 49 | 22 | 51-74 | 47-50 | < 47 | 18 |
|  | RAVi max | 22-53 | 54-63 | > 63 | 23 | 19-61 | 62-69 | > 69 | 19 |
|  | RAEF | 41-70 | 36-40 | < 36 | 23 | 35-69 | 31-34 | < 31 | 19 |
| 50-59 | LVEDVi | 53-88 | 89-97 | > 97 | 66 | 54-95 | 96-102 | > 102 | 49 |
|  | LVESVi | 14-34 | 35-39 | > 39 | 66 | 14-39 | 40-42 | > 42 | 49 |
|  | LVSVi | 34-60 | 32-33 | < 32 | 66 | 33-60 | 29-32 | < 29 | 49 |
|  | LVEF | 55-77 | 51-54 | < 51 | 66 | 53-72 | 50-52 | < 50 | 48 |
|  | LVCOi | 2.1-4.1 | 02-2 | < 2 | 66 | 2.2-4.3 | 2-2.1 | < 2 | 49 |
|  | LVMi | 33-54 | 55-60 | > 60 | 66 | 39-67 | 68-73 | > 73 | 49 |
|  | LVM/LVEDV | 0.5-0.8 |  | > 0.8 | 66 | 0.5-0.9 | 01-1 | > 1 | 49 |
|  | RVEDVi | 51-90 | 91-99 | > 99 | 66 | 56-101 | 102-108 | > 108 | 49 |
|  | RVESVi | 15-38 | 39-43 | > 43 | 66 | 17-45 | 46-48 | > 48 | 49 |
|  | RVEF | 52-74 | 48-51 | < 48 | 66 | 48-70 | 46-47 | < 46 | 49 |
|  | RVCOi | 1.9-3.8 |  | < 1.8 | 66 | 1.9-4.2 | 1.6-1.8 | < 1.6 | 49 |
|  | RVSVi | 31-58 | 28-30 | < 28 | 66 | 29-62 | 23-28 | < 23 | 49 |
|  | LAVi max | 22-50 | 51-56 | > 56 | 60 | 21-46 | 47-50 | > 50 | 40 |
|  | LAEF | 51-76 | 48-50 | < 48 | 58 | 50-77 | 46-49 | < 46 | 40 |
|  | RAVi max | 23-58 | 59-69 | > 69 | 63 | 20-63 | 64-71 | > 71 | 46 |
|  | RAEF | 40-68 | 36-39 | < 36 | 63 | 35-71 | 31-34 | < 31 | 46 |
| 60-69 | LVEDVi | 51-85 | 86-93 | > 93 | 52 | 51-94 | 95-102 | > 102 | 37 |
|  | LVESVi | 13-32 | 33-37 | > 37 | 52 | 12-38 | 39-41 | > 41 | 37 |
|  | LVSVi | 33-59 | 29-32 | < 29 | 52 | 32-61 | 29-31 | < 29 | 37 |
|  | LVEF | 55-78 | 50-54 | < 50 | 52 | 53-73 | 50-52 | < 50 | 35 |
|  | LVCOi | 2.1-3.8 | 02-2 | < 2 | 52 | 1.9-4.2 | 1.7-1.8 | < 1.7 | 37 |
|  | LVMi | 34-55 | 56-62 | > 62 | 52 | 39-69 | 70-80 | > 80 | 37 |
|  | LVM/LVEDV | 0.5-0.8 | 0.9-1 | > 1 | 52 | 0.6-1 | 1.1-1.1 | > 1.1 | 37 |
|  | RVEDVi | 49-88 | 89-96 | > 96 | 52 | 54-99 | 100-107 | > 107 | 36 |
|  | RVESVi | 14-36 | 37-40 | > 40 | 52 | 16-44 | 45-48 | > 48 | 37 |
|  | RVEF | 53-74 | 49-52 | < 49 | 52 | 49-70 | 46-48 | < 46 | 36 |
|  | RVCOi | 2-3.7 | 1.8-1.9 | < 1.8 | 52 | 1.7-4.2 | 1.5-1.6 | < 1.5 | 37 |
|  | RVSVi | 31-57 | 28-30 | < 28 | 52 | 27-61 | 22-26 | < 22 | 37 |
|  | LAVi max | 21-49 | 50-55 | > 55 | 47 | 19-46 | 47-50 | > 50 | 30 |
|  | LAEF | 49-76 | 45-48 | < 45 | 46 | 48-80 | 44-47 | < 44 | 30 |
|  | RAVi max | 23-63 | 64-76 | > 76 | 46 | 20-65 | 66-73 | > 73 | 35 |
|  | RAEF | 40-66 | 36-39 | < 36 | 46 | 35-72 | 32-34 | < 32 | 35 |
| 70+ | LVEDVi | 50-83 | 84-90 | > 90 | 12 |  |  |  |  |
|  | LVESVi | 12-31 | 32-35 | > 35 | 12 |  |  |  |  |
|  | LVSVi | 30-59 | 25-29 | < 25 | 12 |  |  |  |  |
|  | LVEF | 55-79 | 50-54 | < 50 | 12 |  |  |  |  |
|  | LVCOi | 2-3.9 | 1.8-1.9 | < 1.8 | 12 |  |  |  |  |
|  | LVMi | 35-56 | 57-63 | > 63 | 12 |  |  |  |  |
|  | LVM/LVEDV | 0.5-1 | 1.1-1.3 | > 1.3 | 12 |  |  |  |  |
|  | RVEDVi | 47-86 | 87-93 | > 93 | 12 |  |  |  |  |
|  | RVESVi | 14-34 | 35-38 | > 38 | 12 |  |  |  |  |
|  | RVEF | 54-74 | 50-53 | < 50 | 12 |  |  |  |  |
|  | RVCOi | 2-3.6 | 1.8-1.9 | < 1.8 | 12 |  |  |  |  |
|  | RVSVi | 31-56 | 28-30 | < 28 | 12 |  |  |  |  |
|  | LAVi max | 21-48 | 49-53 | > 53 | 11 |  |  |  |  |
|  | LAEF | 47-78 | 42-46 | < 42 | 12 |  |  |  |  |
|  | RAVi max | 23-68 | 69-82 | > 82 | 11 |  |  |  |  |
|  | RAEF | 39-65 | 36-38 | < 36 | 11 |  |  |  |  |

### 40. Table: Reference ranges and centile-based categories for cardiac chamber measurements derived using cvi42 software (papillary segmentation), indexed to BSA, in a Mixed adult population

|  |  | **Female** | | | | **Male** | | | |
| --- | --- | --- | --- | --- | --- | --- | --- | --- | --- |
| **Age group** | **Metrics** | **Reference range** | **Mild** | **At least moderate** | **n** | **Reference range** | **Mild** | **At least moderate** | **n** |
| 50-59 | LVEDVi | 53-93 | 94-101 | > 101 | 134 | 59-104 | 105-111 | > 111 | 65 |
|  | LVESVi | 15-35 | 36-39 | > 39 | 134 | 16-42 | 43-45 | > 45 | 66 |
|  | LVSVi | 33-64 | 30-32 | < 30 | 133 | 36-69 | 32-35 | < 32 | 65 |
|  | LVEF | 54-76 | 51-53 | < 51 | 134 | 52-74 | 48-51 | < 48 | 66 |
|  | LVCOi | 2-4.1 | 1.8-1.9 | < 1.8 | 132 | 2.1-4.6 | 1.9-2 | < 1.9 | 66 |
|  | LVMi | 32-55 | 56-60 | > 60 | 134 | 41-73 | 74-80 | > 80 | 65 |
|  | LVM/LVEDV | 0.5-0.8 |  | > 0.8 | 134 | 0.5-0.9 | 1-1.1 | > 1.1 | 64 |
|  | RVEDVi | 53-98 | 99-107 | > 107 | 134 | 62-120 | 121-132 | > 132 | 65 |
|  | RVESVi | 17-39 | 40-44 | > 44 | 134 | 22-48 | 49-52 | > 52 | 65 |
|  | RVEF | 53-72 | 50-52 | < 50 | 134 | 51-70 | 48-50 | < 48 | 66 |
|  | RVCOi | 1.9-4.1 | 1.7-1.8 | < 1.7 | 133 | 2.2-4.6 | 2-2.1 | < 2 | 66 |
|  | RVSVi | 32-60 | 28-31 | < 28 | 133 | 36-73 | 32-35 | < 32 | 66 |
|  | LAVi max | 21-48 | 49-51 | > 51 | 121 | 24-57 | 58-64 | > 64 | 53 |
|  | LAEF | 55-78 | 52-54 | < 52 | 120 | 54-78 | 51-53 | < 51 | 53 |
|  | RAVi max | 21-63 | 64-73 | > 73 | 128 | 21-68 | 69-76 | > 76 | 64 |
|  | RAEF | 37-64 | 31-36 | < 31 | 124 | 35-63 | 31-34 | < 31 | 64 |
| 60-69 | LVEDVi | 48-89 | 90-96 | > 96 | 110 | 52-98 | 99-106 | > 106 | 59 |
|  | LVESVi | 12-34 | 35-38 | > 38 | 110 | 15-42 | 43-47 | > 47 | 58 |
|  | LVSVi | 31-59 | 27-30 | < 27 | 110 | 33-63 | 29-32 | < 29 | 59 |
|  | LVEF | 54-76 | 50-53 | < 50 | 108 | 51-74 | 47-50 | < 47 | 58 |
|  | LVCOi | 02-4 | 1.8-1.9 | < 1.8 | 110 | 2-4.3 | 1.8-1.9 | < 1.8 | 59 |
|  | LVMi | 31-54 | 55-58 | > 58 | 110 | 39-71 | 72-78 | > 78 | 59 |
|  | LVM/LVEDV | 0.5-0.8 | 0.9-0.9 | > 0.9 | 110 | 0.5-1 | 1.1-1.1 | > 1.1 | 59 |
|  | RVEDVi | 47-93 | 94-100 | > 100 | 110 | 55-113 | 114-124 | > 124 | 58 |
|  | RVESVi | 16-38 | 39-42 | > 42 | 110 | 17-48 | 49-52 | > 52 | 58 |
|  | RVEF | 53-72 | 49-52 | < 49 | 110 | 50-69 | 47-49 | < 47 | 58 |
|  | RVCOi | 1.9-4 | 1.7-1.8 | < 1.7 | 110 | 2-4.4 | 1.8-1.9 | < 1.8 | 59 |
|  | RVSVi | 28-60 | 24-27 | < 24 | 110 | 32-68 | 27-31 | < 27 | 59 |
|  | LAVi max | 22-51 | 52-56 | > 56 | 100 | 21-56 | 57-63 | > 63 | 52 |
|  | LAEF | 52-79 | 49-51 | < 49 | 98 | 50-79 | 47-49 | < 47 | 52 |
|  | RAVi max | 20-62 | 63-70 | > 70 | 103 | 18-71 | 72-79 | > 79 | 57 |
|  | RAEF | 35-68 | 29-34 | < 29 | 103 | 36-68 | 33-35 | < 33 | 57 |
| 70+ | LVEDVi | 46-82 | 83-87 | > 87 | 51 | 46-94 | 95-101 | > 101 | 33 |
|  | LVESVi | 9-33 | 34-37 | > 37 | 51 | 14-45 | 46-52 | > 52 | 34 |
|  | LVSVi | 28-54 | 24-27 | < 24 | 50 | 30-59 | 25-29 | < 25 | 34 |
|  | LVEF | 54-76 | 49-53 | < 49 | 51 | 50-73 | 47-49 | < 47 | 34 |
|  | LVCOi | 1.9-4 |  | < 1.8 | 51 | 1.9-3.9 | 1.7-1.8 | < 1.7 | 35 |
|  | LVMi | 31-52 | 53-56 | > 56 | 51 | 38-70 | 71-76 | > 76 | 35 |
|  | LVM/LVEDV | 0.5-0.8 | 0.9-0.9 | > 0.9 | 51 | 0.5-1 | 1.1-1.2 | > 1.2 | 33 |
|  | RVEDVi | 51-81 | 82-85 | > 85 | 51 | 49-107 | 108-117 | > 117 | 35 |
|  | RVESVi | 16-35 | 36-38 | > 38 | 51 | 13-47 | 48-52 | > 52 | 35 |
|  | RVEF | 52-72 | 48-51 | < 48 | 51 | 49-69 | 46-48 | < 46 | 35 |
|  | RVCOi | 1.9-3.8 | 1.7-1.8 | < 1.7 | 51 | 1.8-4.1 | 1.6-1.7 | < 1.6 | 35 |
|  | RVSVi | 31-54 | 27-30 | < 27 | 50 | 28-64 | 22-27 | < 22 | 35 |
|  | LAVi max | 22-55 | 56-61 | > 61 | 45 | 18-55 | 56-63 | > 63 | 32 |
|  | LAEF | 50-80 | 47-49 | < 47 | 44 | 46-81 | 42-45 | < 42 | 30 |
|  | RAVi max | 19-61 | 62-68 | > 68 | 47 | 17-72 | 73-81 | > 81 | 31 |
|  | RAEF | 33-72 | 28-32 | < 28 | 46 | 36-76 | 34-35 | < 34 | 31 |

### 41. Table: Reference ranges and centile-based categories for cardiac chamber measurements derived using suiteHEART software (smooth segmentation), indexed to BSA, in a White adult population

|  |  | **Female** | | | | **Male** | | | |
| --- | --- | --- | --- | --- | --- | --- | --- | --- | --- |
| **Age group** | **Metrics** | **Reference range** | **Mild** | **At least moderate** | **n** | **Reference range** | **Mild** | **At least moderate** | **n** |
| 18-29 | LVEDVi | 63-109 | 110-116 | > 116 | 101 | 73-122 | 123-128 | > 128 | 95 |
|  | LVESVi | 24-48 | 49-53 | > 53 | 101 | 28-52 | 53-56 | > 56 | 84 |
|  | LVSVi | 36-66 | 31-35 | < 31 | 101 | 40-73 | 35-39 | < 35 | 100 |
|  | LVEF | 50-68 | 47-49 | < 47 | 101 | 47-67 | 44-46 | < 44 | 104 |
|  | LVMi | 34-55 | 56-60 | > 60 | 101 | 45-77 | 78-85 | > 85 | 104 |
|  | LVM/LVEDV | 0.4-0.7 | 0.8-0.8 | > 0.8 | 101 | 0.5-0.8 | 0.9-1 | > 1 | 95 |
|  | RVEDVi | 60-119 | 120-130 | > 130 | 101 | 76-134 | 135-145 | > 145 | 93 |
|  | RVESVi | 25-55 | 56-62 | > 62 | 100 | 29-59 | 60-64 | > 64 | 90 |
|  | RVEF | 47-68 | 44-46 | < 44 | 99 | 45-66 | 42-44 | < 42 | 102 |
|  | RVSVi | 34-74 | 30-33 | < 30 | 100 | 38-77 | 34-37 | < 34 | 97 |
|  | LAESV | 18-42 | 43-46 | > 46 | 65 | 19-50 | 51-57 | > 57 | 64 |
|  | LAEF | 57-85 | 52-56 | < 52 | 65 | 55-79 | 51-54 | < 51 | 62 |
|  | RAESV | 23-48 | 49-52 | > 52 | 77 | 24-60 | 61-66 | > 66 | 87 |
|  | RAEF | 42-75 | 38-41 | < 38 | 72 | 35-71 | 31-34 | < 31 | 85 |
| 30-39 | LVEDVi | 60-106 | 107-114 | > 114 | 131 | 64-111 | 112-118 | > 118 | 127 |
|  | LVESVi | 22-47 | 48-52 | > 52 | 131 | 25-50 | 51-54 | > 54 | 123 |
|  | LVSVi | 34-64 | 30-33 | < 30 | 130 | 35-67 | 31-34 | < 31 | 128 |
|  | LVEF | 50-68 | 47-49 | < 47 | 131 | 48-68 | 45-47 | < 45 | 129 |
|  | LVMi | 34-57 | 58-62 | > 62 | 131 | 44-76 | 77-83 | > 83 | 128 |
|  | LVM/LVEDV | 0.4-0.8 | 0.9-0.9 | > 0.9 | 131 | 0.5-0.9 | 01-1 | > 1 | 128 |
|  | RVEDVi | 58-113 | 114-124 | > 124 | 131 | 68-124 | 125-134 | > 134 | 125 |
|  | RVESVi | 23-51 | 52-58 | > 58 | 131 | 27-57 | 58-61 | > 61 | 126 |
|  | RVEF | 48-69 | 45-47 | < 45 | 131 | 45-67 | 42-44 | < 42 | 128 |
|  | RVSVi | 32-70 | 29-31 | < 29 | 131 | 35-73 | 30-34 | < 30 | 125 |
|  | LAESV | 19-45 | 46-50 | > 50 | 95 | 18-49 | 50-56 | > 56 | 86 |
|  | LAEF | 57-84 | 53-56 | < 53 | 95 | 55-80 | 51-54 | < 51 | 87 |
|  | RAESV | 23-50 | 51-55 | > 55 | 113 | 23-60 | 61-67 | > 67 | 104 |
|  | RAEF | 41-75 | 37-40 | < 37 | 108 | 36-72 | 31-35 | < 31 | 103 |
| 40-49 | LVEDVi | 58-105 | 106-113 | > 113 | 521 | 63-114 | 115-122 | > 122 | 423 |
|  | LVESVi | 20-45 | 46-50 | > 50 | 521 | 23-50 | 51-54 | > 54 | 422 |
|  | LVSVi | 34-63 | 29-33 | < 29 | 521 | 35-69 | 30-34 | < 30 | 430 |
|  | LVEF | 51-70 | 48-50 | < 48 | 520 | 49-69 | 46-48 | < 46 | 443 |
|  | LVMi | 34-57 | 58-62 | > 62 | 521 | 43-75 | 76-82 | > 82 | 441 |
|  | LVM/LVEDV | 0.4-0.8 | 0.9-0.9 | > 0.9 | 521 | 0.5-0.9 | 1-1.1 | > 1.1 | 420 |
|  | RVEDVi | 56-108 | 109-118 | > 118 | 521 | 67-126 | 127-136 | > 136 | 428 |
|  | RVESVi | 21-47 | 48-53 | > 53 | 521 | 25-56 | 57-61 | > 61 | 422 |
|  | RVEF | 49-71 | 46-48 | < 46 | 518 | 45-69 | 42-44 | < 42 | 442 |
|  | RVSVi | 32-68 | 28-31 | < 28 | 519 | 34-74 | 29-33 | < 29 | 423 |
|  | LAESV | 19-48 | 49-53 | > 53 | 453 | 18-49 | 50-56 | > 56 | 374 |
|  | LAEF | 57-83 | 53-56 | < 53 | 453 | 55-81 | 51-54 | < 51 | 374 |
|  | RAESV | 22-52 | 53-58 | > 58 | 452 | 23-64 | 65-71 | > 71 | 362 |
|  | RAEF | 41-75 | 36-40 | < 36 | 445 | 36-73 | 31-35 | < 31 | 360 |
| 50-59 | LVEDVi | 55-100 | 101-108 | > 108 | 912 | 58-110 | 111-118 | > 118 | 805 |
|  | LVESVi | 18-42 | 43-47 | > 47 | 911 | 20-47 | 48-52 | > 52 | 801 |
|  | LVSVi | 33-63 | 29-32 | < 29 | 912 | 34-68 | 29-33 | < 29 | 820 |
|  | LVEF | 53-72 | 49-52 | < 49 | 912 | 50-71 | 47-49 | < 47 | 833 |
|  | LVMi | 33-57 | 58-62 | > 62 | 912 | 42-73 | 74-81 | > 81 | 825 |
|  | LVM/LVEDV | 0.4-0.8 | 0.9-1 | > 1 | 912 | 0.5-1 | 1.1-1.1 | > 1.1 | 795 |
|  | RVEDVi | 54-103 | 104-113 | > 113 | 910 | 60-116 | 117-126 | > 126 | 811 |
|  | RVESVi | 18-43 | 44-49 | > 49 | 910 | 22-53 | 54-58 | > 58 | 804 |
|  | RVEF | 50-72 | 46-49 | < 46 | 909 | 46-70 | 42-45 | < 42 | 823 |
|  | RVSVi | 31-65 | 27-30 | < 27 | 906 | 31-71 | 26-30 | < 26 | 818 |
|  | LAESV | 19-48 | 49-54 | > 54 | 825 | 17-49 | 50-56 | > 56 | 700 |
|  | LAEF | 56-83 | 51-55 | < 51 | 823 | 54-82 | 50-53 | < 50 | 703 |
|  | RAESV | 21-54 | 55-61 | > 61 | 769 | 21-60 | 61-68 | > 68 | 604 |
|  | RAEF | 40-75 | 35-39 | < 35 | 760 | 36-74 | 31-35 | < 31 | 604 |
| 60-69 | LVEDVi | 51-96 | 97-105 | > 105 | 920 | 55-108 | 109-118 | > 118 | 789 |
|  | LVESVi | 16-39 | 40-45 | > 45 | 918 | 19-46 | 47-51 | > 51 | 785 |
|  | LVSVi | 32-61 | 28-31 | < 28 | 920 | 32-67 | 27-31 | < 27 | 792 |
|  | LVEF | 53-73 | 49-52 | < 49 | 917 | 50-71 | 47-49 | < 47 | 796 |
|  | LVMi | 33-58 | 59-64 | > 64 | 920 | 41-72 | 73-79 | > 79 | 798 |
|  | LVM/LVEDV | 0.5-0.9 | 01-1 | > 1 | 920 | 0.5-1 | 1.1-1.2 | > 1.2 | 781 |
|  | RVEDVi | 51-99 | 100-109 | > 109 | 918 | 57-114 | 115-123 | > 123 | 791 |
|  | RVESVi | 16-41 | 42-47 | > 47 | 919 | 20-51 | 52-57 | > 57 | 788 |
|  | RVEF | 51-74 | 46-50 | < 46 | 919 | 46-71 | 42-45 | < 42 | 794 |
|  | RVSVi | 30-64 | 26-29 | < 26 | 916 | 30-70 | 24-29 | < 24 | 793 |
|  | LAESV | 18-49 | 50-56 | > 56 | 809 | 16-49 | 50-57 | > 57 | 652 |
|  | LAEF | 54-83 | 48-53 | < 48 | 807 | 53-83 | 48-52 | < 48 | 654 |
|  | RAESV | 20-57 | 58-66 | > 66 | 721 | 21-63 | 64-72 | > 72 | 521 |
|  | RAEF | 41-75 | 35-40 | < 35 | 699 | 37-74 | 32-36 | < 32 | 496 |
| 70+ | LVEDVi | 50-95 | 96-105 | > 105 | 1099 | 52-108 | 109-119 | > 119 | 1242 |
|  | LVESVi | 15-39 | 40-45 | > 45 | 1098 | 18-46 | 47-52 | > 52 | 1227 |
|  | LVSVi | 31-60 | 28-30 | < 28 | 1101 | 30-65 | 24-29 | < 24 | 1253 |
|  | LVEF | 53-73 | 49-52 | < 49 | 1098 | 50-71 | 46-49 | < 46 | 1241 |
|  | LVMi | 33-59 | 60-66 | > 66 | 1101 | 40-71 | 72-78 | > 78 | 1248 |
|  | LVM/LVEDV | 0.5-0.9 | 1-1.1 | > 1.1 | 1099 | 0.5-1 | 1.1-1.2 | > 1.2 | 1235 |
|  | RVEDVi | 50-99 | 100-110 | > 110 | 1100 | 55-112 | 113-122 | > 122 | 1242 |
|  | RVESVi | 15-40 | 41-47 | > 47 | 1100 | 20-51 | 52-57 | > 57 | 1239 |
|  | RVEF | 50-75 | 44-49 | < 44 | 1096 | 45-72 | 41-44 | < 41 | 1228 |
|  | RVSVi | 29-64 | 24-28 | < 24 | 1097 | 28-69 | 21-27 | < 21 | 1244 |
|  | LAESV | 17-52 | 53-61 | > 61 | 947 | 15-50 | 51-58 | > 58 | 953 |
|  | LAEF | 50-83 | 44-49 | < 44 | 938 | 51-84 | 45-50 | < 45 | 943 |
|  | RAESV | 20-61 | 62-73 | > 73 | 769 | 22-66 | 67-76 | > 76 | 631 |
|  | RAEF | 42-77 | 37-41 | < 37 | 753 | 37-75 | 32-36 | < 32 | 619 |

Footnote: All measurements were derived using suiteHEART software with smooth endocardial segmentation; papillary muscles and trabeculations were excluded from left ventricular mass measurements. Volumetric and mass measurements were indexed to height. Reference ranges are defined as the interval between the 2.5th and 97.5th centiles derived from age-specific generalized additive models for location, scale and shape (GAMLSS) using the Box–Cox Cole–Green distribution. Centile-based categories beyond the reference range are derived from LMS-based thresholds (P0.5, P2.5, P97.5, and P99.5) and describe the position of a measurement within the reference distribution rather than disease severity or diagnostic abnormality. Reference ranges are based on a White adult population.

Abbreviations: LV = left ventricle; RV = right ventricle; LA = left atrium; RA = right atrium; EDV = end-diastolic volume; ESV = end-systolic volume; SV = stroke volume; EF = ejection fraction; CO = cardiac output.

### 42. Table: Reference ranges and centile-based categories for cardiac chamber measurements derived using suiteHEART software (smooth segmentation), indexed to BSA, in a Black adult population

|  |  | **Female** | | | | **Male** | | | |
| --- | --- | --- | --- | --- | --- | --- | --- | --- | --- |
| **Age group** | **Metrics** | **Reference range** | **Mild** | **At least moderate** | **n** | **Reference range** | **Mild** | **At least moderate** | **n** |
| 40-49 | LVEDVi | 55-98 | 99-104 | > 104 | 10 |  |  |  |  |
|  | LVESVi | 17-44 | 45-47 | > 47 | 10 |  |  |  |  |
|  | LVSVi | 34-61 | 29-33 | < 29 | 10 |  |  |  |  |
|  | LVEF | 52-72 | 50-51 | < 50 | 10 |  |  |  |  |
|  | LVMi | 35-63 | 64-76 | > 76 | 10 |  |  |  |  |
|  | LVM/LVEDV | 0.4-0.8 | 0.9-1 | > 1 | 10 |  |  |  |  |
|  | RVEDVi | 51-101 | 102-109 | > 109 | 10 |  |  |  |  |
|  | RVESVi | 19-47 | 48-54 | > 54 | 10 |  |  |  |  |
|  | RVEF | 47-69 | 43-46 | < 43 | 10 |  |  |  |  |
|  | RVSVi | 28-62 | 21-27 | < 21 | 10 |  |  |  |  |
|  | LAESVi |  |  |  |  |  |  |  |  |
|  | LAEF |  |  |  |  |  |  |  |  |
|  | RAESVi |  |  |  |  |  |  |  |  |
|  | RAEF |  |  |  |  |  |  |  |  |
| 50-59 | LVEDVi | 54-97 | 98-104 | > 104 | 95 | 59-111 | 112-123 | > 123 | 74 |
|  | LVESVi | 17-42 | 43-46 | > 46 | 95 | 21-47 | 48-53 | > 53 | 73 |
|  | LVSVi | 34-61 | 29-33 | < 29 | 95 | 36-68 | 32-35 | < 32 | 75 |
|  | LVEF | 52-73 | 50-51 | < 50 | 95 | 50-70 | 47-49 | < 47 | 76 |
|  | LVMi | 35-62 | 63-72 | > 72 | 95 | 42-77 | 78-87 | > 87 | 77 |
|  | LVM/LVEDV | 0.4-0.9 | 01-1 | > 1 | 95 | 0.5-0.9 | 01-1 | > 1 | 74 |
|  | RVEDVi | 53-101 | 102-110 | > 110 | 95 | 65-114 | 115-125 | > 125 | 77 |
|  | RVESVi | 18-47 | 48-54 | > 54 | 95 | 25-50 | 51-58 | > 58 | 74 |
|  | RVEF | 48-71 | 43-47 | < 43 | 95 | 47-69 | 43-46 | < 43 | 75 |
|  | RVSVi | 30-62 | 25-29 | < 25 | 95 | 34-68 | 29-33 | < 29 | 76 |
|  | LAESVi | 20-45 | 46-49 | > 49 | 88 | 18-46 | 47-53 | > 53 | 67 |
|  | LAEF | 59-79 | 56-58 | < 56 | 88 | 56-83 | 53-55 | < 53 | 67 |
|  | RAESVi | 20-58 | 59-66 | > 66 | 84 | 24-57 | 58-62 | > 62 | 64 |
|  | RAEF | 38-69 | 32-37 | < 32 | 84 | 38-65 | 35-37 | < 35 | 63 |
| 60-69 | LVEDVi | 53-96 | 97-104 | > 104 | 65 | 60-114 | 115-127 | > 127 | 48 |
|  | LVESVi | 16-40 | 41-45 | > 45 | 65 | 19-51 | 52-57 | > 57 | 47 |
|  | LVSVi | 33-61 | 29-32 | < 29 | 65 | 35-67 | 32-34 | < 32 | 48 |
|  | LVEF | 53-73 | 49-52 | < 49 | 65 | 49-71 | 44-48 | < 44 | 48 |
|  | LVMi | 37-63 | 64-70 | > 70 | 65 | 42-76 | 77-87 | > 87 | 48 |
|  | LVM/LVEDV | 0.5-0.9 | 1-1.1 | > 1.1 | 65 | 0.5-1 | 1.1-1.1 | > 1.1 | 48 |
|  | RVEDVi | 55-101 | 102-113 | > 113 | 65 | 61-112 | 113-123 | > 123 | 48 |
|  | RVESVi | 17-46 | 47-54 | > 54 | 65 | 21-52 | 53-61 | > 61 | 47 |
|  | RVEF | 48-74 | 43-47 | < 43 | 64 | 46-73 | 42-45 | < 42 | 48 |
|  | RVSVi | 32-63 | 28-31 | < 28 | 65 | 32-67 | 26-31 | < 26 | 48 |
|  | LAESVi | 18-49 | 50-55 | > 55 | 60 | 17-50 | 51-58 | > 58 | 42 |
|  | LAEF | 54-81 | 49-53 | < 49 | 61 | 52-85 | 48-51 | < 48 | 42 |
|  | RAESVi | 18-51 | 52-55 | > 55 | 52 | 21-62 | 63-68 | > 68 | 40 |
|  | RAEF | 32-66 | 21-31 | < 21 | 53 | 36-71 | 32-35 | < 32 | 41 |
| 70+ | LVEDVi | 51-96 | 97-110 | > 110 | 18 | 54-104 | 105-115 | > 115 | 14 |
|  | LVESVi | 16-41 | 42-48 | > 48 | 18 | 15-45 | 46-52 | > 52 | 14 |
|  | LVSVi | 32-61 | 29-31 | < 29 | 18 | 35-66 | 32-34 | < 32 | 14 |
|  | LVEF | 52-73 | 45-51 | < 45 | 18 | 46-72 | 38-45 | < 38 | 14 |
|  | LVMi | 38-64 | 65-69 | > 69 | 18 | 43-76 | 77-87 | > 87 | 14 |
|  | LVM/LVEDV | 0.5-1 | 1.1-1.1 | > 1.1 | 18 | 0.5-1.1 | 1.2-1.3 | > 1.3 | 14 |
|  | RVEDVi | 56-102 | 103-117 | > 117 | 18 | 56-111 | 112-121 | > 121 | 14 |
|  | RVESVi | 16-46 | 47-54 | > 54 | 18 | 15-53 | 54-62 | > 62 | 14 |
|  | RVEF | 48-76 | 43-47 | < 43 | 18 | 45-79 | 42-44 | < 42 | 14 |
|  | RVSVi | 32-67 | 29-31 | < 29 | 18 | 30-66 | 23-29 | < 23 | 14 |
|  | LAESVi | 15-55 | 56-64 | > 64 | 16 |  |  |  |  |
|  | LAEF | 47-82 | 39-46 | < 39 | 17 |  |  |  |  |
|  | RAESVi | 18-48 | 49-50 | > 50 | 13 |  |  |  |  |
|  | RAEF | 35-71 | 25-34 | < 25 | 12 |  |  |  |  |

Footnote: All measurements were derived using suiteHEART software with smooth endocardial segmentation; papillary muscles and trabeculations were excluded from left ventricular mass measurements. Volumetric and mass measurements were indexed to height. Reference ranges are defined as the interval between the 2.5th and 97.5th centiles derived from age-specific generalized additive models for location, scale and shape (GAMLSS) using the Box–Cox Cole–Green distribution. Centile-based categories beyond the reference range are derived from LMS-based thresholds (P0.5, P2.5, P97.5, and P99.5) and describe the position of a measurement within the reference distribution rather than disease severity or diagnostic abnormality. Reference ranges are based on a Black adult population.

Abbreviations: LV = left ventricle; RV = right ventricle; LA = left atrium; RA = right atrium; EDV = end-diastolic volume; ESV = end-systolic volume; SV = stroke volume; EF = ejection fraction; CO = cardiac output.

### 43. Table: Reference ranges and centile-based categories for cardiac chamber measurements derived using suiteHEART software (smooth segmentation), indexed to BSA, in an Asian adult population

|  |  | **Female** | | | | **Male** | | | |
| --- | --- | --- | --- | --- | --- | --- | --- | --- | --- |
| **Age group** | **Metrics** | **Reference range** | **Mild** | **At least moderate** | **n** | **Reference range** | **Mild** | **At least moderate** | **n** |
| 40-49 | LVEDVi | 54-88 | 89-96 | > 96 | 10 | 62-106 | 107-121 | > 121 | 20 |
|  | LVESVi | 18-37 | 38-45 | > 45 | 10 | 20-43 | 44-49 | > 49 | 20 |
|  | LVSVi | 31-53 | 25-30 | < 25 | 10 | 36-65 | 32-35 | < 32 | 20 |
|  | LVEF | 54-72 | 50-53 | < 50 | 10 | 53-71 | 50-52 | < 50 | 20 |
|  | LVMi | 30-51 | 52-57 | > 57 | 10 | 36-61 | 62-66 | > 66 | 20 |
|  | LVM/LVEDV | 0.4-0.8 | 0.9-1 | > 1 | 10 | 0.5-0.8 | 0.9-0.9 | > 0.9 | 20 |
|  | RVEDVi | 51-84 | 85-91 | > 91 | 10 | 61-103 | 104-109 | > 109 | 20 |
|  | RVESVi | 15-36 | 37-40 | > 40 | 10 | 21-49 | 50-54 | > 54 | 20 |
|  | RVEF | 49-76 | 45-48 | < 45 | 10 | 45-69 | 40-44 | < 40 | 20 |
|  | RVSVi | 31-60 | 29-30 | < 29 | 10 | 34-63 | 30-33 | < 30 | 20 |
|  | LAESVi | 19-40 | 41-43 | > 43 | 10 | 15-42 | 43-48 | > 48 | 19 |
|  | LAEF | 59-85 | 55-58 | < 55 | 10 | 61-81 | 57-60 | < 57 | 19 |
|  | RAESVi | 20-42 | 43-45 | > 45 | 10 | 19-52 | 53-57 | > 57 | 17 |
|  | RAEF | 39-69 | 31-38 | < 31 | 10 | 35-71 | 30-34 | < 30 | 17 |
| 50-59 | LVEDVi | 52-89 | 90-98 | > 98 | 83 | 57-100 | 101-110 | > 110 | 132 |
|  | LVESVi | 17-39 | 40-49 | > 49 | 83 | 19-43 | 44-49 | > 49 | 132 |
|  | LVSVi | 31-54 | 27-30 | < 27 | 83 | 33-63 | 29-32 | < 29 | 132 |
|  | LVEF | 53-73 | 48-52 | < 48 | 83 | 52-71 | 49-51 | < 49 | 131 |
|  | LVMi | 30-54 | 55-61 | > 61 | 83 | 36-64 | 65-70 | > 70 | 131 |
|  | LVM/LVEDV | 0.4-0.8 | 0.9-1 | > 1 | 83 | 0.5-0.9 | 01-1 | > 1 | 131 |
|  | RVEDVi | 50-88 | 89-97 | > 97 | 83 | 57-103 | 104-110 | > 110 | 131 |
|  | RVESVi | 16-37 | 38-41 | > 41 | 83 | 20-48 | 49-53 | > 53 | 131 |
|  | RVEF | 50-76 | 46-49 | < 46 | 82 | 46-70 | 41-45 | < 41 | 131 |
|  | RVSVi | 31-62 | 28-30 | < 28 | 82 | 32-63 | 28-31 | < 28 | 131 |
|  | LAESVi | 19-42 | 43-46 | > 46 | 77 | 16-44 | 45-49 | > 49 | 121 |
|  | LAEF | 58-84 | 53-57 | < 53 | 77 | 58-81 | 54-57 | < 54 | 122 |
|  | RAESVi | 20-46 | 47-52 | > 52 | 68 | 19-54 | 55-60 | > 60 | 118 |
|  | RAEF | 38-69 | 30-37 | < 30 | 68 | 38-71 | 33-37 | < 33 | 115 |
| 60-69 | LVEDVi | 50-91 | 92-102 | > 102 | 63 | 50-98 | 99-106 | > 106 | 118 |
|  | LVESVi | 15-40 | 41-50 | > 50 | 63 | 17-43 | 44-50 | > 50 | 118 |
|  | LVSVi | 31-56 | 28-30 | < 28 | 63 | 31-62 | 27-30 | < 27 | 118 |
|  | LVEF | 53-74 | 48-52 | < 48 | 63 | 51-72 | 47-50 | < 47 | 119 |
|  | LVMi | 30-59 | 60-69 | > 69 | 63 | 36-68 | 69-76 | > 76 | 119 |
|  | LVM/LVEDV | 0.5-0.9 | 01-1 | > 1 | 63 | 0.5-1 | 1.1-1.1 | > 1.1 | 118 |
|  | RVEDVi | 48-95 | 96-107 | > 107 | 63 | 52-103 | 104-112 | > 112 | 119 |
|  | RVESVi | 16-38 | 39-44 | > 44 | 63 | 18-46 | 47-51 | > 51 | 118 |
|  | RVEF | 52-75 | 48-51 | < 48 | 63 | 47-72 | 42-46 | < 42 | 119 |
|  | RVSVi | 29-64 | 26-28 | < 26 | 63 | 29-64 | 24-28 | < 24 | 119 |
|  | LAESVi | 19-46 | 47-52 | > 52 | 60 | 16-46 | 47-52 | > 52 | 111 |
|  | LAEF | 55-82 | 51-54 | < 51 | 60 | 55-82 | 51-54 | < 51 | 111 |
|  | RAESVi | 21-57 | 58-70 | > 70 | 51 | 19-57 | 58-65 | > 65 | 86 |
|  | RAEF | 37-72 | 29-36 | < 29 | 50 | 40-71 | 37-39 | < 37 | 86 |
| 70+ | LVEDVi | 47-94 | 95-107 | > 107 | 28 | 48-100 | 101-111 | > 111 | 40 |
|  | LVESVi | 15-36 | 37-42 | > 42 | 28 | 16-43 | 44-51 | > 51 | 40 |
|  | LVSVi | 31-61 | 29-30 | < 29 | 28 | 29-62 | 25-28 | < 25 | 40 |
|  | LVEF | 57-76 | 55-56 | < 55 | 28 | 50-72 | 44-49 | < 44 | 40 |
|  | LVMi | 30-65 | 66-78 | > 78 | 28 | 37-68 | 69-79 | > 79 | 40 |
|  | LVM/LVEDV | 0.5-0.9 | 1-1.1 | > 1.1 | 28 | 0.5-1.1 | 1.2-1.4 | > 1.4 | 40 |
|  | RVEDVi | 46-104 | 105-119 | > 119 | 27 | 48-104 | 105-114 | > 114 | 40 |
|  | RVESVi | 16-40 | 41-48 | > 48 | 27 | 17-44 | 45-50 | > 50 | 40 |
|  | RVEF | 53-75 | 50-52 | < 50 | 27 | 48-73 | 43-47 | < 43 | 39 |
|  | RVSVi | 27-66 | 23-26 | < 23 | 27 | 26-65 | 20-25 | < 20 | 40 |
|  | LAESVi | 19-52 | 53-62 | > 62 | 29 | 17-48 | 49-55 | > 55 | 34 |
|  | LAEF | 52-81 | 48-51 | < 48 | 28 | 52-83 | 47-51 | < 47 | 34 |
|  | RAESVi | 19-65 | 66-80 | > 80 | 21 | 19-61 | 62-71 | > 71 | 26 |
|  | RAEF | 39-79 | 30-38 | < 30 | 19 | 43-71 | 40-42 | < 40 | 25 |

Footnote: All measurements were derived using suiteHEART software with smooth endocardial segmentation; papillary muscles and trabeculations were excluded from left ventricular mass measurements. Volumetric and mass measurements were indexed to height. Reference ranges are defined as the interval between the 2.5th and 97.5th centiles derived from age-specific generalized additive models for location, scale and shape (GAMLSS) using the Box–Cox Cole–Green distribution. Centile-based categories beyond the reference range are derived from LMS-based thresholds (P0.5, P2.5, P97.5, and P99.5) and describe the position of a measurement within the reference distribution rather than disease severity or diagnostic abnormality. Reference ranges are based on an Asian adult population.

Abbreviations: LV = left ventricle; RV = right ventricle; LA = left atrium; RA = right atrium; EDV = end-diastolic volume; ESV = end-systolic volume; SV = stroke volume; EF = ejection fraction; CO = cardiac output.

### 44. Table: Reference ranges and centile-based categories for cardiac chamber measurements derived using suiteHEART software (smooth segmentation), indexed to BSA, in a Chinese adult population

|  |  | **Female** | | | | | **Male** | | | |
| --- | --- | --- | --- | --- | --- | --- | --- | --- | --- | --- |
| **Age group** | **Metrics** | **Reference range** | **Mild** | **At least moderate** | **n** | **Reference range** | | **Mild** | **At least moderate** | **n** |
| 18-29 | LVEDVi | 64-108 | 109-124 | > 124 | 16 | 72-104 | | 105-109 | > 109 | 19 |
|  | LVESVi | 22-43 | 44-49 | > 49 | 16 | 27-43 | | 44-46 | > 46 | 18 |
|  | LVSVi | 38-68 | 35-37 | < 35 | 16 | 43-64 | | 39-42 | < 39 | 19 |
|  | LVEF | 54-70 | 52-53 | < 52 | 16 | 51-66 | | 46-50 | < 46 | 19 |
|  | LVMi | 27-49 | 50-55 | > 55 | 16 | 40-61 | | 62-64 | > 64 | 19 |
|  | LVM/LVEDV | 0.4-0.6 |  | > 0.6 | 16 | 0.5-0.7 | | 0.8-0.8 | > 0.8 | 19 |
|  | RVEDVi | 67-112 | 113-124 | > 124 | 16 | 77-118 | | 119-123 | > 123 | 19 |
|  | RVESVi | 25-55 | 56-62 | > 62 | 16 | 33-58 | | 59-64 | > 64 | 19 |
|  | RVEF | 45-67 | 41-44 | < 41 | 16 | 48-64 | | 45-47 | < 45 | 19 |
|  | RVSVi | 35-64 | 31-34 | < 31 | 16 | 44-68 | | 41-43 | < 41 | 19 |
|  | LAESVi | 20-48 | 49-54 | > 54 | 12 | 21-44 | | 45-48 | > 48 | 15 |
|  | LAEF | 55-79 | 49-54 | < 49 | 12 | 53-77 | | 50-52 | < 50 | 15 |
|  | RAESVi | 22-46 | 47-51 | > 51 | 11 | 24-63 | | 64-80 | > 80 | 15 |
|  | RAEF | 47-69 | 45-46 | < 45 | 11 | 39-64 | | 33-38 | < 33 | 14 |
| 30-39 | LVEDVi | 61-103 | 104-116 | > 116 | 18 | 68-103 | | 104-109 | > 109 | 17 |
|  | LVESVi | 20-42 | 43-48 | > 48 | 18 | 25-43 | | 44-47 | > 47 | 17 |
|  | LVSVi | 37-65 | 34-36 | < 34 | 18 | 40-64 | | 36-39 | < 36 | 17 |
|  | LVEF | 55-71 | 52-54 | < 52 | 18 | 49-67 | | 42-48 | < 42 | 17 |
|  | LVMi | 29-51 | 52-58 | > 58 | 18 | 40-62 | | 63-67 | > 67 | 17 |
|  | LVM/LVEDV | 0.4-0.6 | 0.7-0.7 | > 0.7 | 18 | 0.5-0.8 | |  | > 0.8 | 17 |
|  | RVEDVi | 63-109 | 110-120 | > 120 | 18 | 73-115 | | 116-121 | > 121 | 17 |
|  | RVESVi | 23-51 | 52-57 | > 57 | 18 | 29-58 | | 59-64 | > 64 | 17 |
|  | RVEF | 46-69 | 42-45 | < 42 | 18 | 47-65 | | 44-46 | < 44 | 17 |
|  | RVSVi | 33-64 | 29-32 | < 29 | 18 | 41-68 | | 38-40 | < 38 | 17 |
|  | LAESVi | 20-48 | 49-54 | > 54 | 16 | 20-43 | | 44-47 | > 47 | 17 |
|  | LAEF | 55-79 | 49-54 | < 49 | 16 | 53-78 | | 50-52 | < 50 | 17 |
|  | RAESVi | 22-48 | 49-54 | > 54 | 16 | 24-62 | | 63-74 | > 74 | 17 |
|  | RAEF | 48-71 | 45-47 | < 45 | 14 | 39-66 | | 33-38 | < 33 | 17 |
| 40-49 | LVEDVi | 59-98 | 99-109 | > 109 | 24 | 65-103 | | 104-109 | > 109 | 20 |
|  | LVESVi | 19-40 | 41-46 | > 46 | 24 | 24-43 | | 44-48 | > 48 | 20 |
|  | LVSVi | 37-62 | 34-36 | < 34 | 24 | 37-64 | | 33-36 | < 33 | 20 |
|  | LVEF | 55-72 | 52-54 | < 52 | 24 | 52-70 | | 50-51 | < 50 | 20 |
|  | LVMi | 31-54 | 55-63 | > 63 | 24 | 40-64 | | 65-70 | > 70 | 20 |
|  | LVM/LVEDV | 0.4-0.7 |  | > 0.7 | 24 | 0.5-0.8 | | 0.9-0.9 | > 0.9 | 20 |
|  | RVEDVi | 59-104 | 105-115 | > 115 | 24 | 69-113 | | 114-120 | > 120 | 20 |
|  | RVESVi | 20-47 | 48-53 | > 53 | 24 | 25-56 | | 57-63 | > 63 | 20 |
|  | RVEF | 47-71 | 42-46 | < 42 | 24 | 47-67 | | 44-46 | < 44 | 20 |
|  | RVSVi | 32-63 | 28-31 | < 28 | 24 | 38-68 | | 35-37 | < 35 | 20 |
|  | LAESVi | 20-48 | 49-54 | > 54 | 20 | 18-43 | | 44-47 | > 47 | 19 |
|  | LAEF | 55-80 | 50-54 | < 50 | 20 | 53-80 | | 49-52 | < 49 | 19 |
|  | RAESVi | 23-51 | 52-58 | > 58 | 21 | 23-61 | | 62-71 | > 71 | 20 |
|  | RAEF | 48-73 | 45-47 | < 45 | 20 | 39-68 | | 33-38 | < 33 | 20 |
| 50-59 | LVEDVi | 56-94 | 95-102 | > 102 | 66 | 61-103 | | 104-111 | > 111 | 49 |
|  | LVESVi | 18-39 | 40-45 | > 45 | 66 | 23-44 | | 45-50 | > 50 | 49 |
|  | LVSVi | 36-59 | 33-35 | < 33 | 66 | 34-65 | | 30-33 | < 30 | 49 |
|  | LVEF | 55-73 | 52-54 | < 52 | 66 | 52-71 | | 49-51 | < 49 | 49 |
|  | LVMi | 32-57 | 58-69 | > 69 | 66 | 40-67 | | 68-76 | > 76 | 49 |
|  | LVM/LVEDV | 0.4-0.8 | 0.9-0.9 | > 0.9 | 66 | 0.5-0.9 | | 01-1 | > 1 | 49 |
|  | RVEDVi | 55-100 | 101-110 | > 110 | 66 | 65-111 | | 112-121 | > 121 | 49 |
|  | RVESVi | 17-42 | 43-48 | > 48 | 66 | 23-53 | | 54-58 | > 58 | 49 |
|  | RVEF | 48-72 | 43-47 | < 43 | 66 | 46-70 | | 43-45 | < 43 | 49 |
|  | RVSVi | 31-63 | 26-30 | < 26 | 66 | 35-68 | | 31-34 | < 31 | 49 |
|  | LAESVi | 20-48 | 49-55 | > 55 | 59 | 17-42 | | 43-46 | > 46 | 45 |
|  | LAEF | 55-80 | 50-54 | < 50 | 58 | 53-81 | | 48-52 | < 48 | 45 |
|  | RAESVi | 23-54 | 55-62 | > 62 | 61 | 22-60 | | 61-68 | > 68 | 44 |
|  | RAEF | 47-74 | 44-46 | < 44 | 60 | 39-70 | | 33-38 | < 33 | 44 |
| 60-69 | LVEDVi | 54-89 | 90-97 | > 97 | 52 | 58-104 | | 105-116 | > 116 | 37 |
|  | LVESVi | 16-37 | 38-43 | > 43 | 52 | 21-45 | | 46-53 | > 53 | 37 |
|  | LVSVi | 34-59 | 31-33 | < 31 | 52 | 31-67 | | 27-30 | < 27 | 37 |
|  | LVEF | 55-74 | 51-54 | < 51 | 52 | 50-71 | | 46-49 | < 46 | 37 |
|  | LVMi | 34-60 | 61-76 | > 76 | 52 | 39-72 | | 73-87 | > 87 | 37 |
|  | LVM/LVEDV | 0.5-0.9 | 01-1 | > 1 | 52 | 0.5-0.9 | | 01-1 | > 1 | 37 |
|  | RVEDVi | 50-95 | 96-104 | > 104 | 52 | 61-112 | | 113-125 | > 125 | 37 |
|  | RVESVi | 15-39 | 40-45 | > 45 | 52 | 21-47 | | 48-52 | > 52 | 37 |
|  | RVEF | 49-74 | 43-48 | < 43 | 51 | 46-73 | | 43-45 | < 43 | 37 |
|  | RVSVi | 29-63 | 25-28 | < 25 | 52 | 32-70 | | 28-31 | < 28 | 37 |
|  | LAESVi | 19-49 | 50-58 | > 58 | 48 | 16-41 | | 42-45 | > 45 | 31 |
|  | LAEF | 54-81 | 49-53 | < 49 | 48 | 52-83 | | 47-51 | < 47 | 31 |
|  | RAESVi | 24-57 | 58-67 | > 67 | 45 | 20-60 | | 61-67 | > 67 | 32 |
|  | RAEF | 46-74 | 42-45 | < 42 | 45 | 39-72 | | 33-38 | < 33 | 31 |
| 70+ | LVEDVi | 52-86 | 87-92 | > 92 | 12 |  | |  |  |  |
|  | LVESVi | 15-35 | 36-41 | > 41 | 12 |  | |  |  |  |
|  | LVSVi | 31-60 | 28-30 | < 28 | 12 |  | |  |  |  |
|  | LVEF | 54-75 | 50-53 | < 50 | 12 |  | |  |  |  |
|  | LVMi | 35-62 | 63-83 | > 83 | 12 |  | |  |  |  |
|  | LVM/LVEDV | 0.5-1 | 1.1-1.2 | > 1.2 | 12 |  | |  |  |  |
|  | RVEDVi | 46-91 | 92-100 | > 100 | 12 |  | |  |  |  |
|  | RVESVi | 14-37 | 38-42 | > 42 | 12 |  | |  |  |  |
|  | RVEF | 50-76 | 43-49 | < 43 | 12 |  | |  |  |  |
|  | RVSVi | 28-63 | 24-27 | < 24 | 12 |  | |  |  |  |
|  | LAESVi | 18-53 | 54-65 | > 65 | 12 |  | |  |  |  |
|  | LAEF | 54-82 | 48-53 | < 48 | 12 |  | |  |  |  |
|  | RAESVi | 24-61 | 62-73 | > 73 | 10 |  | |  |  |  |
|  | RAEF | 44-73 | 41-43 | < 41 | 10 |  | |  |  |  |

Footnote: All measurements were derived using suiteHEART software with smooth endocardial segmentation; papillary muscles and trabeculations were excluded from left ventricular mass measurements. Volumetric and mass measurements were indexed to height. Reference ranges are defined as the interval between the 2.5th and 97.5th centiles derived from age-specific generalized additive models for location, scale and shape (GAMLSS) using the Box–Cox Cole–Green distribution. Centile-based categories beyond the reference range are derived from LMS-based thresholds (P0.5, P2.5, P97.5, and P99.5) and describe the position of a measurement within the reference distribution rather than disease severity or diagnostic abnormality. Reference ranges are based on an Asian adult population.

Abbreviations: LV = left ventricle; RV = right ventricle; LA = left atrium; RA = right atrium; EDV = end-diastolic volume; ESV = end-systolic volume; SV = stroke volume; EF = ejection fraction; CO = cardiac output.

### 45. Table: Reference ranges and centile-based categories for cardiac chamber measurements derived using suiteHEART software (smooth segmentation), indexed to BSA, in a Mixed adult population

|  |  | **Female** | | | | **Male** | | | |
| --- | --- | --- | --- | --- | --- | --- | --- | --- | --- |
| **Age group** | **Metrics** | **Reference range** | **Mild** | **At least moderate** | **n** | **Reference range** | **Mild** | **At least moderate** | **n** |
| 50-59 | LVEDVi | 57-96 | 97-104 | > 104 | 134 | 60-111 | 112-119 | > 119 | 65 |
|  | LVESVi | 17-38 | 39-42 | > 42 | 134 | 21-46 | 47-49 | > 49 | 66 |
|  | LVSVi | 35-62 | 33-34 | < 33 | 133 | 36-74 | 31-35 | < 31 | 66 |
|  | LVEF | 54-74 | 52-53 | < 52 | 134 | 52-71 | 48-51 | < 48 | 66 |
|  | LVMi | 30-56 | 57-61 | > 61 | 134 | 40-76 | 77-86 | > 86 | 64 |
|  | LVM/LVEDV | 0.4-0.8 | 0.9-0.9 | > 0.9 | 134 | 0.5-1 | 1.1-1.3 | > 1.3 | 63 |
|  | RVEDVi | 53-94 | 95-100 | > 100 | 134 | 65-118 | 119-128 | > 128 | 65 |
|  | RVESVi | 17-40 | 41-44 | > 44 | 134 | 23-52 | 53-57 | > 57 | 65 |
|  | RVEF | 52-73 | 49-51 | < 49 | 134 | 46-71 | 40-45 | < 40 | 66 |
|  | RVSVi | 31-61 | 26-30 | < 26 | 134 | 35-73 | 30-34 | < 30 | 66 |
|  | LAESVi | 18-44 | 45-48 | > 48 | 122 | 18-50 | 51-57 | > 57 | 53 |
|  | LAEF | 57-80 | 51-56 | < 51 | 122 | 56-81 | 51-55 | < 51 | 54 |
|  | RAESVi | 21-55 | 56-63 | > 63 | 117 | 24-60 | 61-67 | > 67 | 54 |
|  | RAEF | 41-72 | 37-40 | < 37 | 117 | 37-72 | 32-36 | < 32 | 54 |
| 60-69 | LVEDVi | 51-94 | 95-102 | > 102 | 110 | 54-107 | 108-115 | > 115 | 59 |
|  | LVESVi | 15-38 | 39-43 | > 43 | 110 | 19-45 | 46-51 | > 51 | 58 |
|  | LVSVi | 33-61 | 29-32 | < 29 | 110 | 32-67 | 27-31 | < 27 | 59 |
|  | LVEF | 54-74 | 51-53 | < 51 | 110 | 50-71 | 45-49 | < 45 | 59 |
|  | LVMi | 31-58 | 59-64 | > 64 | 110 | 40-73 | 74-81 | > 81 | 59 |
|  | LVM/LVEDV | 0.4-0.8 | 0.9-1 | > 1 | 110 | 0.5-1.1 | 1.2-1.3 | > 1.3 | 59 |
|  | RVEDVi | 49-99 | 100-108 | > 108 | 110 | 58-118 | 119-131 | > 131 | 57 |
|  | RVESVi | 17-40 | 41-44 | > 44 | 110 | 22-55 | 56-64 | > 64 | 58 |
|  | RVEF | 52-74 | 47-51 | < 47 | 110 | 47-71 | 43-46 | < 43 | 58 |
|  | RVSVi | 28-65 | 22-27 | < 22 | 110 | 30-71 | 25-29 | < 25 | 58 |
|  | LAESVi | 18-47 | 48-52 | > 52 | 100 | 19-45 | 46-50 | > 50 | 53 |
|  | LAEF | 55-81 | 50-54 | < 50 | 100 | 52-81 | 48-51 | < 48 | 53 |
|  | RAESVi | 23-56 | 57-64 | > 64 | 93 | 22-62 | 63-69 | > 69 | 40 |
|  | RAEF | 40-73 | 35-39 | < 35 | 93 | 37-72 | 32-36 | < 32 | 40 |
| 70+ | LVEDVi | 48-96 | 97-106 | > 106 | 51 | 49-103 | 104-112 | > 112 | 33 |
|  | LVESVi | 14-36 | 37-40 | > 40 | 51 | 17-47 | 48-55 | > 55 | 33 |
|  | LVSVi | 29-61 | 24-28 | < 24 | 51 | 29-61 | 22-28 | < 22 | 34 |
|  | LVEF | 53-75 | 49-52 | < 49 | 51 | 48-70 | 40-47 | < 40 | 35 |
|  | LVMi | 31-60 | 61-67 | > 67 | 51 | 40-71 | 72-78 | > 78 | 35 |
|  | LVM/LVEDV | 0.5-0.9 | 01-1 | > 1 | 51 | 0.5-1.1 | 1.2-1.3 | > 1.3 | 33 |
|  | RVEDVi | 55-92 | 93-99 | > 99 | 51 | 53-120 | 121-136 | > 136 | 34 |
|  | RVESVi | 17-36 | 37-41 | > 41 | 51 | 23-56 | 57-69 | > 69 | 35 |
|  | RVEF | 50-75 | 44-49 | < 44 | 51 | 47-72 | 44-46 | < 44 | 35 |
|  | RVSVi | 30-64 | 25-29 | < 25 | 51 | 26-70 | 21-25 | < 21 | 33 |
|  | LAESVi | 17-50 | 51-55 | > 55 | 43 | 15-51 | 52-61 | > 61 | 34 |
|  | LAEF | 53-83 | 49-52 | < 49 | 42 | 50-83 | 45-49 | < 45 | 34 |
|  | RAESVi | 25-54 | 55-62 | > 62 | 36 | 19-64 | 65-72 | > 72 | 21 |
|  | RAEF | 38-74 | 33-37 | < 33 | 34 | 35-68 | 30-34 | < 30 | 21 |

Footnote: All measurements were derived using suiteHEART software with smooth endocardial segmentation; papillary muscles and trabeculations were excluded from left ventricular mass measurements. Volumetric and mass measurements were indexed to height. Reference ranges are defined as the interval between the 2.5th and 97.5th centiles derived from age-specific generalized additive models for location, scale and shape (GAMLSS) using the Box–Cox Cole–Green distribution. Centile-based categories beyond the reference range are derived from LMS-based thresholds (P0.5, P2.5, P97.5, and P99.5) and describe the position of a measurement within the reference distribution rather than disease severity or diagnostic abnormality. Reference ranges are based on a Mixed adult population.

Abbreviations: LV = left ventricle; RV = right ventricle; LA = left atrium; RA = right atrium; EDV = end-diastolic volume; ESV = end-systolic volume; SV = stroke volume; EF = ejection fraction; CO = cardiac output.

### 46. Table: Reference ranges and centile-based categories for cardiac chamber measurements derived using suiteHEART software (anatomical segmentation), indexed to BSA, in a White adult population

|  |  | **Female** | | | | **Male** | | | |
| --- | --- | --- | --- | --- | --- | --- | --- | --- | --- |
| **Age group** | **Metrics** | **Reference range** | **Mild** | **At least moderate** | **n** | **Reference range** | **Mild** | **At least moderate** | **n** |
| 18-29 | LVEDVi | 55-99 | 100-107 | > 107 | 101 | 64-112 | 113-120 | > 120 | 101 |
|  | LVESVi | 17-39 | 40-44 | > 44 | 101 | 19-47 | 48-52 | > 52 | 103 |
|  | LVSVi | 35-65 | 30-34 | < 30 | 101 | 39-73 | 34-38 | < 34 | 101 |
|  | LVEF | 55-74 | 51-54 | < 51 | 101 | 52-73 | 49-51 | < 49 | 104 |
|  | LVMi | 40-66 | 67-71 | > 71 | 101 | 53-85 | 86-91 | > 91 | 101 |
|  | LVM/LVEDV | 0.5-0.9 | 1-1.1 | > 1.1 | 101 | 0.6-1.1 |  | > 1.2 | 99 |
|  | RVEDVi | 60-112 | 113-120 | > 120 | 101 | 72-125 | 126-133 | > 133 | 98 |
|  | RVESVi | 23-51 | 52-57 | > 57 | 100 | 26-59 | 60-64 | > 64 | 98 |
|  | RVEF | 47-70 | 44-46 | < 44 | 99 | 46-68 | 43-45 | < 43 | 102 |
|  | RVSVi | 35-70 | 32-34 | < 32 | 100 | 40-76 | 34-39 | < 34 | 101 |
|  | LAESV | 18-42 | 43-46 | > 46 | 65 | 19-50 | 51-57 | > 57 | 64 |
|  | LAEF | 57-85 | 52-56 | < 52 | 65 | 55-79 | 51-54 | < 51 | 62 |
|  | RAESV | 23-48 | 49-52 | > 52 | 77 | 24-60 | 61-66 | > 66 | 87 |
|  | RAEF | 42-75 | 38-41 | < 38 | 72 | 35-71 | 31-34 | < 31 | 85 |
| 30-39 | LVEDVi | 52-95 | 96-103 | > 103 | 131 | 55-100 | 101-107 | > 107 | 128 |
|  | LVESVi | 15-37 | 38-42 | > 42 | 131 | 16-41 | 42-46 | > 46 | 128 |
|  | LVSVi | 33-63 | 29-32 | < 29 | 130 | 34-67 | 30-33 | < 30 | 128 |
|  | LVEF | 56-76 | 52-55 | < 52 | 131 | 53-75 | 50-52 | < 50 | 129 |
|  | LVMi | 40-67 | 68-72 | > 72 | 131 | 52-84 | 85-90 | > 90 | 125 |
|  | LVM/LVEDV | 0.6-1 | 1.1-1.2 | > 1.2 | 131 | 0.6-1.2 | 1.3-1.3 | > 1.3 | 126 |
|  | RVEDVi | 58-109 | 110-117 | > 117 | 131 | 65-120 | 121-128 | > 128 | 127 |
|  | RVESVi | 21-49 | 50-54 | > 54 | 131 | 23-55 | 56-60 | > 60 | 128 |
|  | RVEF | 49-71 | 45-48 | < 45 | 131 | 47-71 | 43-46 | < 43 | 129 |
|  | RVSVi | 34-69 | 30-33 | < 30 | 131 | 37-74 | 31-36 | < 31 | 126 |
|  | LAESV | 19-45 | 46-50 | > 50 | 95 | 18-49 | 50-56 | > 56 | 86 |
|  | LAEF | 57-84 | 53-56 | < 53 | 95 | 55-80 | 51-54 | < 51 | 87 |
|  | RAESV | 23-50 | 51-55 | > 55 | 113 | 23-60 | 61-67 | > 67 | 104 |
|  | RAEF | 41-75 | 37-40 | < 37 | 108 | 36-72 | 31-35 | < 31 | 103 |
| 40-49 | LVEDVi | 51-94 | 95-101 | > 101 | 521 | 54-103 | 104-111 | > 111 | 437 |
|  | LVESVi | 13-35 | 36-40 | > 40 | 521 | 14-41 | 42-46 | > 46 | 440 |
|  | LVSVi | 33-62 | 29-32 | < 29 | 521 | 34-68 | 29-33 | < 29 | 433 |
|  | LVEF | 57-78 | 53-56 | < 53 | 520 | 54-77 | 51-53 | < 51 | 442 |
|  | LVMi | 40-67 | 68-73 | > 73 | 521 | 51-83 | 84-89 | > 89 | 414 |
|  | LVM/LVEDV | 0.6-1.1 |  | > 1.2 | 521 | 0.6-1.2 | 1.3-1.4 | > 1.4 | 411 |
|  | RVEDVi | 54-104 | 105-113 | > 113 | 521 | 62-120 | 121-129 | > 129 | 434 |
|  | RVESVi | 18-44 | 45-50 | > 50 | 521 | 20-53 | 54-59 | > 59 | 441 |
|  | RVEF | 51-73 | 47-50 | < 47 | 517 | 48-72 | 43-47 | < 43 | 442 |
|  | RVSVi | 32-67 | 28-31 | < 28 | 519 | 35-75 | 29-34 | < 29 | 428 |
|  | LAESV | 19-48 | 49-53 | > 53 | 453 | 18-49 | 50-56 | > 56 | 374 |
|  | LAEF | 57-83 | 53-56 | < 53 | 453 | 55-81 | 51-54 | < 51 | 374 |
|  | RAESV | 22-52 | 53-58 | > 58 | 452 | 23-64 | 65-71 | > 71 | 362 |
|  | RAEF | 41-75 | 36-40 | < 36 | 445 | 36-73 | 31-35 | < 31 | 360 |
| 50-59 | LVEDVi | 48-90 | 91-98 | > 98 | 912 | 50-100 | 101-109 | > 109 | 831 |
|  | LVESVi | 11-33 | 34-38 | > 38 | 912 | 12-39 | 40-44 | > 44 | 835 |
|  | LVSVi | 33-62 | 29-32 | < 29 | 912 | 33-68 | 28-32 | < 28 | 825 |
|  | LVEF | 58-80 | 54-57 | < 54 | 911 | 56-79 | 51-55 | < 51 | 833 |
|  | LVMi | 40-67 | 68-74 | > 74 | 911 | 49-82 | 83-88 | > 88 | 772 |
|  | LVM/LVEDV | 0.6-1.1 | 1.2-1.3 | > 1.3 | 911 | 0.7-1.3 | 1.4-1.5 | > 1.5 | 770 |
|  | RVEDVi | 49-96 | 97-105 | > 105 | 910 | 54-109 | 110-119 | > 119 | 826 |
|  | RVESVi | 15-39 | 40-44 | > 44 | 910 | 17-48 | 49-54 | > 54 | 826 |
|  | RVEF | 52-75 | 48-51 | < 48 | 908 | 48-74 | 43-47 | < 43 | 831 |
|  | RVSVi | 30-64 | 26-29 | < 26 | 908 | 31-69 | 25-30 | < 25 | 824 |
|  | LAESV | 19-48 | 49-54 | > 54 | 825 | 17-49 | 50-56 | > 56 | 700 |
|  | LAEF | 56-83 | 51-55 | < 51 | 823 | 54-82 | 50-53 | < 50 | 703 |
|  | RAESV | 21-54 | 55-61 | > 61 | 769 | 21-60 | 61-68 | > 68 | 604 |
|  | RAEF | 40-75 | 35-39 | < 35 | 760 | 36-74 | 31-35 | < 31 | 604 |
| 60-69 | LVEDVi | 45-86 | 87-95 | > 95 | 920 | 47-99 | 100-108 | > 108 | 804 |
|  | LVESVi | 10-32 | 33-37 | > 37 | 919 | 11-37 | 38-43 | > 43 | 801 |
|  | LVSVi | 32-60 | 28-31 | < 28 | 920 | 31-66 | 26-30 | < 26 | 793 |
|  | LVEF | 59-81 | 54-58 | < 54 | 916 | 56-80 | 52-55 | < 52 | 803 |
|  | LVMi | 40-68 | 69-75 | > 75 | 919 | 48-81 | 82-87 | > 87 | 761 |
|  | LVM/LVEDV | 0.6-1.2 | 1.3-1.4 | > 1.4 | 919 | 0.7-1.3 | 1.4-1.6 | > 1.6 | 759 |
|  | RVEDVi | 47-92 | 93-102 | > 102 | 918 | 51-108 | 109-118 | > 118 | 803 |
|  | RVESVi | 13-36 | 37-41 | > 41 | 919 | 15-46 | 47-52 | > 52 | 804 |
|  | RVEF | 53-77 | 47-52 | < 47 | 915 | 48-76 | 43-47 | < 43 | 799 |
|  | RVSVi | 29-62 | 25-28 | < 25 | 917 | 29-69 | 23-28 | < 23 | 795 |
|  | LAESV | 18-49 | 50-56 | > 56 | 809 | 16-49 | 50-57 | > 57 | 652 |
|  | LAEF | 54-83 | 48-53 | < 48 | 807 | 53-83 | 48-52 | < 48 | 654 |
|  | RAESV | 20-57 | 58-66 | > 66 | 721 | 21-63 | 64-72 | > 72 | 521 |
|  | RAEF | 41-75 | 35-40 | < 35 | 699 | 37-74 | 32-36 | < 32 | 496 |
| 70+ | LVEDVi | 43-85 | 86-94 | > 94 | 1101 | 44-97 | 98-108 | > 108 | 1254 |
|  | LVESVi | 8-30 | 31-36 | > 36 | 1101 | 10-38 | 39-45 | > 45 | 1250 |
|  | LVSVi | 31-59 | 28-30 | < 28 | 1101 | 30-64 | 24-29 | < 24 | 1254 |
|  | LVEF | 59-83 | 55-58 | < 55 | 1086 | 56-80 | 51-55 | < 51 | 1242 |
|  | LVMi | 39-69 | 70-76 | > 76 | 1100 | 47-80 | 81-86 | > 86 | 1224 |
|  | LVM/LVEDV | 0.6-1.3 | 1.4-1.5 | > 1.5 | 1100 | 0.7-1.4 | 1.5-1.7 | > 1.7 | 1222 |
|  | RVEDVi | 46-92 | 93-102 | > 102 | 1100 | 50-106 | 107-116 | > 116 | 1252 |
|  | RVESVi | 12-36 | 37-42 | > 42 | 1100 | 15-45 | 46-52 | > 52 | 1253 |
|  | RVEF | 52-78 | 46-51 | < 46 | 1086 | 48-76 | 42-47 | < 42 | 1236 |
|  | RVSVi | 28-63 | 24-27 | < 24 | 1098 | 27-69 | 20-26 | < 20 | 1247 |
|  | LAESV | 17-52 | 53-61 | > 61 | 947 | 15-50 | 51-58 | > 58 | 953 |
|  | LAEF | 50-83 | 44-49 | < 44 | 938 | 51-84 | 45-50 | < 45 | 943 |
|  | RAESV | 20-61 | 62-73 | > 73 | 769 | 22-66 | 67-76 | > 76 | 631 |
|  | RAEF | 42-77 | 37-41 | < 37 | 753 | 37-75 | 32-36 | < 32 | 619 |

Footnote: All measurements were derived using suiteHEART software with anatomical endocardial segmentation; papillary muscles and trabeculations were included in left ventricular mass measurements. Volumetric and mass measurements were indexed to height. Reference ranges are defined as the interval between the 2.5th and 97.5th centiles derived from age-specific generalized additive models for location, scale and shape (GAMLSS) using the Box–Cox Cole–Green distribution. Centile-based categories beyond the reference range are derived from LMS-based thresholds (P0.5, P2.5, P97.5, and P99.5) and describe the position of a measurement within the reference distribution rather than disease severity or diagnostic abnormality. Reference ranges are based on a White adult population.

Abbreviations: LV = left ventricle; RV = right ventricle; LA = left atrium; RA = right atrium; EDV = end-diastolic volume; ESV = end-systolic volume; SV = stroke volume; EF = ejection fraction; CO = cardiac output.

### 47. Table: Reference ranges and centile-based categories for cardiac chamber measurements derived using suitheart software (anatomical segmentation), indexed to BSA, in a Black adult population

|  |  | **Female** | | | | **Male** | | | |
| --- | --- | --- | --- | --- | --- | --- | --- | --- | --- |
| **Age group** | **Metrics** | **Reference range** | **Mild** | **At least moderate** | **n** | **Reference range** | **Mild** | **At least moderate** | **n** |
| 40-49 | LVEDVi | 48-88 | 89-94 | > 94 | 10 |  |  |  |  |
|  | LVESVi | 10-35 | 36-38 | > 38 | 10 |  |  |  |  |
|  | LVSVi | 34-60 | 29-33 | < 29 | 10 |  |  |  |  |
|  | LVEF | 57-79 | 53-56 | < 53 | 10 |  |  |  |  |
|  | LVMi | 42-75 | 76-91 | > 91 | 10 |  |  |  |  |
|  | LVM/LVEDV | 0.6-1.2 | 1.3-1.4 | > 1.4 | 10 |  |  |  |  |
|  | RVEDVi | 47-95 | 96-103 | > 103 | 10 |  |  |  |  |
|  | RVESVi | 15-42 | 43-48 | > 48 | 10 |  |  |  |  |
|  | RVEF | 48-73 | 41-47 | < 41 | 10 |  |  |  |  |
|  | RVSVi | 28-60 | 23-27 | < 23 | 10 |  |  |  |  |
|  | LAESVi |  |  |  |  |  |  |  |  |
|  | LAEF |  |  |  |  |  |  |  |  |
|  | RAESVi |  |  |  |  |  |  |  |  |
|  | RAEF |  |  |  |  |  |  |  |  |
| 50-59 | LVEDVi | 47-87 | 88-93 | > 93 | 95 | 51-100 | 101-110 | > 110 | 77 |
|  | LVESVi | 9-32 | 33-36 | > 36 | 95 | 12-37 | 38-42 | > 42 | 76 |
|  | LVSVi | 33-60 | 29-32 | < 29 | 95 | 35-68 | 32-34 | < 32 | 75 |
|  | LVEF | 58-80 | 55-57 | < 55 | 92 | 56-80 | 52-55 | < 52 | 76 |
|  | LVMi | 43-75 | 76-86 | > 86 | 95 | 50-83 | 84-90 | > 90 | 68 |
|  | LVM/LVEDV | 0.6-1.2 | 1.3-1.4 | > 1.4 | 95 | 0.7-1.3 | 1.4-1.5 | > 1.5 | 68 |
|  | RVEDVi | 49-95 | 96-103 | > 103 | 95 | 59-103 | 104-113 | > 113 | 77 |
|  | RVESVi | 15-42 | 43-48 | > 48 | 95 | 19-46 | 47-56 | > 56 | 77 |
|  | RVEF | 50-75 | 45-49 | < 45 | 95 | 48-74 | 41-47 | < 41 | 77 |
|  | RVSVi | 29-61 | 25-28 | < 25 | 95 | 33-66 | 28-32 | < 28 | 76 |
|  | LAESVi | 20-45 | 46-49 | > 49 | 88 | 18-46 | 47-53 | > 53 | 67 |
|  | LAEF | 59-79 | 56-58 | < 56 | 88 | 56-83 | 53-55 | < 53 | 67 |
|  | RAESVi | 20-58 | 59-66 | > 66 | 84 | 24-57 | 58-62 | > 62 | 64 |
|  | RAEF | 38-69 | 32-37 | < 32 | 84 | 38-65 | 35-37 | < 35 | 63 |
| 60-69 | LVEDVi | 45-84 | 85-92 | > 92 | 65 | 50-100 | 101-111 | > 111 | 48 |
|  | LVESVi | 8-29 | 30-34 | > 34 | 65 | 10-41 | 42-48 | > 48 | 48 |
|  | LVSVi | 33-60 | 29-32 | < 29 | 65 | 35-66 | 32-34 | < 32 | 48 |
|  | LVEF | 60-83 | 56-59 | < 56 | 64 | 48-81 | 36-47 | < 36 | 48 |
|  | LVMi | 44-75 | 76-83 | > 83 | 65 | 51-84 | 85-91 | > 91 | 44 |
|  | LVM/LVEDV | 0.7-1.3 | 1.4-1.5 | > 1.5 | 65 | 0.7-1.3 | 1.4-1.5 | > 1.5 | 44 |
|  | RVEDVi | 51-94 | 95-105 | > 105 | 65 | 55-104 | 105-114 | > 114 | 48 |
|  | RVESVi | 13-41 | 42-49 | > 49 | 65 | 15-50 | 51-61 | > 61 | 48 |
|  | RVEF | 51-79 | 47-50 | < 47 | 64 | 48-77 | 42-47 | < 42 | 47 |
|  | RVSVi | 31-62 | 27-30 | < 27 | 65 | 31-65 | 26-30 | < 26 | 48 |
|  | LAESVi | 18-49 | 50-55 | > 55 | 60 | 17-50 | 51-58 | > 58 | 42 |
|  | LAEF | 54-81 | 49-53 | < 49 | 61 | 52-85 | 48-51 | < 48 | 42 |
|  | RAESVi | 18-51 | 52-55 | > 55 | 52 | 21-62 | 63-68 | > 68 | 40 |
|  | RAEF | 32-66 | 21-31 | < 21 | 53 | 36-71 | 32-35 | < 32 | 41 |
| 70+ | LVEDVi | 42-83 | 84-93 | > 93 | 18 | 45-90 | 91-100 | > 100 | 14 |
|  | LVESVi | 8-30 | 31-35 | > 35 | 18 | 8-39 | 40-48 | > 48 | 14 |
|  | LVSVi | 32-60 | 29-31 | < 29 | 18 | 35-65 | 32-34 | < 32 | 14 |
|  | LVEF | 60-87 | 55-59 | < 55 | 18 | 53-82 | 44-52 | < 44 | 14 |
|  | LVMi | 45-76 | 77-82 | > 82 | 18 | 50-82 | 83-89 | > 89 | 14 |
|  | LVM/LVEDV | 0.8-1.4 | 1.5-1.6 | > 1.6 | 18 | 0.7-1.4 | 1.5-1.6 | > 1.6 | 14 |
|  | RVEDVi | 52-94 | 95-108 | > 108 | 18 | 50-104 | 105-114 | > 114 | 14 |
|  | RVESVi | 12-41 | 42-50 | > 50 | 18 | 11-52 | 53-63 | > 63 | 14 |
|  | RVEF | 48-80 | 37-47 | < 37 | 18 | 48-83 | 43-47 | < 43 | 14 |
|  | RVSVi | 32-65 | 29-31 | < 29 | 18 | 30-65 | 23-29 | < 23 | 14 |
|  | LAESVi | 15-55 | 56-64 | > 64 | 16 |  |  |  |  |
|  | LAEF | 47-82 | 39-46 | < 39 | 17 |  |  |  |  |
|  | RAESVi | 18-48 | 49-50 | > 50 | 13 |  |  |  |  |
|  | RAEF | 35-71 | 25-34 | < 25 | 12 |  |  |  |  |

Footnote: All measurements were derived using suiteHEART software with anatomical endocardial segmentation; papillary muscles and trabeculations were included in left ventricular mass measurements. Volumetric and mass measurements were indexed to height. Reference ranges are defined as the interval between the 2.5th and 97.5th centiles derived from age-specific generalized additive models for location, scale and shape (GAMLSS) using the Box–Cox Cole–Green distribution. Centile-based categories beyond the reference range are derived from LMS-based thresholds (P0.5, P2.5, P97.5, and P99.5) and describe the position of a measurement within the reference distribution rather than disease severity or diagnostic abnormality. Reference ranges are based on a Black adult population.

Abbreviations: LV = left ventricle; RV = right ventricle; LA = left atrium; RA = right atrium; EDV = end-diastolic volume; ESV = end-systolic volume; SV = stroke volume; EF = ejection fraction; CO = cardiac output.

### 48. Table: Reference ranges and centile-based categories for cardiac chamber measurements derived using suiteHEART software (anatomical segmentation), indexed to BSA, in an Asian adult population

|  |  | **Female** | | | | **Male** | | | |
| --- | --- | --- | --- | --- | --- | --- | --- | --- | --- |
| **Age group** | **Metrics** | **Reference range** | **Mild** | **At least moderate** | **n** | **Reference range** | **Mild** | **At least moderate** | **n** |
| 40-49 | LVEDVi | 47-80 | 81-87 | > 87 | 10 | 55-95 | 96-108 | > 108 | 20 |
|  | LVESVi | 10-34 | 35-43 | > 43 | 10 | 13-33 | 34-38 | > 38 | 20 |
|  | LVSVi | 31-53 | 25-30 | < 25 | 10 | 37-65 | 34-36 | < 34 | 20 |
|  | LVEF | 59-80 | 54-58 | < 54 | 10 | 59-79 | 55-58 | < 55 | 20 |
|  | LVMi | 35-58 | 59-65 | > 65 | 10 | 43-72 | 73-77 | > 77 | 20 |
|  | LVM/LVEDV | 0.6-1.1 | 1.2-1.3 | > 1.3 | 10 | 0.6-1.1 |  | > 1.2 | 20 |
|  | RVEDVi | 47-79 | 80-86 | > 86 | 10 | 56-96 | 97-102 | > 102 | 20 |
|  | RVESVi | 13-33 | 34-38 | > 38 | 10 | 16-43 | 44-47 | > 47 | 20 |
|  | RVEF | 51-78 | 46-50 | < 46 | 10 | 48-72 | 43-47 | < 43 | 19 |
|  | RVSVi | 30-57 | 28-29 | < 28 | 10 | 33-62 | 29-32 | < 29 | 20 |
|  | LAESVi | 19-40 | 41-43 | > 43 | 10 | 15-42 | 43-48 | > 48 | 19 |
|  | LAEF | 59-85 | 55-58 | < 55 | 10 | 61-81 | 57-60 | < 57 | 19 |
|  | RAESVi | 20-42 | 43-45 | > 45 | 10 | 19-52 | 53-57 | > 57 | 17 |
|  | RAEF | 39-69 | 31-38 | < 31 | 10 | 35-71 | 30-34 | < 30 | 17 |
| 50-59 | LVEDVi | 46-80 | 81-88 | > 88 | 83 | 50-90 | 91-98 | > 98 | 132 |
|  | LVESVi | 10-33 | 34-41 | > 41 | 83 | 12-34 | 35-39 | > 39 | 132 |
|  | LVSVi | 31-54 | 27-30 | < 27 | 83 | 33-62 | 30-32 | < 30 | 132 |
|  | LVEF | 58-81 | 52-57 | < 52 | 83 | 58-80 | 54-57 | < 54 | 132 |
|  | LVMi | 36-63 | 64-71 | > 71 | 83 | 42-75 | 76-81 | > 81 | 130 |
|  | LVM/LVEDV | 0.6-1.1 | 1.2-1.3 | > 1.3 | 83 | 0.7-1.2 | 1.3-1.3 | > 1.3 | 130 |
|  | RVEDVi | 46-82 | 83-90 | > 90 | 83 | 52-95 | 96-102 | > 102 | 131 |
|  | RVESVi | 13-33 | 34-38 | > 38 | 83 | 15-42 | 43-47 | > 47 | 131 |
|  | RVEF | 52-78 | 47-51 | < 47 | 82 | 48-74 | 43-47 | < 43 | 131 |
|  | RVSVi | 30-59 | 28-29 | < 28 | 82 | 31-62 | 27-30 | < 27 | 131 |
|  | LAESVi | 19-42 | 43-46 | > 46 | 77 | 16-44 | 45-49 | > 49 | 121 |
|  | LAEF | 58-84 | 53-57 | < 53 | 77 | 58-81 | 54-57 | < 54 | 122 |
|  | RAESVi | 20-46 | 47-52 | > 52 | 68 | 19-54 | 55-60 | > 60 | 118 |
|  | RAEF | 38-69 | 30-37 | < 30 | 68 | 38-71 | 33-37 | < 33 | 115 |
| 60-69 | LVEDVi | 43-81 | 82-90 | > 90 | 63 | 43-87 | 88-94 | > 94 | 118 |
|  | LVESVi | 9-30 | 31-38 | > 38 | 63 | 10-34 | 35-41 | > 41 | 119 |
|  | LVSVi | 31-56 | 28-30 | < 28 | 63 | 30-60 | 26-29 | < 26 | 118 |
|  | LVEF | 58-83 | 53-57 | < 53 | 63 | 57-81 | 51-56 | < 51 | 119 |
|  | LVMi | 37-70 | 71-81 | > 81 | 63 | 42-77 | 78-85 | > 85 | 116 |
|  | LVM/LVEDV | 0.6-1.2 | 1.3-1.4 | > 1.4 | 63 | 0.7-1.3 | 1.4-1.5 | > 1.5 | 116 |
|  | RVEDVi | 44-89 | 90-99 | > 99 | 63 | 48-95 | 96-103 | > 103 | 119 |
|  | RVESVi | 13-33 | 34-38 | > 38 | 63 | 14-42 | 43-48 | > 48 | 119 |
|  | RVEF | 54-78 | 49-53 | < 49 | 63 | 49-76 | 43-48 | < 43 | 119 |
|  | RVSVi | 29-62 | 26-28 | < 26 | 63 | 28-63 | 24-27 | < 24 | 119 |
|  | LAESVi | 19-46 | 47-52 | > 52 | 60 | 16-46 | 47-52 | > 52 | 111 |
|  | LAEF | 55-82 | 51-54 | < 51 | 60 | 55-82 | 51-54 | < 51 | 111 |
|  | RAESVi | 21-57 | 58-70 | > 70 | 51 | 19-57 | 58-65 | > 65 | 86 |
|  | RAEF | 37-72 | 29-36 | < 29 | 50 | 40-71 | 37-39 | < 37 | 86 |
| 70+ | LVEDVi | 41-82 | 83-92 | > 92 | 28 | 40-88 | 89-97 | > 97 | 40 |
|  | LVESVi | 8-28 | 29-35 | > 35 | 28 | 9-35 | 36-44 | > 44 | 40 |
|  | LVSVi | 31-60 | 29-30 | < 29 | 28 | 30-63 | 26-29 | < 26 | 40 |
|  | LVEF | 63-83 | 60-62 | < 60 | 27 | 54-82 | 47-53 | < 47 | 40 |
|  | LVMi | 36-75 | 76-91 | > 91 | 28 | 43-77 | 78-86 | > 86 | 40 |
|  | LVM/LVEDV | 0.7-1.3 | 1.4-1.5 | > 1.5 | 28 | 0.7-1.5 | 1.6-1.8 | > 1.8 | 40 |
|  | RVEDVi | 42-96 | 97-109 | > 109 | 27 | 43-95 | 96-104 | > 104 | 40 |
|  | RVESVi | 12-33 | 34-39 | > 39 | 27 | 14-42 | 43-51 | > 51 | 40 |
|  | RVEF | 55-78 | 51-54 | < 51 | 27 | 49-78 | 43-48 | < 43 | 40 |
|  | RVSVi | 26-64 | 22-25 | < 22 | 27 | 25-63 | 20-24 | < 20 | 40 |
|  | LAESVi | 19-52 | 53-62 | > 62 | 29 | 17-48 | 49-55 | > 55 | 34 |
|  | LAEF | 52-81 | 48-51 | < 48 | 28 | 52-83 | 47-51 | < 47 | 34 |
|  | RAESVi | 19-65 | 66-80 | > 80 | 21 | 19-61 | 62-71 | > 71 | 26 |
|  | RAEF | 39-79 | 30-38 | < 30 | 19 | 43-71 | 40-42 | < 40 | 25 |

Footnote: All measurements were derived using suiteHEART software with anatomical endocardial segmentation; papillary muscles and trabeculations were included in left ventricular mass measurements. Volumetric and mass measurements were indexed to height. Reference ranges are defined as the interval between the 2.5th and 97.5th centiles derived from age-specific generalized additive models for location, scale and shape (GAMLSS) using the Box–Cox Cole–Green distribution. Centile-based categories beyond the reference range are derived from LMS-based thresholds (P0.5, P2.5, P97.5, and P99.5) and describe the position of a measurement within the reference distribution rather than disease severity or diagnostic abnormality. Reference ranges are based on an Asian adult population.

Abbreviations: LV = left ventricle; RV = right ventricle; LA = left atrium; RA = right atrium; EDV = end-diastolic volume; ESV = end-systolic volume; SV = stroke volume; EF = ejection fraction; CO = cardiac output.

### 49. Table: Reference ranges and centile-based categories for cardiac chamber measurements derived using suiteHEART software (anatomical segmentation), indexed to BSA, in a Chinese adult population

|  |  | **Female** | | | | | **Male** | | | |
| --- | --- | --- | --- | --- | --- | --- | --- | --- | --- | --- |
| **Age group** | **Metrics** | **Reference range** | **Mild** | **At least moderate** | **n** | **Reference range** | | **Mild** | **At least moderate** | **n** |
| 18-29 | LVEDVi | 58-98 | 99-112 | > 112 | 16 | 64-95 | | 96-100 | > 100 | 19 |
|  | LVESVi | 17-39 | 40-49 | > 49 | 16 | 20-40 | | 41-50 | > 50 | 19 |
|  | LVSVi | 37-67 | 35-36 | < 35 | 16 | 43-63 | | 39-42 | < 39 | 19 |
|  | LVEF | 58-76 | 56-57 | < 56 | 16 | 56-73 | | 51-55 | < 51 | 19 |
|  | LVMi | 33-59 | 60-68 | > 68 | 16 | 49-76 | | 77-83 | > 83 | 19 |
|  | LVM/LVEDV | 0.5-0.8 | 0.9-0.9 | > 0.9 | 16 | 0.6-1 | | 1.1-1.1 | > 1.1 | 19 |
|  | RVEDVi | 64-107 | 108-119 | > 119 | 16 | 73-110 | | 111-114 | > 114 | 19 |
|  | RVESVi | 24-51 | 52-58 | > 58 | 16 | 28-54 | | 55-60 | > 60 | 19 |
|  | RVEF | 45-68 | 41-44 | < 41 | 16 | 48-66 | | 45-47 | < 45 | 19 |
|  | RVSVi | 33-62 | 29-32 | < 29 | 16 | 42-65 | | 38-41 | < 38 | 19 |
|  | LAESVi | 20-48 | 49-54 | > 54 | 12 | 21-44 | | 45-48 | > 48 | 15 |
|  | LAEF | 55-79 | 49-54 | < 49 | 12 | 53-77 | | 50-52 | < 50 | 15 |
|  | RAESVi | 22-46 | 47-51 | > 51 | 11 | 24-63 | | 64-80 | > 80 | 15 |
|  | RAEF | 47-69 | 45-46 | < 45 | 11 | 39-64 | | 33-38 | < 33 | 14 |
| 30-39 | LVEDVi | 56-94 | 95-105 | > 105 | 18 | 61-94 | | 95-99 | > 99 | 17 |
|  | LVESVi | 15-36 | 37-45 | > 45 | 18 | 18-39 | | 40-48 | > 48 | 17 |
|  | LVSVi | 37-64 | 34-36 | < 34 | 18 | 40-63 | | 36-39 | < 36 | 17 |
|  | LVEF | 59-77 | 56-58 | < 56 | 18 | 56-75 | | 51-55 | < 51 | 17 |
|  | LVMi | 35-62 | 63-72 | > 72 | 18 | 48-77 | | 78-85 | > 85 | 17 |
|  | LVM/LVEDV | 0.5-0.8 | 0.9-0.9 | > 0.9 | 18 | 0.6-1 | | 1.1-1.2 | > 1.2 | 17 |
|  | RVEDVi | 60-103 | 104-114 | > 114 | 18 | 69-107 | | 108-112 | > 112 | 17 |
|  | RVESVi | 21-48 | 49-54 | > 54 | 18 | 25-53 | | 54-59 | > 59 | 17 |
|  | RVEF | 47-70 | 42-46 | < 42 | 18 | 48-68 | | 45-47 | < 45 | 17 |
|  | RVSVi | 32-62 | 28-31 | < 28 | 18 | 40-64 | | 36-39 | < 36 | 17 |
|  | LAESVi | 20-48 | 49-54 | > 54 | 16 | 20-43 | | 44-47 | > 47 | 17 |
|  | LAEF | 55-79 | 49-54 | < 49 | 16 | 53-78 | | 50-52 | < 50 | 17 |
|  | RAESVi | 22-48 | 49-54 | > 54 | 16 | 24-62 | | 63-74 | > 74 | 17 |
|  | RAEF | 48-71 | 45-47 | < 45 | 14 | 39-66 | | 33-38 | < 33 | 17 |
| 40-49 | LVEDVi | 53-89 | 90-98 | > 98 | 24 | 57-93 | | 94-99 | > 99 | 20 |
|  | LVESVi | 13-33 | 34-39 | > 39 | 24 | 17-37 | | 38-45 | > 45 | 20 |
|  | LVSVi | 36-61 | 34-35 | < 34 | 24 | 37-63 | | 33-36 | < 33 | 20 |
|  | LVEF | 60-79 | 57-59 | < 57 | 24 | 56-76 | | 52-55 | < 52 | 20 |
|  | LVMi | 37-65 | 66-77 | > 77 | 24 | 48-78 | | 79-88 | > 88 | 20 |
|  | LVM/LVEDV | 0.5-0.9 | 01-1 | > 1 | 24 | 0.6-1.1 | | 1.2-1.3 | > 1.3 | 20 |
|  | RVEDVi | 55-99 | 100-109 | > 109 | 24 | 64-104 | | 105-111 | > 111 | 20 |
|  | RVESVi | 17-43 | 44-49 | > 49 | 24 | 22-51 | | 52-56 | > 56 | 20 |
|  | RVEF | 48-73 | 43-47 | < 43 | 24 | 48-71 | | 45-47 | < 45 | 20 |
|  | RVSVi | 31-61 | 27-30 | < 27 | 24 | 37-64 | | 33-36 | < 33 | 20 |
|  | LAESVi | 20-48 | 49-54 | > 54 | 20 | 18-43 | | 44-47 | > 47 | 19 |
|  | LAEF | 55-80 | 50-54 | < 50 | 20 | 53-80 | | 49-52 | < 49 | 19 |
|  | RAESVi | 23-51 | 52-58 | > 58 | 21 | 23-61 | | 62-71 | > 71 | 20 |
|  | RAEF | 48-73 | 45-47 | < 45 | 20 | 39-68 | | 33-38 | < 33 | 20 |
| 50-59 | LVEDVi | 49-84 | 85-92 | > 92 | 66 | 53-92 | | 93-99 | > 99 | 49 |
|  | LVESVi | 11-30 | 31-35 | > 35 | 66 | 15-36 | | 37-42 | > 42 | 49 |
|  | LVSVi | 36-59 | 33-35 | < 33 | 66 | 34-64 | | 30-33 | < 30 | 49 |
|  | LVEF | 61-81 | 57-60 | < 57 | 66 | 56-78 | | 52-55 | < 52 | 49 |
|  | LVMi | 39-68 | 69-83 | > 83 | 66 | 47-80 | | 81-90 | > 90 | 49 |
|  | LVM/LVEDV | 0.6-1 | 1.1-1.2 | > 1.2 | 66 | 0.6-1.2 | | 1.3-1.3 | > 1.3 | 49 |
|  | RVEDVi | 51-94 | 95-103 | > 103 | 66 | 60-103 | | 104-111 | > 111 | 49 |
|  | RVESVi | 15-39 | 40-45 | > 45 | 66 | 19-47 | | 48-53 | > 53 | 49 |
|  | RVEF | 50-75 | 44-49 | < 44 | 66 | 48-74 | | 45-47 | < 45 | 49 |
|  | RVSVi | 30-61 | 26-29 | < 26 | 66 | 34-65 | | 30-33 | < 30 | 49 |
|  | LAESVi | 20-48 | 49-55 | > 55 | 59 | 17-42 | | 43-46 | > 46 | 45 |
|  | LAEF | 55-80 | 50-54 | < 50 | 58 | 53-81 | | 48-52 | < 48 | 45 |
|  | RAESVi | 23-54 | 55-62 | > 62 | 61 | 22-60 | | 61-68 | > 68 | 44 |
|  | RAEF | 47-74 | 44-46 | < 44 | 60 | 39-70 | | 33-38 | < 33 | 44 |
| 60-69 | LVEDVi | 46-80 | 81-87 | > 87 | 52 | 49-92 | | 93-101 | > 101 | 37 |
|  | LVESVi | 8-27 | 28-30 | > 30 | 52 | 13-34 | | 35-40 | > 40 | 37 |
|  | LVSVi | 34-59 | 31-33 | < 31 | 52 | 31-65 | | 27-30 | < 27 | 37 |
|  | LVEF | 61-83 | 57-60 | < 57 | 51 | 57-81 | | 53-56 | < 53 | 37 |
|  | LVMi | 41-70 | 71-88 | > 88 | 52 | 47-81 | | 82-94 | > 94 | 37 |
|  | LVM/LVEDV | 0.6-1.2 | 1.3-1.5 | > 1.5 | 52 | 0.6-1.3 | |  | > 1.4 | 37 |
|  | RVEDVi | 46-89 | 90-98 | > 98 | 52 | 56-103 | | 104-115 | > 115 | 37 |
|  | RVESVi | 12-35 | 36-41 | > 41 | 52 | 17-43 | | 44-47 | > 47 | 37 |
|  | RVEF | 52-78 | 45-51 | < 45 | 51 | 48-77 | | 45-47 | < 45 | 37 |
|  | RVSVi | 28-61 | 24-27 | < 24 | 52 | 31-68 | | 27-30 | < 27 | 37 |
|  | LAESVi | 19-49 | 50-58 | > 58 | 48 | 16-41 | | 42-45 | > 45 | 31 |
|  | LAEF | 54-81 | 49-53 | < 49 | 48 | 52-83 | | 47-51 | < 47 | 31 |
|  | RAESVi | 24-57 | 58-67 | > 67 | 45 | 20-60 | | 61-67 | > 67 | 32 |
|  | RAEF | 46-74 | 42-45 | < 42 | 45 | 39-72 | | 33-38 | < 33 | 31 |
| 70+ | LVEDVi | 43-76 | 77-82 | > 82 | 12 |  | |  |  |  |
|  | LVESVi | 6-24 | 25-27 | > 27 | 12 |  | |  |  |  |
|  | LVSVi | 32-60 | 28-31 | < 28 | 12 |  | |  |  |  |
|  | LVEF | 61-85 | 55-60 | < 55 | 12 |  | |  |  |  |
|  | LVMi | 43-73 | 74-93 | > 93 | 12 |  | |  |  |  |
|  | LVM/LVEDV | 0.7-1.4 | 1.5-1.7 | > 1.7 | 12 |  | |  |  |  |
|  | RVEDVi | 42-85 | 86-93 | > 93 | 12 |  | |  |  |  |
|  | RVESVi | 10-33 | 34-38 | > 38 | 12 |  | |  |  |  |
|  | RVEF | 53-80 | 46-52 | < 46 | 12 |  | |  |  |  |
|  | RVSVi | 27-61 | 23-26 | < 23 | 12 |  | |  |  |  |
|  | LAESVi | 18-53 | 54-65 | > 65 | 12 |  | |  |  |  |
|  | LAEF | 54-82 | 48-53 | < 48 | 12 |  | |  |  |  |
|  | RAESVi | 24-61 | 62-73 | > 73 | 10 |  | |  |  |  |
|  | RAEF | 44-73 | 41-43 | < 41 | 10 |  | |  |  |  |

Footnote: All measurements were derived using suiteHEART software with anatomical endocardial segmentation; papillary muscles and trabeculations were included in left ventricular mass measurements. Volumetric and mass measurements were indexed to height. Reference ranges are defined as the interval between the 2.5th and 97.5th centiles derived from age-specific generalized additive models for location, scale and shape (GAMLSS) using the Box–Cox Cole–Green distribution. Centile-based categories beyond the reference range are derived from LMS-based thresholds (P0.5, P2.5, P97.5, and P99.5) and describe the position of a measurement within the reference distribution rather than disease severity or diagnostic abnormality. Reference ranges are based on a Chinese adult population.

Abbreviations: LV = left ventricle; RV = right ventricle; LA = left atrium; RA = right atrium; EDV = end-diastolic volume; ESV = end-systolic volume; SV = stroke volume; EF = ejection fraction; CO = cardiac output.

### 50. Table: Reference ranges and centile-based categories for cardiac chamber measurements derived using suitheart software (anatomical segmentation), indexed to BSA, in a Mixed adult population

|  |  | **Female** | | | | **Male** | | | |
| --- | --- | --- | --- | --- | --- | --- | --- | --- | --- |
| **Age group** | **Metrics** | **Reference range** | **Mild** | **At least moderate** | **n** | **Reference range** | **Mild** | **At least moderate** | **n** |
| 50-59 | LVEDVi | 51-87 | 88-95 | > 95 | 134 | 53-104 | 105-113 | > 113 | 66 |
|  | LVESVi | 10-30 | 31-33 | > 33 | 134 | 11-35 | 36-38 | > 38 | 66 |
|  | LVSVi | 35-61 | 32-34 | < 32 | 133 | 36-73 | 31-35 | < 31 | 66 |
|  | LVEF | 60-81 | 56-59 | < 56 | 133 | 58-82 | 55-57 | < 55 | 66 |
|  | LVMi | 37-67 | 68-73 | > 73 | 134 | 47-82 | 83-87 | > 87 | 62 |
|  | LVM/LVEDV | 0.6-1.1 |  | > 1.2 | 134 | 0.6-1.3 | 1.4-1.9 | > 1.9 | 62 |
|  | RVEDVi | 49-89 | 90-96 | > 96 | 134 | 59-110 | 111-119 | > 119 | 66 |
|  | RVESVi | 14-37 | 38-41 | > 41 | 134 | 17-45 | 46-49 | > 49 | 66 |
|  | RVEF | 54-76 | 51-53 | < 51 | 133 | 47-76 | 40-46 | < 40 | 66 |
|  | RVSVi | 30-60 | 25-29 | < 25 | 134 | 34-72 | 29-33 | < 29 | 66 |
|  | LAESVi | 18-44 | 45-48 | > 48 | 122 | 18-50 | 51-57 | > 57 | 53 |
|  | LAEF | 57-80 | 51-56 | < 51 | 122 | 56-81 | 51-55 | < 51 | 54 |
|  | RAESVi | 21-55 | 56-63 | > 63 | 117 | 24-60 | 61-67 | > 67 | 54 |
|  | RAEF | 41-72 | 37-40 | < 37 | 117 | 37-72 | 32-36 | < 32 | 54 |
| 60-69 | LVEDVi | 44-84 | 85-90 | > 90 | 110 | 46-98 | 99-108 | > 108 | 59 |
|  | LVESVi | 8-29 | 30-33 | > 33 | 110 | 11-36 | 37-41 | > 41 | 59 |
|  | LVSVi | 32-61 | 29-31 | < 29 | 110 | 32-66 | 26-31 | < 26 | 59 |
|  | LVEF | 60-83 | 55-59 | < 55 | 109 | 56-80 | 51-55 | < 51 | 59 |
|  | LVMi | 37-69 | 70-75 | > 75 | 110 | 47-82 | 83-88 | > 88 | 58 |
|  | LVM/LVEDV | 0.6-1.1 | 1.2-1.3 | > 1.3 | 110 | 0.7-1.4 | 1.5-1.8 | > 1.8 | 58 |
|  | RVEDVi | 45-93 | 94-101 | > 101 | 110 | 52-112 | 113-124 | > 124 | 59 |
|  | RVESVi | 14-34 | 35-39 | > 39 | 110 | 17-48 | 49-56 | > 56 | 59 |
|  | RVEF | 54-77 | 50-53 | < 50 | 110 | 49-75 | 45-48 | < 45 | 59 |
|  | RVSVi | 28-63 | 22-27 | < 22 | 110 | 30-70 | 24-29 | < 24 | 58 |
|  | LAESVi | 18-47 | 48-52 | > 52 | 100 | 19-45 | 46-50 | > 50 | 53 |
|  | LAEF | 55-81 | 50-54 | < 50 | 100 | 52-81 | 48-51 | < 48 | 53 |
|  | RAESVi | 23-56 | 57-64 | > 64 | 93 | 22-62 | 63-69 | > 69 | 40 |
|  | RAEF | 40-73 | 35-39 | < 35 | 93 | 37-72 | 32-36 | < 32 | 40 |
| 70+ | LVEDVi | 42-86 | 87-95 | > 95 | 50 | 41-93 | 94-102 | > 102 | 34 |
|  | LVESVi | 8-28 | 29-32 | > 32 | 51 | 10-40 | 41-51 | > 51 | 34 |
|  | LVSVi | 28-59 | 23-27 | < 23 | 51 | 28-60 | 22-27 | < 22 | 34 |
|  | LVEF | 59-84 | 54-58 | < 54 | 51 | 53-79 | 43-52 | < 43 | 35 |
|  | LVMi | 38-70 | 71-77 | > 77 | 51 | 47-83 | 84-90 | > 90 | 33 |
|  | LVM/LVEDV | 0.6-1.2 | 1.3-1.3 | > 1.3 | 50 | 0.6-1.5 | 1.6-1.8 | > 1.8 | 33 |
|  | RVEDVi | 50-86 | 87-92 | > 92 | 51 | 46-116 | 117-132 | > 132 | 35 |
|  | RVESVi | 13-32 | 33-37 | > 37 | 51 | 18-50 | 51-64 | > 64 | 35 |
|  | RVEF | 54-79 | 48-53 | < 48 | 51 | 50-74 | 47-49 | < 47 | 35 |
|  | RVSVi | 29-63 | 24-28 | < 24 | 51 | 26-69 | 20-25 | < 20 | 33 |
|  | LAESVi | 17-50 | 51-55 | > 55 | 43 | 15-51 | 52-61 | > 61 | 34 |
|  | LAEF | 53-83 | 49-52 | < 49 | 42 | 50-83 | 45-49 | < 45 | 34 |
|  | RAESVi | 25-54 | 55-62 | > 62 | 36 | 19-64 | 65-72 | > 72 | 21 |
|  | RAEF | 38-74 | 33-37 | < 33 | 34 | 35-68 | 30-34 | < 30 | 21 |

Footnote: All measurements were derived using suiteHEART software with anatomical endocardial segmentation; papillary muscles and trabeculations were included in left ventricular mass measurements. Volumetric and mass measurements were indexed to height. Reference ranges are defined as the interval between the 2.5th and 97.5th centiles derived from age-specific generalized additive models for location, scale and shape (GAMLSS) using the Box–Cox Cole–Green distribution. Centile-based categories beyond the reference range are derived from LMS-based thresholds (P0.5, P2.5, P97.5, and P99.5) and describe the position of a measurement within the reference distribution rather than disease severity or diagnostic abnormality. Reference ranges are based on a Mixed adult population.

Abbreviations: LV = left ventricle; RV = right ventricle; LA = left atrium; RA = right atrium; EDV = end-diastolic volume; ESV = end-systolic volume; SV = stroke volume; EF = ejection fraction; CO = cardiac output.
